# Comparative Effectiveness of Semaglutide and Tirzepatide for Weight Loss in Adults with Overweight and Obesity in the US: A Real-World Evidence Study

**DOI:** 10.1101/2023.11.21.23298775

**Authors:** Patricia J Rodriguez, Brianna M Goodwin Cartwright, Samuel Gratzl, Rajdeep Brar, Charlotte Baker, Ty J. Gluckman, Nicholas L Stucky

## Abstract

**Background:** Both tirzepatide and semaglutide have been shown to reduce weight for patients with overweight or obesity in randomized controlled trials (RCTs). While tirzepatide appears to provide greater weight loss than semaglutide in this population, head-to-head RCTs are not yet available. Accordingly, we sought to compare on-treatment weight loss in a real-world setting for adults with overweight or obesity initiated on tirzepatide or semaglutide.

**Methods:** Adults with overweight or obesity first dispensed semaglutide or tirzepatide between May 2022 and September 2023 were identified from Truveta Data, a large electronic health record (EHR) dataset linked with comprehensive pharmacy dispensing data. The study cohort was restricted to patients with no prior GLP-1 RA use, who initiated a formulation of semaglutide or tirzepatide labelled for type 2 diabetes mellitus (T2D) (a proxy for dose), received regular care in the previous year, had a GLP-1 RA prescription written in the 60 days prior to initiation, and had an available baseline weight. On treatment weight changes in a propensity score matched population were compared for outcomes of time to 5%, 10%, and 15% weight loss, and percentage change in weight by 3, 6, and 12 months. For all outcomes, we conducted subgroup analyses stratified by T2D (on-label users) and sensitivity analyses using inverse probability of treatment weighting.

**Results:** A total of 41,223 patients met our cohort definition (semaglutide: 32,030; tirzepatide: 9,193). Propensity score matching produced an analytic cohort of 18,386 patients with good balance on all baseline covariates. At treatment initiation, the mean(SD) age was 52.0 (12.9) years, 70.5% of patients were female, 51.7% had T2D, and mean(SD) weight was 110 (25.7) kg. A larger proportion of patients on tirzepatide, compared to semaglutide, achieved weight reductions ≥5% (81.8% vs. 64.6%), ≥10% (62.1% vs. 38.0%), and ≥15% (42.3% vs 19.3%) within 1 year on treatment, yielding hazard ratios of 1.76 (1.68 - 1.85) for 5%, 2.42 (2.25 - 2.59) for 10%, and 3.04 (2.73 - 3.38) for 15% weight loss. Patients on tirzepatide experienced larger changes in percentage of body weight lost at 3 months on treatment (difference [95% CI]) (−2.3% [-2.5%, -2.2%]), 6 months on treatment (−4.3% [-4.6%, -3.9%]), and 12 months on treatment (−7.2% [-8.3%, -6.2%]). Hazards for all gastrointestinal (GI) adverse events were similar between groups.

**Conclusion:** In a real-world population of US adults with overweight or obesity initiated on tirzepatide or semaglutide formulations labelled for T2D, patients on tirzepatide were significantly more likely to achieve 5%, 10% and 15% weight loss and experience larger reductions in weight at 3, 6, and 12 months. Findings were robust to analytic choice and consistent among populations stratified by T2D. Future work is needed to understand whether these findings result in a differential impact on other outcomes, including rates of adverse cardiovascular events.

## Introduction

Overweight and obesity are highly prevalent conditions associated with increased morbidity and mortality [1–3]. Historically, pharmacologic treatments for weight reduction (anti-obesity medications [AOMs]) have been limited in number, poor tolerated, and modest in their impact on weight [4–6]. However, newer therapies, including the glucagon-like peptide 1 receptor agonist (GLP-1 RA) semaglutide and the dual GLP-1 RA/gastric inhibitory polypeptide (GIP) agonist tirzepatide, have demonstrated substantial weight reduction in patients with obesity, with and without type 2 diabetes mellitus (T2D), in randomized controlled trials (RCTs) [7–10].

While tirzepatide produces greater weight loss than semaglutide in patients with T2D [11], head-to-head trials evaluating these therapies in patients with overweight or obesity are not yet available. Further, it remains unclear whether the magnitude of weight loss observed in real-world settings mirrors that observed in RCTs, given well-described differences between these populations [12–14]. Finally, because these medications are costly and insurance coverage is limited for patients without T2D, real-world adherence may be reduced, attenuating the treatment effect in real-world settings.

We aimed to compare on-treatment weight change between tirzepatide and semaglutide (injectable) labelled for T2D in a large, real-world population. We quantify differences in (1) likelihood of achieving 5%, 10%, and 15% weight loss, and (2) percentage change in body weight at 3, 6, and 12 months on treatment. We also compared the risk of bowel obstruction, cholecystitis, cholelithiasis, gastroenteritis, gastroparesis, and pancreatitis between tirzepatide and semaglutide.

## Methods

### Study Design

New users of tirzepatide or injectable semaglutide with overweight or obesity (regardless of T2D) were enrolled in our study at the time of their first dispensed prescription. The first dispense was considered the treatment initiation date and served as the index date for our study. New users were defined as those having no previous dispense of any GLP-1 RA or GLP-1 RA/GIP agonist (supplement). Only adult patients with regular interactions with the health care system and an available baseline weight were included (detailed inclusion and exclusion criteria in Study Population below). Patients were followed for weight loss and gastrointestinal adverse event outcomes until the first of discontinuation of therapy, GLP-1 RA switching, or administrative censoring.

### Data

This study used a subset of Truveta Data. Truveta provides access to continuously updated and linked electronic health record (EHR) from a collective of US health care systems, including structured information on demographics, encounters, diagnoses, vital signs (e.g., weight, BMI, blood pressure), medication requests (prescriptions), medication administration, laboratory and diagnostic tests and results (e.g., HbA1c tests and values), and procedures. Updated EHR data are provided daily to Truveta by constituent health care systems. In addition to EHR data for care delivered within Truveta constituent health care systems, medication dispensing and social drivers of health (SDOH) information are made available through linked third-party data. Medication dispense (via e-prescribing data) includes fills for prescriptions written both within and outside Truveta constituent health care systems, providing greater observability into patients’ medication history. Medication dispense histories are updated at the time of the encounter, and include fill dates, NDC or RxNorm codes, quantity dispensed, and days of medication supplied. SDOH data include individual-level factors, including income and education.

Data are normalized into a common data model through syntactic and semantic normalization. Truveta Data are then de-identified by expert determination under the HIPAA Privacy Rule. Once de-identified, data are available for analysis in R or Python using Truveta Studio. Data for this study were accessed on November 3, 2023.

### Study population and exposure

We identified adults first dispensed tirzepatide or injectable semaglutide labelled for T2D (as Mounjaro or Ozempic, respectively) between May 2022 (the month of tirzepatide approval for T2D) and September 30, 2023, who had overweight (body mass index [BMI] ≥ 27 *kg/m*^2^or a diagnosis code indicating BMI ≥ 27 *kg/m*^2^) or obesity (BMI ≥ 30 *kg/m*^2^ or a diagnosis code for obesity) in the 12 months before their index date. We required a complete negative history of GLP-1 RA and GLP-1 RA/GIP agonist use. To improve outcome observability, we limited our analysis to patients with regular interactions with the health care system during the year prior to their index date: at least one encounter, observation, or medication request in each consecutive 6-month period preceding the index date. We required a GLP-1 RA or GLP-1 RA/GIP agonist prescription written and a baseline weight measurement in the 60 days before the index date. A 60-day window was selected because insurance denials and appeals processes for these medications may result in unusually long times between GLP-1 RA prescribing and filling. Of note, the GLP-1 RA or GLP-1 RA/GIP agonist prescribed was not required to match the medication first dispensed, given that drug shortages during the study period [15] may have resulted in substitutions at the pharmacy. Patients were categorized according to the medication dispensed. Additional exclusions were made for patients with missing sex and those with no follow-up time.

We relied on brand as a proxy for target dose. The standard dose is 0.5 mg for injectable semaglutide labelled for T2D and 5mg for tirzepatide (labelled exclusively for T2D at the time of this analysis). Both drugs follow dose escalation schedules, with initiation at the lowest dose and titration to the highest tolerated dose over weeks to months.

### Patient Comorbidities and Covariates

Patients were classified as having T2D if they had a T2D diagnosis, were prescribed, administered, or dispensed insulin or a dipeptidyl peptidase 4 (DPP-4) inhibitor, or had an HbA1c level ≥7.5% in the two years before their index date. Baseline patient demographics, clinical comorbidities, use of other anti-diabetic medication (ADM) and AOM, and history of bariatric surgery in the 2 years before the index date were assessed. Several steps were taken to standardize weight data from the EHR, including the removal of apparent data entry or unit conversation errors (described in supplement). The most recent weight within the 60 days before index date was considered the baseline value. Codes for all data definitions are provided in the supplement.

### Weight Outcomes

Our primary estimand of interest was on-treatment weight loss. Therefore, patients were censored at the first of: treatment discontinuation (≥30 days without medication on hand), GLP-1 RA or GLP-1 RA/GIP agonist switching (change to a difference substance; brand changes were allowed), or administrative censoring (last encounter).

Propensity scores were used to balance treatment groups on measured variables. Propensity scores estimated the probability of initiating tirzepatide, compared to semaglutide, as a function of demographic, clinical, and utilization characteristics. Patients were then matched using 1:1 nearest neighbor propensity score (PS) matching. Balance was assessed by standardized mean differences, with an acceptable threshold of 0.1. To provide further control for residual confounding, age, presence of T2D, and baseline weight were included as covariates in all parametric and semi-parametric models.

Percentage changes in body weight were calculated as: (follow-up weight - baseline weight)/baseline weight. Event probabilities for 5%, 10%, and 15% weight loss and median time to each event were extracted from Kaplan-Meier models. Differences in the hazard of achieving the specified weight loss threshold for those on tirzepatide compared to semaglutide were estimated using Cox proportional hazards models.

For weight change at 3, 6, and 12 months, we considered the subpopulation still at risk (not yet censored) at the timepoint of interest. The weight value nearest to the timepoint, up to 45 days before or after, was considered the outcome value. For at-risk patients without a weight value in this window, multiple imputation was used to impute weight change. Imputation used information from the full at risk-population and all measured covariates and outcomes. Within each (m = 5) imputed dataset of patients still at risk at the timepoint of interest, propensity score matching was re-applied as described above, and differences in percentage weight loss were estimated using linear models. Estimates were then pooled across imputations using Rubin’s rules [16].

### Sensitivity Analyses

Several sensitivity analyses were performed to test the robustness of findings. First, we replicated all analyses using inverse probability of treatment weighting (IPTW), rather than 1:1 propensity score matching. Second, we conducted stratified analyses for patients with and without T2D, replicating the full process described above for each strata. Finally, we conducted a modified intention-to-treat (ITT) analysis, where censoring time was based on administrative censoring only. The modified ITT analysis included all available follow-up weights regardless of whether the patient was still on treatment.

### Safety Outcomes

Incidence rates of gastrointestinal adverse events (AEs) (bowel obstruction, cholecystitis, cholelithiasis, gastroenteritis, gastroparesis, and pancreatitis) per 1,000 person years at risk were calculated, censoring patients at the time of switching, discontinuation, or administrative censoring. Patients with a history of the specified AE in the 12 months before index were not considered at risk and were excluded from analyses of the specified AE. Differences in the hazard of each AE between tirzepatide and semaglutide were estimated using Cox proportional hazards models.

### Stats program and packages used

All analyses were conducted in R using the following packages: rlang, arrow, dplyr, tidyr, lubridate, forcats, table1, cobalt, MatchIt, WeightIt, mice, MatchThem, survey, survival, ggsurvfit, broom, ggplot2, and xtable [17–32].

## Results

### Patient Characteristics

In total, 41,223 patients met our inclusion criteria (tirzepatide: 9,193, semaglutide: 32,030) (Figure 1). Prior to propensity score matching, patients initiated on tirzepatide, compared to semaglutide, were younger and a higher proportion were female, white, and had at least a college education (Table 1). Patients initiated on tirzepatide had a lower prevalence of T2D and most other comorbidities. Despite clinical and demographic differences, mean baseline weight was similar between groups (110 kg for tirzepatide vs. 109 kg for semaglutide), with measurement occurring an average (median) of 9.3 (4.0) days before treatment initiation. The 1:1 PS matched analytic cohort included 18,386 patients, with standardized mean differences for all variables <0.05.

**Table 1:**
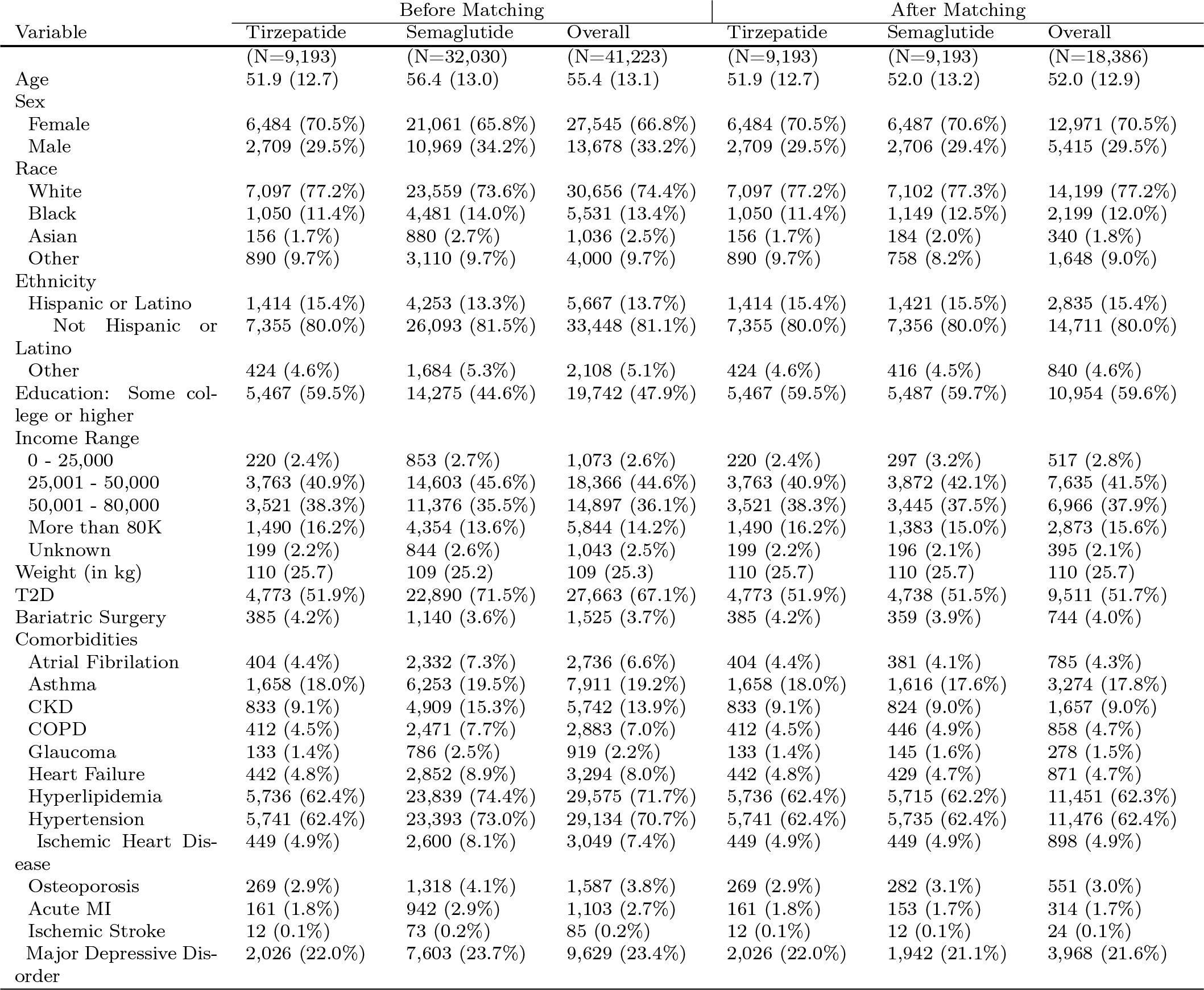
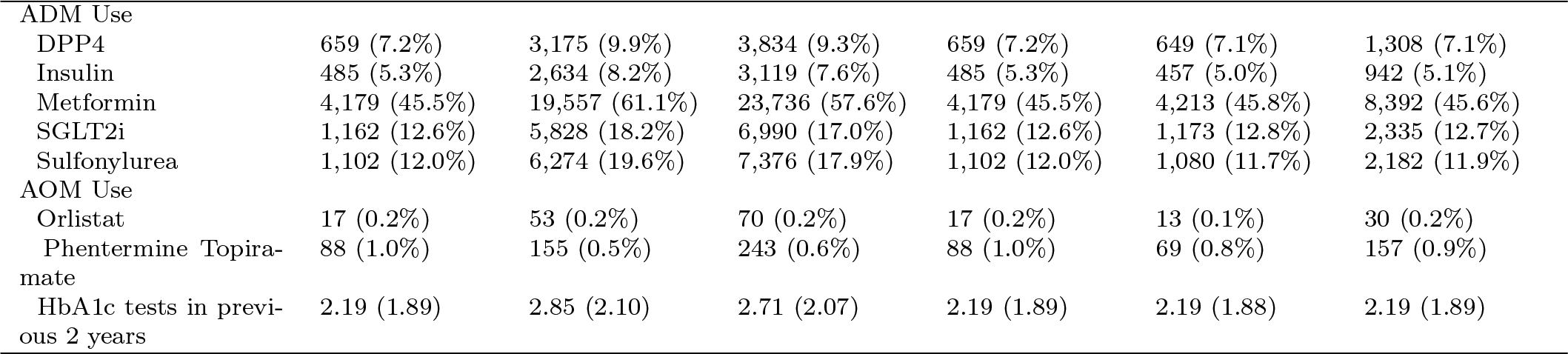
Characteristics of study population before and after propensity score matching. Quantitative variables expressed as mean(standard deviation). Categorical variables expressed as number(percentage). Abbreviations: ADM = anti-diabetic medication, AOM = anti-obesity medication, Black = Black or African American, BMI = body mass index, CKD = chronic kidney disease, COPD = chronic obstructive pulmonary disease, DPP4 = dipeptidyl peptidase 4 inhibitor, MI = myocardial infarction, SD = standard deviation, SGLT2i = sodium/glucose cotransporter-2 inhibitor, T2D = type 2 diabetes mellitus.

**Figure 1:**
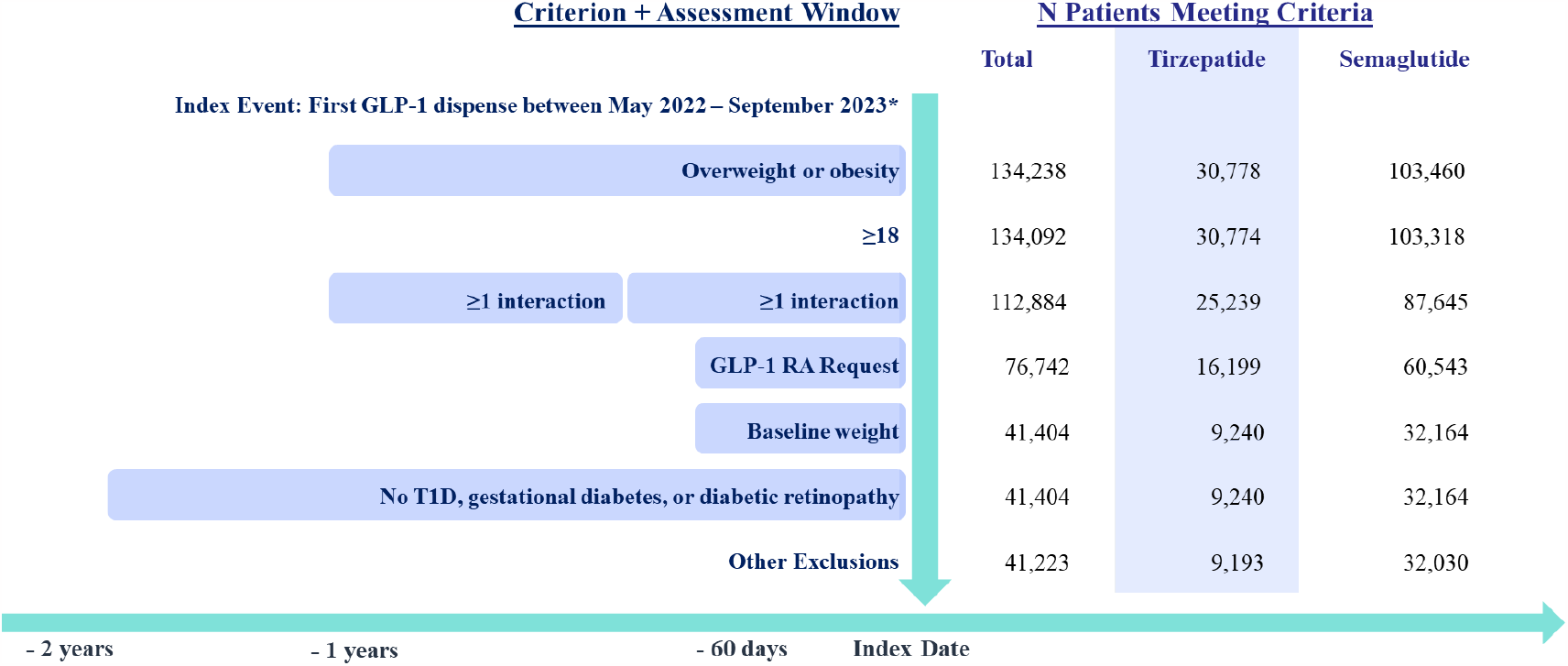
Consort diagram. Other exclusions include missing sex and no follow-up time.

The mean (median) duration of follow-up on treatment was 167 (133) days. Follow-up was ended by discontinuation for 55.1% (55.7% for tirzepatide and 54.4% for semaglutide), medication switching for 0.9% (1.4% for tirzepatide and 0.4% for semaglutide) and administrative censoring for 44.1% (42.9% for tirzepatide and 45.2% for semaglutide). The mean (median) duration of follow-up with administrative censoring alone (modified ITT analysis) was 263 (267) days.

### Time to Weight Change

Among the matched population at risk (on treatment), 81.8% (95% CI: 79.8% -83.7%) of patients on tirzepatide vs. 66.8% (64.6% -68.8%) on semaglutide achieved ≥5% weight loss, 62.1% (59.7% -64.3%) vs. 38.0% (35.7% -40.2%) achieved ≥10% weight loss, and 42.3% (39.8% -44.6%) vs. 19.3% (17.4% -21.2%) achieved ≥15% weight loss within 365 days (Figure 2). The median time to 5% weight loss was 134 (131-140) days for tirzepatide and 215 (205-226) days for semaglutide. Resulting hazard ratios comparing tirzepatide to semaglutide were 1.76 (1.68 - 1.85) for 5% weight loss, 2.42 (2.25 - 2.59) for ≥10% weight loss and 3.04 (2.73 - 3.38) for ≥15% weight loss (Figure 4).

**Figure 2:**
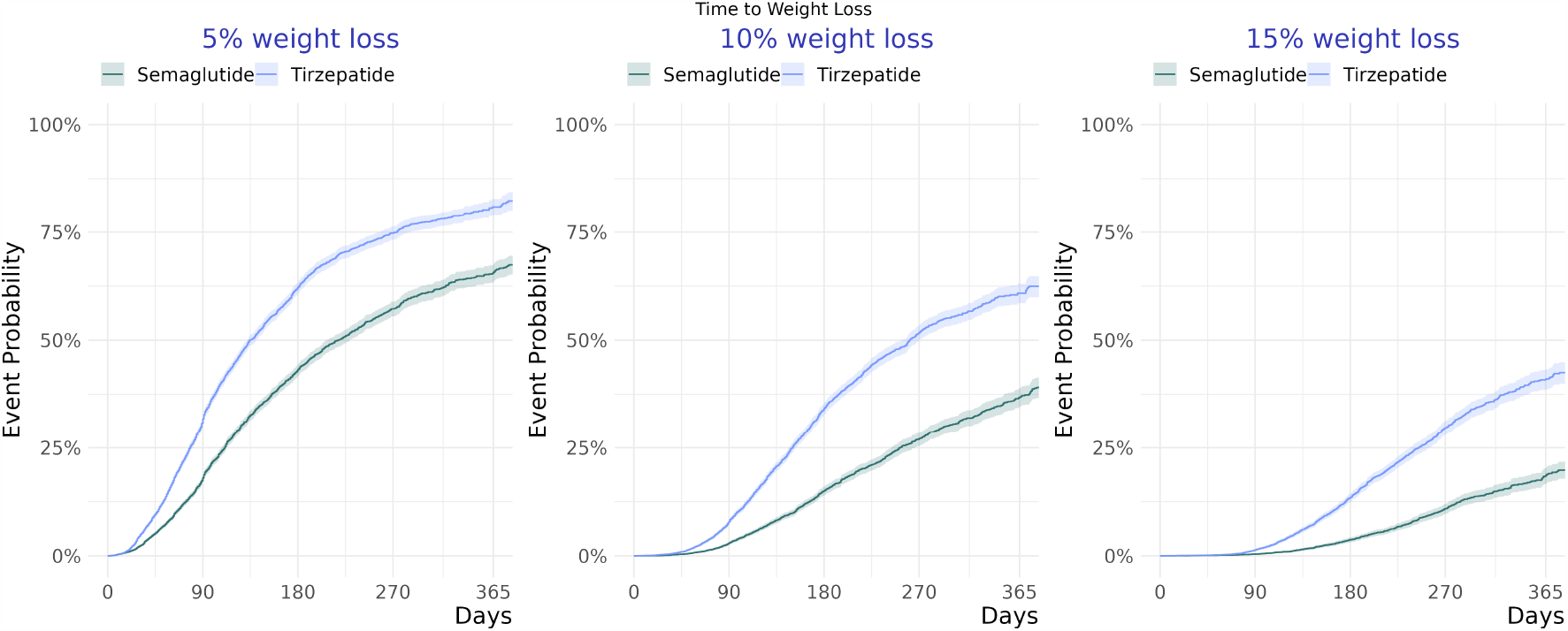
Time to weight reduction for propensity-score matched patients on treatment. Y-axis represents the event probability (1-survival probability).

### Percentage Change in Body Weight

The mean percentage change in body weight was -5.9% (−6.0%, -5.8%) for tirzepatide vs. -3.6% (−3.7%, -3.5%) for semaglutide at 3 months on treatment, -10.1% (−10.4%, -9.9%) vs. -5.9% (−6.1%, -5.6%) at 6 months on treatment, and -15.2% (−16.0%, -14.4%) vs. -7.9% (−8.6%, -7.2%) at 12 months on treatment (Figure 3). After adjusting for residual confounding, the absolute difference in weight loss between tirzepatide and semaglutide was -2.3% (−2.5%, -2.2%), -4.3% (−4.6%, -3.9%), and -7.2% (−8.3%, -6.2%) at 3, 6, and 12 months on treatment, respectively (Figure 5).

**Figure 3:**
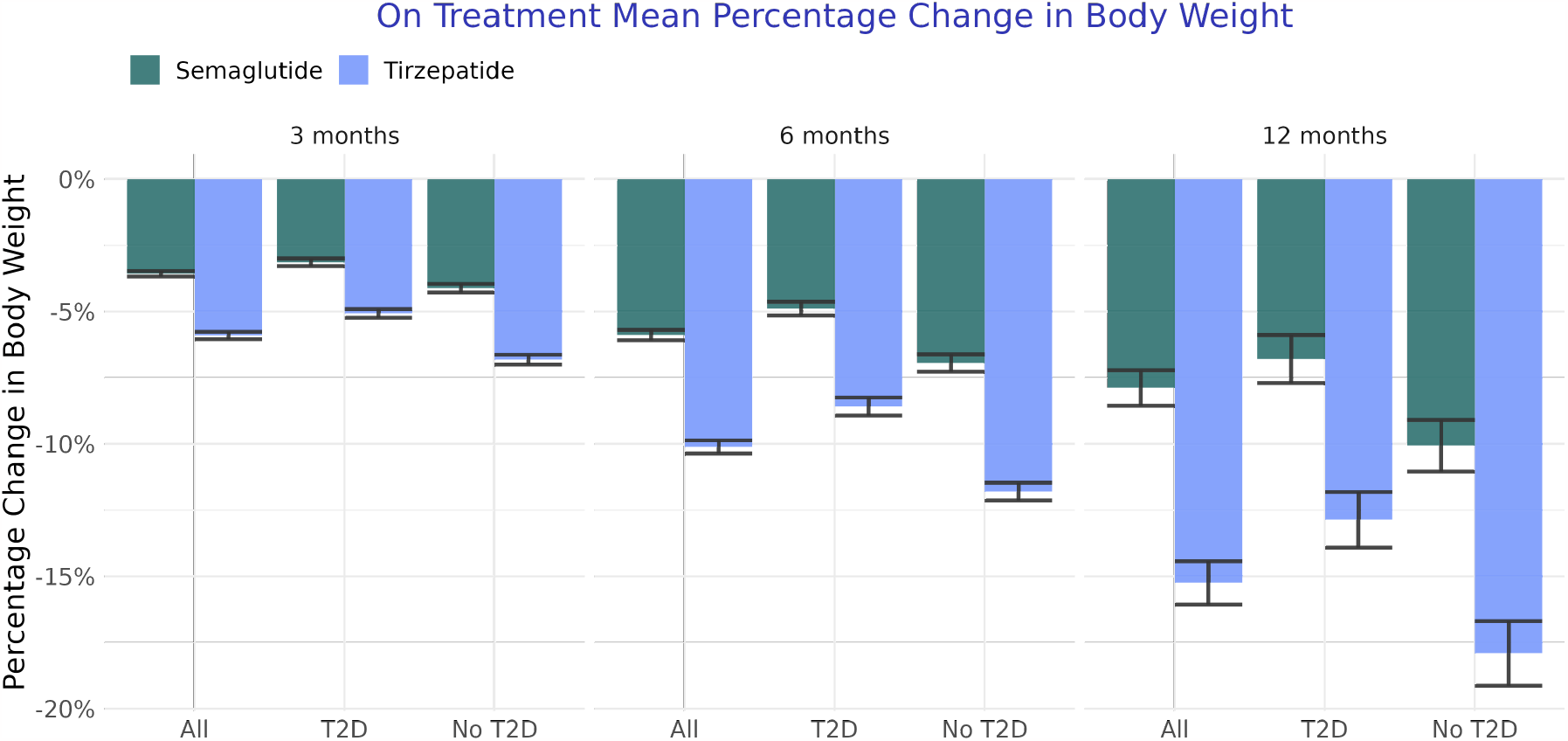
Mean percentage change in body weight at 3, 6 and 12 months on treatment for the overall population, those with T2D, and those without T2D.

**Figure 4:**
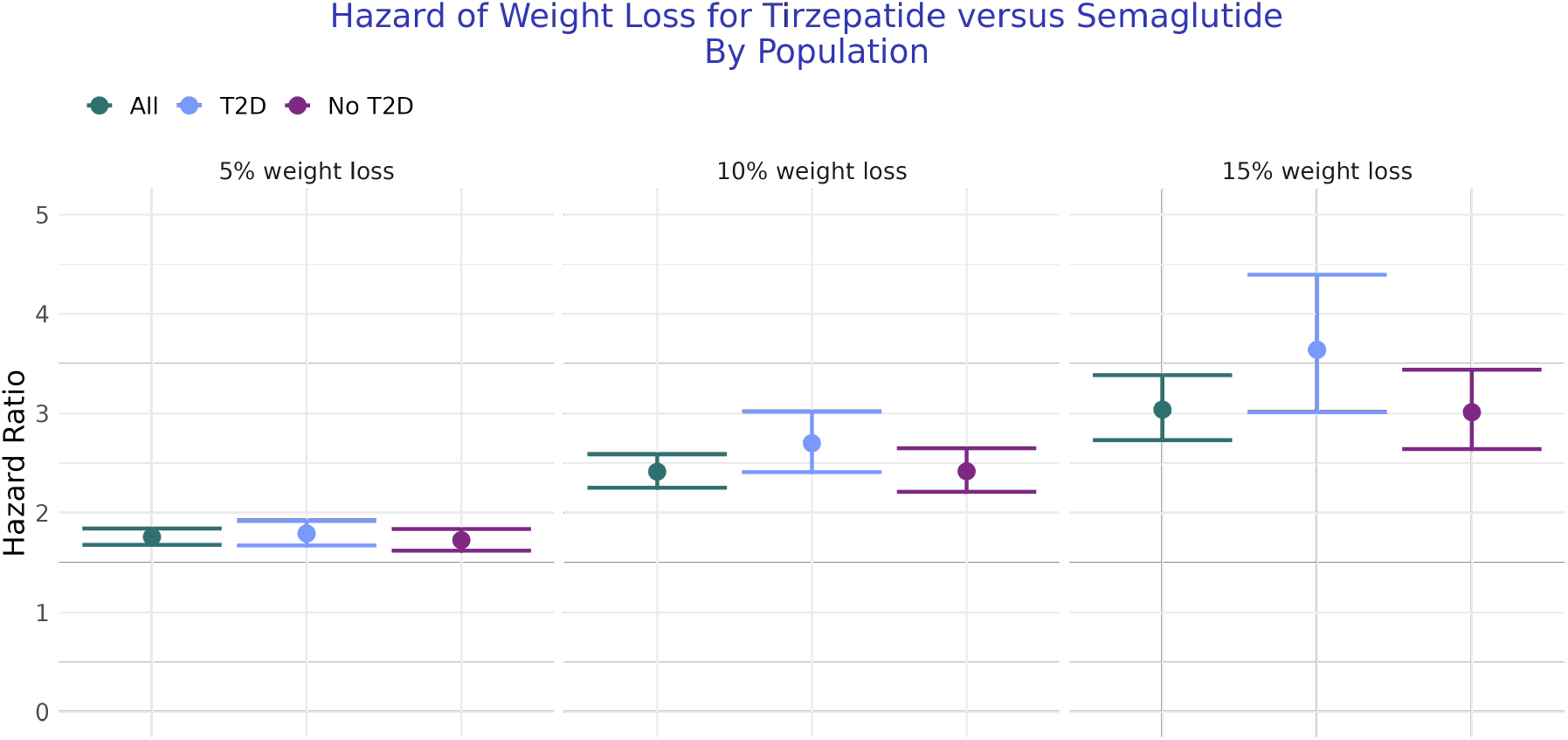
Treatment effects (hazard Ratios) for the overall population, T2D population, and population without T2D. Estimates compare the hazard of achieving ≥5%, ≥10%, and ≥15% weight loss for patients on treatment with tirzepatide vs on treatment with semaglutide, among PS-matched populations. Hazard ratios >1 indicate higher effectiveness of tirzepatide for reaching weight loss threshold. Points represent point estimates; lines represent 95% confidence interval.)

**Figure 5:**
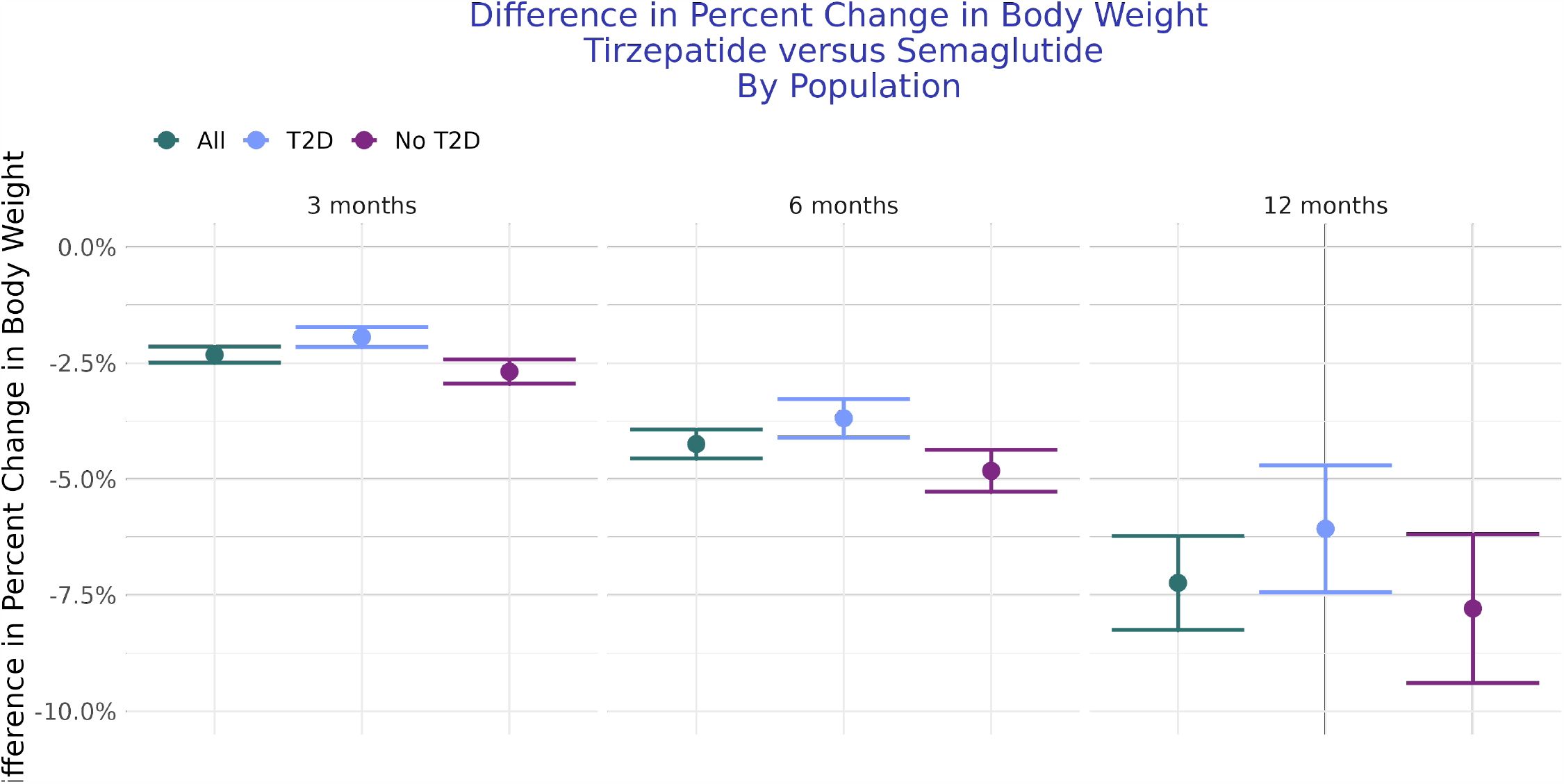
Treatment effects (absolute differences in body weight change) for the overall population, T2D population, and population without T2D. Estimates compare the absolute difference in body weight change at 3 months, 6 months, and 12 months on treatment with tirzepatide vs. semaglutide, among PS-matched populations still on treatment at the timepoint. Negative differences indicate greater weight loss for tirzepatide. Points represent point estimates; lines represent 95% confidence interval.

### Sensitivity Analyses

Modified ITT analyses resulted in fewer patients achieving weight loss targets, smaller weight reductions, and slightly attenuated treatment effects, though tirzepatide remained significantly more effective than semaglutide in all analyses (supplement). A smaller proportion achieved weight loss ≥5%, 71.1% (69.9% -72.3%) on tirzepatide and 56.3% (54.9% -57.6%) on semaglutide, resulting in a hazard ratio of 1.6 (1.6-1.7). Similarly, mean percentage changes in body weight were smaller: -5.3% (−5.4%, -5.2%) for tirzepatide vs. -3.3% (−3.4%, -3.2%) for semaglutide at 3 months, -8.2% (−8.4%, -8.0%) for tirzepatide vs. -4.9% (−5%, -4.7%) for semaglutide at 6 months, and -11.2% (−11.7%, -10.8%) for tirzepatide vs. -6.0% (−6.4%, -5.6%) for semaglutide at 12 months. After adjusting for residual confounding, the difference in weight loss between tirzepatide and semaglutide was -2.0% (−2.2%, -1.8%) at 3 months, -3.4% (−3.6%, -3.1%) at 6 months, and -5.2% (−5.9%, -4.6%) at 12 months.

Sensitivity analyses using inverse probability of treatment weighting produced highly similar results (supplement).

### Subgroup Analyses

In stratified analyses, those without T2D experienced larger reductions in body weight than those with T2D for tirzepatide and semaglutide alike (supplement). Tirzepatide remained significantly more effective in lowering weight than semaglutide in all analyses among those with and without T2D (Figures 4 and 5).

### Gastrointestinal Adverse Events

We observed no significant differences in the risk of GI AEs. Gastroenteritis was the most common GI AE, with 21.0 and 19.1 cases per 1,000 person-years at risk for tirzepatide and semaglutide, respectively (Table 2).

**Table 2:** Gastrointestinal Adverse Event Rates Per 1000 person-years.

| Adverse Event | Tirzepatide | Semaglutide | Harzard Ratio (CI) |
| --- | --- | --- | --- |
| Bowel Obstruction | 6.52 | 3.95 | 1.65 (0.86-3.16) |
| Cholecystitis | 6.79 | 5.00 | 1.33 (0.73-2.42) |
| Cholelithiasis | 13.41 | 15.39 | 0.88 (0.6-1.28) |
| Gastroenteritis | 21.09 | 19.14 | 1.12 (0.81-1.55) |
| Gastroparesis | 4.06 | 4.73 | 0.86 (0.43-1.7) |
| Pancreatitis | 3.79 | 3.41 | 1.08 (0.51-2.29) |

## Discussion

In this large, real-world analysis of US adults with overweight or obesity initiated on tirzepatide or semaglutide, those on tirzepatide were more likely to achieve 5%, 10% and 15% weight loss and experienced larger reductions in body weight at 3, 6, and 12 months. To our knowledge, this study represents the first real-world comparative effectiveness study of tirzepatide and semaglutide in adults with overweight or obesity. Treatment effects were consistent in direction and significance between methodological approaches (PS matching, IPTW, modified ITT) and within the subgroup of patients with T2D. No significant differences in the incidence of GI AEs were observed.

Findings in this study are broadly consistent with existing evidence from RCTs. Among placebo-controlled trials of patients with obesity, treatment with tirzepatide at 10 mg per week resulted in 82% and 96% of individuals with and without T2D achieving 5% body weight by 72 weeks, respectively (efficiacy estimands)[9, 10]. Likewise, among similarly designed placebo-controlled trials, treatment with semaglutide at 2.4 mg per week resulted in 73% and 92% of individuals with and without T2D achieving ≥5% body weight by 68 weeks, respectively (efficacy estimands) [8, 33]. While data from head-to-head trials are more limited, a single study that evaluated the glucose-lowering effect of tirzepatide (5 mg per week) compared to semaglutide (1 mg per week) when added to metformin in patients with T2D found that 5% weight loss was achieved by 69% and 58%, respectively [11]. Importantly, a trial comparing tirzepatide to semaglutide in patients with overweight or obesity, but without T2D is underway (SURMOUNT-5, NCT05822830); however, results are not expected until late 2024.

This study has several strengths. First, the analysis included a large cohort of patients with overweight and obesity evaluated after tirzepatide’s approval (May 2022). In fact, it’s likely that the weight reduction observed in our study was greater than that found in previous real-world studies because such studies ended before semaglutide and/or tirzepatide were available [34, 35]. Second, treatment effects were consistent in direction and significance between estimands (on treatment vs. ITT), subgroups (with vs. without T2D), and methodological approaches (PS matching, IPTW, complete case analysis). Third, our study included some populations who were ineligible for clinical trials, such as those with active or unstable major depressive disorder (MDD) in the previous 2 years. MDD was common in our population (22% of patients had a history in previous 2 years) suggesting clinical trials may have excluded a large proportion of patients using these medications in real-world settings. Finally use of prescribing and dispensing data allowed us to include populations without T2D, which may not be captured in pharmacy claims data alone given off-label use of these medications may not be covered by insurers.

Our study is also subject to several limitations. Unlike many clinical endpoints, weight loss is directly observable to patients, which may result in informative censoring [35, 36]. Those observing desired weight loss may be more likely to continue treatment, while those observing no weight change may be more likely to discontinue or switch drugs. While a modified ITT analysis inclusive of post-discontinuation weights showed smaller reductions in weight, differences between tirzepatide and semaglutide were similar. Generalizability of the ITT estimand may be limited, however, given medication shortages during the study period which may have resulted in higher discontinuation than expected under normal conditions. In addition, it’s possible that unmeasured confounding, especially the degree of motivation for weight loss, exists. A substantial amount of unmeasured confounding, though, would be required to negate the treatment effects observed in this study. Finally, this study included medications labelled for T2D only. Future studies should be conducted to compare the effects of medications labelled for weight loss, which have higher doses on average.

## Conclusions

In this large, propensity-matched, real-world analysis, individuals with overweight or obesity treated with tirzepatide were significantly more likely to achieve clinically meaningful weight loss and larger reductions in body weight compared to those treated with semaglutide. Consistent treatment effects were observed in subgroups with and without T2D. Future work is needed to compare the effect of tirzepatide and semaglutide on other key end points (e.g., reduction in adverse cardiovascular events) [6, 37].

## Supporting information

Supplement A: Methods and Results

Supplement B: Data Definitions

## Data Availability

The data used in this study are available to all Truveta subscribers and may be accessed at studio.truveta.com.

## Acknowledgements

The authors thank Elisabetta Patorno for her substantial contributions to the study design.

## Results

## References

[1] Cynthia L. Ogden, Tala H. Fakhouri, Margaret D. Carroll, Craig M. Hales, Cheryl D. Fryar, Xianfen Li, and David S. Freedman. Prevalence of obesity among adults, by household income and education — united states, 2011–2014. MMWR. Morbidity and Mortality Weekly Report, 66(50):1369–1373, dec 2017. doi: 10.15585/mmwr.mm6650a1. URL https://doi.org/10.15585%2Fmmwr.mm6650a1.

[2] Zahra Raisi-Estabragh, Ofer Kobo, Jennifer H. Mieres, Renee P. Bullock-Palmer, Harriette G.C. Van Spall, Khadijah Breathett, and Mamas A. Mamas. Racial disparities in obesity-related cardiovascular mortality in the united states: Temporal trends from 1999 to 2020. Journal of the American Heart Association, 12(18), sep 2023. doi: 10.1161/jaha.122.028409. URL https://doi.org/10.1161%2Fjaha.122.028409.

[3] Rena R. Wing, Wei Lang, Thomas A. Wadden, Monika Safford, William C. Knowler, Alain G. Bertoni, James O. Hill, Frederick L. Brancati, Anne Peters, and Lynne Wagenknecht and. Benefits of modest weight loss in improving cardiovascular risk factors in overweight and obese individuals with type 2 diabetes. Diabetes Care, 34(7):1481–1486, jun 2011. doi: 10.2337/dc10-2415. URL https://doi.org/10.2337%2Fdc10-2415.

[4] Ken Fujioka. Safety and tolerability of medications approved for chronic weight management. Obesity, 23(S1), apr 2015. doi: 10.1002/oby.21094. URL https://doi.org/10.1002%2Foby.21094.

[5] Nithushi R. Samaranayake, Kwok L. Ong, Raymond Y.H. Leung, and Bernard M.Y. Cheung. Management of obesity in the national health and nutrition examination survey (NHANES), 2007–2008. Annals of Epidemiology, 22(5):349–353, may 2012. doi: 10.1016/j.annepidem.2012.01.001. URL https://doi.org/10.1016%2Fj.annepidem.2012.01.001.

[6] Muhammad Shariq Usman, Melanie Davies, Michael E Hall, Subodh Verma, Stefan D Anker, Julio Rosenstock, and Javed Butler. The cardiovascular effects of novel weight loss therapies. European Heart Journal, November 2023. ISSN 1522-9645. doi: 10.1093/eurheartj/ehad664. URL http://dx.doi.org/10.1093/eurheartj/ehad664.

[7] W. Timothy Garvey, Rachel L. Batterham, Meena Bhatta, Silvio Buscemi, Louise N. Christensen, Juan P. Frias, Esteban Jódar, Kristian Kandler, Georgia Rigas, Thomas A. Wadden, and Sean Wharton and. Two-year effects of semaglutide in adults with overweight or obesity: the STEP 5 trial. Nature Medicine, 28(10):2083–2091, oct 2022. doi: 10.1038/s41591-022-02026-4. URL https://doi.org/10.1038%2Fs41591-022-02026-4.

[8] Melanie Davies, Louise Færch, Ole K Jeppesen, Arash Pakseresht, Sue D Pedersen, Leigh Perreault, Julio Rosenstock, Iichiro Shimomura, Adie Viljoen, Thomas A Wadden, and Ildiko Lingvay. Semaglutide 2·4 mg once a week in adults with overweight or obesity, and type 2 diabetes (STEP 2): a randomised, double-blind, double-dummy, placebo-controlled, phase 3 trial. The Lancet, 397(10278):971–984, mar 2021. doi: 10.1016/s0140-6736(21)00213-0. URL https://doi.org/10.1016%2Fs0140-6736%2821%2900213-0.

[9] Ania M Jastreboff, Louis J Aronne, Nadia N Ahmad, Sean Wharton, Lisa Connery, Breno Alves, Arihiro Kiyosue, Zhang, Mathijs C Bunck, et al. Tirzepatide once weekly for the treatment of obesity. New England Journal of Medicine, 387(3):205–216, 2022.

[10] W Timothy Garvey, Juan P Frias, Ania M Jastreboff, Carel W le Roux, Naveed Sattar, Diego Aizenberg, Huzhang Mao, Shuyu Zhang, Nadia N Ahmad, Mathijs C Bunck, et al. Tirzepatide once weekly for the treatment of obesity in people with type 2 diabetes (surmount-2): a double-blind, randomised, multicentre, placebo-controlled, phase 3 trial. The Lancet, 2023.

[11] Juan P Frias, Melanie J Davies, Julio Rosenstock, Federico C Pérez Manghi, Laura Fernández Landó, Brandon K Bergman, Bing Liu, Xuewei Cui, and Katelyn Brown. Tirzepatide versus semaglutide once weekly in patients with type 2 diabetes. New England Journal of Medicine, 385(6):503–515, 2021.

[12] Crystal N. Johnson-Mann, Julie S. Cupka, Alexandra Ro, Andrea E. Davidson, Brooke A. Armfield, Frank Miralles, Asena Markal, Kiara E. Fierman, Victoria Hough, Mackenzie Newsom, Isha Verma, Abdul-Vehab Dozic, and Azra Bihorac. A systematic review on participant diversity in clinical trials—have we made progress for the management of obesity and its metabolic sequelae in diet, drug, and surgical trials. Journal of Racial and Ethnic Health Disparities, dec 2022. doi: 10.1007/s40615-022-01487-0. URL https://doi.org/10.1007%2Fs40615-022-01487-0.

[13] Luther T. Clark, Laurence Watkins, Ileana L. Piña, Mary Elmer, Ola Akinboboye, Millicent Gorham, Brenda Jamerson, Cassandra McCullough, Christine Pierre, Adam B. Polis, Gary Puckrein, and Jeanne M. Regnante. Increasing diversity in clinical trials: Overcoming critical barriers. Current Problems in Cardiology, 44(5):148–172, may 2019. doi: 10.1016/j.cpcardiol.2018.11.002. URL https://doi.org/10.1016%2Fj.cpcardiol.2018.11.002.

[14] Amelia J. Averitt, Chunhua Weng, Patrick Ryan, and Adler Perotte. Translating evidence into practice: eligibility criteria fail to eliminate clinically significant differences between real-world and study populations. npj Digital Medicine, 3(1), may 2020. doi: 10.1038/s41746-020-0277-8. URL https://doi.org/10.1038%2Fs41746-020-0277-8.

[15] H Beba, C Ranns, C Hambling, J Diggle, and P Brown. Pcds consensus statement: A strategy for managing the supply shortage of the glp-1 ras ozempic and trulicity. Diabetes & Primary Care, 24(5), 2022.

[16] Donald B. Rubin. Multiple Imputation for Nonresponse in Surveys. Wiley, jun 1987. doi: 10.1002/9780470316696. URL https://doi.org/10.1002%2F9780470316696.

[17] Lionel Henry and Hadley Wickham. rlang: Functions for Base Types and Core R and Tidyverse Features, 2023. URL https://CRAN.R-project.org/package=rlang. R package version 1.1.0.

[18] Neal Richardson, Ian Cook, Nic Crane, Jonathan Keane, Romain François, Jeroen Ooms, and Apache Arrow. arrow: Integration to Apache ‘Arrow’, 2022. https://github.com/apache/arrow/.

[19] Hadley Wickham, Romain François, Lionel Henry, Kirill Müller, and Davis Vaughan. dplyr: A Grammar of Data Manipulation, 2023. URL https://CRAN.R-project.org/package=dplyr. R package version 1.1.0.

[20] Vitalie Spinu, Garrett Grolemund, and Hadley Wickham. lubridate: Make Dealing with Dates a Little Easier, 2023. URL https://CRAN.R-project.org/package=lubridate. R package version 1.9.1.

[21] Hadley Wickham. forcats: Tools for Working with Categorical Variables (Factors), 2021. URL https://CRAN.R-project.org/package=forcats. R package version 0.5.1.

[22] Benjamin Rich. table1: Tables of Descriptive Statistics in HTML, 2021. URL https://github.com/benjaminrich/table1. R package version 1.4.2.

[23] Noah Greifer. cobalt: Covariate Balance Tables and Plots, 2023. URL https://CRAN.R-pro ject.org/package=cobalt. R package version 4.5.1.

[24] Daniel Ho, Kosuke Imai, Gary King, Elizabeth Stuart, and Noah Greifer. MatchIt: Nonparametric Preprocessing for Parametric Causal Inference, 2022. URL https://CRAN.R-project.org/package=MatchIt. R package version 4.5.0.

[25] Noah Greifer. WeightIt: Weighting for Covariate Balance in Observational Studies, 2023. URL https://CRAN.R-project.org/package=WeightIt. R package version 0.14.2.

[26] Farhad Pishgar and Noah Greifer. MatchThem: Matching and Weighting Multiply Imputed Datasets, 2023. URL https://github.com/FarhadPishgar/MatchThem. R package version 1.1.0.

[27] Thomas Lumley. survey: Analysis of Complex Survey Samples, 2023. URL http://r-surve y.r-forge.r-project.org/survey/. R package version 4.2-1.

[28] Terry M Therneau. survival: Survival Analysis, 2023. URL https://github.com/therneau/survival. R package version 3.5-5.

[29] Daniel D. Sjoberg, Mark Baillie, Charlotta Fruechtenicht, Steven Haesendonckx, and Tim Treis. ggsurvfit: Flexible Time-to-Event Figures, 2023. URL https://CRAN.R-project.org/package=ggsurvfit. R package version 1.0.0.

[30] David Robinson, Alex Hayes, and Simon Couch. broom: Convert Statistical Objects into Tidy Tibbles, 2022. URL https://CRAN.R-project.org/package=broom. R package version 1.0.1.

[31] Hadley Wickham, Winston Chang, Lionel Henry, Thomas Lin Pedersen, Kohske Takahashi, Claus Wilke, Kara Woo, Hiroaki Yutani, and Dewey Dunnington. ggplot2: Create Elegant Data Visualisations Using the Grammar of Graphics, 2022. URL https://CRAN.R-project.org/package=ggplot2. R package version 3.4.0.

[32] David B. Dahl, David Scott, Charles Roosen, Arni Magnusson, and Jonathan Swinton. xtable: Export Tables to LaTeX or HTML, 2019. URL http://xtable.r-forge.r-project.org/. R package version 1.8-4.

[33] John PH Wilding, Rachel L Batterham, Salvatore Calanna, Melanie Davies, Luc F Van Gaal, Ildiko Lingvay, Barbara M McGowan, Julio Rosenstock, Marie TD Tran, Thomas A Wadden, et al. Once-weekly semaglutide in adults with overweight or obesity. New England Journal of Medicine, 384(11):989–1002, 2021.

[34] Tracey Weiss, Lingfeng Yang, Richard D Carr, Sampriti Pal, Baanie Sawhney, Robert Boggs, Swapnil Rajpathak, and Kristy Iglay. Real-world weight change, adherence, and discontinuation among patients with type 2 diabetes initiating glucagon-like peptide-1 receptor agonists in the UK. BMJ Open Diabetes Research &amp Care, 10(1):e002517, jan 2022. doi: 10.1136/bmjdrc-2021-002517. URL https://doi.org/10.1136%2Fbmjdrc-2021-002517.

[35] Emily Durden, Michael Liang, Robert Fowler, Ulrik Haagen Panton, and Emina Mocevic. The effect of early response to GLP-1 RA therapy on long-term adherence and persistence among type 2 diabetes patients in the united states. Journal of Managed Care &amp Specialty Pharmacy, 25(6):669–680, jun 2019. doi: 10.18553/jmcp.2019.18429. URL https://doi.org/10.18553%2Fjmcp.2019.18429.

[36] Sean Wharton, Arne Astrup, Lars Endahl, Michael E. J. Lean, Altynai Satylganova, Dorthe Skovgaard, Thomas A. Wadden, and John P. H. Wilding. Estimating and reporting treatment effects in clinical trials for weight management: using estimands to interpret effects of intercurrent events and missing data. International Journal of Obesity, 45(5):923–933, jan 2021. doi: 10.1038/s41366-020-00733-x. URL https://doi.org/10.1038%2Fs41366-020-00733-x.

[37] A. Michael Lincoff, Kirstine Brown-Frandsen, Helen M. Colhoun, John Deanfield, Scott S. Emerson, Sille Esbjerg, Søren Hardt-Lindberg, G. Kees Hovingh, Steven E. Kahn, Robert F. Kushner, Ildiko Lingvay, Tugce K. Oral, Marie M. Michelsen, Jorge Plutzky, Christoffer W. Tornøe, and Donna H. Ryan. Semaglutide and cardiovascular outcomes in obesity without diabetes. New England Journal of Medicine, 0(0):null, 0. doi: 10.1056/NEJMoa2307563. URL https://doi.org/10.1056/NEJMoa2307563.

