## Supplement A: Methods and Results for "Comparative Effectiveness of Semaglutide and Tirzepatide for Weight Loss in Adults with Overweight and Obesity in the US: A Real-World Evidence Study"

### Supplemental Material

#### Contents


**1 Quantitative Measure Pre-processing** **2**
1.1 Weight 2

**2 Censoring** **3**

**3 Sensitivity Analyses** **4**
3.1 Time to Weight Loss 4
3.2 Change in Body weight 5

**4 Subgroup Analyses** **6**
4.1 Time to Weight Loss 6
4.1.1 Survival Curves 6
4.1.2 Proportion Achieving Weight Loss within 1 Year 8
4.1.3 Treatment Effects 8
4.2 Change in Body weight 10
4.2.1 Treatment Effects, Change in Body Weight 10

### 1 Quantitative Measure Pre-processing

#### 1.1 Weight

Several steps were taken to pre-process weight data. First, weights observations with null/missing values were removed.

In instances where a value was recorded but the unit was missing, we assumed values between 40,824 - 317,520 were grams, values between 1,440 - 11,200 were ounces, values greater than 317 were pounds, and values less than 125 were kilograms. These ranges correspond to values of 90 -700 lbs. For this study of adults with overweight or obesity, values equivalent to <90 or >700 pounds were assumed to be entered in error.

For remaining observations where a unit could not be assumed based on value, we assumed the unit based on consistency with other patient values. These were predominately values >125 and <300, which could plausibly correspond to either pounds or kilograms in our population. For each patient, we calculated the mean and standard deviation of weights with known or assumed units in the 15 months before to 15 months after the index date. For each observation with a value but no unit, we calculated the distance from the mean assuming the observation was measured in (a) pounds and (b) kilograms. We then assigned the unit as whichever was closest to the mean, so long as it fell within a plausible distance. We assumed a plausible distance of 30%. We then removed any additional values where a unit could not be assigned.

Finally, after reviewing high-variance patient trajectories to identify common patterns, we removed outliers in the patient trajectory that likely represented data entry errors. The primary pattern was a weight bounce, where one weight value was highly inconsistent with both the previous and subsequent weight values. These largely appeared to be incorrect entry of pounds as kilograms and vice versa, resulting in approximate halving or doubling of a single observation (e.g., day -60: 402 pounds, day 0: 400 pounds, day 45: 180 pounds, day 70: 390 pounds). If the preceding weight value was within 365 days and the absolute change was >40% *and* the subsequent weight value was within 45 days and the absolute change was >40%, we removed the value. We also removed instances where one value is >40% different from the next two weight values and all are within 365 days. This was intended to capture cases of bounce, where the first observation was the aberrant value. Next, we removed biologically implausible changes in a short period, those where the preceding weight was within 5 days and absolute change was > 10% *and* the next weight value is within 5 days and absolute change was > 10%. We explored the use of interquartile based approaches for outlier detection but found these to have poor specificity.

### 2 Censoring

Enrollment and follow-up times are given in Figures 1 and 2. Follow-up times were somewhat shorter for patients initiating medications in Fall 2022, when shortages were reported. Follow-up times increased somewhat for patients initiating in December 2022, around the time some shortages of semaglutide were reduced.

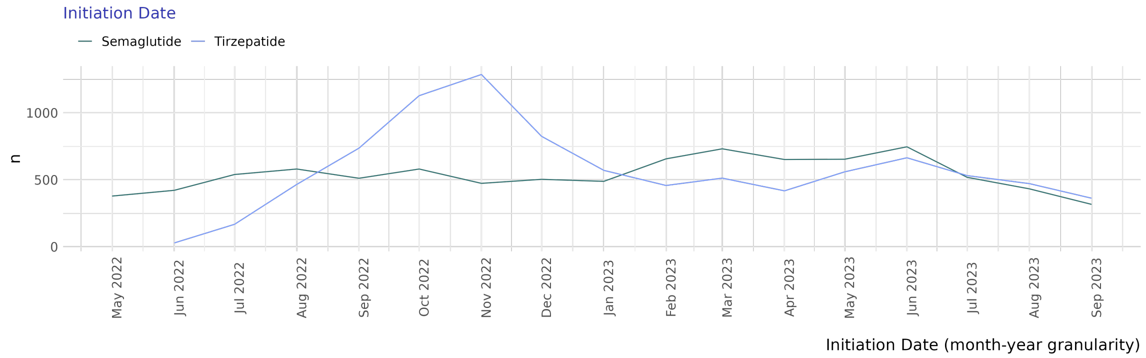


Figure 1: Distribution of Initiation Time by group

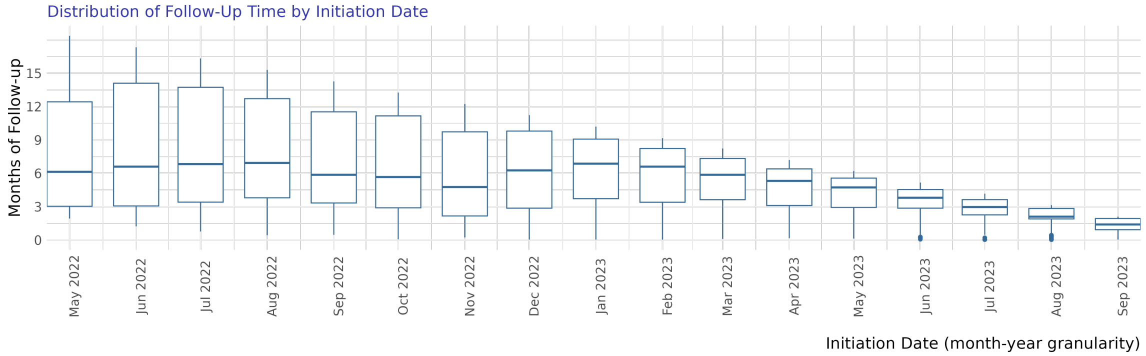


Figure 2: Distribution of follow-up time by initiation date.

### 3 Sensitivity Analyses

Sensitivity analyses included use of inverse probability of treatment weighting (IPTW), complete case analysis (e.g., no imputation), and modified intention-to-treat (ITT) analysis.

#### 3.1 Time to Weight Loss

In modified intention-to-treat analyses where patients were administratively censored only (discontinuation and switching were ignored), fewer patients achieved weight reductions 5%, 10%, and 15% (Figure 3). Treatment effects were somewhat attenuated (Figure 4).

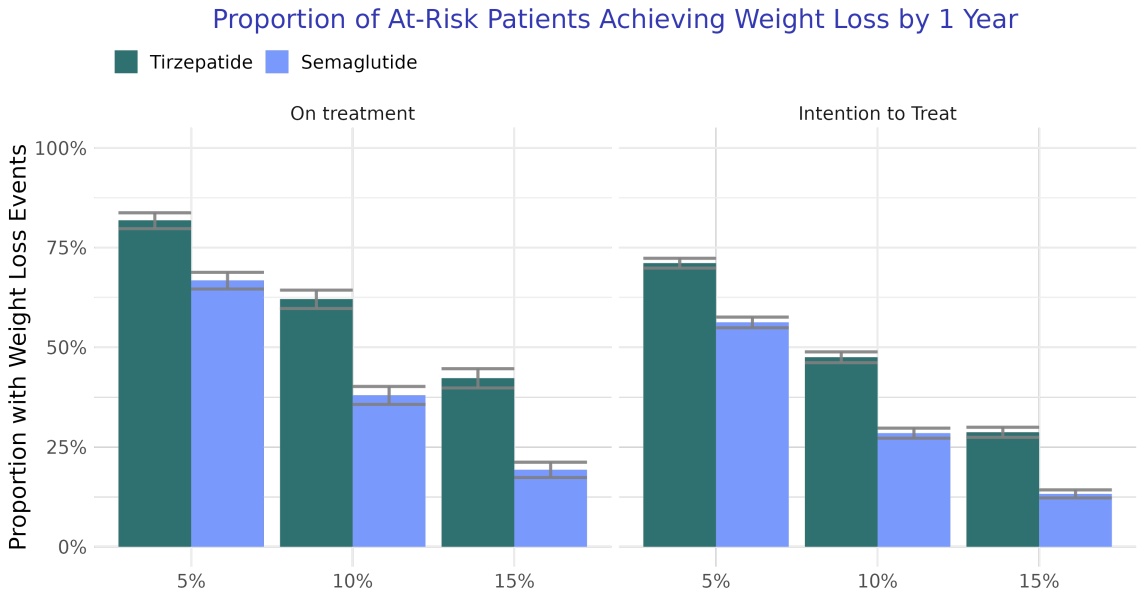


Figure 3: Proportion of at-risk patients achieving weight loss targets by one year for on treatment and intention to treat analyses

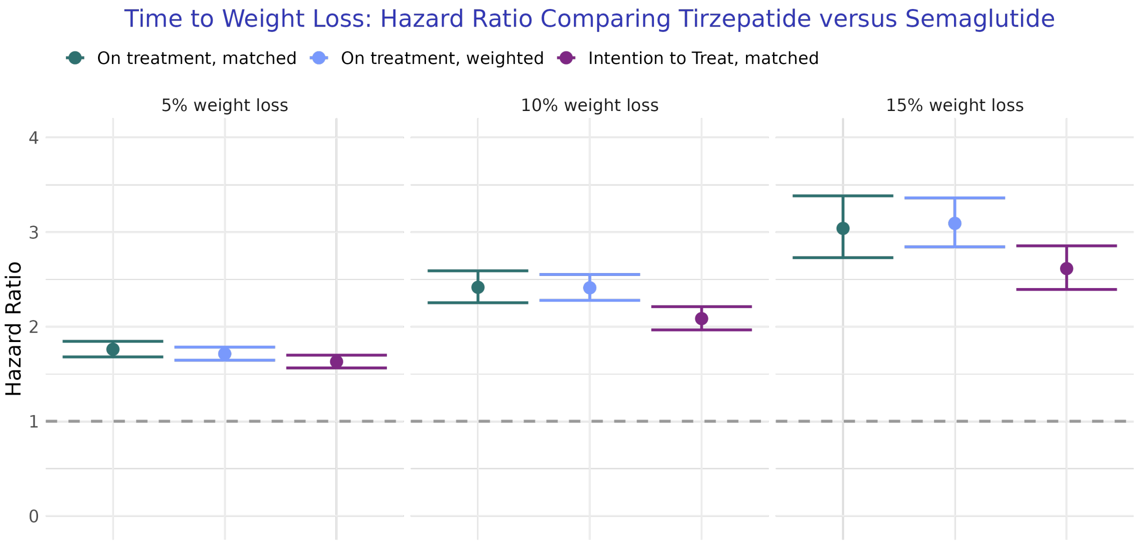


Figure 4: Hazard ratio comparing Tirzepatide vs Semaglutide for achieving weight loss targets under different analytic approaches

#### 3.2 Change in Body weight

Modified intention-to-treat analyses of percentage changes in body weight included patients not yet administratively censored at the timepoint. Both pre- and post- discontinuation weights were included. Reductions in weight were smaller at all timepoints under this analysis (Figure 5). Treatment effects were somewhat attenuated (Figure 6).

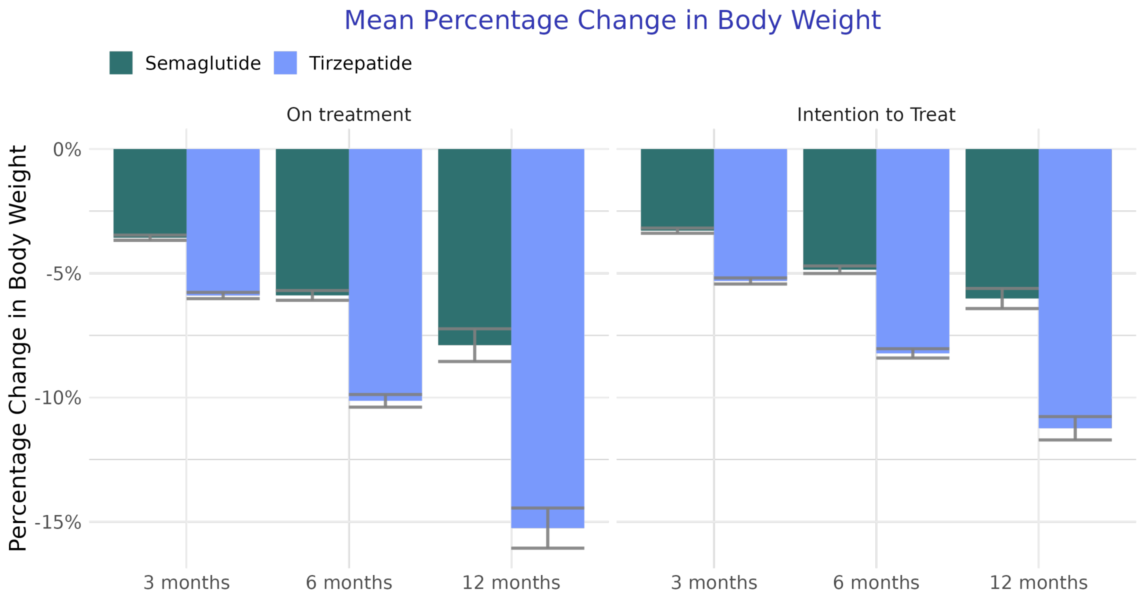


Figure 5: Mean change in body weight for tirzepatide and semaglutide groups under on treatment and modified intention to treat analyses.

### 4 Subgroup Analyses

#### 4.1 Time to Weight Loss

##### 4.1.1 Survival Curves

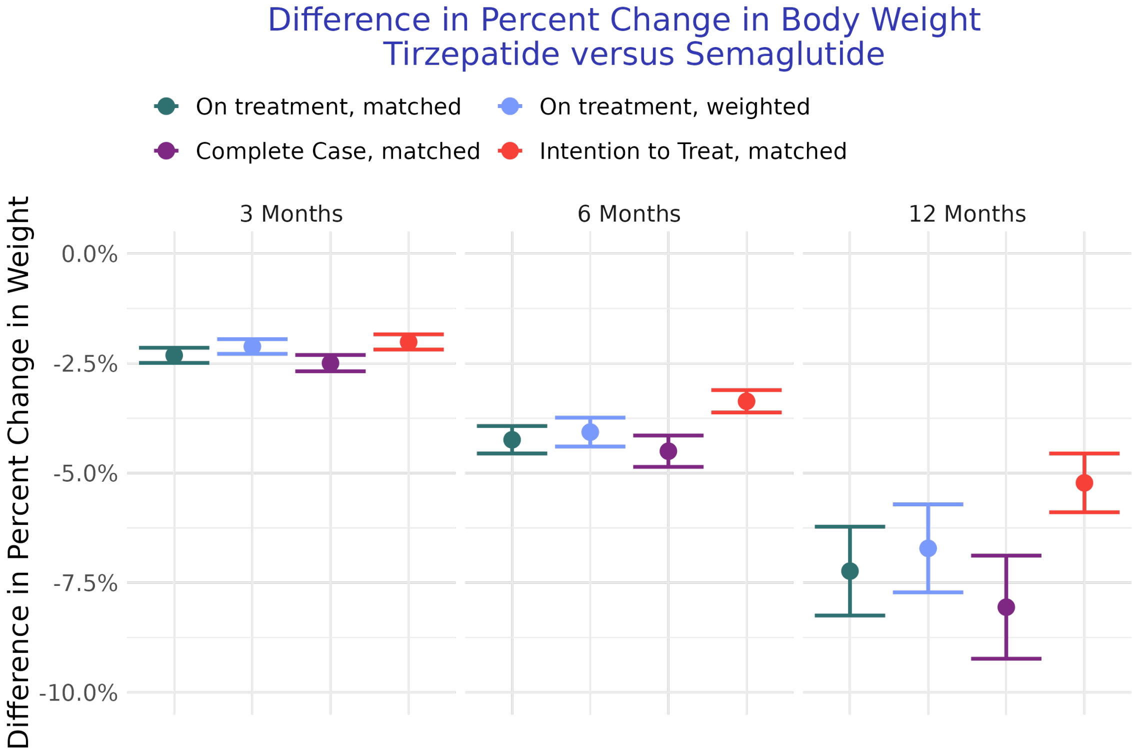


Figure 6: Difference in percent change in body weight comparing tirzepatide to semaglutide under different analytic approached

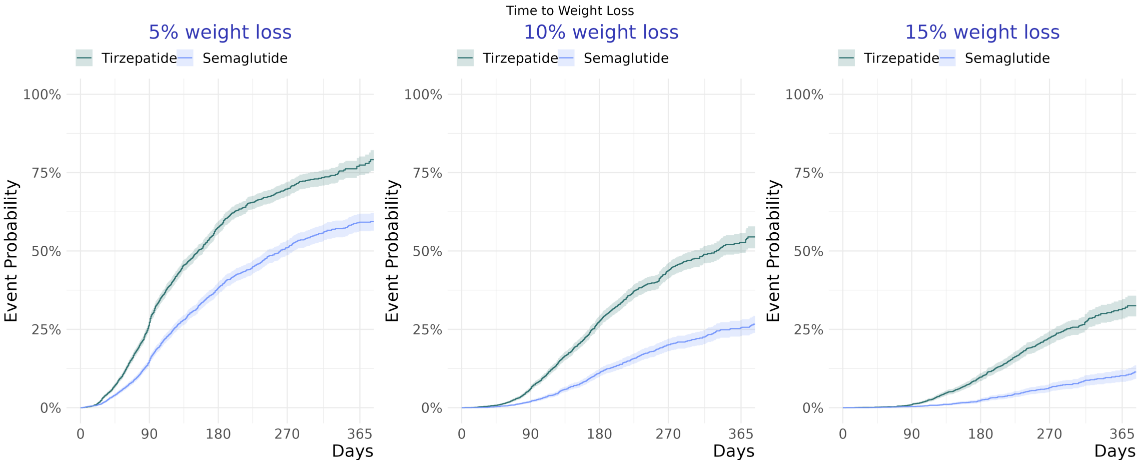


Figure 7: Time to weight loss, for patients with T2D.

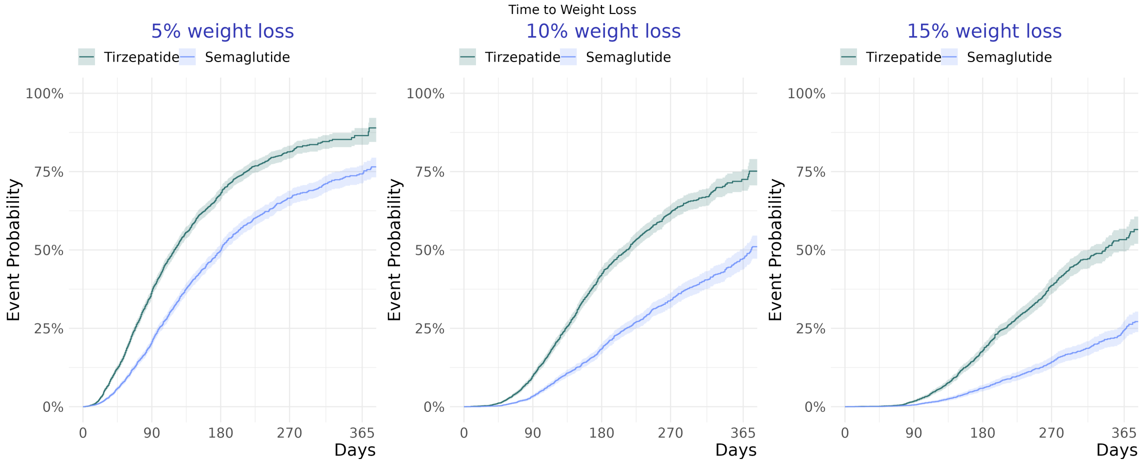


Figure 8: Time to weight loss, for patients without T2D

##### 4.1.2 Proportion Achieving Weight Loss within 1 Year

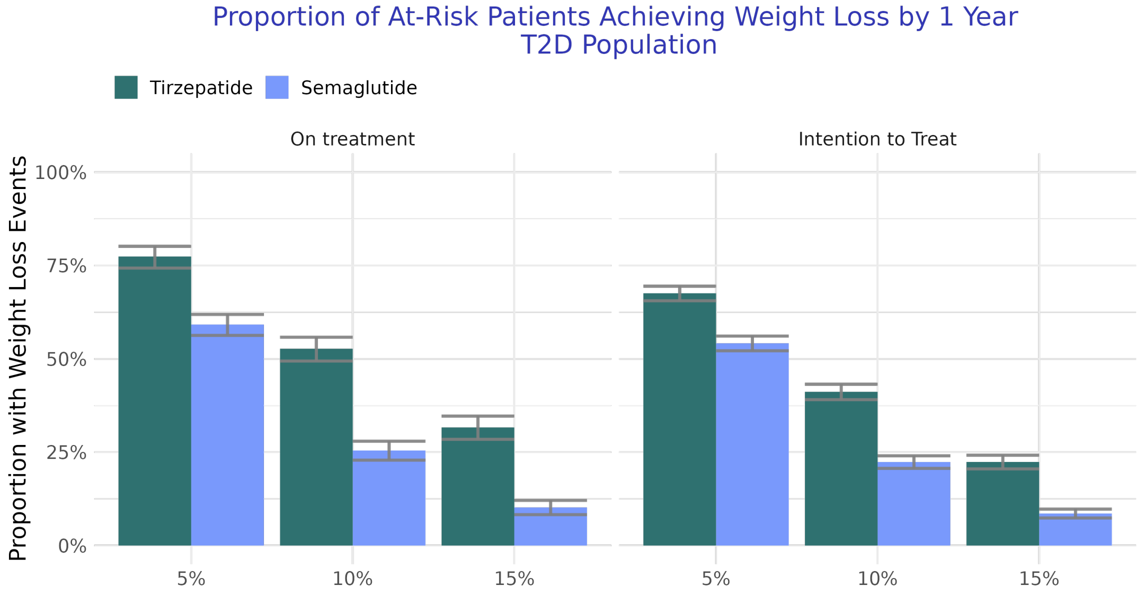


Figure 9: Proportion achieving weight loss, for patients with T2D using on treatment and intention to treat censoring approaches.

##### 4.1.3 Treatment Effects

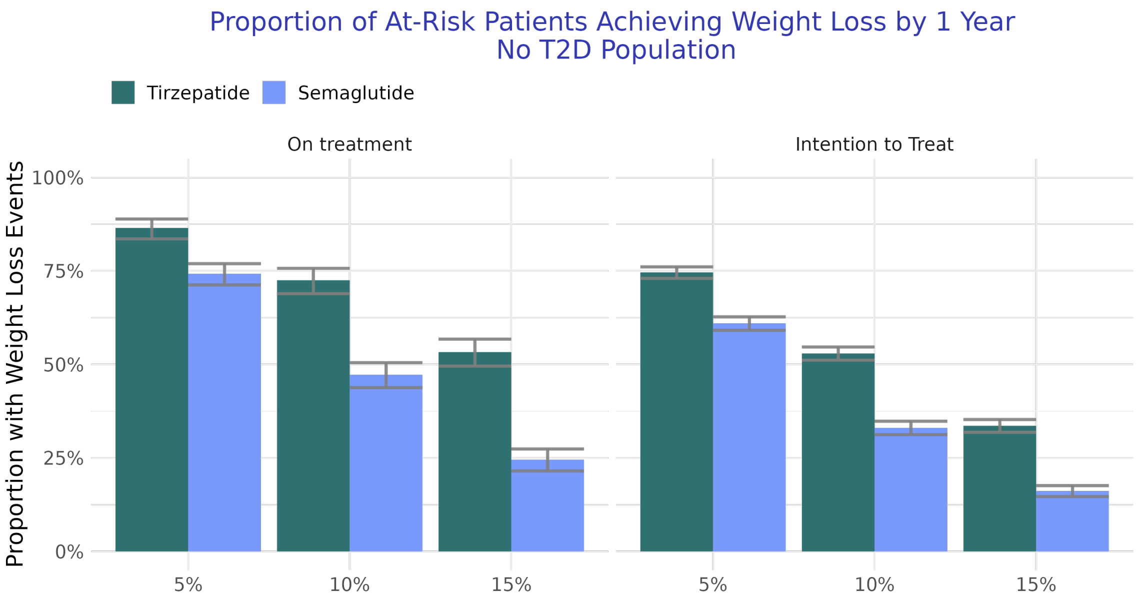


Figure 10: Proportion achieving weight loss, for patients without T2D using on treatment and intention to treat censoring approaches.

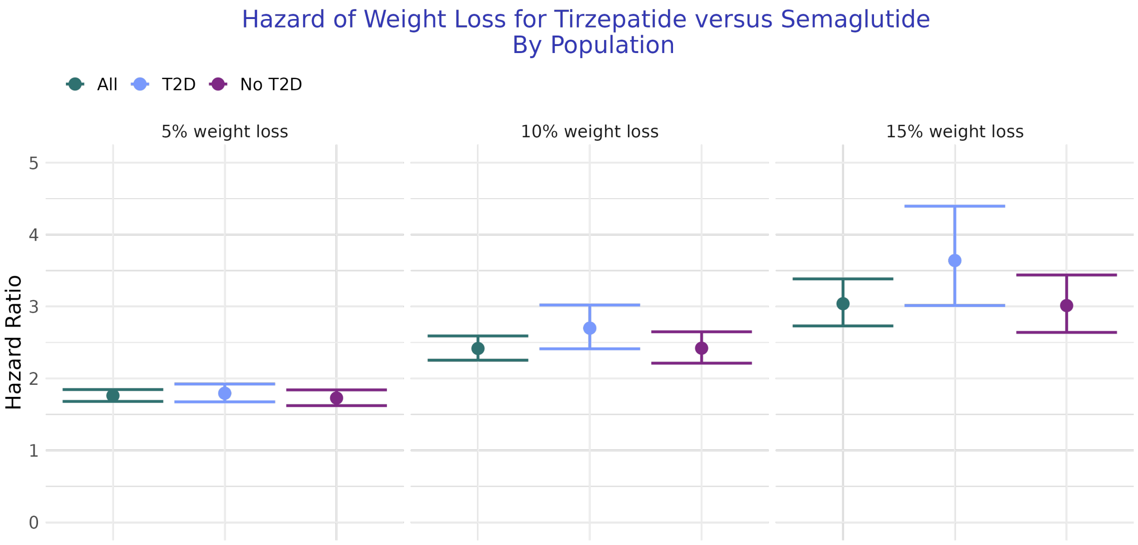


Figure 11: Harzard ratio comparing tirzepatide to semaglutide for different populations

#### 4.2 Change in Body weight

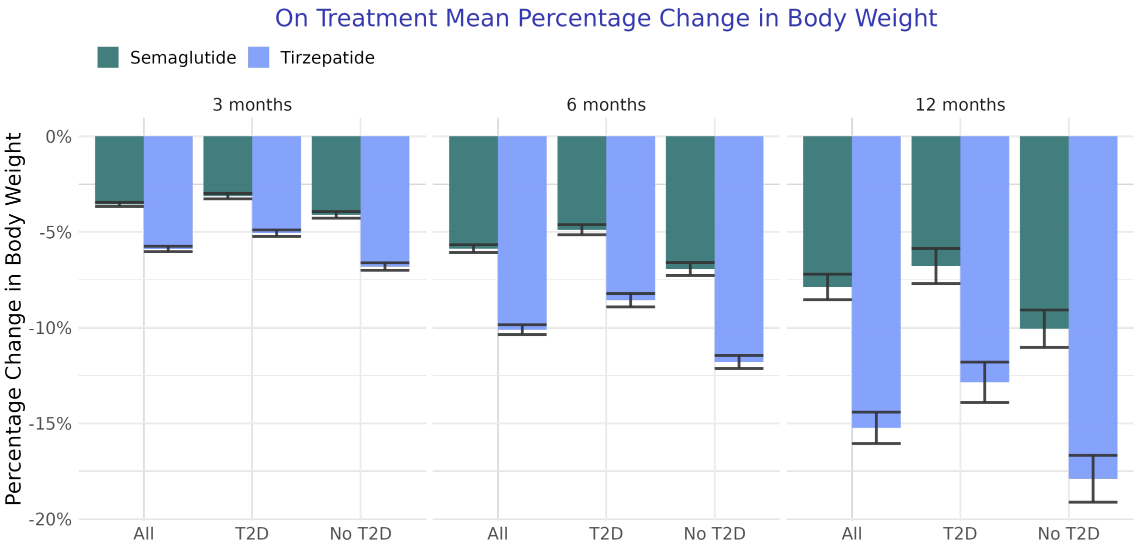


Figure 12: Difference in percent change in bodyweight comparing Tirzepatide to Semaglutide under different analytic approached

##### 4.2.1 Treatment Effects, Change in Body Weight

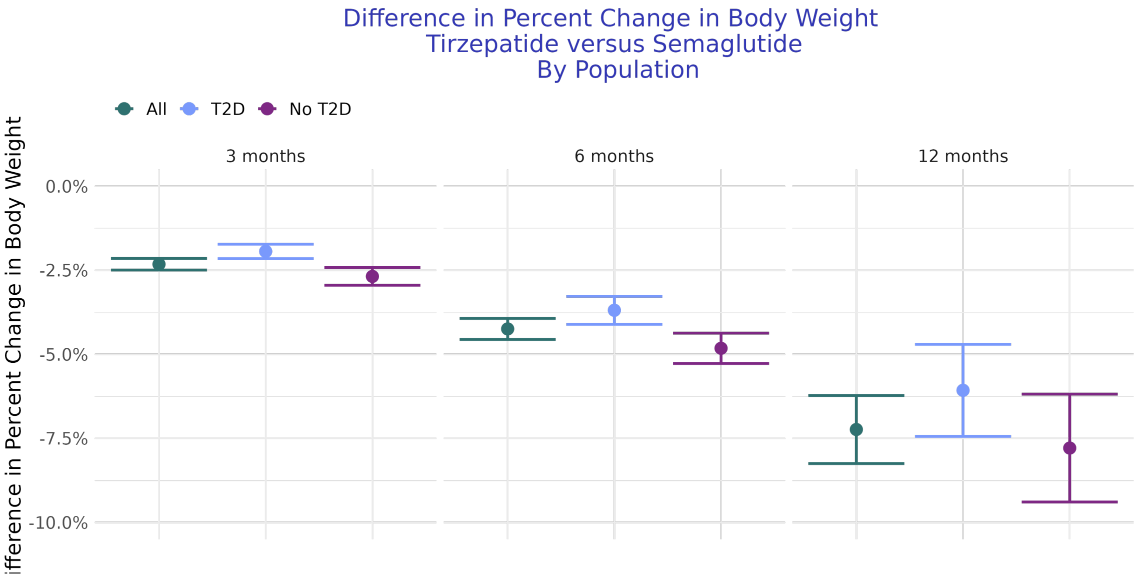


Figure 13: Difference in percent change in bodyweight comparing Tirzepatide to Semaglutide under different analytic approached
