## Supplement B: Data Definitions for "Comparative Effectiveness of Semaglutide and Tirzepatide for Weight Loss in Adults with Overweight and Obesity in the US: A Real-World Evidence Study"

### Supplemental Material: Data Definitions

#### 0.1 GLP1 RA Medications

The following codes were used to identify previous use of any GLP-1 RA or GLP-1 RA/GIP agonist.

Table 1: Concept codes used to identify GLP-1 RA medications.

| CodeSystem | ConceptCode | ConceptName |
|---|---|---|
| RxNorm | 1991302 | semaglutide |
| RxNorm | 2553504 | semaglutide Auto-Injector [Wegovy] |
| RxNorm | 2553506 | 0.5 ML semaglutide 0.5 MG/ML Auto-Injector [Wegovy] |
| RxNorm | 2553604 | semaglutide 1 MG/ML Auto-Injector |
| RxNorm | 2553606 | semaglutide 2 MG/ML |
| RxNorm | 2553700 | semaglutide 2 MG/ML Auto-Injector [Wegovy] |
| RxNorm | 2553900 | semaglutide 2.27 MG/ML |
| RxNorm | 2553902 | semaglutide 2.27 MG/ML [Wegovy] |
| RxNorm | 2553904 | semaglutide 2.27 MG/ML Auto-Injector |
| RxNorm | 2554103 | semaglutide 3.2 MG/ML [Wegovy] |
| RxNorm | 2553601 | 0.5 ML semaglutide 1 MG/ML Auto-Injector |
| RxNorm | 2553608 | semaglutide 2 MG/ML [Wegovy] |
| RxNorm | 2554101 | semaglutide 3.2 MG/ML |
| RxNorm | 2553501 | 0.5 ML semaglutide 0.5 MG/ML Auto-Injector |
| RxNorm | 2553905 | semaglutide 2.27 MG/ML Auto-Injector [Wegovy] |
| RxNorm | 2554102 | 0.75 ML semaglutide 3.2 MG/ML Auto-Injector |
| RxNorm | 2553400 | semaglutide 0.5 MG/ML |
| RxNorm | 2553505 | Wegovy Injectable Product |
| RxNorm | 2553507 | semaglutide 0.5 MG/ML Auto-Injector |
| RxNorm | 2553605 | semaglutide 1 MG/ML Auto-Injector [Wegovy] |
| RxNorm | 2553607 | semaglutide 2 MG/ML Auto-Injector |
| RxNorm | 2553901 | 0.75 ML semaglutide 2.27 MG/ML Auto-Injector |
| RxNorm | 2553903 | 0.75 ML semaglutide 2.27 MG/ML Auto-Injector [Wegovy] |
| RxNorm | 2554104 | 0.75 ML semaglutide 3.2 MG/ML Auto-Injector [Wegovy] |
| RxNorm | 2553502 | Wegovy |
| RxNorm | 2553803 | 0.5 ML semaglutide 2 MG/ML Auto-Injector [Wegovy] |
| RxNorm | 2554105 | semaglutide 3.2 MG/ML Auto-Injector |
| RxNorm | 2553503 | semaglutide 0.5 MG/ML [Wegovy] |
| RxNorm | 2553802 | 0.5 ML semaglutide 2 MG/ML Auto-Injector |
| RxNorm | 2554106 | semaglutide 3.2 MG/ML Auto-Injector [Wegovy] |
| RxNorm | 2553508 | semaglutide 0.5 MG/ML Auto-Injector [Wegovy] |
| RxNorm | 2553600 | semaglutide 1 MG/ML |
| RxNorm | 2553602 | semaglutide 1 MG/ML [Wegovy] |
| RxNorm | 2553500 | semaglutide Auto-Injector |
| RxNorm | 2553603 | 0.5 ML semaglutide 1 MG/ML Auto-Injector [Wegovy] |
| RxNorm | 2599362 | 3 ML semaglutide 2.68 MG/ML Pen Injector |
| RxNorm | 2599365 | 3 ML semaglutide 2.68 MG/ML Pen Injector [Ozempic] |
| RxNorm | 2599361 | semaglutide 2.68 MG/ML |
| RxNorm | 2599364 | semaglutide 2.68 MG/ML [Ozempic] |

Continued on next page

**Table1 – continued from previous page**

| CodeSystem | ConceptCode | ConceptName |
|---|---|---|
| RxNorm | 2599363 | semaglutide 2.68 MG/ML Pen Injector |
| RxNorm | 2599366 | semaglutide 2.68 MG/ML Pen Injector [Ozempic] |
| RxNorm | 1991303 | semaglutide 1.34 MG/ML |
| RxNorm | 1991304 | semaglutide Injectable Product |
| RxNorm | 1991305 | semaglutide Pen Injector |
| RxNorm | 1991306 | 0.25 MG, 0.5 MG Dose 1.5 ML semaglutide 1.34 MG/ML Pen Injector |
| RxNorm | 1991307 | Ozempic |
| RxNorm | 1991308 | semaglutide 1.34 MG/ML [Ozempic] |
| RxNorm | 1991309 | semaglutide Pen Injector [Ozempic] |
| RxNorm | 1991310 | Ozempic Injectable Product |
| RxNorm | 1991311 | 0.25 MG, 0.5 MG Dose 1.5 ML semaglutide 1.34 MG/ML Pen Injector [Ozempic] |
| RxNorm | 1991316 | 1 MG Dose 1.5 ML semaglutide 1.34 MG/ML Pen Injector |
| RxNorm | 1991317 | 1 MG Dose 1.5 ML semaglutide 1.34 MG/ML Pen Injector [Ozempic] |
| RxNorm | 2619152 | 0.25 MG, 0.5 MG Dose 3 ML semaglutide 0.68 MG/ML Pen Injector |
| RxNorm | 2619154 | 0.25 MG, 0.5 MG Dose 3 ML semaglutide 0.68 MG/ML Pen Injector [Ozempic] |
| RxNorm | 2200654 | semaglutide 3 MG Oral Tablet [Rybelsus] |
| RxNorm | 2200655 | semaglutide 7 MG |
| RxNorm | 2200656 | semaglutide 7 MG Oral Tablet |
| RxNorm | 2200657 | semaglutide 7 MG [Rybelsus] |
| RxNorm | 2200658 | semaglutide 7 MG Oral Tablet [Rybelsus] |
| RxNorm | 2200640 | semaglutide 14 MG |
| RxNorm | 2200641 | semaglutide Oral Product |
| RxNorm | 2200642 | semaglutide Pill |
| RxNorm | 2200643 | semaglutide Oral Tablet |
| RxNorm | 2200644 | semaglutide 14 MG Oral Tablet |
| RxNorm | 2200645 | Rybelsus |
| RxNorm | 2200646 | semaglutide 14 MG [Rybelsus] |
| RxNorm | 2200647 | semaglutide Oral Tablet [Rybelsus] |
| RxNorm | 2200648 | Rybelsus Oral Product |
| RxNorm | 2200649 | Rybelsus Pill |
| RxNorm | 2200650 | semaglutide 14 MG Oral Tablet [Rybelsus] |
| RxNorm | 2200651 | semaglutide 3 MG |
| RxNorm | 2200652 | semaglutide 3 MG Oral Tablet |
| RxNorm | 2200653 | semaglutide 3 MG [Rybelsus] |
| RxNorm | 2398841 | 3 ML semaglutide 1.34 MG/ML Pen Injector |
| RxNorm | 2398842 | 3 ML semaglutide 1.34 MG/ML Pen Injector [Ozempic] |
| RxNorm | 2398843 | semaglutide 1.34 MG/ML Pen Injector |
| RxNorm | 2398844 | semaglutide 1.34 MG/ML Pen Injector [Ozempic] |

Continued on next page

**Table1 – continued from previous page**

| CodeSystem | ConceptCode | ConceptName |
|---|---|---|
| RxNorm | 2619151 | semaglutide 0.68 MG/ML |
| RxNorm | 2619153 | semaglutide 0.68 MG/ML [Ozempic] |
| SNOMED CT | 764283003 | Semaglutide |
| SNOMED CT | 764284009 | Semaglutide-containing product |
| SNOMED CT | 770768004 | Semaglutide-containing product in parenteral dose form |
| SNOMED CT | 764285005 | Semaglutide-containing product in oral dose form |
| SNOMED CT | 777514008 | Semaglutide only product |
| SNOMED CT | 780439007 | Semaglutide only product in oral dose form |
| SNOMED CT | 780440009 | Semaglutide only product in parenteral dose form |
| SNOMED CT | 782102009 | Semaglutide 1.34 mg/mL solution for injection |
| SNOMED CT | 1003647000 | Semaglutide 3 mg oral tablet |
| SNOMED CT | 1003648005 | Semaglutide 7 mg oral tablet |
| SNOMED CT | 1003649002 | Semaglutide 14 mg oral tablet |
| NDC | 500905949 | semaglutide 1.34mg/mL SUBCUTANEOUS INJECTION, SOLUTION |
| NDC | 00169477290 | 3 ML semaglutide 2.68 MG/ML Pen Injector [Ozempic] |
| NDC | 00169477211 | 3 ML semaglutide 2.68 MG/ML Pen Injector [Ozempic] |
| NDC | 001694772 | semaglutide 2.68mg/mL SUBCUTANEOUS INJECTION, SOLUTION |
| NDC | 50090594900 | 3 ML semaglutide 1.34 MG/ML Pen Injector [Ozempic] |
| NDC | 00169477297 | 3 ML semaglutide 2.68 MG/ML Pen Injector [Ozempic] |
| NDC | 00169477212 | 3 ML semaglutide 2.68 MG/ML Pen Injector [Ozempic] |
| NDC | 705182143 | semaglutide 1.34mg/mL SUBCUTANEOUS INJECTION, SOLUTION |
| NDC | 70518214300 | 0.25 MG, 0.5 MG Dose 1.5 ML semaglutide 1.34 MG/ML Pen Injector [Ozempic] |
| NDC | 001694130 | semaglutide 1.34mg/mL SUBCUTANEOUS INJECTION, SOLUTION |
| NDC | 00169413001 | 3 ML semaglutide 1.34 MG/ML Pen Injector [Ozempic] |
| NDC | 00169413013 | 3 ML semaglutide 1.34 MG/ML Pen Injector [Ozempic] |
| NDC | 001694132 | semaglutide 1.34mg/mL SUBCUTANEOUS INJECTION, SOLUTION |
| NDC | 00169413211 | 0.25 MG, 0.5 MG Dose 1.5 ML semaglutide 1.34 MG/ML Pen Injector [Ozempic] |
| NDC | 00169413212 | 0.25 MG, 0.5 MG Dose 1.5 ML semaglutide 1.34 MG/ML Pen Injector [Ozempic] |
| NDC | 00169413290 | 0.25 MG, 0.5 MG Dose 1.5 ML semaglutide 1.34 MG/ML Pen Injector [Ozempic] |
| NDC | 00169413297 | 0.25 MG, 0.5 MG Dose 1.5 ML semaglutide 1.34 MG/ML Pen Injector [Ozempic] |
| NDC | 001694136 | semaglutide 1.34mg/mL SUBCUTANEOUS INJECTION, SOLUTION |

Continued on next page

**Table1 – continued from previous page**

| CodeSystem | ConceptCode | ConceptName |
|---|---|---|
| NDC | 00169413602 | 1 MG Dose 1.5 ML semaglutide 1.34 MG/ML Pen Injector [Ozempic] |
| NDC | 00169413611 | 1 MG Dose 1.5 ML semaglutide 1.34 MG/ML Pen Injector [Ozempic] |
| NDC | 001694303 | oral semaglutide 3mg/1 ORAL TABLET |
| NDC | 00169430301 | semaglutide 3 MG Oral Tablet [Rybelsus] |
| NDC | 00169430313 | semaglutide 3 MG Oral Tablet [Rybelsus] |
| NDC | 00169430330 | semaglutide 3 MG Oral Tablet [Rybelsus] |
| NDC | 00169430390 | semaglutide 3 MG Oral Tablet [Rybelsus] |
| NDC | 00169430393 | semaglutide 3 MG Oral Tablet [Rybelsus] |
| NDC | 00169430399 | semaglutide 3 MG Oral Tablet [Rybelsus] |
| NDC | 001694307 | oral semaglutide 7mg/1 ORAL TABLET |
| NDC | 00169430701 | semaglutide 7 MG Oral Tablet [Rybelsus] |
| NDC | 00169430713 | semaglutide 7 MG Oral Tablet [Rybelsus] |
| NDC | 00169430730 | semaglutide 7 MG Oral Tablet [Rybelsus] |
| NDC | 001694314 | oral semaglutide 14mg/1 ORAL TABLET |
| NDC | 00169431401 | semaglutide 14 MG Oral Tablet [Rybelsus] |
| NDC | 00169431413 | semaglutide 14 MG Oral Tablet [Rybelsus] |
| NDC | 00169431430 | semaglutide 14 MG Oral Tablet [Rybelsus] |
| NDC | 001694501 | semaglutide 1mg/.5mL SUBCUTANEOUS INJECTION, SOLUTION |
| NDC | 00169450101 | 0.5 ML semaglutide 2 MG/ML Auto-Injector [Wegovy] |
| NDC | 00169450114 | 0.5 ML semaglutide 2 MG/ML Auto-Injector [Wegovy] |
| NDC | 001694505 | semaglutide .5mg/.5mL SUBCUTANEOUS INJECTION, SOLUTION |
| NDC | 00169450501 | 0.5 ML semaglutide 1 MG/ML Auto-Injector [Wegovy] |
| NDC | 00169450514 | 0.5 ML semaglutide 1 MG/ML Auto-Injector [Wegovy] |
| NDC | 001694517 | semaglutide 1.7mg/.75mL SUBCUTANEOUS INJECTION, SOLUTION |
| NDC | 00169451701 | 0.75 ML semaglutide 2.27 MG/ML Auto-Injector [Wegovy] |
| NDC | 00169451714 | 0.75 ML semaglutide 2.27 MG/ML Auto-Injector [Wegovy] |
| NDC | 001694524 | semaglutide 2.4mg/.75mL SUBCUTANEOUS INJECTION, SOLUTION |
| NDC | 00169452401 | 0.75 ML semaglutide 3.2 MG/ML Auto-Injector [Wegovy] |
| NDC | 00169452414 | 0.75 ML semaglutide 3.2 MG/ML Auto-Injector [Wegovy] |
| NDC | 001694525 | semaglutide .25mg/.5mL SUBCUTANEOUS INJECTION, SOLUTION |
| NDC | 00169452501 | 0.5 ML semaglutide 0.5 MG/ML Auto-Injector [Wegovy] |
| NDC | 00169452514 | 0.5 ML semaglutide 0.5 MG/ML Auto-Injector [Wegovy] |
| NDC | 00169452590 | 0.5 ML semaglutide 0.5 MG/ML Auto-Injector [Wegovy] |
| NDC | 00169452594 | 0.5 ML semaglutide 0.5 MG/ML Auto-Injector [Wegovy] |
| NDC | 500905138 | semaglutide 1.34mg/mL SUBCUTANEOUS INJECTION, SOLUTION |

Continued on next page

**Table1 – continued from previous page**

| CodeSystem | ConceptCode | ConceptName |
|---|---|---|
| NDC | 50090513800 | 0.25 MG, 0.5 MG Dose 1.5 ML semaglutide 1.34 MG/ML Pen Injector [Ozempic] |
| NDC | 500905139 | semaglutide 1.34mg/mL SUBCUTANEOUS INJECTION, SOLUTION |
| NDC | 50090513900 | 1 MG Dose 1.5 ML semaglutide 1.34 MG/ML Pen Injector [Ozempic] |
| NDC | 500905824 | semaglutide .25mg/.5mL SUBCUTANEOUS INJECTION, SOLUTION |
| NDC | 50090582400 | 0.5 ML semaglutide 0.5 MG/ML Auto-Injector [Wegovy] |
| RxNorm | 2553502 | Wegovy |
| RxNorm | 2553504 | semaglutide Auto-Injector [Wegovy] |
| RxNorm | 2553506 | 0.5 ML semaglutide 0.5 MG/ML Auto-Injector [Wegovy] |
| RxNorm | 2553700 | semaglutide 2 MG/ML Auto-Injector [Wegovy] |
| RxNorm | 2553902 | semaglutide 2.27 MG/ML [Wegovy] |
| RxNorm | 2554103 | semaglutide 3.2 MG/ML [Wegovy] |
| RxNorm | 2553608 | semaglutide 2 MG/ML [Wegovy] |
| RxNorm | 2553905 | semaglutide 2.27 MG/ML Auto-Injector [Wegovy] |
| RxNorm | 2553505 | Wegovy Injectable Product |
| RxNorm | 2553605 | semaglutide 1 MG/ML Auto-Injector [Wegovy] |
| RxNorm | 2553903 | 0.75 ML semaglutide 2.27 MG/ML Auto-Injector [Wegovy] |
| RxNorm | 2554104 | 0.75 ML semaglutide 3.2 MG/ML Auto-Injector [Wegovy] |
| RxNorm | 2553803 | 0.5 ML semaglutide 2 MG/ML Auto-Injector [Wegovy] |
| RxNorm | 2553503 | semaglutide 0.5 MG/ML [Wegovy] |
| RxNorm | 2554106 | semaglutide 3.2 MG/ML Auto-Injector [Wegovy] |
| RxNorm | 2553508 | semaglutide 0.5 MG/ML Auto-Injector [Wegovy] |
| RxNorm | 2553602 | semaglutide 1 MG/ML [Wegovy] |
| RxNorm | 2553603 | 0.5 ML semaglutide 1 MG/ML Auto-Injector [Wegovy] |
| NDC | 001694501 | semaglutide 1mg/.5mL SUBCUTANEOUS INJECTION, SOLUTION |
| NDC | 00169450101 | 0.5 ML semaglutide 2 MG/ML Auto-Injector [Wegovy] |
| NDC | 00169450114 | 0.5 ML semaglutide 2 MG/ML Auto-Injector [Wegovy] |
| NDC | 001694505 | semaglutide .5mg/.5mL SUBCUTANEOUS INJECTION, SOLUTION |
| NDC | 00169450501 | 0.5 ML semaglutide 1 MG/ML Auto-Injector [Wegovy] |
| NDC | 00169450514 | 0.5 ML semaglutide 1 MG/ML Auto-Injector [Wegovy] |
| NDC | 001694517 | semaglutide 1.7mg/.75mL SUBCUTANEOUS INJECTION, SOLUTION |
| NDC | 00169451701 | 0.75 ML semaglutide 2.27 MG/ML Auto-Injector [Wegovy] |
| NDC | 00169451714 | 0.75 ML semaglutide 2.27 MG/ML Auto-Injector [Wegovy] |
| NDC | 001694524 | semaglutide 2.4mg/.75mL SUBCUTANEOUS INJECTION, SOLUTION |
| NDC | 00169452401 | 0.75 ML semaglutide 3.2 MG/ML Auto-Injector [Wegovy] |
| NDC | 00169452414 | 0.75 ML semaglutide 3.2 MG/ML Auto-Injector [Wegovy] |

Continued on next page

**Table1 – continued from previous page**

| CodeSystem | ConceptCode | ConceptName |
|---|---|---|
| NDC | 001694525 | semaglutide .25mg/.5mL SUBCUTANEOUS INJECTION, SOLUTION |
| NDC | 00169452501 | 0.5 ML semaglutide 0.5 MG/ML Auto-Injector [Wegovy] |
| NDC | 00169452514 | 0.5 ML semaglutide 0.5 MG/ML Auto-Injector [Wegovy] |
| NDC | 00169452590 | 0.5 ML semaglutide 0.5 MG/ML Auto-Injector [Wegovy] |
| NDC | 00169452594 | 0.5 ML semaglutide 0.5 MG/ML Auto-Injector [Wegovy] |
| NDC | 500905824 | semaglutide .25mg/.5mL SUBCUTANEOUS INJECTION, SOLUTION |
| NDC | 50090582400 | 0.5 ML semaglutide 0.5 MG/ML Auto-Injector [Wegovy] |
| RxNorm | 1991307 | Ozempic |
| RxNorm | 2599365 | 3 ML semaglutide 2.68 MG/ML Pen Injector [Ozempic] |
| RxNorm | 2599366 | semaglutide 2.68 MG/ML Pen Injector [Ozempic] |
| RxNorm | 1991308 | semaglutide 1.34 MG/ML [Ozempic] |
| RxNorm | 1991309 | semaglutide Pen Injector [Ozempic] |
| RxNorm | 1991310 | Ozempic Injectable Product |
| RxNorm | 1991311 | 0.25 MG, 0.5 MG Dose 1.5 ML semaglutide 1.34 MG/ML Pen Injector [Ozempic] |
| RxNorm | 1991317 | 1 MG Dose 1.5 ML semaglutide 1.34 MG/ML Pen Injector [Ozempic] |
| RxNorm | 2619154 | 0.25 MG, 0.5 MG Dose 3 ML semaglutide 0.68 MG/ML Pen Injector [Ozempic] |
| RxNorm | 2398842 | 3 ML semaglutide 1.34 MG/ML Pen Injector [Ozempic] |
| RxNorm | 2398844 | semaglutide 1.34 MG/ML Pen Injector [Ozempic] |
| NDC | 500905949 | semaglutide 1.34mg/mL SUBCUTANEOUS INJECTION, SOLUTION |
| NDC | 00169477290 | 3 ML semaglutide 2.68 MG/ML Pen Injector [Ozempic] |
| NDC | 00169477211 | 3 ML semaglutide 2.68 MG/ML Pen Injector [Ozempic] |
| NDC | 001694772 | semaglutide 2.68mg/mL SUBCUTANEOUS INJECTION, SOLUTION |
| NDC | 50090594900 | 3 ML semaglutide 1.34 MG/ML Pen Injector [Ozempic] |
| NDC | 00169477297 | 3 ML semaglutide 2.68 MG/ML Pen Injector [Ozempic] |
| NDC | 00169477212 | 3 ML semaglutide 2.68 MG/ML Pen Injector [Ozempic] |
| NDC | 705182143 | semaglutide 1.34mg/mL SUBCUTANEOUS INJECTION, SOLUTION |
| NDC | 70518214300 | 0.25 MG, 0.5 MG Dose 1.5 ML semaglutide 1.34 MG/ML Pen Injector [Ozempic] |
| NDC | 001694130 | semaglutide 1.34mg/mL SUBCUTANEOUS INJECTION, SOLUTION |
| NDC | 00169413001 | 3 ML semaglutide 1.34 MG/ML Pen Injector [Ozempic] |
| NDC | 00169413013 | 3 ML semaglutide 1.34 MG/ML Pen Injector [Ozempic] |
| NDC | 001694132 | semaglutide 1.34mg/mL SUBCUTANEOUS INJECTION, SOLUTION |

Continued on next page

**Table1 – continued from previous page**

| CodeSystem | ConceptCode | ConceptName |
|---|---|---|
| NDC | 00169413211 | 0.25 MG, 0.5 MG Dose 1.5 ML semaglutide 1.34 MG/ML Pen Injector [Ozempic] |
| NDC | 00169413212 | 0.25 MG, 0.5 MG Dose 1.5 ML semaglutide 1.34 MG/ML Pen Injector [Ozempic] |
| NDC | 00169413290 | 0.25 MG, 0.5 MG Dose 1.5 ML semaglutide 1.34 MG/ML Pen Injector [Ozempic] |
| NDC | 00169413297 | 0.25 MG, 0.5 MG Dose 1.5 ML semaglutide 1.34 MG/ML Pen Injector [Ozempic] |
| NDC | 001694136 | semaglutide 1.34mg/mL SUBCUTANEOUS INJECTION, SOLUTION |
| NDC | 00169413602 | 1 MG Dose 1.5 ML semaglutide 1.34 MG/ML Pen Injector [Ozempic] |
| NDC | 00169413611 | 1 MG Dose 1.5 ML semaglutide 1.34 MG/ML Pen Injector [Ozempic] |
| NDC | 500905138 | semaglutide 1.34mg/mL SUBCUTANEOUS INJECTION, SOLUTION |
| NDC | 50090513800 | 0.25 MG, 0.5 MG Dose 1.5 ML semaglutide 1.34 MG/ML Pen Injector [Ozempic] |
| NDC | 500905139 | semaglutide 1.34mg/mL SUBCUTANEOUS INJECTION, SOLUTION |
| NDC | 50090513900 | 1 MG Dose 1.5 ML semaglutide 1.34 MG/ML Pen Injector [Ozempic] |
| RxNorm | 2601723 | tirzepatide |
| RxNorm | 2601742 | tirzepatide 10 MG/ML |
| RxNorm | 2601745 | tirzepatide 10 MG/ML [Mounjaro] |
| RxNorm | 2601744 | tirzepatide 10 MG/ML Auto-Injector |
| RxNorm | 2601747 | tirzepatide 10 MG/ML Auto-Injector [Mounjaro] |
| RxNorm | 2601778 | tirzepatide 15 MG/ML |
| RxNorm | 2601781 | tirzepatide 15 MG/ML [Mounjaro] |
| RxNorm | 2601780 | tirzepatide 15 MG/ML Auto-Injector |
| RxNorm | 2601783 | tirzepatide 15 MG/ML Auto-Injector [Mounjaro] |
| RxNorm | 2601736 | Mounjaro Injectable Product |
| RxNorm | 2601759 | tirzepatide 30 MG/ML Auto-Injector [Mounjaro] |
| RxNorm | 2601760 | tirzepatide 5 MG/ML |
| RxNorm | 2601763 | tirzepatide 5 MG/ML [Mounjaro] |
| RxNorm | 2601762 | tirzepatide 5 MG/ML Auto-Injector |
| RxNorm | 2601765 | tirzepatide 5 MG/ML Auto-Injector [Mounjaro] |
| RxNorm | 2601731 | tirzepatide Auto-Injector |
| RxNorm | 2601737 | tirzepatide Auto-Injector [Mounjaro] |
| RxNorm | 2601730 | tirzepatide Injectable Product |
| RxNorm | 2601743 | 0.5 ML tirzepatide 10 MG/ML Auto-Injector |
| RxNorm | 2601746 | 0.5 ML tirzepatide 10 MG/ML Auto-Injector [Mounjaro] |
| RxNorm | 2601784 | 0.5 ML tirzepatide 15 MG/ML Auto-Injector |

Continued on next page

**Table1 – continued from previous page**

| CodeSystem | ConceptCode | ConceptName |
|---|---|---|
| RxNorm | 2601785 | 0.5 ML tirzepatide 15 MG/ML Auto-Injector [Mounjaro] |
| RxNorm | 2601767 | 0.5 ML tirzepatide 20 MG/ML Auto-Injector |
| RxNorm | 2601770 | 0.5 ML tirzepatide 20 MG/ML Auto-Injector [Mounjaro] |
| RxNorm | 2601773 | 0.5 ML tirzepatide 25 MG/ML Auto-Injector |
| RxNorm | 2601761 | 0.5 ML tirzepatide 5 MG/ML Auto-Injector |
| RxNorm | 2601764 | 0.5 ML tirzepatide 5 MG/ML Auto-Injector [Mounjaro] |
| RxNorm | 2601755 | 0.5 ML tirzepatide 30 MG/ML Auto-Injector |
| RxNorm | 2644401 | 0.5 ML tirzepatide 10 MG/ML Injection |
| RxNorm | 2644403 | 0.5 ML tirzepatide 10 MG/ML Injection [Mounjaro] |
| RxNorm | 2644396 | 0.5 ML tirzepatide 15 MG/ML Injection |
| RxNorm | 2644399 | 0.5 ML tirzepatide 15 MG/ML Injection [Mounjaro] |
| RxNorm | 2644417 | 0.5 ML tirzepatide 20 MG/ML Injection |
| RxNorm | 2644419 | 0.5 ML tirzepatide 20 MG/ML Injection [Mounjaro] |
| RxNorm | 2644413 | 0.5 ML tirzepatide 25 MG/ML Injection |
| RxNorm | 2644415 | 0.5 ML tirzepatide 25 MG/ML Injection [Mounjaro] |
| RxNorm | 2644409 | 0.5 ML tirzepatide 30 MG/ML Injection |
| RxNorm | 2644411 | 0.5 ML tirzepatide 30 MG/ML Injection [Mounjaro] |
| RxNorm | 2644405 | 0.5 ML tirzepatide 5 MG/ML Injection |
| RxNorm | 2644407 | 0.5 ML tirzepatide 5 MG/ML Injection [Mounjaro] |
| RxNorm | 2601769 | tirzepatide 20 MG/ML [Mounjaro] |
| RxNorm | 2601757 | tirzepatide 30 MG/ML [Mounjaro] |
| RxNorm | 2644402 | tirzepatide 10 MG/ML Injection |
| RxNorm | 2644404 | tirzepatide 10 MG/ML Injection [Mounjaro] |
| RxNorm | 2644397 | tirzepatide 15 MG/ML Injection |
| RxNorm | 2644400 | tirzepatide 15 MG/ML Injection [Mounjaro] |
| RxNorm | 2644418 | tirzepatide 20 MG/ML Injection |
| RxNorm | 2644420 | tirzepatide 20 MG/ML Injection [Mounjaro] |
| RxNorm | 2644414 | tirzepatide 25 MG/ML Injection |
| RxNorm | 2644416 | tirzepatide 25 MG/ML Injection [Mounjaro] |
| RxNorm | 2644410 | tirzepatide 30 MG/ML Injection |
| RxNorm | 2644412 | tirzepatide 30 MG/ML Injection [Mounjaro] |
| RxNorm | 2644406 | tirzepatide 5 MG/ML Injection |
| RxNorm | 2644408 | tirzepatide 5 MG/ML Injection [Mounjaro] |
| RxNorm | 2644395 | tirzepatide Injection |
| RxNorm | 2644398 | tirzepatide Injection [Mounjaro] |
| RxNorm | 2601776 | 0.5 ML tirzepatide 25 MG/ML Auto-Injector [Mounjaro] |
| RxNorm | 2601756 | tirzepatide 30 MG/ML Auto-Injector |
| RxNorm | 2601771 | tirzepatide 20 MG/ML Auto-Injector [Mounjaro] |
| RxNorm | 2601772 | tirzepatide 25 MG/ML |
| RxNorm | 2601775 | tirzepatide 25 MG/ML [Mounjaro] |
| RxNorm | 2601774 | tirzepatide 25 MG/ML Auto-Injector |
| RxNorm | 2601768 | tirzepatide 20 MG/ML Auto-Injector |
| RxNorm | 2601777 | tirzepatide 25 MG/ML Auto-Injector [Mounjaro] |

Continued on next page

**Table1 – continued from previous page**

| CodeSystem | ConceptCode | ConceptName |
|---|---|---|
| RxNorm | 2601754 | tirzepatide 30 MG/ML |
| RxNorm | 2601766 | tirzepatide 20 MG/ML |
| RxNorm | 2601758 | 0.5 ML tirzepatide 30 MG/ML Auto-Injector [Mounjaro] |
| NDC | 00002150661 | tirzepatide 2.5mg/.5mL SUBCUTANEOUS INJECTION, SOLUTION |
| NDC | 00002147180 | 0.5 ML tirzepatide 20 MG/ML Auto-Injector [Mounjaro] |
| NDC | 00002146080 | 0.5 ML tirzepatide 25 MG/ML Auto-Injector [Mounjaro] |
| NDC | 00002150680 | 0.5 ML tirzepatide 5 MG/ML Auto-Injector [Mounjaro] |
| NDC | 00002145780 | 0.5 ML tirzepatide 30 MG/ML Auto-Injector [Mounjaro] |
| NDC | 000021471 | tirzepatide 10mg/.5mL SUBCUTANEOUS INJECTION, SOLUTION |
| NDC | 000021460 | tirzepatide 12.5mg/.5mL SUBCUTANEOUS INJECTION, SOLUTION |
| NDC | 000021484 | tirzepatide 7.5mg/.5mL SUBCUTANEOUS INJECTION, SOLUTION |
| NDC | 00002146001 | 0.5 ML tirzepatide 25 MG/ML Auto-Injector [Mounjaro] |
| NDC | 00002149580 | 0.5 ML tirzepatide 10 MG/ML Auto-Injector [Mounjaro] |
| NDC | 00002148480 | 0.5 ML tirzepatide 15 MG/ML Auto-Injector [Mounjaro] |
| NDC | 000021506 | tirzepatide 2.5mg/.5mL SUBCUTANEOUS INJECTION, SOLUTION |
| NDC | 00002150601 | 0.5 ML tirzepatide 5 MG/ML Auto-Injector [Mounjaro] |
| NDC | 00002145701 | 0.5 ML tirzepatide 30 MG/ML Auto-Injector [Mounjaro] |
| NDC | 00002147101 | 0.5 ML tirzepatide 20 MG/ML Auto-Injector [Mounjaro] |
| NDC | 00002148401 | 0.5 ML tirzepatide 15 MG/ML Auto-Injector [Mounjaro] |
| NDC | 00002149501 | 0.5 ML tirzepatide 10 MG/ML Auto-Injector [Mounjaro] |
| NDC | 000021457 | tirzepatide 15mg/.5mL SUBCUTANEOUS INJECTION, SOLUTION |
| NDC | 000021495 | tirzepatide 5mg/.5mL SUBCUTANEOUS INJECTION, SOLUTION |
| RxNorm | 2601745 | tirzepatide 10 MG/ML [Mounjaro] |
| RxNorm | 2601747 | tirzepatide 10 MG/ML Auto-Injector [Mounjaro] |
| RxNorm | 2601781 | tirzepatide 15 MG/ML [Mounjaro] |
| RxNorm | 2601783 | tirzepatide 15 MG/ML Auto-Injector [Mounjaro] |
| RxNorm | 2601734 | Mounjaro |
| RxNorm | 2601759 | tirzepatide 30 MG/ML Auto-Injector [Mounjaro] |
| RxNorm | 2601763 | tirzepatide 5 MG/ML [Mounjaro] |
| RxNorm | 2601765 | tirzepatide 5 MG/ML Auto-Injector [Mounjaro] |
| RxNorm | 2601737 | tirzepatide Auto-Injector [Mounjaro] |
| RxNorm | 2601746 | 0.5 ML tirzepatide 10 MG/ML Auto-Injector [Mounjaro] |
| RxNorm | 2601785 | 0.5 ML tirzepatide 15 MG/ML Auto-Injector [Mounjaro] |
| RxNorm | 2601770 | 0.5 ML tirzepatide 20 MG/ML Auto-Injector [Mounjaro] |
| RxNorm | 2601764 | 0.5 ML tirzepatide 5 MG/ML Auto-Injector [Mounjaro] |
| RxNorm | 2601769 | tirzepatide 20 MG/ML [Mounjaro] |

Continued on next page

**Table1 – continued from previous page**

| CodeSystem | ConceptCode | ConceptName |
|---|---|---|
| RxNorm | 2601757 | tirzepatide 30 MG/ML [Mounjaro] |
| RxNorm | 2601776 | 0.5 ML tirzepatide 25 MG/ML Auto-Injector [Mounjaro] |
| RxNorm | 2601771 | tirzepatide 20 MG/ML Auto-Injector [Mounjaro] |
| RxNorm | 2601777 | tirzepatide 25 MG/ML Auto-Injector [Mounjaro] |
| RxNorm | 2601758 | 0.5 ML tirzepatide 30 MG/ML Auto-Injector [Mounjaro] |
| RxNorm | 2644403 | 0.5 ML tirzepatide 10 MG/ML Injection [Mounjaro] |
| RxNorm | 2644399 | 0.5 ML tirzepatide 15 MG/ML Injection [Mounjaro] |
| RxNorm | 2644419 | 0.5 ML tirzepatide 20 MG/ML Injection [Mounjaro] |
| RxNorm | 2644411 | 0.5 ML tirzepatide 30 MG/ML Injection [Mounjaro] |
| RxNorm | 2644407 | 0.5 ML tirzepatide 5 MG/ML Injection [Mounjaro] |
| RxNorm | 2644404 | tirzepatide 10 MG/ML Injection [Mounjaro] |
| RxNorm | 2644400 | tirzepatide 15 MG/ML Injection [Mounjaro] |
| RxNorm | 2644420 | tirzepatide 20 MG/ML Injection [Mounjaro] |
| RxNorm | 2644412 | tirzepatide 30 MG/ML Injection [Mounjaro] |
| RxNorm | 2644408 | tirzepatide 5 MG/ML Injection [Mounjaro] |
| NDC | 00002150661 | tirzepatide 2.5mg/.5mL SUBCUTANEOUS INJECTION, SOLUTION |
| NDC | 00002147180 | 0.5 ML tirzepatide 20 MG/ML Auto-Injector [Mounjaro] |
| NDC | 00002146080 | 0.5 ML tirzepatide 25 MG/ML Auto-Injector [Mounjaro] |
| NDC | 00002150680 | 0.5 ML tirzepatide 5 MG/ML Auto-Injector [Mounjaro] |
| NDC | 00002145780 | 0.5 ML tirzepatide 30 MG/ML Auto-Injector [Mounjaro] |
| NDC | 000021471 | tirzepatide 10mg/.5mL SUBCUTANEOUS INJECTION, SOLUTION |
| NDC | 000021460 | tirzepatide 12.5mg/.5mL SUBCUTANEOUS INJECTION, SOLUTION |
| NDC | 000021484 | tirzepatide 7.5mg/.5mL SUBCUTANEOUS INJECTION, SOLUTION |
| NDC | 00002146001 | 0.5 ML tirzepatide 25 MG/ML Auto-Injector [Mounjaro] |
| NDC | 00002149580 | 0.5 ML tirzepatide 10 MG/ML Auto-Injector [Mounjaro] |
| NDC | 00002148480 | 0.5 ML tirzepatide 15 MG/ML Auto-Injector [Mounjaro] |
| NDC | 000021506 | tirzepatide 2.5mg/.5mL SUBCUTANEOUS INJECTION, SOLUTION |
| NDC | 00002150601 | 0.5 ML tirzepatide 5 MG/ML Auto-Injector [Mounjaro] |
| NDC | 00002145701 | 0.5 ML tirzepatide 30 MG/ML Auto-Injector [Mounjaro] |
| NDC | 00002147101 | 0.5 ML tirzepatide 20 MG/ML Auto-Injector [Mounjaro] |
| NDC | 00002148401 | 0.5 ML tirzepatide 15 MG/ML Auto-Injector [Mounjaro] |
| NDC | 00002149501 | 0.5 ML tirzepatide 10 MG/ML Auto-Injector [Mounjaro] |
| NDC | 000021457 | tirzepatide 15mg/.5mL SUBCUTANEOUS INJECTION, SOLUTION |
| NDC | 000021495 | tirzepatide 5mg/.5mL SUBCUTANEOUS INJECTION, SOLUTION |
| RxNorm | 1551291 | dulaglutide |
| RxNorm | 1551294 | dulaglutide Prefilled Syringe |

Continued on next page

**Table1 – continued from previous page**

| CodeSystem | ConceptCode | ConceptName |
|---|---|---|
| RxNorm | 1551298 | dulaglutide Prefilled Syringe [Trulicity] |
| RxNorm | 1551292 | dulaglutide 1.5 MG/ML |
| RxNorm | 1551293 | dulaglutide Injectable Product |
| RxNorm | 1551295 | 0.5 ML dulaglutide 1.5 MG/ML Auto-Injector |
| RxNorm | 1551296 | Trulicity |
| RxNorm | 1551297 | dulaglutide 1.5 MG/ML [Trulicity] |
| RxNorm | 1551299 | Trulicity Injectable Product |
| RxNorm | 1551300 | 0.5 ML dulaglutide 1.5 MG/ML Auto-Injector [Trulicity] |
| RxNorm | 1551301 | dulaglutide 1.5 MG/ML Auto-Injector |
| RxNorm | 1551302 | dulaglutide 1.5 MG/ML Auto-Injector [Trulicity] |
| RxNorm | 1551303 | dulaglutide 3 MG/ML |
| RxNorm | 1551304 | 0.5 ML dulaglutide 3 MG/ML Auto-Injector |
| RxNorm | 1551305 | dulaglutide 3 MG/ML [Trulicity] |
| RxNorm | 1551306 | 0.5 ML dulaglutide 3 MG/ML Auto-Injector [Trulicity] |
| RxNorm | 1551307 | dulaglutide 3 MG/ML Auto-Injector |
| RxNorm | 1551308 | dulaglutide 3 MG/ML Auto-Injector [Trulicity] |
| RxNorm | 1649584 | dulaglutide Auto-Injector |
| RxNorm | 1649586 | dulaglutide Auto-Injector [Trulicity] |
| RxNorm | 2395776 | dulaglutide 6 MG/ML |
| RxNorm | 2395777 | 0.5 ML dulaglutide 6 MG/ML Auto-Injector |
| RxNorm | 2395778 | dulaglutide 6 MG/ML [Trulicity] |
| RxNorm | 2395779 | 0.5 ML dulaglutide 6 MG/ML Auto-Injector [Trulicity] |
| RxNorm | 2395780 | dulaglutide 6 MG/ML Auto-Injector |
| RxNorm | 2395781 | dulaglutide 6 MG/ML Auto-Injector [Trulicity] |
| RxNorm | 2395782 | dulaglutide 9 MG/ML |
| RxNorm | 2395783 | 0.5 ML dulaglutide 9 MG/ML Auto-Injector |
| RxNorm | 2395784 | dulaglutide 9 MG/ML [Trulicity] |
| RxNorm | 2395785 | 0.5 ML dulaglutide 9 MG/ML Auto-Injector [Trulicity] |
| RxNorm | 2395786 | dulaglutide 9 MG/ML Auto-Injector |
| RxNorm | 2395787 | dulaglutide 9 MG/ML Auto-Injector [Trulicity] |
| SNOMED CT | 714080005 | Dulaglutide |
| SNOMED CT | 714081009 | Dulaglutide-containing product |
| SNOMED CT | 775712006 | Dulaglutide only product |
| SNOMED CT | 1010536005 | Dulaglutide 1.5 mg/mL solution for injection |
| SNOMED CT | 1010537001 | Dulaglutide 3 mg/mL solution for injection |
| SNOMED CT | 1010538006 | Dulaglutide 6 mg/mL solution for injection |
| SNOMED CT | 1010539003 | Dulaglutide 9 mg/mL solution for injection |
| SNOMED CT | 1010540001 | Dulaglutide-containing product in parenteral dose form |
| SNOMED CT | 1010541002 | Dulaglutide only product in parenteral dose form |
| NDC | 545680433 | dulaglutide .75mg/.5mL SUBCUTANEOUS INJECTION, SOLUTION [trulicity] |
| NDC | 54568043363 | 0.5 ML dulaglutide 1.5 MG/ML Auto-Injector [Trulicity] |
| NDC | 54568043371 | 0.5 ML dulaglutide 1.5 MG/ML Auto-Injector [Trulicity] |

Continued on next page

**Table1 – continued from previous page**

| CodeSystem | ConceptCode | ConceptName |
|---|---|---|
| NDC | 545680434 | dulaglutide 1.5mg/.5mL SUBCUTANEOUS INJECTION, SOLUTION [trulicity] |
| NDC | 54568043463 | 0.5 ML dulaglutide 3 MG/ML Auto-Injector [Trulicity] |
| NDC | 54568043471 | 0.5 ML dulaglutide 3 MG/ML Auto-Injector [Trulicity] |
| NDC | 000021433 | dulaglutide .75mg/.5mL SUBCUTANEOUS INJECTION, SOLUTION |
| NDC | 00002143301 | 0.5 ML dulaglutide 1.5 MG/ML Auto-Injector [Trulicity] |
| NDC | 00002143361 | 0.5 ML dulaglutide 1.5 MG/ML Auto-Injector [Trulicity] |
| NDC | 00002143380 | 0.5 ML dulaglutide 1.5 MG/ML Auto-Injector [Trulicity] |
| NDC | 000021434 | dulaglutide 1.5mg/.5mL SUBCUTANEOUS INJECTION, SOLUTION |
| NDC | 00002143401 | 0.5 ML dulaglutide 3 MG/ML Auto-Injector [Trulicity] |
| NDC | 00002143461 | 0.5 ML dulaglutide 3 MG/ML Auto-Injector [Trulicity] |
| NDC | 00002143480 | 0.5 ML dulaglutide 3 MG/ML Auto-Injector [Trulicity] |
| NDC | 000022236 | dulaglutide 3mg/.5mL SUBCUTANEOUS INJECTION, SOLUTION |
| NDC | 00002223601 | 0.5 ML dulaglutide 6 MG/ML Auto-Injector [Trulicity] |
| NDC | 00002223661 | 0.5 ML dulaglutide 6 MG/ML Auto-Injector [Trulicity] |
| NDC | 00002223680 | 0.5 ML dulaglutide 6 MG/ML Auto-Injector [Trulicity] |
| NDC | 000023182 | dulaglutide 4.5mg/.5mL SUBCUTANEOUS INJECTION, SOLUTION |
| NDC | 00002318201 | 0.5 ML dulaglutide 9 MG/ML Auto-Injector [Trulicity] |
| NDC | 00002318261 | 0.5 ML dulaglutide 9 MG/ML Auto-Injector [Trulicity] |
| NDC | 00002318280 | 0.5 ML dulaglutide 9 MG/ML Auto-Injector [Trulicity] |
| NDC | 500903483 | dulaglutide 1.5mg/.5mL SUBCUTANEOUS INJECTION, SOLUTION |
| NDC | 50090348300 | 0.5 ML dulaglutide 3 MG/ML Auto-Injector [Trulicity] |
| NDC | 500903484 | dulaglutide .75mg/.5mL SUBCUTANEOUS INJECTION, SOLUTION |
| NDC | 50090348400 | 0.5 ML dulaglutide 1.5 MG/ML Auto-Injector [Trulicity] |
| NDC | 500905467 | dulaglutide 3mg/.5mL SUBCUTANEOUS INJECTION, SOLUTION |
| NDC | 50090546700 | 0.5 ML dulaglutide 6 MG/ML Auto-Injector [Trulicity] |
| RxNorm | 1551296 | Trulicity |
| RxNorm | 1551298 | dulaglutide Prefilled Syringe [Trulicity] |
| RxNorm | 1551297 | dulaglutide 1.5 MG/ML [Trulicity] |
| RxNorm | 1551299 | Trulicity Injectable Product |
| RxNorm | 1551300 | 0.5 ML dulaglutide 1.5 MG/ML Auto-Injector [Trulicity] |
| RxNorm | 1551302 | dulaglutide 1.5 MG/ML Auto-Injector [Trulicity] |
| RxNorm | 1551305 | dulaglutide 3 MG/ML [Trulicity] |
| RxNorm | 1551306 | 0.5 ML dulaglutide 3 MG/ML Auto-Injector [Trulicity] |
| RxNorm | 1551308 | dulaglutide 3 MG/ML Auto-Injector [Trulicity] |
| RxNorm | 1649586 | dulaglutide Auto-Injector [Trulicity] |

Continued on next page

**Table1 – continued from previous page**

| CodeSystem | ConceptCode | ConceptName |
|---|---|---|
| RxNorm | 2395778 | dulaglutide 6 MG/ML [Trulicity] |
| RxNorm | 2395779 | 0.5 ML dulaglutide 6 MG/ML Auto-Injector [Trulicity] |
| RxNorm | 2395781 | dulaglutide 6 MG/ML Auto-Injector [Trulicity] |
| RxNorm | 2395784 | dulaglutide 9 MG/ML [Trulicity] |
| RxNorm | 2395785 | 0.5 ML dulaglutide 9 MG/ML Auto-Injector [Trulicity] |
| RxNorm | 2395787 | dulaglutide 9 MG/ML Auto-Injector [Trulicity] |
| NDC | 545680433 | dulaglutide .75mg/.5mL SUBCUTANEOUS INJECTION, SOLUTION [trulicity] |
| NDC | 54568043363 | 0.5 ML dulaglutide 1.5 MG/ML Auto-Injector [Trulicity] |
| NDC | 54568043371 | 0.5 ML dulaglutide 1.5 MG/ML Auto-Injector [Trulicity] |
| NDC | 545680434 | dulaglutide 1.5mg/.5mL SUBCUTANEOUS INJECTION, SOLUTION [trulicity] |
| NDC | 54568043463 | 0.5 ML dulaglutide 3 MG/ML Auto-Injector [Trulicity] |
| NDC | 54568043471 | 0.5 ML dulaglutide 3 MG/ML Auto-Injector [Trulicity] |
| NDC | 000021433 | dulaglutide .75mg/.5mL SUBCUTANEOUS INJECTION, SOLUTION |
| NDC | 00002143301 | 0.5 ML dulaglutide 1.5 MG/ML Auto-Injector [Trulicity] |
| NDC | 00002143361 | 0.5 ML dulaglutide 1.5 MG/ML Auto-Injector [Trulicity] |
| NDC | 00002143380 | 0.5 ML dulaglutide 1.5 MG/ML Auto-Injector [Trulicity] |
| NDC | 000021434 | dulaglutide 1.5mg/.5mL SUBCUTANEOUS INJECTION, SOLUTION |
| NDC | 00002143401 | 0.5 ML dulaglutide 3 MG/ML Auto-Injector [Trulicity] |
| NDC | 00002143461 | 0.5 ML dulaglutide 3 MG/ML Auto-Injector [Trulicity] |
| NDC | 00002143480 | 0.5 ML dulaglutide 3 MG/ML Auto-Injector [Trulicity] |
| NDC | 000022236 | dulaglutide 3mg/.5mL SUBCUTANEOUS INJECTION, SOLUTION |
| NDC | 00002223601 | 0.5 ML dulaglutide 6 MG/ML Auto-Injector [Trulicity] |
| NDC | 00002223661 | 0.5 ML dulaglutide 6 MG/ML Auto-Injector [Trulicity] |
| NDC | 00002223680 | 0.5 ML dulaglutide 6 MG/ML Auto-Injector [Trulicity] |
| NDC | 000023182 | dulaglutide 4.5mg/.5mL SUBCUTANEOUS INJECTION, SOLUTION |
| NDC | 00002318201 | 0.5 ML dulaglutide 9 MG/ML Auto-Injector [Trulicity] |
| NDC | 00002318261 | 0.5 ML dulaglutide 9 MG/ML Auto-Injector [Trulicity] |
| NDC | 00002318280 | 0.5 ML dulaglutide 9 MG/ML Auto-Injector [Trulicity] |
| NDC | 500903483 | dulaglutide 1.5mg/.5mL SUBCUTANEOUS INJECTION, SOLUTION |
| NDC | 50090348300 | 0.5 ML dulaglutide 3 MG/ML Auto-Injector [Trulicity] |
| NDC | 500903484 | dulaglutide .75mg/.5mL SUBCUTANEOUS INJECTION, SOLUTION |
| NDC | 50090348400 | 0.5 ML dulaglutide 1.5 MG/ML Auto-Injector [Trulicity] |
| NDC | 500905467 | dulaglutide 3mg/.5mL SUBCUTANEOUS INJECTION, SOLUTION |
| NDC | 50090546700 | 0.5 ML dulaglutide 6 MG/ML Auto-Injector [Trulicity] |

Continued on next page

**Table1 – continued from previous page**

| CodeSystem | ConceptCode | ConceptName |
|---|---|---|
| RxNorm | 1440051 | lixisenatide |
| RxNorm | 1440054 | Lixisenatide Injectable Solution |
| RxNorm | 1440055 | Lixisenatide 0.1 MG/ML Injectable Solution |
| RxNorm | 1440057 | Lixisenatide 0.05 MG/ML Injectable Solution |
| RxNorm | 1440052 | lixisenatide 0.1 MG/ML |
| RxNorm | 1440053 | lixisenatide Injectable Product |
| RxNorm | 1440056 | lixisenatide 0.05 MG/ML |
| RxNorm | 1858991 | lixisenatide 0.033 MG/ML |
| RxNorm | 1858992 | insulin glargine / lixisenatide Injectable Product |
| RxNorm | 1858993 | insulin glargine / lixisenatide Pen Injector |
| RxNorm | 1858994 | insulin glargine / lixisenatide |
| RxNorm | 1858995 | 3 ML insulin glargine 100 UNT/ML / lixisenatide 0.033 MG/ML Pen Injector |
| RxNorm | 1858996 | Soliqua |
| RxNorm | 1858997 | insulin glargine 100 UNT/ML / lixisenatide 0.033 MG/ML [Soliqua] |
| RxNorm | 1858998 | insulin glargine / lixisenatide Pen Injector [Soliqua] |
| RxNorm | 1858999 | Soliqua Injectable Product |
| RxNorm | 1859000 | 3 ML insulin glargine 100 UNT/ML / lixisenatide 0.033 MG/ML Pen Injector [Soliqua] |
| RxNorm | 1859001 | insulin glargine 100 UNT/ML / lixisenatide 0.033 MG/ML Pen Injector |
| RxNorm | 1859002 | insulin glargine 100 UNT/ML / lixisenatide 0.033 MG/ML Pen Injector [Soliqua] |
| RxNorm | 1803885 | lixisenatide Pen Injector |
| RxNorm | 1803886 | lixisenatide 0.05 MG/ML Pen Injector |
| RxNorm | 1803887 | Adlyxin |
| RxNorm | 1803888 | lixisenatide 0.05 MG/ML [Adlyxin] |
| RxNorm | 1803889 | lixisenatide Pen Injector [Adlyxin] |
| RxNorm | 1803890 | Adlyxin Injectable Product |
| RxNorm | 1803891 | lixisenatide 0.05 MG/ML Pen Injector [Adlyxin] |
| RxNorm | 1803892 | 3 ML lixisenatide 0.05 MG/ML Pen Injector |
| RxNorm | 1803893 | 3 ML lixisenatide 0.05 MG/ML Pen Injector [Adlyxin] |
| RxNorm | 1803894 | 3 ML lixisenatide 0.1 MG/ML Pen Injector |
| RxNorm | 1803895 | lixisenatide 0.1 MG/ML [Adlyxin] |
| RxNorm | 1803896 | 3 ML lixisenatide 0.1 MG/ML Pen Injector [Adlyxin] |
| RxNorm | 1803897 | lixisenatide 0.1 MG/ML Pen Injector |
| RxNorm | 1803898 | lixisenatide 0.1 MG/ML Pen Injector [Adlyxin] |
| RxNorm | 1803902 | {1 (3 ML lixisenatide 0.05 MG/ML Pen Injector) / 1 (3 ML lixisenatide 0.1 MG/ML Pen Injector) } Pack |
| RxNorm | 1803903 | {1 (3 ML lixisenatide 0.05 MG/ML Pen Injector [Adlyxin]) / 1 (3 ML lixisenatide 0.1 MG/ML Pen Injector [Adlyxin]) } Pack [Adlyxin Starter Kit] |

Continued on next page

**Table1 – continued from previous page**

| CodeSystem | ConceptCode | ConceptName |
|---|---|---|
| SNOMED CT | 763570007 | Lixisenatide-containing product |
| SNOMED CT | 764330009 | Lixisenatide-containing product in parenteral dose form |
| SNOMED CT | 708808004 | Lixisenatide |
| SNOMED CT | 776560001 | Lixisenatide only product |
| SNOMED CT | 779728007 | Lixisenatide only product in parenteral dose form |
| NDC | 00024574000 | 3 ML lixisenatide 0.1 MG/ML Pen Injector [Adlyxin] |
| NDC | 00024574101 | 3 ML lixisenatide 0.05 MG/ML Pen Injector [Adlyxin] |
| NDC | 000245745 | lixisenatide KIT |
| NDC | 00024574502 | {1 (3 ML lixisenatide 0.05 MG/ML Pen Injector [Adlyxin]) / 1 (3 ML lixisenatide 0.1 MG/ML Pen Injector [Adlyxin]) } Pack [Adlyxin Starter Kit] |
| NDC | 000245747 | lixisenatide 100ug/mL SUBCUTANEOUS INJECTION, SOLUTION |
| NDC | 00024574702 | 3 ML lixisenatide 0.1 MG/ML Pen Injector [Adlyxin] |
| NDC | 000245761 | insulin glargine and lixisenatide 100U/mL / 33ug/mL SUBCUTANEOUS INJECTION, SOLUTION |
| NDC | 00024576101 | 3 ML insulin glargine 100 UNT/ML / lixisenatide 0.033 MG/ML Pen Injector [Soliqua] |
| NDC | 00024576102 | 3 ML insulin glargine 100 UNT/ML / lixisenatide 0.033 MG/ML Pen Injector [Soliqua] |
| NDC | 00024576105 | 3 ML insulin glargine 100 UNT/ML / lixisenatide 0.033 MG/ML Pen Injector [Soliqua] |
| NDC | 00024576302 | 3 ML insulin glargine 100 UNT/ML / lixisenatide 0.033 MG/ML Pen Injector [Soliqua] |
| RxNorm | 1858996 | Soliqua |
| RxNorm | 1858997 | insulin glargine 100 UNT/ML / lixisenatide 0.033 MG/ML [Soliqua] |
| RxNorm | 1858998 | insulin glargine / lixisenatide Pen Injector [Soliqua] |
| RxNorm | 1858999 | Soliqua Injectable Product |
| RxNorm | 1859000 | 3 ML insulin glargine 100 UNT/ML / lixisenatide 0.033 MG/ML Pen Injector [Soliqua] |
| RxNorm | 1859002 | insulin glargine 100 UNT/ML / lixisenatide 0.033 MG/ML Pen Injector [Soliqua] |
| NDC | 000245761 | insulin glargine and lixisenatide 100U/mL / 33ug/mL SUBCUTANEOUS INJECTION, SOLUTION |
| NDC | 00024576101 | 3 ML insulin glargine 100 UNT/ML / lixisenatide 0.033 MG/ML Pen Injector [Soliqua] |
| NDC | 00024576102 | 3 ML insulin glargine 100 UNT/ML / lixisenatide 0.033 MG/ML Pen Injector [Soliqua] |
| NDC | 00024576105 | 3 ML insulin glargine 100 UNT/ML / lixisenatide 0.033 MG/ML Pen Injector [Soliqua] |
| NDC | 00024576302 | 3 ML insulin glargine 100 UNT/ML / lixisenatide 0.033 MG/ML Pen Injector [Soliqua] |

Continued on next page

**Table1 – continued from previous page**

| CodeSystem | ConceptCode | ConceptName |
|---|---|---|
| RxNorm | 1803887 | Adlyxin |
| RxNorm | 1803888 | lixisenatide 0.05 MG/ML [Adlyxin] |
| RxNorm | 1803889 | lixisenatide Pen Injector [Adlyxin] |
| RxNorm | 1803890 | Adlyxin Injectable Product |
| RxNorm | 1803891 | lixisenatide 0.05 MG/ML Pen Injector [Adlyxin] |
| RxNorm | 1803893 | 3 ML lixisenatide 0.05 MG/ML Pen Injector [Adlyxin] |
| RxNorm | 1803895 | lixisenatide 0.1 MG/ML [Adlyxin] |
| RxNorm | 1803896 | 3 ML lixisenatide 0.1 MG/ML Pen Injector [Adlyxin] |
| RxNorm | 1803898 | lixisenatide 0.1 MG/ML Pen Injector [Adlyxin] |
| RxNorm | 1803903 | {1 (3 ML lixisenatide 0.05 MG/ML Pen Injector [Adlyxin]) / 1 (3 ML lixisenatide 0.1 MG/ML Pen Injector [Adlyxin]) } Pack [Adlyxin Starter Kit] |
| NDC | 00024574000 | 3 ML lixisenatide 0.1 MG/ML Pen Injector [Adlyxin] |
| NDC | 00024574101 | 3 ML lixisenatide 0.05 MG/ML Pen Injector [Adlyxin] |
| NDC | 000245745 | lixisenatide KIT |
| NDC | 00024574502 | {1 (3 ML lixisenatide 0.05 MG/ML Pen Injector [Adlyxin]) / 1 (3 ML lixisenatide 0.1 MG/ML Pen Injector [Adlyxin]) } Pack [Adlyxin Starter Kit] |
| NDC | 000245747 | lixisenatide 100ug/mL SUBCUTANEOUS INJECTION, SOLUTION |
| NDC | 00024574702 | 3 ML lixisenatide 0.1 MG/ML Pen Injector [Adlyxin] |
| RxNorm | 475968 | liraglutide |
| RxNorm | 1598266 | liraglutide Prefilled Syringe [Saxenda] |
| RxNorm | 897125 | liraglutide Prefilled Syringe [Victoza] |
| RxNorm | 897121 | liraglutide Prefilled Syringe |
| RxNorm | 897120 | liraglutide 6 MG/ML |
| RxNorm | 897122 | 3 ML liraglutide 6 MG/ML Pen Injector |
| RxNorm | 897123 | Victoza |
| RxNorm | 897124 | liraglutide 6 MG/ML [Victoza] |
| RxNorm | 897126 | 3 ML liraglutide 6 MG/ML Pen Injector [Victoza] |
| RxNorm | 1186578 | Victoza Injectable Product |
| RxNorm | 1163230 | liraglutide Injectable Product |
| RxNorm | 1360105 | liraglutide 6 MG/ML Pen Injector |
| RxNorm | 1360495 | liraglutide 6 MG/ML Pen Injector [Victoza] |
| RxNorm | 1598264 | Saxenda |
| RxNorm | 1598265 | liraglutide 6 MG/ML [Saxenda] |
| RxNorm | 1598267 | Saxenda Injectable Product |
| RxNorm | 1598268 | 3 ML liraglutide 6 MG/ML Pen Injector [Saxenda] |
| RxNorm | 1598269 | liraglutide 6 MG/ML Pen Injector [Saxenda] |
| RxNorm | 1653594 | liraglutide Pen Injector |
| RxNorm | 1653597 | liraglutide Pen Injector [Victoza] |
| RxNorm | 1653600 | liraglutide Pen Injector [Saxenda] |
| RxNorm | 1727493 | insulin degludec / liraglutide |

Continued on next page

**Table1 – continued from previous page**

| CodeSystem | ConceptCode | ConceptName |
|---|---|---|
| RxNorm | 1860164 | liraglutide 3.6 MG/ML |
| RxNorm | 1860165 | insulin degludec / liraglutide Injectable Product |
| RxNorm | 1860166 | insulin degludec / liraglutide Pen Injector |
| RxNorm | 1860167 | 3 ML insulin degludec 100 UNT/ML / liraglutide 3.6 MG/ML Pen Injector |
| RxNorm | 1860168 | Xultophy |
| RxNorm | 1860169 | insulin degludec 100 UNT/ML / liraglutide 3.6 MG/ML [Xultophy] |
| RxNorm | 1860170 | insulin degludec / liraglutide Pen Injector [Xultophy] |
| RxNorm | 1860171 | Xultophy Injectable Product |
| RxNorm | 1860172 | 3 ML insulin degludec 100 UNT/ML / liraglutide 3.6 MG/ML Pen Injector [Xultophy] |
| RxNorm | 1860173 | insulin degludec 100 UNT/ML / liraglutide 3.6 MG/ML Pen Injector |
| RxNorm | 1860174 | insulin degludec 100 UNT/ML / liraglutide 3.6 MG/ML Pen Injector [Xultophy] |
| NDC | 54569650700 | 3 ML liraglutide 6 MG/ML Pen Injector [Victoza] |
| NDC | 001692800 | liraglutide 6mg/mL SUBCUTANEOUS INJECTION, SOLUTION |
| NDC | 00169280013 | 3 ML liraglutide 6 MG/ML Pen Injector [Saxenda] |
| NDC | 00169280015 | 3 ML liraglutide 6 MG/ML Pen Injector [Saxenda] |
| NDC | 00169280090 | 3 ML liraglutide 6 MG/ML Pen Injector [Saxenda] |
| NDC | 00169280097 | 3 ML liraglutide 6 MG/ML Pen Injector [Saxenda] |
| NDC | 001692911 | (insulin degludec and liraglutide) 3.6mg/mL / 100[iU]/mL SUBCUTANEOUS INJECTION, SOLUTION |
| NDC | 00169291115 | 3 ML insulin degludec 100 UNT/ML / liraglutide 3.6 MG/ML Pen Injector [Xultophy] |
| NDC | 00169291190 | 3 ML insulin degludec 100 UNT/ML / liraglutide 3.6 MG/ML Pen Injector [Xultophy] |
| NDC | 00169291197 | 3 ML insulin degludec 100 UNT/ML / liraglutide 3.6 MG/ML Pen Injector [Xultophy] |
| NDC | 001694060 | liraglutide 6mg/mL SUBCUTANEOUS INJECTION |
| NDC | 00169406012 | 3 ML liraglutide 6 MG/ML Pen Injector [Victoza] |
| NDC | 00169406013 | 3 ML liraglutide 6 MG/ML Pen Injector [Victoza] |
| NDC | 00169406090 | 3 ML liraglutide 6 MG/ML Pen Injector [Victoza] |
| NDC | 00169406097 | 3 ML liraglutide 6 MG/ML Pen Injector [Victoza] |
| NDC | 00169406098 | 3 ML liraglutide 6 MG/ML Pen Injector [Victoza] |
| NDC | 00169406099 | 3 ML liraglutide 6 MG/ML Pen Injector [Victoza] |
| NDC | 500902853 | liraglutide 6mg/mL SUBCUTANEOUS INJECTION |
| NDC | 50090285300 | 3 ML liraglutide 6 MG/ML Pen Injector [Victoza] |
| NDC | 500904257 | liraglutide 6mg/mL SUBCUTANEOUS INJECTION, SOLUTION |
| NDC | 50090425700 | 3 ML liraglutide 6 MG/ML Pen Injector [Saxenda] |

Continued on next page

**Table1 – continued from previous page**

| CodeSystem | ConceptCode | ConceptName |
|---|---|---|
| NDC | 500904503 | liraglutide 6mg/mL SUBCUTANEOUS INJECTION |
| NDC | 50090450300 | 3 ML liraglutide 6 MG/ML Pen Injector [Victoza] |
| RxNorm | 897123 | Victoza |
| RxNorm | 897125 | liraglutide Prefilled Syringe [Victoza] |
| RxNorm | 1186578 | Victoza Injectable Product |
| RxNorm | 1360495 | liraglutide 6 MG/ML Pen Injector [Victoza] |
| RxNorm | 1653597 | liraglutide Pen Injector [Victoza] |
| RxNorm | 897124 | liraglutide 6 MG/ML [Victoza] |
| RxNorm | 897126 | 3 ML liraglutide 6 MG/ML Pen Injector [Victoza] |
| NDC | 54569650700 | 3 ML liraglutide 6 MG/ML Pen Injector [Victoza] |
| NDC | 001694060 | liraglutide 6mg/mL SUBCUTANEOUS INJECTION |
| NDC | 00169406012 | 3 ML liraglutide 6 MG/ML Pen Injector [Victoza] |
| NDC | 00169406013 | 3 ML liraglutide 6 MG/ML Pen Injector [Victoza] |
| NDC | 00169406090 | 3 ML liraglutide 6 MG/ML Pen Injector [Victoza] |
| NDC | 00169406097 | 3 ML liraglutide 6 MG/ML Pen Injector [Victoza] |
| NDC | 00169406098 | 3 ML liraglutide 6 MG/ML Pen Injector [Victoza] |
| NDC | 00169406099 | 3 ML liraglutide 6 MG/ML Pen Injector [Victoza] |
| NDC | 500902853 | liraglutide 6mg/mL SUBCUTANEOUS INJECTION |
| NDC | 50090285300 | 3 ML liraglutide 6 MG/ML Pen Injector [Victoza] |
| NDC | 500904503 | liraglutide 6mg/mL SUBCUTANEOUS INJECTION |
| NDC | 50090450300 | 3 ML liraglutide 6 MG/ML Pen Injector [Victoza] |
| RxNorm | 1598264 | Saxenda |
| RxNorm | 1598266 | liraglutide Prefilled Syringe [Saxenda] |
| RxNorm | 1598265 | liraglutide 6 MG/ML [Saxenda] |
| RxNorm | 1598267 | Saxenda Injectable Product |
| RxNorm | 1598268 | 3 ML liraglutide 6 MG/ML Pen Injector [Saxenda] |
| RxNorm | 1598269 | liraglutide 6 MG/ML Pen Injector [Saxenda] |
| RxNorm | 1653600 | liraglutide Pen Injector [Saxenda] |
| NDC | 001692800 | liraglutide 6mg/mL SUBCUTANEOUS INJECTION, SOLUTION |
| NDC | 00169280013 | 3 ML liraglutide 6 MG/ML Pen Injector [Saxenda] |
| NDC | 00169280015 | 3 ML liraglutide 6 MG/ML Pen Injector [Saxenda] |
| NDC | 00169280090 | 3 ML liraglutide 6 MG/ML Pen Injector [Saxenda] |
| NDC | 00169280097 | 3 ML liraglutide 6 MG/ML Pen Injector [Saxenda] |
| NDC | 500904257 | liraglutide 6mg/mL SUBCUTANEOUS INJECTION, SOLUTION |
| NDC | 50090425700 | 3 ML liraglutide 6 MG/ML Pen Injector [Saxenda] |
| RxNorm | 60548 | exenatide |
| RxNorm | 1242962 | exenatide Injectable Suspension |
| RxNorm | 1544917 | exenatide Prefilled Syringe [Bydureon] |
| RxNorm | 604750 | exenatide 0.250 MG/ML Injectable Solution |
| RxNorm | 604749 | exenatide Injectable Solution |
| RxNorm | 604752 | exenatide 0.250 MG/ML [Byetta] |

Continued on next page

**Table1 – continued from previous page**

| CodeSystem | ConceptCode | ConceptName |
|---|---|---|
| RxNorm | 604748 | exenatide 0.250 MG/ML |
| RxNorm | 604753 | exenatide Injectable Solution [Byetta] |
| RxNorm | 604754 | exenatide 0.250 MG/ML Injectable Solution [Byetta] |
| RxNorm | 847909 | exenatide Prefilled Syringe |
| RxNorm | 847912 | exenatide Prefilled Syringe [Byetta] |
| RxNorm | 1163790 | exenatide Injectable Product |
| RxNorm | 1169415 | Byetta Injectable Product |
| RxNorm | 1242961 | exenatide 3.08 MG/ML |
| RxNorm | 1242963 | exenatide 2 MG Injection |
| RxNorm | 1242964 | Bydureon |
| RxNorm | 1242965 | exenatide 3.08 MG/ML [Bydureon] |
| RxNorm | 1242967 | Bydureon Injectable Product |
| RxNorm | 1242968 | exenatide 2 MG Injection [Bydureon] |
| RxNorm | 1359802 | exenatide 0.005 MG/ACTUAT Pen Injector [Byetta] |
| RxNorm | 1359979 | exenatide 0.01 MG/ACTUAT Pen Injector [Byetta] |
| RxNorm | 1360454 | exenatide 0.01 MG/ACTUAT Pen Injector |
| RxNorm | 1359640 | exenatide 0.005 MG/ACTUAT Pen Injector |
| RxNorm | 1544919 | exenatide 3.08 MG/ML Pen Injector |
| RxNorm | 1544920 | exenatide 3.08 MG/ML Pen Injector [Bydureon] |
| RxNorm | 1544916 | 0.65 ML exenatide 3.08 MG/ML Pen Injector |
| RxNorm | 1544918 | 0.65 ML exenatide 3.08 MG/ML Pen Injector [Bydureon] |
| RxNorm | 1653610 | exenatide 2 MG |
| RxNorm | 1653611 | exenatide Injection |
| RxNorm | 1653613 | exenatide 2 MG [Bydureon] |
| RxNorm | 1653614 | exenatide Injection [Bydureon] |
| RxNorm | 1653616 | exenatide Pen Injector |
| RxNorm | 1653619 | exenatide Pen Injector [Bydureon] |
| RxNorm | 1653625 | exenatide Pen Injector [Byetta] |
| RxNorm | 1990864 | exenatide 2.35 MG/ML |
| RxNorm | 1990865 | exenatide Auto-Injector |
| RxNorm | 1990866 | 0.85 ML exenatide 2.35 MG/ML Auto-Injector |
| RxNorm | 1990867 | exenatide 2.35 MG/ML [Bydureon] |
| RxNorm | 1990868 | exenatide Auto-Injector [Bydureon] |
| RxNorm | 1990869 | 0.85 ML exenatide 2.35 MG/ML Auto-Injector [Bydureon] |
| RxNorm | 1990870 | exenatide 2.35 MG/ML Auto-Injector |
| RxNorm | 1990871 | exenatide 2.35 MG/ML Auto-Injector [Bydureon] |
| RxNorm | 847908 | exenatide 0.01 MG/ACTUAT |
| RxNorm | 847910 | 60 ACTUAT exenatide 0.01 MG/ACTUAT Pen Injector |
| RxNorm | 847911 | exenatide 0.01 MG/ACTUAT [Byetta] |
| RxNorm | 847913 | 60 ACTUAT exenatide 0.01 MG/ACTUAT Pen Injector [Byetta] |
| RxNorm | 847914 | exenatide 0.005 MG/ACTUAT |
| RxNorm | 847915 | 60 ACTUAT exenatide 0.005 MG/ACTUAT Pen Injector |

Continued on next page

**Table1 – continued from previous page**

| CodeSystem | ConceptCode | ConceptName |
|---|---|---|
| RxNorm | 847916 | exenatide 0.005 MG/ACTUAT [Byetta] |
| RxNorm | 847917 | 60 ACTUAT exenatide 0.005 MG/ACTUAT Pen Injector [Byetta] |
| RxNorm | 744863 | exenatide 0.25 MG/ML Injectable Solution |
| RxNorm | 744862 | exenatide 0.25 MG/ML |
| RxNorm | 744864 | exenatide 0.25 MG/ML [Byetta] |
| RxNorm | 744865 | exenatide 0.25 MG/ML Injectable Solution [Byetta] |
| RxNorm | 1242966 | exenatide Injectable Suspension [Bydureon] |
| RxNorm | 604751 | Byetta |
| SNOMED CT | 416525003 | Exenatide 250mcg/mL injection solution prefilled pen |
| SNOMED CT | 416859008 | Exenatide |
| SNOMED CT | 417734003 | Exenatide-containing product |
| SNOMED CT | 438958002 | Exenatide 250micrograms/mL injection solution 2.4mL pre-filled pen |
| SNOMED CT | 440246006 | Exenatide 250micrograms/mL injection solution 1.2mL pre-filled pen |
| SNOMED CT | 775913009 | Exenatide only product |
| SNOMED CT | 1155637009 | Exenatide 250 microgram/mL solution for injection |
| SNOMED CT | 1155638004 | Exenatide-containing product in parenteral dose form |
| SNOMED CT | 1155639007 | Exenatide only product in parenteral dose form |
| SNOMED CT | 1237218000 | Exenatide 2 mg powder for prolonged-release suspension for injection vial |
| NDC | 003106512 | exenatide 250ug/mL SUBCUTANEOUS INJECTION |
| NDC | 00310651201 | 60 ACTUAT exenatide 0.005 MG/ACTUAT Pen Injector [Byetta] |
| NDC | 00310651285 | 60 ACTUAT exenatide 0.005 MG/ACTUAT Pen Injector [Byetta] |
| NDC | 003106520 | exenatide KIT |
| NDC | 00310652004 | exenatide 2 MG Injection [Bydureon] |
| NDC | 003106524 | exenatide 250ug/mL SUBCUTANEOUS INJECTION |
| NDC | 00310652401 | 60 ACTUAT exenatide 0.01 MG/ACTUAT Pen Injector [Byetta] |
| NDC | 003106530 | exenatide 2mg/.65mL SUBCUTANEOUS INJECTION, SUSPENSION, EXTENDED RELEASE |
| NDC | 00310653001 | 0.65 ML exenatide 3.08 MG/ML Pen Injector [Bydureon] |
| NDC | 00310653004 | 0.65 ML exenatide 3.08 MG/ML Pen Injector [Bydureon] |
| NDC | 00310653085 | 0.65 ML exenatide 3.08 MG/ML Pen Injector [Bydureon] |
| NDC | 003106540 | exenatide 2mg/.85mL SUBCUTANEOUS INJECTION, SUSPENSION, EXTENDED RELEASE |
| NDC | 00310654001 | 0.85 ML exenatide 2.35 MG/ML Auto-Injector [Bydureon] |
| NDC | 00310654004 | 0.85 ML exenatide 2.35 MG/ML Auto-Injector [Bydureon] |
| NDC | 00310654085 | 0.85 ML exenatide 2.35 MG/ML Auto-Injector [Bydureon] |
| NDC | 548685384 | exenatide 250ug/mL SUBCUTANEOUS INJECTION |

Continued on next page

**Table1 – continued from previous page**

| CodeSystem | ConceptCode | ConceptName |
|---|---|---|
| NDC | 54868538400 | 60 ACTUAT exenatide 0.005 MG/ACTUAT Pen Injector [Byetta] |
| NDC | 54868538401 | 60 ACTUAT exenatide 0.01 MG/ACTUAT Pen Injector [Byetta] |
| NDC | 54868538402 | 60 ACTUAT exenatide 0.005 MG/ACTUAT Prefilled Syringe [Byetta] |
| NDC | 68258894701 | 60 ACTUAT exenatide 0.005 MG/ACTUAT Prefilled Syringe [Byetta] |
| NDC | 68258894802 | 60 ACTUAT exenatide 0.01 MG/ACTUAT Prefilled Syringe [Byetta] |
| NDC | 00002021007 | 60 ACTUAT exenatide 0.005 MG/ACTUAT Prefilled Syringe [Byetta] |
| NDC | 00002021008 | 60 ACTUAT exenatide 0.01 MG/ACTUAT Prefilled Syringe [Byetta] |
| NDC | 00002021009 | 60 ACTUAT exenatide 0.005 MG/ACTUAT Prefilled Syringe [Byetta] |
| NDC | 66029021007 | 60 ACTUAT exenatide 0.005 MG/ACTUAT Prefilled Syringe [Byetta] |
| NDC | 66029021008 | 60 ACTUAT exenatide 0.01 MG/ACTUAT Prefilled Syringe [Byetta] |
| NDC | 667800210 | exenatide 250ug/mL SUBCUTANEOUS INJECTION [byetta] |
| NDC | 66780021007 | 60 ACTUAT exenatide 0.005 MG/ACTUAT Pen Injector [Byetta] |
| NDC | 66780021008 | 60 ACTUAT exenatide 0.01 MG/ACTUAT Prefilled Syringe [Byetta] |
| NDC | 66780021009 | 60 ACTUAT exenatide 0.005 MG/ACTUAT Pen Injector [Byetta] |
| NDC | 667800212 | exenatide 250ug/mL SUBCUTANEOUS INJECTION [byetta] |
| NDC | 66780021201 | 60 ACTUAT exenatide 0.01 MG/ACTUAT Pen Injector [Byetta] |
| NDC | 667800219 | exenatide KIT [bydureon] |
| NDC | 66780021902 | exenatide 2 MG Injection [Bydureon] |
| NDC | 66780021904 | exenatide 2 MG Injection [Bydureon] |
| NDC | 667800226 | exenatide KIT [bydureon] |
| NDC | 66780022601 | exenatide 2 MG Injection [Bydureon] |
| NDC | 66914103504 | 60 ACTUAT exenatide 0.005 MG/ACTUAT Prefilled Syringe [Byetta] |
| NDC | 66914103505 | 60 ACTUAT exenatide 0.01 MG/ACTUAT Prefilled Syringe [Byetta] |
| RxNorm | 1242964 | Bydureon |
| RxNorm | 1544917 | exenatide Prefilled Syringe [Bydureon] |

Continued on next page

**Table1 – continued from previous page**

| CodeSystem | ConceptCode | ConceptName |
|---|---|---|
| RxNorm | 1242965 | exenatide 3.08 MG/ML [Bydureon] |
| RxNorm | 1242967 | Bydureon Injectable Product |
| RxNorm | 1242968 | exenatide 2 MG Injection [Bydureon] |
| RxNorm | 1544920 | exenatide 3.08 MG/ML Pen Injector [Bydureon] |
| RxNorm | 1544918 | 0.65 ML exenatide 3.08 MG/ML Pen Injector [Bydureon] |
| RxNorm | 1653613 | exenatide 2 MG [Bydureon] |
| RxNorm | 1653614 | exenatide Injection [Bydureon] |
| RxNorm | 1653619 | exenatide Pen Injector [Bydureon] |
| RxNorm | 1990867 | exenatide 2.35 MG/ML [Bydureon] |
| RxNorm | 1990868 | exenatide Auto-Injector [Bydureon] |
| RxNorm | 1990869 | 0.85 ML exenatide 2.35 MG/ML Auto-Injector [Bydureon] |
| RxNorm | 1990871 | exenatide 2.35 MG/ML Auto-Injector [Bydureon] |
| RxNorm | 1242966 | exenatide Injectable Suspension [Bydureon] |
| NDC | 003106520 | exenatide KIT |
| NDC | 00310652004 | exenatide 2 MG Injection [Bydureon] |
| NDC | 003106530 | exenatide 2mg/.65mL SUBCUTANEOUS INJECTION, SUSPENSION, EXTENDED RELEASE |
| NDC | 00310653001 | 0.65 ML exenatide 3.08 MG/ML Pen Injector [Bydureon] |
| NDC | 00310653004 | 0.65 ML exenatide 3.08 MG/ML Pen Injector [Bydureon] |
| NDC | 00310653085 | 0.65 ML exenatide 3.08 MG/ML Pen Injector [Bydureon] |
| NDC | 003106540 | exenatide 2mg/.85mL SUBCUTANEOUS INJECTION, SUSPENSION, EXTENDED RELEASE |
| NDC | 00310654001 | 0.85 ML exenatide 2.35 MG/ML Auto-Injector [Bydureon] |
| NDC | 00310654004 | 0.85 ML exenatide 2.35 MG/ML Auto-Injector [Bydureon] |
| NDC | 00310654085 | 0.85 ML exenatide 2.35 MG/ML Auto-Injector [Bydureon] |
| NDC | 667800219 | exenatide KIT [bydureon] |
| NDC | 66780021902 | exenatide 2 MG Injection [Bydureon] |
| NDC | 66780021904 | exenatide 2 MG Injection [Bydureon] |
| NDC | 667800226 | exenatide KIT [bydureon] |
| NDC | 66780022601 | exenatide 2 MG Injection [Bydureon] |
| RxNorm | 604751 | Byetta |
| RxNorm | 604753 | exenatide Injectable Solution [Byetta] |
| RxNorm | 604754 | exenatide 0.250 MG/ML Injectable Solution [Byetta] |
| RxNorm | 847912 | exenatide Prefilled Syringe [Byetta] |
| RxNorm | 1169415 | Byetta Injectable Product |
| RxNorm | 1359802 | exenatide 0.005 MG/ACTUAT Pen Injector [Byetta] |
| RxNorm | 1359979 | exenatide 0.01 MG/ACTUAT Pen Injector [Byetta] |
| RxNorm | 1653625 | exenatide Pen Injector [Byetta] |
| RxNorm | 847911 | exenatide 0.01 MG/ACTUAT [Byetta] |
| RxNorm | 847913 | 60 ACTUAT exenatide 0.01 MG/ACTUAT Pen Injector [Byetta] |
| RxNorm | 847916 | exenatide 0.005 MG/ACTUAT [Byetta] |

Continued on next page

**Table1 – continued from previous page**

| CodeSystem | ConceptCode | ConceptName |
|---|---|---|
| RxNorm | 847917 | 60 ACTUAT exenatide 0.005 MG/ACTUAT Pen Injector [Byetta] |
| RxNorm | 744864 | exenatide 0.25 MG/ML [Byetta] |
| RxNorm | 744865 | exenatide 0.25 MG/ML Injectable Solution [Byetta] |
| NDC | 003106512 | exenatide 250ug/mL SUBCUTANEOUS INJECTION |
| NDC | 00310651201 | 60 ACTUAT exenatide 0.005 MG/ACTUAT Pen Injector [Byetta] |
| NDC | 00310651285 | 60 ACTUAT exenatide 0.005 MG/ACTUAT Pen Injector [Byetta] |
| NDC | 003106524 | exenatide 250ug/mL SUBCUTANEOUS INJECTION |
| NDC | 00310652401 | 60 ACTUAT exenatide 0.01 MG/ACTUAT Pen Injector [Byetta] |
| NDC | 548685384 | exenatide 250ug/mL SUBCUTANEOUS INJECTION |
| NDC | 54868538400 | 60 ACTUAT exenatide 0.005 MG/ACTUAT Pen Injector [Byetta] |
| NDC | 54868538401 | 60 ACTUAT exenatide 0.01 MG/ACTUAT Pen Injector [Byetta] |
| NDC | 54868538402 | 60 ACTUAT exenatide 0.005 MG/ACTUAT Prefilled Syringe [Byetta] |
| NDC | 68258894701 | 60 ACTUAT exenatide 0.005 MG/ACTUAT Prefilled Syringe [Byetta] |
| NDC | 68258894802 | 60 ACTUAT exenatide 0.01 MG/ACTUAT Prefilled Syringe [Byetta] |
| NDC | 00002021007 | 60 ACTUAT exenatide 0.005 MG/ACTUAT Prefilled Syringe [Byetta] |
| NDC | 00002021008 | 60 ACTUAT exenatide 0.01 MG/ACTUAT Prefilled Syringe [Byetta] |
| NDC | 00002021009 | 60 ACTUAT exenatide 0.005 MG/ACTUAT Prefilled Syringe [Byetta] |
| NDC | 66029021007 | 60 ACTUAT exenatide 0.005 MG/ACTUAT Prefilled Syringe [Byetta] |
| NDC | 66029021008 | 60 ACTUAT exenatide 0.01 MG/ACTUAT Prefilled Syringe [Byetta] |
| NDC | 667800210 | exenatide 250ug/mL SUBCUTANEOUS INJECTION [byetta] |
| NDC | 66780021007 | 60 ACTUAT exenatide 0.005 MG/ACTUAT Pen Injector [Byetta] |
| NDC | 66780021008 | 60 ACTUAT exenatide 0.01 MG/ACTUAT Prefilled Syringe [Byetta] |
| NDC | 66780021009 | 60 ACTUAT exenatide 0.005 MG/ACTUAT Pen Injector [Byetta] |
| NDC | 667800212 | exenatide 250ug/mL SUBCUTANEOUS INJECTION [byetta] |

Continued on next page

**Table1 – continued from previous page**

| CodeSystem | ConceptCode | ConceptName |
|---|---|---|
| NDC | 66780021201 | 60 ACTUAT exenatide 0.01 MG/ACTUAT Pen Injector [Byetta] |
| NDC | 66914103504 | 60 ACTUAT exenatide 0.005 MG/ACTUAT Prefilled Syringe [Byetta] |
| NDC | 66914103505 | 60 ACTUAT exenatide 0.01 MG/ACTUAT Prefilled Syringe [Byetta] |

#### 0.2 Obesity

Table 2: Concept codes used to identify obesity conditions.

| CodeSystem | ConceptCode | ConceptName |
|---|---|---|
| ICD10CM | E66.01 | Morbid (severe) obesity due to excess calories |
| ICD10CM | E66.09 | Other obesity due to excess calories |
| ICD10CM | E66.1 | Drug-induced obesity |
| ICD10CM | E66.2 | Morbid (severe) obesity with alveolar hypoventilation |
| ICD10CM | E66.3 | Overweight |
| ICD10CM | E66.8 | Other obesity |
| ICD10CM | E66.9 | Obesity, unspecified |
| ICD10CM | Z68.30 | Body mass index (BMI) 30.0-30.9, adult |
| ICD10CM | Z68.31 | Body mass index (BMI) 31.0-31.9, adult |
| ICD10CM | Z68.32 | Body mass index (BMI) 32.0-32.9, adult |
| ICD10CM | Z68.33 | Body mass index (BMI) 33.0-33.9, adult |
| ICD10CM | Z68.34 | Body mass index (BMI) 34.0-34.9, adult |
| ICD10CM | Z68.35 | Body mass index (BMI) 35.0-35.9, adult |
| ICD10CM | Z68.36 | Body mass index (BMI) 36.0-36.9, adult |
| ICD10CM | Z68.37 | Body mass index (BMI) 37.0-37.9, adult |
| ICD10CM | Z68.38 | Body mass index (BMI) 38.0-38.9, adult |
| ICD10CM | Z68.39 | Body mass index (BMI) 39.0-39.9, adult |
| ICD10CM | Z68.41 | Body mass index (BMI) 40.0-44.9, adult |
| ICD10CM | Z68.42 | Body mass index (BMI) 45.0-49.9, adult |
| ICD10CM | Z68.43 | Body mass index (BMI) 50.0-59.9, adult |
| ICD10CM | Z68.44 | Body mass index (BMI) 60.0-69.9, adult |
| ICD10CM | Z68.45 | Body mass index (BMI) 70 or greater, adult |
| SNOMED CT | 83911000119104 | Severe obesity |
| SNOMED CT | 190965006 | Drug-induced obesity |
| SNOMED CT | 190966007 | Extreme obesity with alveolar hypoventilation |
| SNOMED CT | 162864005 | Body mass index 30+ - obesity |
| SNOMED CT | 238131007 | Overweight |
| SNOMED CT | 238136002 | Morbid obesity |
| SNOMED CT | 408512008 | Body mass index 40+ - severely obese |

Continued on next page

**Table2 – continued from previous page**

| CodeSystem | ConceptCode | ConceptName |
|---|---|---|
| SNOMED CT | 414915002 | Obese |
| SNOMED CT | 414916001 | Obesity |
| SNOMED CT | 415530009 | Simple obesity |
| SNOMED CT | 1187531009 | Obesity due to pituitary disease |
| SNOMED CT | 5036006 | Hypogonadal obesity |
| SNOMED CT | 1229946007 | MAGEL2-related Prader-Willi-like syndrome |
| SNOMED CT | 1208987006 | PHIP-related behavioral problems, intellectual disability, obesity, dysmorphic features syndrome |
| SNOMED CT | 1229943004 | SIM1-related Prader-Willi-like syndrome |
| SNOMED CT | 763350002 | Intellectual disability, obesity, brain malformation, facial dysmorphism syndrome |
| SNOMED CT | 770680004 | Prader-Willi-like syndrome |
| SNOMED CT | 770750002 | Intellectual disability, seizures, macrocephaly, obesity syndrome |
| SNOMED CT | 773663004 | Rapid-onset childhood obesity, hypothalamic dysfunction, hypoventilation, autonomic dysregulation syndrome |
| SNOMED CT | 774102003 | Intellectual disability, obesity, prognathism, eye and skin anomalies syndrome |
| SNOMED CT | 776204008 | Colobomatous microphthalmia, obesity, hypogenitalism, intellectual disability syndrome |
| SNOMED CT | 783549006 | Obesity due to CEP19 deficiency |
| SNOMED CT | 783556000 | Severe early-onset obesity insulin resistance syndrome due to SH2B1 deficiency |
| SNOMED CT | 783719006 | Obesity due to SIM1 deficiency |
| SNOMED CT | 785722006 | Obesity due to leptin receptor gene deficiency |
| SNOMED CT | 788996008 | Obesity in adolescence |
| SNOMED CT | 819948005 | Obese class III |
| SNOMED CT | 171000119107 | Maternal obesity complicating pregnancy, childbirth and the puerperium, antepartum |
| SNOMED CT | 443371000124107 | Obese class I |
| SNOMED CT | 443381000124105 | Obese class II |
| SNOMED CT | 461341000124106 | Lower body obesity |
| SNOMED CT | 1076701000119104 | Hypertrophy of fat pad of right knee |
| SNOMED CT | 1076711000119101 | Hypertrophy of fat pad of left knee |
| SNOMED CT | 10750551000119100 | Obesity in mother complicating childbirth |
| SNOMED CT | 15750121000119108 | Severe obesity complicating pregnancy |
| SNOMED CT | 722037004 | Intellectual disability, epileptic seizures, hypogonadism and hypogenitalism, microcephaly, obesity syndrome |
| SNOMED CT | 162690006 | O/E - obese |
| SNOMED CT | 162863004 | Body mass index 25-29 - overweight |
| SNOMED CT | 238132000 | Android obesity |
| SNOMED CT | 238133005 | Gynecoid obesity |
| SNOMED CT | 238134004 | Generalized obesity |

Continued on next page

**Table2 – continued from previous page**

| CodeSystem | ConceptCode | ConceptName |
|---|---|---|
| SNOMED CT | 238135003 | Fat pad syndrome |
| SNOMED CT | 248311001 | Central obesity |
| SNOMED CT | 248312008 | Peripheral obesity |
| SNOMED CT | 360566006 | Buffalo obesity |
| SNOMED CT | 414438005 | Hyperplastic obesity |
| SNOMED CT | 414917005 | Obesity by adipocyte growth pattern |
| SNOMED CT | 414918000 | Obesity by age of onset |
| SNOMED CT | 414919008 | Obesity by contributing factors |
| SNOMED CT | 414920002 | Obesity by fat distribution pattern |
| SNOMED CT | 44772007 | Maternal obesity syndrome |
| SNOMED CT | 53146006 | Hypothyroid obesity |
| SNOMED CT | 57337005 | Steatopygia |
| SNOMED CT | 62999006 | Adiposogenital dystrophy |
| SNOMED CT | 63702009 | Alstrom syndrome |
| SNOMED CT | 72894001 | Hypertrophy of fat pad of knee |
| SNOMED CT | 80660001 | Mauriac's syndrome |
| SNOMED CT | 444862003 | Childhood obesity |
| SNOMED CT | 450451007 | Overweight in childhood |
| SNOMED CT | 700150001 | Congenital leptin deficiency |
| SNOMED CT | 702949005 | Proopiomelanocortin deficiency syndrome |
| SNOMED CT | 715628009 | MORM syndrome |
| SNOMED CT | 717269008 | Obesity due to melanocortin 4 receptor deficiency |
| SNOMED CT | 717761005 | Choroideremia with deafness and obesity syndrome |
| SNOMED CT | 719160009 | Syndromic X-linked intellectual disability type 7 |
| SNOMED CT | 719834005 | Wilson Turner syndrome |
| SNOMED CT | 721231007 | Hydrocephalus with obesity and hypogonadism syndrome |
| SNOMED CT | 722051004 | Obesity, colitis, hypothyroidism, cardiac hypertrophy, developmental delay syndrome |
| SNOMED CT | 722053001 | Obesity due to prohormone convertase I deficiency |
| SNOMED CT | 722595002 | Overweight in adulthood with body mass index of 25 or more but less than 30 |
| SNOMED CT | 722596001 | Obesity caused by energy imbalance |
| SNOMED CT | 724137002 | MOMO syndrome |
| SNOMED CT | 268915006 | O/E - weight 10-20% over ideal |
| SNOMED CT | 268916007 | O/E - weight greater than 20% over ideal |
| SNOMED CT | 270486005 | Localized adiposity |
| SNOMED CT | 275947003 | O/E - overweight |
| SNOMED CT | 290439001 | Familial obesity |
| SNOMED CT | 292464007 | Constitutional obesity |
| SNOMED CT | 293481008 | Hyperplastic-hypertrophic obesity |
| SNOMED CT | 294493008 | Lifelong obesity |
| SNOMED CT | 295509007 | Hypertrophic obesity |
| SNOMED CT | 296526005 | Adult-onset obesity |

Continued on next page

**Table2 – continued from previous page**

| CodeSystem | ConceptCode | ConceptName |
|---|---|---|
| SNOMED CT | 297500005 | Endogenous obesity |
| SNOMED CT | 298464002 | Obesity of endocrine origin |
| SNOMED CT | 1003380001 | 6q16 microdeletion syndrome |
| SNOMED CT | 1255335006 | X-linked intellectual disability, short stature, overweight syndrome |
| SNOMED CT | 82793005 | Hypothalamic obesity |
| SNOMED CT | 111036000 | Hyperinsulinar obesity |
| SNOMED CT | 1260134001 | Spastic paraplegia, intellectual disability, nystagmus, obesity syndrome |
| SNOMED CT | 1260139006 | Genetic non-syndromic obesity |

#### 0.3 Overweight

Table 3: Concept codes used to identify overweight conditions.

| CodeSystem | ConceptCode | ConceptName |
|---|---|---|
| ICD10CM | E66.3 | Overweight |
| ICD10CM | Z68.27 | Body mass index (BMI) 27.0-27.9, adult |
| ICD10CM | Z68.28 | Body mass index (BMI) 28.0-28.9, adult |
| ICD10CM | Z68.29 | Body mass index (BMI) 29.0-29.9, adult |
| SNOMED CT | 238131007 | Overweight |
| SNOMED CT | 162863004 | Body mass index 25-29 - overweight |
| SNOMED CT | 722595002 | Overweight in adulthood with body mass index of 25 or more but less than 30 |
| SNOMED CT | 450451007 | Overweight in childhood |
| SNOMED CT | 268915006 | O/E - weight 10-20% over ideal |
| SNOMED CT | 268916007 | O/E - weight greater than 20% over ideal |
| SNOMED CT | 275947003 | O/E - overweight |

#### 0.4 Comborbidities

##### 0.4.1 Asthma

Table 4: Concept codes used to identify asthma conditions.

| CodeSystem | ConceptCode | ConceptName |
|---|---|---|
| ICD10CM | J45 | Asthma |
| ICD10CM | J45.2 | Mild intermittent asthma |
| ICD10CM | J45.20 | Mild intermittent asthma, uncomplicated |

Continued on next page

**Table4 – continued from previous page**

| CodeSystem | ConceptCode | ConceptName |
|---|---|---|
| ICD10CM | J45.21 | Mild intermittent asthma with (acute) exacerbation |
| ICD10CM | J45.22 | Mild intermittent asthma with status asthmaticus |
| ICD10CM | J45.3 | Mild persistent asthma |
| ICD10CM | J45.30 | Mild persistent asthma, uncomplicated |
| ICD10CM | J45.31 | Mild persistent asthma with (acute) exacerbation |
| ICD10CM | J45.32 | Mild persistent asthma with status asthmaticus |
| ICD10CM | J45.4 | Moderate persistent asthma |
| ICD10CM | J45.40 | Moderate persistent asthma, uncomplicated |
| ICD10CM | J45.41 | Moderate persistent asthma with (acute) exacerbation |
| ICD10CM | J45.42 | Moderate persistent asthma with status asthmaticus |
| ICD10CM | J45.5 | Severe persistent asthma |
| ICD10CM | J45.50 | Severe persistent asthma, uncomplicated |
| ICD10CM | J45.51 | Severe persistent asthma with (acute) exacerbation |
| ICD10CM | J45.52 | Severe persistent asthma with status asthmaticus |
| ICD10CM | J45.9 | Other and unspecified asthma |
| ICD10CM | J45.90 | Unspecified asthma |
| ICD10CM | J45.901 | Unspecified asthma with (acute) exacerbation |
| ICD10CM | J45.902 | Unspecified asthma with status asthmaticus |
| ICD10CM | J45.909 | Unspecified asthma, uncomplicated |
| ICD10CM | J45.99 | Other asthma |
| ICD10CM | J45.990 | Exercise induced bronchospasm |
| ICD10CM | J45.991 | Cough variant asthma |
| ICD10CM | J45.998 | Other asthma |
| SNOMED CT | 1751000119100 | Acute exacerbation of chronic obstructive airways disease with asthma |
| SNOMED CT | 99031000119107 | Acute exacerbation of asthma co-occurrent with allergic rhinitis |
| SNOMED CT | 124991000119109 | Severe persistent asthma co-occurrent with allergic rhinitis |
| SNOMED CT | 125001000119103 | Moderate persistent asthma co-occurrent with allergic rhinitis |
| SNOMED CT | 10674711000119105 | Acute severe exacerbation of asthma co-occurrent with allergic rhinitis |
| SNOMED CT | 10675431000119106 | Severe persistent allergic asthma |
| SNOMED CT | 10675471000119109 | Acute severe exacerbation of severe persistent allergic asthma |
| SNOMED CT | 10675551000119104 | Acute severe exacerbation of severe persistent asthma co-occurrent with allergic rhinitis |
| SNOMED CT | 10675911000119109 | Acute severe exacerbation of mild persistent allergic asthma |
| SNOMED CT | 10675991000119100 | Acute severe exacerbation of mild persistent allergic asthma co-occurrent with allergic rhinitis |
| SNOMED CT | 10676391000119108 | Moderate persistent allergic asthma |
| SNOMED CT | 10676431000119103 | Acute severe exacerbation of moderate persistent allergic asthma |

Continued on next page

**Table4 – continued from previous page**

| CodeSystem | ConceptCode | ConceptName |
|---|---|---|
| SNOMED CT | 10676511000119109 | Acute severe exacerbation of moderate persistent asthma co-occurrent with allergic rhinitis |
| SNOMED CT | 10692721000119102 | Chronic obstructive asthma co-occurrent with acute exacerbation of asthma |
| SNOMED CT | 370219009 | Moderate asthma |
| SNOMED CT | 370221004 | Severe asthma |
| SNOMED CT | 426656000 | Severe persistent asthma |
| SNOMED CT | 427295004 | Moderate persistent asthma |
| SNOMED CT | 442025000 | Acute exacerbation of chronic asthmatic bronchitis |
| SNOMED CT | 707446004 | Exacerbation of moderate persistent asthma |
| SNOMED CT | 707447008 | Exacerbation of severe persistent asthma |
| SNOMED CT | 707512002 | Uncomplicated moderate persistent asthma |
| SNOMED CT | 707513007 | Uncomplicated severe persistent asthma |
| SNOMED CT | 707979007 | Acute severe exacerbation of severe persistent asthma |
| SNOMED CT | 707980005 | Acute severe exacerbation of moderate persistent asthma |
| SNOMED CT | 707981009 | Acute severe exacerbation of mild persistent asthma |
| SNOMED CT | 708090002 | Acute severe exacerbation of asthma |
| SNOMED CT | 708093000 | Acute exacerbation of allergic asthma |
| SNOMED CT | 708094006 | Acute exacerbation of intrinsic asthma |
| SNOMED CT | 708095007 | Acute severe exacerbation of immunoglobin E-mediated allergic asthma |
| SNOMED CT | 708096008 | Acute severe exacerbation of intrinsic asthma |
| SNOMED CT | 733858005 | Acute severe refractory exacerbation of asthma |
| SNOMED CT | 734904007 | Life threatening acute exacerbation of asthma |
| SNOMED CT | 782513000 | Acute severe exacerbation of allergic asthma |
| SNOMED CT | 786836003 | Near fatal asthma |
| SNOMED CT | 829976001 | Thunderstorm asthma |
| SNOMED CT | 135171000119106 | Acute exacerbation of moderate persistent asthma |
| SNOMED CT | 10675391000119101 | Severe controlled persistent asthma |
| SNOMED CT | 10675751000119107 | Severe uncontrolled persistent asthma |
| SNOMED CT | 10676271000119104 | Acute exacerbation of moderate persistent allergic asthma |

##### 0.4.2 Atrial Fibrillation

Table 5: Concept codes used to identify atrial fibrillation flutter conditions.

| CodeSystem | ConceptCode | ConceptName |
|---|---|---|
| ICD10CM | I48.0 | Paroxysmal atrial fibrillation |
| ICD10CM | I48.11 | Longstanding persistent atrial fibrillation |
| ICD10CM | I48.19 | Other persistent atrial fibrillation |
| ICD10CM | I48.20 | Chronic atrial fibrillation, unspecified |

Continued on next page

**Table5 – continued from previous page**

| CodeSystem | ConceptCode | ConceptName |
|---|---|---|
| ICD10CM | I48.21 | Permanent atrial fibrillation |
| ICD10CM | I48.3 | Typical atrial flutter |
| ICD10CM | I48.4 | Atypical atrial flutter |
| ICD10CM | I48.91 | Unspecified atrial fibrillation |
| ICD10CM | I48.92 | Unspecified atrial flutter |
| ICD9CM | 427.31 | Atrial fibrillation |
| ICD9CM | 427.32 | Atrial flutter |
| SNOMED CT | 5370000 | Atrial flutter |
| SNOMED CT | 120041000119109 | Atrial fibrillation with rapid ventricular response |
| SNOMED CT | 15964901000119107 | Atypical atrial flutter |
| SNOMED CT | 195080001 | Atrial fibrillation and flutter |
| SNOMED CT | 233910005 | Lone atrial fibrillation |
| SNOMED CT | 233911009 | Non-rheumatic atrial fibrillation |
| SNOMED CT | 314208002 | Rapid atrial fibrillation |
| SNOMED CT | 425615007 | Chronic atrial flutter |
| SNOMED CT | 426749004 | Chronic atrial fibrillation |
| SNOMED CT | 426814001 | Transient cerebral ischemia due to atrial fibrillation |
| SNOMED CT | 49436004 | Atrial fibrillation |
| SNOMED CT | 427665004 | Paroxysmal atrial flutter |
| SNOMED CT | 440028005 | Permanent atrial fibrillation |
| SNOMED CT | 440059007 | Persistent atrial fibrillation |
| SNOMED CT | 706923002 | Longstanding persistent atrial fibrillation |
| SNOMED CT | 715395008 | Familial atrial fibrillation |
| SNOMED CT | 720448006 | Typical atrial flutter |
| SNOMED CT | 282825002 | Paroxysmal atrial fibrillation |
| SNOMED CT | 300996004 | Controlled atrial fibrillation |
| SNOMED CT | 762247006 | Preexcited atrial fibrillation |
| SNOMED CT | 280797561000119107 | Permanent atrial fibrillation due to heart valve disorder |
| SNOMED CT | 313377641000119105 | Atrial fibrillation due to heart valve disorder |
| SNOMED CT | 467643831000119105 | Paroxysmal atrial fibrillation due to heart valve disorder |
| SNOMED CT | 489609371000119104 | Persistent atrial fibrillation due to heart valve disorder |
| SNOMED CT | 715560009 | Idiopathic neonatal atrial flutter |
| SNOMED CT | 1010405004 | Paroxysmal atrial fibrillation with rapid ventricular response |

##### 0.4.3 Bariatric Surgery

Table 6: Concept codes used to identify bariatric surgery.

| CodeSystem | ConceptCode | ConceptName |
|---|---|---|
| ICD10CM | Z98.84 | Bariatric surgery status |

Continued on next page

**Table6 – continued from previous page**

| CodeSystem | ConceptCode | ConceptName |
|---|---|---|
| SNOMED CT | 329281000119107 | History of sleeve gastrectomy |
| SNOMED CT | 427074001 | Laparoscopic sleeve gastrectomy |
| SNOMED CT | 430715008 | Bariatric operative procedure |
| SNOMED CT | 442338001 | Bypass of stomach |
| SNOMED CT | 608848006 | History of bariatric surgical procedure |
| SNOMED CT | 18692006 | Bypass gastroenterostomy |
| SNOMED CT | 30803004 | Printen and Mason operation, high gastric bypass |
| SNOMED CT | 1264335004 | Endoscopic ultrasonography guided gastrojejunostomy and insertion of stent |
| SNOMED CT | 1172524007 | Primary obesity surgery endoluminal 2 |
| SNOMED CT | 173587003 | Bypass of esophagus using stomach |
| SNOMED CT | 173748000 | Bypass of stomach by anastomosis of stomach to transposed jejunum |
| SNOMED CT | 173750008 | Conversion to anastomosis of stomach to transposed jejunum |
| SNOMED CT | 173751007 | Conversion from previous anastomosis of stomach to transposed jejunum |
| SNOMED CT | 173857001 | Bypass of duodenum by anastomosis of stomach to jejunum |
| SNOMED CT | 235281005 | Anterior gastrojejunostomy |
| SNOMED CT | 235282003 | Anastomosis of stomach to ileum |
| SNOMED CT | 870378000 | Endoscopic sleeve gastroplasty |
| SNOMED CT | 173747005 | Connection of stomach to transposed jejunum |
| SNOMED CT | 433024005 | Gastrojejunostomy using fluoroscopic guidance |
| SNOMED CT | 443906008 | Bypass of stomach with short limb Roux-en-Y gastroenterostomy |
| SNOMED CT | 708629005 | Laparoscopic bypass gastrojejunostomy |
| SNOMED CT | 708983005 | Laparoscopic bypass of stomach |
| SNOMED CT | 314593003 | Posterior gastrojejunostomy |
| SNOMED CT | 358575006 | Gastroduodenostomy |
| SNOMED CT | 359529008 | Jaboulay operation, gastroduodenostomy |
| SNOMED CT | 265365007 | Bypass of stomach by anastomosis of stomach to duodenum |
| SNOMED CT | 287834004 | Gastroenterostomy - no gastrectomy |
| SNOMED CT | 45753003 | Cholecystenterostomy with gastroenterostomy |
| SNOMED CT | 49245001 | Bypass gastrojejunostomy |
| SNOMED CT | 57245002 | Pylorostomy |
| ICD9CM | V45.86 | Bariatric surgery status |

###### 0.4.4 Chronic Kidney Disease

Table 7: Concept codes used to identify chronic kidney disease conditions.

| CodeSystem | ConceptCode | ConceptName |
|---|---|---|
| ICD10CM | D63.1 | Anemia in chronic kidney disease |
| ICD10CM | E08.22 | Diabetes mellitus due to underlying condition with diabetic chronic kidney disease |
| ICD10CM | E09.22 | Drug or chemical induced diabetes mellitus with diabetic chronic kidney disease |
| ICD10CM | E13.22 | Other specified diabetes mellitus with diabetic chronic kidney disease |
| ICD10CM | I12 | Hypertensive chronic kidney disease |
| ICD10CM | I12.0 | Hypertensive chronic kidney disease with stage 5 chronic kidney disease or end stage renal disease |
| ICD10CM | I12.9 | Hypertensive chronic kidney disease with stage 1 through stage 4 chronic kidney disease, or unspecified chronic kidney disease |
| ICD10CM | I13 | Hypertensive heart and chronic kidney disease |
| ICD10CM | I13.0 | Hypertensive heart and chronic kidney disease with heart failure and stage 1 through stage 4 chronic kidney disease, or unspecified chronic kidney disease |
| ICD10CM | I13.1 | Hypertensive heart and chronic kidney disease without heart failure |
| ICD10CM | I13.10 | Hypertensive heart and chronic kidney disease without heart failure, with stage 1 through stage 4 chronic kidney disease, or unspecified chronic kidney disease |
| ICD10CM | I13.11 | Hypertensive heart and chronic kidney disease without heart failure, with stage 5 chronic kidney disease, or end stage renal disease |
| ICD10CM | I13.2 | Hypertensive heart and chronic kidney disease with heart failure and with stage 5 chronic kidney disease, or end stage renal disease |
| ICD10CM | N18.30 | Chronic kidney disease, stage 3 unspecified |
| ICD10CM | N18.31 | Chronic kidney disease, stage 3a |
| ICD10CM | N18.32 | Chronic kidney disease, stage 3b |
| ICD10CM | N18 | Chronic kidney disease (CKD) |
| ICD10CM | N18.1 | Chronic kidney disease, stage 1 |
| ICD10CM | N18.2 | Chronic kidney disease, stage 2 (mild) |
| ICD10CM | N18.3 | Chronic kidney disease, stage 3 (moderate) |
| ICD10CM | N18.4 | Chronic kidney disease, stage 4 (severe) |
| ICD10CM | N18.5 | Chronic kidney disease, stage 5 |
| ICD10CM | N18.6 | End stage renal disease |
| ICD10CM | N18.9 | Chronic kidney disease, unspecified |
| ICD10CM | O10.211 | Pre-existing hypertensive chronic kidney disease complicating pregnancy, first trimester |

Continued on next page

**Table7 – continued from previous page**

| CodeSystem | ConceptCode | ConceptName |
| --- | --- | --- |
| ICD10CM | O10.212 | Pre-existing hypertensive chronic kidney disease complicating pregnancy, second trimester |
| ICD10CM | O10.213 | Pre-existing hypertensive chronic kidney disease complicating pregnancy, third trimester |
| ICD10CM | O10.219 | Pre-existing hypertensive chronic kidney disease complicating pregnancy, unspecified trimester |
| ICD10CM | O10.22 | Pre-existing hypertensive chronic kidney disease complicating childbirth |
| ICD10CM | O10.31 | Pre-existing hypertensive heart and chronic kidney disease complicating pregnancy |
| ICD10CM | O10.311 | Pre-existing hypertensive heart and chronic kidney disease complicating pregnancy, first trimester |
| ICD10CM | O10.312 | Pre-existing hypertensive heart and chronic kidney disease complicating pregnancy, second trimester |
| ICD10CM | O10.313 | Pre-existing hypertensive heart and chronic kidney disease complicating pregnancy, third trimester |
| ICD10CM | O10.319 | Pre-existing hypertensive heart and chronic kidney disease complicating pregnancy, unspecified trimester |
| ICD10CM | O10.32 | Pre-existing hypertensive heart and chronic kidney disease complicating childbirth |
| ICD10CM | O10.33 | Pre-existing hypertensive heart and chronic kidney disease complicating the puerperium |
| ICD10CM | Q61.2 | Polycystic kidney, adult type |
| ICD10CM | Q61.3 | Polycystic kidney, unspecified |
| ICD10CM | Q61.8 | Other cystic kidney diseases |
| ICD10CM | Z94.0 | Kidney transplant status |
| SNOMED CT | 709044004 | Chronic kidney disease |
| SNOMED CT | 16726004 | Renal osteodystrophy |
| SNOMED CT | 1208934006 | Sagliker syndrome |
| SNOMED CT | 776416004 | Hyperuricemia, pulmonary hypertension, renal failure, alkalosis syndrome |
| SNOMED CT | 711000119100 | Chronic kidney disease stage 5 due to type 2 diabetes mellitus |
| SNOMED CT | 721000119107 | Chronic kidney disease stage 4 due to type 2 diabetes mellitus |
| SNOMED CT | 731000119105 | Chronic kidney disease stage 3 due to type 2 diabetes mellitus |
| SNOMED CT | 741000119101 | Chronic kidney disease stage 2 due to type 2 diabetes mellitus |
| SNOMED CT | 751000119104 | Chronic kidney disease stage 1 due to type 2 diabetes mellitus |
| SNOMED CT | 771000119108 | Chronic kidney disease due to type 2 diabetes mellitus |

Continued on next page

**Table7 – continued from previous page**

| CodeSystem | ConceptCode | ConceptName |
|---|---|---|
| SNOMED CT | 1801000119106 | Anemia, pre-end stage renal disease on erythropoietin protocol |
| SNOMED CT | 8501000119104 | Hypertensive heart and chronic kidney disease |
| SNOMED CT | 71421000119105 | Hypertension in chronic kidney disease due to type 2 diabetes mellitus |
| SNOMED CT | 71701000119105 | Hypertension in chronic kidney disease due to type 1 diabetes mellitus |
| SNOMED CT | 90721000119101 | Chronic kidney disease stage 1 due to type 1 diabetes mellitus |
| SNOMED CT | 90731000119103 | Chronic kidney disease stage 2 due to type 1 diabetes mellitus |
| SNOMED CT | 90741000119107 | Chronic kidney disease stage 3 due to type 1 diabetes mellitus |
| SNOMED CT | 90751000119109 | Chronic kidney disease stage 4 due to type 1 diabetes mellitus |
| SNOMED CT | 90761000119106 | Chronic kidney disease stage 5 due to type 1 diabetes mellitus |
| SNOMED CT | 90771000119100 | End stage renal disease on dialysis due to type 1 diabetes mellitus |
| SNOMED CT | 90791000119104 | End stage renal disease on dialysis due to type 2 diabetes mellitus |
| SNOMED CT | 96441000119101 | Chronic kidney disease due to type 1 diabetes mellitus |
| SNOMED CT | 96701000119107 | Hypertensive heart AND chronic kidney disease on dialysis |
| SNOMED CT | 96711000119105 | Hypertensive heart AND chronic kidney disease stage 5 |
| SNOMED CT | 96721000119103 | Hypertensive heart AND chronic kidney disease stage 4 |
| SNOMED CT | 96731000119100 | Hypertensive heart AND chronic kidney disease stage 3 |
| SNOMED CT | 96741000119109 | Hypertensive heart AND chronic kidney disease stage 2 |
| SNOMED CT | 96751000119106 | Hypertensive heart AND chronic kidney disease stage 1 |
| SNOMED CT | 104931000119100 | Chronic kidney disease due to hypertension |
| SNOMED CT | 111411000119103 | End stage renal disease due to hypertension |
| SNOMED CT | 117681000119102 | Chronic kidney disease stage 1 due to hypertension |
| SNOMED CT | 118781000119108 | Pre-existing hypertensive chronic kidney disease in mother complicating pregnancy |
| SNOMED CT | 120261000119101 | Benign hypertensive heart disease and chronic renal disease stage 5 |
| SNOMED CT | 127991000119101 | Hypertension concurrent and due to end stage renal disease on dialysis due to type 2 diabetes mellitus |
| SNOMED CT | 128001000119105 | Hypertension concurrent and due to end stage renal disease on dialysis due to type 1 diabetes mellitus |
| SNOMED CT | 129151000119102 | Chronic kidney disease stage 4 due to hypertension |
| SNOMED CT | 129161000119100 | Chronic kidney disease stage 5 due to hypertension |
| SNOMED CT | 129171000119106 | Chronic kidney disease stage 3 due to hypertension |
| SNOMED CT | 129181000119109 | Chronic kidney disease stage 2 due to hypertension |

Continued on next page

**Table7 – continued from previous page**

| CodeSystem | ConceptCode | ConceptName |
|---|---|---|
| SNOMED CT | 140101000119109 | Hypertension in chronic kidney disease stage 5 due to type 2 diabetes mellitus |
| SNOMED CT | 140111000119107 | Hypertension in chronic kidney disease stage 4 due to type 2 diabetes mellitus |
| SNOMED CT | 140121000119100 | Hypertension in chronic kidney disease stage 3 due to type 2 diabetes mellitus |
| SNOMED CT | 140131000119102 | Hypertension in chronic kidney disease stage 2 due to type 2 diabetes mellitus |
| SNOMED CT | 153851000119106 | Malignant hypertensive chronic kidney disease stage 5 |
| SNOMED CT | 153891000119101 | End stage renal disease on dialysis due to hypertension |
| SNOMED CT | 284961000119106 | Chronic kidney disease due to benign hypertension |
| SNOMED CT | 284971000119100 | Chronic kidney disease stage 1 due to benign hypertension |
| SNOMED CT | 284981000119102 | Chronic kidney disease stage 2 due to benign hypertension |
| SNOMED CT | 284991000119104 | Chronic kidney disease stage 3 due to benign hypertension |
| SNOMED CT | 285001000119105 | Chronic kidney disease stage 4 due to benign hypertension |
| SNOMED CT | 285011000119108 | Chronic kidney disease stage 5 due to benign hypertension |
| SNOMED CT | 285041000119107 | Benign hypertensive heart disease and chronic renal disease stage 1 |
| SNOMED CT | 285061000119106 | Benign hypertensive heart disease and chronic renal disease stage 2 |
| SNOMED CT | 285081000119102 | Benign hypertensive heart disease and chronic renal disease stage 3 |
| SNOMED CT | 285101000119109 | Benign hypertensive heart disease and chronic renal disease stage 4 |
| SNOMED CT | 285831000119108 | Malignant hypertensive chronic kidney disease |
| SNOMED CT | 285841000119104 | Malignant hypertensive end stage renal disease |
| SNOMED CT | 285851000119102 | Malignant hypertensive chronic kidney disease stage 1 |
| SNOMED CT | 285861000119100 | Malignant hypertensive chronic kidney disease stage 2 |
| SNOMED CT | 285871000119106 | Malignant hypertensive chronic kidney disease stage 3 |
| SNOMED CT | 285881000119109 | Malignant hypertensive chronic kidney disease stage 4 |
| SNOMED CT | 285911000119109 | Malignant hypertensive heart disease and chronic renal disease stage 3 |
| SNOMED CT | 285921000119102 | Malignant hypertensive heart disease and chronic renal disease stage 4 |
| SNOMED CT | 286371000119107 | Malignant hypertensive end stage renal disease on dialysis |
| SNOMED CT | 434431000124103 | Hypertensive end stage renal disease |
| SNOMED CT | 449631000124102 | Benign hypertensive heart disease and chronic renal disease |
| SNOMED CT | 691421000119108 | Anemia co-occurrent and due to chronic kidney disease stage 3 |
| SNOMED CT | 10757401000119104 | Pre-existing hypertensive heart and chronic kidney disease in mother complicating childbirth |
| SNOMED CT | 10757481000119107 | Pre-existing hypertensive heart and chronic kidney disease in mother complicating pregnancy |

Continued on next page

**Table7 – continued from previous page**

| CodeSystem | ConceptCode | ConceptName |
|---|---|---|
| SNOMED CT | 236433006 | Acute-on-chronic renal failure |
| SNOMED CT | 236434000 | End stage renal failure untreated by renal replacement therapy |
| SNOMED CT | 236435004 | End stage renal failure on dialysis |
| SNOMED CT | 236436003 | End stage renal failure with renal transplant |
| SNOMED CT | 236552002 | Adynamic bone disease |
| SNOMED CT | 425369003 | Chronic progressive renal failure |
| SNOMED CT | 46177005 | End-stage renal disease |
| SNOMED CT | 49708008 | Anemia of chronic renal failure |
| SNOMED CT | 57557005 | Chronic milk alkali syndrome |
| SNOMED CT | 431855005 | Chronic kidney disease stage 1 |
| SNOMED CT | 431856006 | Chronic kidney disease stage 2 |
| SNOMED CT | 431857002 | Chronic kidney disease stage 4 |
| SNOMED CT | 433144002 | Chronic kidney disease stage 3 |
| SNOMED CT | 433146000 | Chronic kidney disease stage 5 |
| SNOMED CT | 700378005 | Chronic kidney disease stage 3A |
| SNOMED CT | 700379002 | Chronic kidney disease stage 3B |
| SNOMED CT | 704667004 | Hypertension concurrent and due to end stage renal disease on dialysis |
| SNOMED CT | 707324008 | Anemia in end stage renal disease |
| SNOMED CT | 708975004 | Mixed renal osteodystrophy |
| SNOMED CT | 712487000 | End stage renal disease due to benign hypertension |
| SNOMED CT | 713313000 | Chronic kidney disease mineral and bone disorder |
| SNOMED CT | 714152005 | Chronic kidney disease stage 5 on dialysis |
| SNOMED CT | 714153000 | Chronic kidney disease stage 5 with transplant |
| SNOMED CT | 722098007 | Chronic kidney disease following donor nephrectomy |
| SNOMED CT | 722149000 | Chronic kidney disease due to and following excision of neoplasm of kidney |
| SNOMED CT | 722150000 | Chronic kidney disease due to systemic infection |
| SNOMED CT | 722467000 | Chronic kidney disease due to traumatic loss of kidney |
| SNOMED CT | 723190009 | Chronic renal insufficiency |
| SNOMED CT | 368421000119108 | Chronic kidney disease stage 1 due to drug induced diabetes mellitus |
| SNOMED CT | 368431000119106 | Chronic kidney disease stage 2 due to drug induced diabetes mellitus |
| SNOMED CT | 368441000119102 | Chronic kidney disease stage 3 due to drug induced diabetes mellitus |
| SNOMED CT | 368451000119100 | Chronic kidney disease stage 4 due to drug induced diabetes mellitus |
| SNOMED CT | 368461000119103 | Chronic kidney disease stage 5 due to drug induced diabetes mellitus |
| SNOMED CT | 368471000119109 | End stage renal disease on dialysis due to drug induced diabetes mellitus |

Continued on next page

**Table7 – continued from previous page**

| CodeSystem | ConceptCode | ConceptName |
|---|---|---|
| SNOMED CT | 897308007 | Chronic kidney disease with osteoporosis |
| SNOMED CT | 897310009 | Renal osteodystrophy with high bone turnover |
| SNOMED CT | 897311008 | Renal osteodystrophy with normal bone turnover |
| SNOMED CT | 897312001 | Renal osteodystrophy with low bone turnover |
| SNOMED CT | 90688005 | Chronic renal failure |
| SNOMED CT | 1217070004 | Renal osteodystrophy due to hyperparathyroidism |

##### 0.4.5 COPD

Table 8: Concept codes used to identify chronic obstructive pulmonary disease conditions.

| CodeSystem | ConceptCode | ConceptName |
|---|---|---|
| ICD10CM | J41 | Simple and mucopurulent chronic bronchitis |
| ICD10CM | J42 | Unspecified chronic bronchitis |
| ICD10CM | J43 | Emphysema |
| ICD10CM | J44.0 | Chronic obstructive pulmonary disease with (acute) lower respiratory infection |
| ICD10CM | J44.1 | Chronic obstructive pulmonary disease with (acute) exacerbation |
| ICD10CM | J44.9 | Chronic obstructive pulmonary disease, unspecified |
| ICD10CM | J41.0 | Simple chronic bronchitis |
| ICD10CM | J41.1 | Mucopurulent chronic bronchitis |
| ICD10CM | J41.8 | Mixed simple and mucopurulent chronic bronchitis |
| ICD10CM | J43.0 | Unilateral pulmonary emphysema [MacLeod's syndrome] |
| ICD10CM | J43.1 | Panlobular emphysema |
| ICD10CM | J43.2 | Centrilobular emphysema |
| ICD10CM | J43.8 | Other emphysema |
| ICD10CM | J43.9 | Emphysema, unspecified |
| SNOMED CT | 13645005 | Chronic obstructive lung disease |
| SNOMED CT | 195951007 | Acute exacerbation of chronic obstructive airways disease |
| SNOMED CT | 196001008 | Chronic obstructive pulmonary disease with acute lower respiratory infection |
| SNOMED CT | 135836000 | End stage chronic obstructive airways disease |
| SNOMED CT | 313296004 | Mild chronic obstructive pulmonary disease |
| SNOMED CT | 313297008 | Moderate chronic obstructive pulmonary disease |
| SNOMED CT | 313299006 | Severe chronic obstructive pulmonary disease |
| SNOMED CT | 708030004 | Pulmonary emphysema co-occurrent with fibrosis of lung |
| SNOMED CT | 285381006 | Acute infective exacerbation of chronic obstructive airways disease |
| SNOMED CT | 86680006 | Ruptured emphysematous bleb of lung |

Continued on next page

**Table8 – continued from previous page**

| CodeSystem | ConceptCode | ConceptName |
|---|---|---|
| SNOMED CT | 87433001 | Pulmonary emphysema |
| SNOMED CT | 4981000 | Panacinar emphysema |
| SNOMED CT | 16003001 | Giant bullous emphysema |
| SNOMED CT | 16846004 | Obstructive emphysema |
| SNOMED CT | 23958009 | Vanishing lung |
| SNOMED CT | 31898008 | Paraseptal emphysema |
| SNOMED CT | 33325001 | Compensatory emphysema |
| SNOMED CT | 1177120001 | Bronchiolitis obliterans syndrome due to and following allogeneic stem cell transplant |
| SNOMED CT | 762618008 | Bronchiolitis obliterans syndrome due to and after lung transplantation |
| SNOMED CT | 836477007 | Chronic emphysema due to vapor |
| SNOMED CT | 840350008 | Chronic obliterative bronchiolitis due to chemical fumes |
| SNOMED CT | 840351007 | Chronic obliterative bronchiolitis due to vapor |
| SNOMED CT | 1751000119100 | Acute exacerbation of chronic obstructive airways disease with asthma |
| SNOMED CT | 106001000119101 | Chronic obstructive lung disease co-occurrent with acute bronchitis |
| SNOMED CT | 10692761000119107 | Asthma-chronic obstructive pulmonary disease overlap syndrome |
| SNOMED CT | 233674008 | Pulmonary emphysema in alpha-1 primary immunodeficiency deficiency |
| SNOMED CT | 195957006 | Chronic bullous emphysema |
| SNOMED CT | 195958001 | Segmental bullous emphysema |
| SNOMED CT | 195959009 | Zonal bullous emphysema |
| SNOMED CT | 196026004 | Chronic emphysema due to chemical fumes |
| SNOMED CT | 233675009 | Toxic emphysema |
| SNOMED CT | 233677001 | Scar emphysema |
| SNOMED CT | 47895001 | Congenital emphysema |
| SNOMED CT | 47938003 | Chronic obliterative bronchiolitis |
| SNOMED CT | 57686001 | Emphysematous bleb of lung |
| SNOMED CT | 60805002 | Hemolytic anemia with emphysema AND cutis laxa |
| SNOMED CT | 66987001 | Congenital lobar emphysema |
| SNOMED CT | 68328006 | Centriacinar emphysema |
| SNOMED CT | 70756004 | Bronchial atresia with segmental pulmonary emphysema |
| SNOMED CT | 77690003 | Interstitial emphysema of lung |
| SNOMED CT | 266355005 | Bullous emphysema with collapse |
| SNOMED CT | 266356006 | Atrophic (senile) emphysema |
| SNOMED CT | 1010333003 | Emphysema of left lung |
| SNOMED CT | 1010334009 | Emphysema of right lung |

##### 0.4.6 Glaucoma

Table 9: Concept codes used to identify glaucoma conditions.

| CodeSystem | ConceptCode | ConceptName |
|---|---|---|
| ICD10CM | H40.10X0 | Unspecified open-angle glaucoma, stage unspecified |
| ICD10CM | H40.10X1 | Unspecified open-angle glaucoma, mild stage |
| ICD10CM | H40.10X2 | Unspecified open-angle glaucoma, moderate stage |
| ICD10CM | H40.10X3 | Unspecified open-angle glaucoma, severe stage |
| ICD10CM | H40.10X4 | Unspecified open-angle glaucoma, indeterminate stage |
| ICD10CM | H40.1110 | Primary open-angle glaucoma, right eye, stage unspecified |
| ICD10CM | H40.1111 | Primary open-angle glaucoma, right eye, mild stage |
| ICD10CM | H40.1112 | Primary open-angle glaucoma, right eye, moderate stage |
| ICD10CM | H40.1113 | Primary open-angle glaucoma, right eye, severe stage |
| ICD10CM | H40.1114 | Primary open-angle glaucoma, right eye, indeterminate stage |
| ICD10CM | H40.1120 | Primary open-angle glaucoma, left eye, stage unspecified |
| ICD10CM | H40.1121 | Primary open-angle glaucoma, left eye, mild stage |
| ICD10CM | H40.1122 | Primary open-angle glaucoma, left eye, moderate stage |
| ICD10CM | H40.1123 | Primary open-angle glaucoma, left eye, severe stage |
| ICD10CM | H40.1124 | Primary open-angle glaucoma, left eye, indeterminate stage |
| ICD10CM | H40.1130 | Primary open-angle glaucoma, bilateral, stage unspecified |
| ICD10CM | H40.1131 | Primary open-angle glaucoma, bilateral, mild stage |
| ICD10CM | H40.1132 | Primary open-angle glaucoma, bilateral, moderate stage |
| ICD10CM | H40.1133 | Primary open-angle glaucoma, bilateral, severe stage |
| ICD10CM | H40.1134 | Primary open-angle glaucoma, bilateral, indeterminate stage |
| ICD10CM | H40.1210 | Low-tension glaucoma, right eye, stage unspecified |
| ICD10CM | H40.1211 | Low-tension glaucoma, right eye, mild stage |
| ICD10CM | H40.1212 | Low-tension glaucoma, right eye, moderate stage |
| ICD10CM | H40.1213 | Low-tension glaucoma, right eye, severe stage |
| ICD10CM | H40.1214 | Low-tension glaucoma, right eye, indeterminate stage |
| ICD10CM | H40.1220 | Low-tension glaucoma, left eye, stage unspecified |
| ICD10CM | H40.1221 | Low-tension glaucoma, left eye, mild stage |
| ICD10CM | H40.1222 | Low-tension glaucoma, left eye, moderate stage |
| ICD10CM | H40.1223 | Low-tension glaucoma, left eye, severe stage |
| ICD10CM | H40.1224 | Low-tension glaucoma, left eye, indeterminate stage |
| ICD10CM | H40.1230 | Low-tension glaucoma, bilateral, stage unspecified |
| ICD10CM | H40.1231 | Low-tension glaucoma, bilateral, mild stage |
| ICD10CM | H40.1232 | Low-tension glaucoma, bilateral, moderate stage |
| ICD10CM | H40.1233 | Low-tension glaucoma, bilateral, severe stage |
| ICD10CM | H40.1234 | Low-tension glaucoma, bilateral, indeterminate stage |
| ICD10CM | H40.1310 | Pigmentary glaucoma, right eye, stage unspecified |
| ICD10CM | H40.1311 | Pigmentary glaucoma, right eye, mild stage |
| ICD10CM | H40.1312 | Pigmentary glaucoma, right eye, moderate stage |
| ICD10CM | H40.1313 | Pigmentary glaucoma, right eye, severe stage |
| ICD10CM | H40.1314 | Pigmentary glaucoma, right eye, indeterminate stage |

Continued on next page

**Table9 – continued from previous page**

| CodeSystem | ConceptCode | ConceptName |
|---|---|---|
| ICD10CM | H40.1320 | Pigmentary glaucoma, left eye, stage unspecified |
| ICD10CM | H40.1321 | Pigmentary glaucoma, left eye, mild stage |
| ICD10CM | H40.1322 | Pigmentary glaucoma, left eye, moderate stage |
| ICD10CM | H40.1323 | Pigmentary glaucoma, left eye, severe stage |
| ICD10CM | H40.1324 | Pigmentary glaucoma, left eye, indeterminate stage |
| ICD10CM | H40.1330 | Pigmentary glaucoma, bilateral, stage unspecified |
| ICD10CM | H40.1331 | Pigmentary glaucoma, bilateral, mild stage |
| ICD10CM | H40.1332 | Pigmentary glaucoma, bilateral, moderate stage |
| ICD10CM | H40.1333 | Pigmentary glaucoma, bilateral, severe stage |
| ICD10CM | H40.1334 | Pigmentary glaucoma, bilateral, indeterminate stage |
| ICD10CM | H40.1410 | Capsular glaucoma with pseudoexfoliation of lens, right eye, stage unspecified |
| ICD10CM | H40.1411 | Capsular glaucoma with pseudoexfoliation of lens, right eye, mild stage |
| ICD10CM | H40.1412 | Capsular glaucoma with pseudoexfoliation of lens, right eye, moderate stage |
| ICD10CM | H40.1413 | Capsular glaucoma with pseudoexfoliation of lens, right eye, severe stage |
| ICD10CM | H40.1414 | Capsular glaucoma with pseudoexfoliation of lens, right eye, indeterminate stage |
| ICD10CM | H40.1420 | Capsular glaucoma with pseudoexfoliation of lens, left eye, stage unspecified |
| ICD10CM | H40.1421 | Capsular glaucoma with pseudoexfoliation of lens, left eye, mild stage |
| ICD10CM | H40.1422 | Capsular glaucoma with pseudoexfoliation of lens, left eye, moderate stage |
| ICD10CM | H40.1423 | Capsular glaucoma with pseudoexfoliation of lens, left eye, severe stage |
| ICD10CM | H40.1424 | Capsular glaucoma with pseudoexfoliation of lens, left eye, indeterminate stage |
| ICD10CM | H40.1430 | Capsular glaucoma with pseudoexfoliation of lens, bilateral, stage unspecified |
| ICD10CM | H40.1431 | Capsular glaucoma with pseudoexfoliation of lens, bilateral, mild stage |
| ICD10CM | H40.1432 | Capsular glaucoma with pseudoexfoliation of lens, bilateral, moderate stage |
| ICD10CM | H40.1433 | Capsular glaucoma with pseudoexfoliation of lens, bilateral, severe stage |
| ICD10CM | H40.1434 | Capsular glaucoma with pseudoexfoliation of lens, bilateral, indeterminate stage |
| ICD10CM | H40.151 | Residual stage of open-angle glaucoma, right eye |
| ICD10CM | H40.152 | Residual stage of open-angle glaucoma, left eye |
| ICD10CM | H40.153 | Residual stage of open-angle glaucoma, bilateral |

Continued on next page

**Table9 – continued from previous page**

| CodeSystem | ConceptCode | ConceptName |
|---|---|---|
| ICD10CM | H40.20X0 | Unspecified primary angle-closure glaucoma, stage unspecified |
| ICD10CM | H40.20X1 | Unspecified primary angle-closure glaucoma, mild stage |
| ICD10CM | H40.20X2 | Unspecified primary angle-closure glaucoma, moderate stage |
| ICD10CM | H40.20X3 | Unspecified primary angle-closure glaucoma, severe stage |
| ICD10CM | H40.20X4 | Unspecified primary angle-closure glaucoma, indeterminate stage |
| ICD10CM | H40.211 | Acute angle-closure glaucoma, right eye |
| ICD10CM | H40.212 | Acute angle-closure glaucoma, left eye |
| ICD10CM | H40.213 | Acute angle-closure glaucoma, bilateral |
| ICD10CM | H40.2210 | Chronic angle-closure glaucoma, right eye, stage unspecified |
| ICD10CM | H40.2211 | Chronic angle-closure glaucoma, right eye, mild stage |
| ICD10CM | H40.2212 | Chronic angle-closure glaucoma, right eye, moderate stage |
| ICD10CM | H40.2213 | Chronic angle-closure glaucoma, right eye, severe stage |
| ICD10CM | H40.2214 | Chronic angle-closure glaucoma, right eye, indeterminate stage |
| ICD10CM | H40.2220 | Chronic angle-closure glaucoma, left eye, stage unspecified |
| ICD10CM | H40.2221 | Chronic angle-closure glaucoma, left eye, mild stage |
| ICD10CM | H40.2222 | Chronic angle-closure glaucoma, left eye, moderate stage |
| ICD10CM | H40.2223 | Chronic angle-closure glaucoma, left eye, severe stage |
| ICD10CM | H40.2224 | Chronic angle-closure glaucoma, left eye, indeterminate stage |
| ICD10CM | H40.2230 | Chronic angle-closure glaucoma, bilateral, stage unspecified |
| ICD10CM | H40.2231 | Chronic angle-closure glaucoma, bilateral, mild stage |
| ICD10CM | H40.2232 | Chronic angle-closure glaucoma, bilateral, moderate stage |
| ICD10CM | H40.2233 | Chronic angle-closure glaucoma, bilateral, severe stage |
| ICD10CM | H40.2234 | Chronic angle-closure glaucoma, bilateral, indeterminate stage |
| ICD10CM | H40.231 | Intermittent angle-closure glaucoma, right eye |
| ICD10CM | H40.232 | Intermittent angle-closure glaucoma, left eye |
| ICD10CM | H40.233 | Intermittent angle-closure glaucoma, bilateral |
| ICD10CM | H40.241 | Residual stage of angle-closure glaucoma, right eye |
| ICD10CM | H40.242 | Residual stage of angle-closure glaucoma, left eye |
| ICD10CM | H40.243 | Residual stage of angle-closure glaucoma, bilateral |
| ICD10CM | H40.31X0 | Glaucoma secondary to eye trauma, right eye, stage unspecified |
| ICD10CM | H40.31X1 | Glaucoma secondary to eye trauma, right eye, mild stage |
| ICD10CM | H40.31X2 | Glaucoma secondary to eye trauma, right eye, moderate stage |
| ICD10CM | H40.31X3 | Glaucoma secondary to eye trauma, right eye, severe stage |
| ICD10CM | H40.31X4 | Glaucoma secondary to eye trauma, right eye, indeterminate stage |

Continued on next page

**Table9 – continued from previous page**

| CodeSystem | ConceptCode | ConceptName |
|---|---|---|
| ICD10CM | H40.32X0 | Glaucoma secondary to eye trauma, left eye, stage unspecified |
| ICD10CM | H40.32X1 | Glaucoma secondary to eye trauma, left eye, mild stage |
| ICD10CM | H40.32X2 | Glaucoma secondary to eye trauma, left eye, moderate stage |
| ICD10CM | H40.32X3 | Glaucoma secondary to eye trauma, left eye, severe stage |
| ICD10CM | H40.32X4 | Glaucoma secondary to eye trauma, left eye, indeterminate stage |
| ICD10CM | H40.33X0 | Glaucoma secondary to eye trauma, bilateral, stage unspecified |
| ICD10CM | H40.33X1 | Glaucoma secondary to eye trauma, bilateral, mild stage |
| ICD10CM | H40.33X2 | Glaucoma secondary to eye trauma, bilateral, moderate stage |
| ICD10CM | H40.33X3 | Glaucoma secondary to eye trauma, bilateral, severe stage |
| ICD10CM | H40.33X4 | Glaucoma secondary to eye trauma, bilateral, indeterminate stage |
| ICD10CM | H40.41X0 | Glaucoma secondary to eye inflammation, right eye, stage unspecified |
| ICD10CM | H40.41X1 | Glaucoma secondary to eye inflammation, right eye, mild stage |
| ICD10CM | H40.41X2 | Glaucoma secondary to eye inflammation, right eye, moderate stage |
| ICD10CM | H40.41X3 | Glaucoma secondary to eye inflammation, right eye, severe stage |
| ICD10CM | H40.41X4 | Glaucoma secondary to eye inflammation, right eye, indeterminate stage |
| ICD10CM | H40.42X0 | Glaucoma secondary to eye inflammation, left eye, stage unspecified |
| ICD10CM | H40.42X1 | Glaucoma secondary to eye inflammation, left eye, mild stage |
| ICD10CM | H40.42X2 | Glaucoma secondary to eye inflammation, left eye, moderate stage |
| ICD10CM | H40.42X3 | Glaucoma secondary to eye inflammation, left eye, severe stage |
| ICD10CM | H40.42X4 | Glaucoma secondary to eye inflammation, left eye, indeterminate stage |
| ICD10CM | H40.43X0 | Glaucoma secondary to eye inflammation, bilateral, stage unspecified |
| ICD10CM | H40.43X1 | Glaucoma secondary to eye inflammation, bilateral, mild stage |
| ICD10CM | H40.43X2 | Glaucoma secondary to eye inflammation, bilateral, moderate stage |
| ICD10CM | H40.43X3 | Glaucoma secondary to eye inflammation, bilateral, severe stage |

Continued on next page

**Table9 – continued from previous page**

| CodeSystem | ConceptCode | ConceptName |
| --- | --- | --- |
| ICD10CM | H40.43X4 | Glaucoma secondary to eye inflammation, bilateral, indeterminate stage |
| ICD10CM | H40.51X0 | Glaucoma secondary to other eye disorders, right eye, stage unspecified |
| ICD10CM | H40.51X1 | Glaucoma secondary to other eye disorders, right eye, mild stage |
| ICD10CM | H40.51X2 | Glaucoma secondary to other eye disorders, right eye, moderate stage |
| ICD10CM | H40.51X3 | Glaucoma secondary to other eye disorders, right eye, severe stage |
| ICD10CM | H40.51X4 | Glaucoma secondary to other eye disorders, right eye, indeterminate stage |
| ICD10CM | H40.52X0 | Glaucoma secondary to other eye disorders, left eye, stage unspecified |
| ICD10CM | H40.52X1 | Glaucoma secondary to other eye disorders, left eye, mild stage |
| ICD10CM | H40.52X2 | Glaucoma secondary to other eye disorders, left eye, moderate stage |
| ICD10CM | H40.52X3 | Glaucoma secondary to other eye disorders, left eye, severe stage |
| ICD10CM | H40.52X4 | Glaucoma secondary to other eye disorders, left eye, indeterminate stage |
| ICD10CM | H40.53X0 | Glaucoma secondary to other eye disorders, bilateral, stage unspecified |
| ICD10CM | H40.53X1 | Glaucoma secondary to other eye disorders, bilateral, mild stage |
| ICD10CM | H40.53X2 | Glaucoma secondary to other eye disorders, bilateral, moderate stage |
| ICD10CM | H40.53X3 | Glaucoma secondary to other eye disorders, bilateral, severe stage |
| ICD10CM | H40.53X4 | Glaucoma secondary to other eye disorders, bilateral, indeterminate stage |
| ICD10CM | H40.61X0 | Glaucoma secondary to drugs, right eye, stage unspecified |
| ICD10CM | H40.61X1 | Glaucoma secondary to drugs, right eye, mild stage |
| ICD10CM | H40.61X2 | Glaucoma secondary to drugs, right eye, moderate stage |
| ICD10CM | H40.61X3 | Glaucoma secondary to drugs, right eye, severe stage |
| ICD10CM | H40.61X4 | Glaucoma secondary to drugs, right eye, indeterminate stage |
| ICD10CM | H40.62X0 | Glaucoma secondary to drugs, left eye, stage unspecified |
| ICD10CM | H40.62X1 | Glaucoma secondary to drugs, left eye, mild stage |
| ICD10CM | H40.62X2 | Glaucoma secondary to drugs, left eye, moderate stage |
| ICD10CM | H40.62X3 | Glaucoma secondary to drugs, left eye, severe stage |
| ICD10CM | H40.62X4 | Glaucoma secondary to drugs, left eye, indeterminate stage |

Continued on next page

**Table9 – continued from previous page**

| CodeSystem | ConceptCode | ConceptName |
|---|---|---|
| ICD10CM | H40.63X0 | Glaucoma secondary to drugs, bilateral, stage unspecified |
| ICD10CM | H40.63X1 | Glaucoma secondary to drugs, bilateral, mild stage |
| ICD10CM | H40.63X2 | Glaucoma secondary to drugs, bilateral, moderate stage |
| ICD10CM | H40.63X3 | Glaucoma secondary to drugs, bilateral, severe stage |
| ICD10CM | H40.63X4 | Glaucoma secondary to drugs, bilateral, indeterminate stage |
| ICD10CM | H40.811 | Glaucoma with increased episcleral venous pressure, right eye |
| ICD10CM | H40.812 | Glaucoma with increased episcleral venous pressure, left eye |
| ICD10CM | H40.813 | Glaucoma with increased episcleral venous pressure, bilateral |
| ICD10CM | H40.821 | Hypersecretion glaucoma, right eye |
| ICD10CM | H40.822 | Hypersecretion glaucoma, left eye |
| ICD10CM | H40.823 | Hypersecretion glaucoma, bilateral |
| ICD10CM | H40.831 | Aqueous misdirection, right eye |
| ICD10CM | H40.832 | Aqueous misdirection, left eye |
| ICD10CM | H40.833 | Aqueous misdirection, bilateral |
| ICD10CM | H40.89 | Other specified glaucoma |
| ICD10CM | Q15.0 | Congenital glaucoma |
| ICD10CM | H40.1510 | Residual stage of open-angle glaucoma, right eye, stage unspecified |
| ICD10CM | H40.1511 | Residual stage of open-angle glaucoma, right eye, mild stage |
| ICD10CM | H40.1512 | Residual stage of open-angle glaucoma, right eye, moderate stage |
| ICD10CM | H40.1513 | Residual stage of open-angle glaucoma, right eye, severe stage |
| ICD10CM | H40.1514 | Residual stage of open-angle glaucoma, right eye, indeterminate stage |
| ICD10CM | H40.1520 | Residual stage of open-angle glaucoma, left eye, stage unspecified |
| ICD10CM | H40.1521 | Residual stage of open-angle glaucoma, left eye, mild stage |
| ICD10CM | H40.1522 | Residual stage of open-angle glaucoma, left eye, moderate stage |
| ICD10CM | H40.1523 | Residual stage of open-angle glaucoma, left eye, severe stage |
| ICD10CM | H40.1524 | Residual stage of open-angle glaucoma, left eye, indeterminate stage |
| ICD10CM | H40.1530 | Residual stage of open-angle glaucoma, bilateral, stage unspecified |
| ICD10CM | H40.1531 | Residual stage of open-angle glaucoma, bilateral, mild stage |
| ICD10CM | H40.1532 | Residual stage of open-angle glaucoma, bilateral, moderate stage |
| ICD10CM | H40.1533 | Residual stage of open-angle glaucoma, bilateral, severe stage |

Continued on next page

**Table9 – continued from previous page**

| CodeSystem | ConceptCode | ConceptName |
|---|---|---|
| ICD10CM | H40.1534 | Residual stage of open-angle glaucoma, bilateral, indeterminate stage |
| SNOMED CT | 1654001 | Corticosteroid-induced open angle glaucoma |
| SNOMED CT | 15374009 | Aphakic glaucoma |
| SNOMED CT | 19144002 | Absolute glaucoma |
| SNOMED CT | 19309007 | Glaucoma associated with vascular disorder |
| SNOMED CT | 21571006 | Secondary angle-closure glaucoma |
| SNOMED CT | 21928008 | Secondary open-angle glaucoma |
| SNOMED CT | 23986001 | Glaucoma |
| SNOMED CT | 27735002 | Glaucoma associated with tumors AND/OR cysts |
| SNOMED CT | 29369005 | Hypersecretion glaucoma |
| SNOMED CT | 30041005 | Acute angle-closure glaucoma |
| SNOMED CT | 32893002 | Phacolytic glaucoma |
| SNOMED CT | 33647009 | Chronic angle-closure glaucoma |
| SNOMED CT | 34623005 | Glaucoma with increased episcleral venous pressure |
| SNOMED CT | 787051000 | Open-angle glaucoma of left eye |
| SNOMED CT | 787052007 | Open-angle glaucoma of right eye |
| SNOMED CT | 24151000119106 | Steroid-induced open angle glaucoma |
| SNOMED CT | 41911000119107 | Glaucoma due to type 2 diabetes mellitus |
| SNOMED CT | 60981000119103 | Glaucoma due to diabetes mellitus type 1 |
| SNOMED CT | 336611000119109 | Acute angle-closure glaucoma of right eye |
| SNOMED CT | 336631000119104 | Absolute glaucoma right eye |
| SNOMED CT | 342221000119104 | Acute angle-closure glaucoma of left eye |
| SNOMED CT | 342241000119105 | Absolute glaucoma left eye |
| SNOMED CT | 347381000119106 | Bilateral acute angle-closure glaucoma |
| SNOMED CT | 347401000119106 | Bilateral absolute glaucoma |
| SNOMED CT | 12239301000119102 | Bilateral open-angle glaucoma |
| SNOMED CT | 12239421000119101 | Bilateral glaucoma |
| SNOMED CT | 12239461000119106 | Glaucoma of left eye |
| SNOMED CT | 12239501000119106 | Glaucoma of right eye |
| SNOMED CT | 15633281000119103 | Bilateral primary open angle glaucoma |
| SNOMED CT | 15633321000119108 | Primary open angle glaucoma of left eye |
| SNOMED CT | 15640441000119104 | Primary open angle glaucoma of right eye |
| SNOMED CT | 15673001000119103 | Congenital glaucoma of bilateral eyes |
| SNOMED CT | 15679801000119105 | Bilateral uveitis-glaucoma-hyphema syndrome of eyes |
| SNOMED CT | 15736441000119108 | Primary angle-closure glaucoma of bilateral eyes |
| SNOMED CT | 15736481000119103 | Primary angle-closure glaucoma of right eye |
| SNOMED CT | 15736521000119103 | Primary angle-closure glaucoma of left eye |
| SNOMED CT | 15736561000119108 | Neovascular glaucoma of bilateral eyes |
| SNOMED CT | 15736601000119108 | Neovascular glaucoma of right eye |
| SNOMED CT | 15736641000119105 | Neovascular glaucoma of left eye |
| SNOMED CT | 15736681000119100 | Narrow-angle glaucoma of right eye |
| SNOMED CT | 15736721000119106 | Bilateral angle-closure glaucoma |

Continued on next page

**Table9 – continued from previous page**

| CodeSystem | ConceptCode | ConceptName |
|---|---|---|
| SNOMED CT | 15736761000119101 | Narrow angle glaucoma of left eye |
| SNOMED CT | 15738841000119105 | Glaucoma of bilateral eyes due to combination of mechanisms |
| SNOMED CT | 15738881000119100 | Glaucoma of left eye due to combination of mechanisms |
| SNOMED CT | 15738921000119107 | Glaucoma of right eye due to combination of mechanisms |
| SNOMED CT | 15739041000119106 | Glaucoma of bilateral eyes due to iris anomaly |
| SNOMED CT | 15739081000119101 | Glaucoma of bilateral eyes due to anterior segment anomaly |
| SNOMED CT | 15739121000119104 | Glaucoma of right eye due to anterior segment anomaly |
| SNOMED CT | 15739161000119109 | Glaucoma of left eye due to anterior segment anomaly |
| SNOMED CT | 15739201000119104 | Glaucoma of left eye due to ocular vascular disorder |
| SNOMED CT | 15739241000119102 | Glaucoma of right eye due to ocular vascular disorder |
| SNOMED CT | 15739281000119107 | Glaucoma of bilateral eyes due to ocular vascular disorder |
| SNOMED CT | 15739321000119102 | Glaucoma of right eye due to chamber angle anomaly |
| SNOMED CT | 15739361000119107 | Glaucoma of bilateral eyes due to chamber angle anomaly |
| SNOMED CT | 15739401000119103 | Glaucoma of left eye due to chamber angle anomaly |
| SNOMED CT | 15739441000119101 | Glaucoma of right eye due to trauma |
| SNOMED CT | 15739481000119106 | Glaucoma of left eye due to trauma |
| SNOMED CT | 15739561000119101 | Glaucoma of right eye due to lens disorder |
| SNOMED CT | 15739641000119104 | Glaucoma of left eye due to lens disorder |
| SNOMED CT | 15739681000119109 | Glaucoma of right eye due to systemic disorder |
| SNOMED CT | 15739721000119103 | Glaucoma of left eye due to systemic disorder |
| SNOMED CT | 15739761000119108 | Glaucoma of bilateral eyes due to systemic disorder |
| SNOMED CT | 15993671000119108 | Open-angle glaucoma of right eye caused by steroid |
| SNOMED CT | 15993711000119107 | Open-angle glaucoma of left eye caused by steroid |
| SNOMED CT | 15993751000119108 | Open-angle glaucoma of bilateral eyes caused by steroid |
| SNOMED CT | 15996831000119101 | Glaucoma caused by Onchocerca volvulus |
| SNOMED CT | 193548006 | Steroid-induced glaucoma glaucomatous stage |
| SNOMED CT | 193552006 | Glaucoma due to chamber angle anomaly |
| SNOMED CT | 193553001 | Glaucoma due to iris anomaly |
| SNOMED CT | 193555008 | Glaucoma due to systemic syndrome |
| SNOMED CT | 193561006 | Secondary angle-closure glaucoma with pupillary block |
| SNOMED CT | 193562004 | Glaucoma due to ocular vascular disorder |
| SNOMED CT | 204113001 | Congenital glaucoma |
| SNOMED CT | 206248004 | Traumatic glaucoma due to birth trauma |
| SNOMED CT | 232081005 | Iatrogenic glaucoma |
| SNOMED CT | 232082003 | Iatrogenic angle-closure glaucoma |
| SNOMED CT | 232083008 | Glaucoma and corneal anomaly |
| SNOMED CT | 232086000 | Neovascular glaucoma |
| SNOMED CT | 232087009 | Glaucoma with intraocular hemorrhage |
| SNOMED CT | 232088004 | Ghost cell glaucoma |
| SNOMED CT | 232090003 | Glaucoma following surgery |
| SNOMED CT | 314017009 | Acute-on-chronic glaucoma |
| SNOMED CT | 314033007 | Uveitic glaucoma |

Continued on next page

**Table9 – continued from previous page**

| CodeSystem | ConceptCode | ConceptName |
|---|---|---|
| SNOMED CT | 314784002 | Secondary angle-closure glaucoma - synechial |
| SNOMED CT | 370504007 | Aqueous humor misdirect |
| SNOMED CT | 392030001 | Hemolytic glaucoma |
| SNOMED CT | 392288006 | Primary angle-closure glaucoma |
| SNOMED CT | 392291006 | Angle-closure glaucoma |
| SNOMED CT | 392300000 | Phacomorphic glaucoma |
| SNOMED CT | 392352004 | Angle recession glaucoma |
| SNOMED CT | 404648005 | Lens particle glaucoma |
| SNOMED CT | 415176004 | Primary congenital glaucoma |
| SNOMED CT | 37155002 | Glaucoma associated with ocular inflammation |
| SNOMED CT | 45623002 | Glaucoma associated with anterior segment anomaly |
| SNOMED CT | 46168003 | Pigmentary glaucoma |
| SNOMED CT | 50485007 | Low tension glaucoma |
| SNOMED CT | 53667005 | Glaucoma associated with systemic syndromes |
| SNOMED CT | 65460003 | Intermittent angle-closure glaucoma |
| SNOMED CT | 66725002 | Glaucoma due to perforating injury |
| SNOMED CT | 66747002 | Glaucoma associated with ocular disorder |
| SNOMED CT | 68241007 | Glaucoma associated with ocular trauma |
| SNOMED CT | 77075001 | Primary open angle glaucoma |
| SNOMED CT | 444863008 | Anatomical narrow angle glaucoma with borderline intraocular pressure |
| SNOMED CT | 698840003 | Neovascular glaucoma due to hyphema |
| SNOMED CT | 713457002 | Neovascular glaucoma due to diabetes mellitus |
| SNOMED CT | 715144004 | Glaucoma caused by silicone oil |
| SNOMED CT | 716166002 | Microcornea with glaucoma and absent frontal sinus syndrome |
| SNOMED CT | 721898008 | Open angle glaucoma of bilateral eyes caused by corticosteroid |
| SNOMED CT | 722321001 | Open angle glaucoma of left eye caused by corticosteroid |
| SNOMED CT | 722329004 | Open angle glaucoma of right eye caused by corticosteroid |
| SNOMED CT | 267625001 | Glaucoma due to ocular tumor or cyst |
| SNOMED CT | 275477002 | Glaucoma due to ocular cyst |
| SNOMED CT | 84333006 | Phacogenic glaucoma |
| SNOMED CT | 84494001 | Open-angle glaucoma |
| SNOMED CT | 89215000 | Postoperative angle-closure glaucoma |
| SNOMED CT | 92829008 | Glaucoma due to combination of mechanisms |
| SNOMED CT | 95213001 | Primary glaucoma due to combination of mechanisms |
| SNOMED CT | 95250000 | Secondary glaucoma due to combination mechanisms |
| SNOMED CT | 95717004 | Secondary glaucoma |
| SNOMED CT | 111513000 | Advanced open-angle glaucoma |
| SNOMED CT | 111514006 | Pseudoexfoliation glaucoma |
| SNOMED CT | 1207009 | Glaucomatous atrophy of optic disc |
| SNOMED CT | 29538005 | Glaucomatocyclitic crisis |

Continued on next page

**Table9 – continued from previous page**

| CodeSystem | ConceptCode | ConceptName |
|---|---|---|
| SNOMED CT | 35472004 | Open angle with borderline intraocular pressure |
| SNOMED CT | 1231271005 | Glaucoma due to and following retinopathy of prematurity |
| SNOMED CT | 1231270006 | Glaucoma following ocular onchocerciasis |
| SNOMED CT | 334961000119105 | Right secondary angle-closure glaucoma due to pupil block |
| SNOMED CT | 349231000119108 | Bilateral aqueous humor misdirection of eyes |
| SNOMED CT | 1231243004 | High pressure primary open-angle glaucoma |
| SNOMED CT | 733086003 | Pseudoprogeria syndrome |
| SNOMED CT | 733116005 | Aniridia, renal agenesis, psychomotor retardation syndrome |
| SNOMED CT | 733455003 | Spastic paraplegia, glaucoma, intellectual disability syndrome |
| SNOMED CT | 736838001 | Cupping of optic disc due to open angle glaucoma |
| SNOMED CT | 737006004 | Cupping of optic discs of bilateral eyes due to open-angle glaucoma |
| SNOMED CT | 770776002 | Borderline angle-closure glaucoma of right eye |
| SNOMED CT | 770777006 | Borderline angle-closure glaucoma of left eye |
| SNOMED CT | 783246000 | Megalocornea, spherophakia, secondary glaucoma syndrome |
| SNOMED CT | 788945006 | Glaucoma caused by contact lens |
| SNOMED CT | 788946007 | Glaucoma due to eye inflammation |
| SNOMED CT | 788947003 | Glaucoma due to intraocular neoplasm |
| SNOMED CT | 788948008 | Glaucoma due to retinal detachment |
| SNOMED CT | 788995007 | Normal pressure primary open-angle glaucoma |
| SNOMED CT | 833278008 | Glaucoma suspect |
| SNOMED CT | 860798008 | Glaucoma due to diabetes mellitus |
| SNOMED CT | 331471000119105 | Uveitic glaucoma of right eye |
| SNOMED CT | 332871000119103 | Pseudoexfoliation glaucoma of right eye |
| SNOMED CT | 333111000119107 | Pigmentary glaucoma of right eye |
| SNOMED CT | 333151000119108 | Phacolytic glaucoma of right eye |
| SNOMED CT | 334321000119101 | Low tension glaucoma of right eye |
| SNOMED CT | 336031000119105 | Anatomic narrow angle glaucoma of right eye |
| SNOMED CT | 337081000119109 | Uveitic glaucoma of left eye |
| SNOMED CT | 338481000119100 | Pseudoexfoliation glaucoma of left eye |
| SNOMED CT | 338721000119108 | Pigmentary glaucoma of left eye |
| SNOMED CT | 338761000119103 | Phacolytic glaucoma of left eye |
| SNOMED CT | 339921000119104 | Low tension glaucoma of left eye |
| SNOMED CT | 341641000119100 | Anatomic narrow angle glaucoma of left eye |
| SNOMED CT | 343161000119109 | Bilateral uveitic glaucoma of eyes |
| SNOMED CT | 344251000119103 | Bilateral pseudoexfoliation glaucoma of eyes |
| SNOMED CT | 344491000119107 | Bilateral pigmentary glaucoma of eyes |
| SNOMED CT | 345291000119109 | Bilateral low tension glaucoma of eyes |
| SNOMED CT | 346861000119107 | Bilateral eye anatomic narrow angle glaucoma |
| SNOMED CT | 348361000119103 | Aphakic glaucoma of right eye |
| SNOMED CT | 348811000119108 | Aphakic glaucoma of left eye |

Continued on next page

**Table9 – continued from previous page**

| CodeSystem | ConceptCode | ConceptName |
|---|---|---|
| SNOMED CT | 349311000119106 | Bilateral aphakic glaucoma |
| SNOMED CT | 678401000119102 | Bilateral glaucoma of eyes caused by drug |
| SNOMED CT | 678411000119104 | Glaucoma of left eye caused by drug |
| SNOMED CT | 678421000119106 | Glaucoma of right eye caused by drug |
| SNOMED CT | 678471000119107 | Aqueous humor misdirection of left eye |
| SNOMED CT | 678481000119105 | Aqueous humor misdirection of right eye |
| SNOMED CT | 15673041000119101 | Congenital glaucoma of right eye |
| SNOMED CT | 15673081000119106 | Congenital glaucoma of left eye |
| SNOMED CT | 15679721000119107 | Uveitis-hyphema-glaucoma syndrome of right eye |
| SNOMED CT | 15679761000119102 | Uveitis-hyphema-glaucoma syndrome of left eye |
| SNOMED CT | 15738761000119100 | Bilateral eye borderline angle-closure glaucoma |
| SNOMED CT | 15739801000119100 | Cupping of left optic disc due to open-angle glaucoma of left eye |
| SNOMED CT | 15739881000119108 | Cupping of right optic disc due to open-angle glaucoma of right eye |
| SNOMED CT | 193531003 | Borderline glaucoma |
| SNOMED CT | 193533000 | Open-angle glaucoma - borderline |
| SNOMED CT | 193534006 | Angle-closure glaucoma - borderline |
| SNOMED CT | 193556009 | Glaucoma in endocrine, nutritional and metabolic diseases |
| SNOMED CT | 232080006 | Plateau iris |
| SNOMED CT | 1285643003 | Neovascular glaucoma due to central retinal vein occlusion |
| SNOMED CT | 392029006 | Uveitis-glaucoma-hyphema syndrome |
| SNOMED CT | 404634005 | Glaucoma associated with iridocorneal endothelial syndrome |
| SNOMED CT | 415501002 | Schwartz ocular syndrome |
| SNOMED CT | 71111008 | Glaucoma of childhood |
| SNOMED CT | 718851007 | Cataract glaucoma syndrome |
| SNOMED CT | 719976001 | Glaucoma and sleep apnea syndrome |
| SNOMED CT | 721974000 | Lowry MacLean syndrome |
| SNOMED CT | 722450007 | GEMSS syndrome |
| SNOMED CT | 723503006 | Retinal degeneration, nanophthalmos, glaucoma syndrome |
| SNOMED CT | 302895007 | Steroid-induced glaucoma - borderline |
| SNOMED CT | 340571000119101 | Left secondary angle-closure glaucoma due to pupil block |
| SNOMED CT | 1196923000 | Neovascular glaucoma due to diabetes mellitus type 1 |
| SNOMED CT | 1003425007 | Glaucoma due to Lowe syndrome |
| SNOMED CT | 1003428009 | Glaucoma due to congenital anomaly of eye |
| SNOMED CT | 1003657004 | Congenital glaucoma of left eye |
| SNOMED CT | 15995551000119104 | Bilateral glaucoma of eyes due to plateau iris |
| SNOMED CT | 334761000119107 | Intermittent angle-closure glaucoma of right eye |
| SNOMED CT | 340361000119101 | Intermittent angle-closure glaucoma of left eye |
| SNOMED CT | 345701000119106 | Bilateral intermittent angle-closure glaucoma |
| SNOMED CT | 678431000119109 | Bilateral glaucoma of eyes due to increased episcleral venous pressure |

Continued on next page

**Table9 – continued from previous page**

| CodeSystem | ConceptCode | ConceptName |
|---|---|---|
| SNOMED CT | 678441000119100 | Glaucoma of left eye due to increased episcleral venous pressure |
| SNOMED CT | 678451000119103 | Glaucoma of right eye due to increased episcleral venous pressure |
| SNOMED CT | 890423003 | Secondary childhood glaucoma following congenital cataract surgery |
| SNOMED CT | 897585008 | Bilateral primary congenital glaucoma |
| SNOMED CT | 1137633000 | Secondary angle-closure glaucoma due to iridocorneal endothelial syndrome |
| SNOMED CT | 345881000119107 | Bilateral secondary angle-closure glaucoma due to pupil block |
| SNOMED CT | 93435005 | Drug-induced glaucoma |
| SNOMED CT | 1196922005 | Neovascular glaucoma due to diabetes mellitus type 2 |

##### 0.4.7 Heart Failure

Table 10: Concept codes used to identify heart failure conditions.

| CodeSystem | ConceptCode | ConceptName |
|---|---|---|
| ICD10CM | I11.0 | Hypertensive heart disease with heart failure |
| ICD10CM | I13.0 | Hypertensive heart and chronic kidney disease with heart failure and stage 1 through stage 4 chronic kidney disease, or unspecified chronic kidney disease |
| ICD10CM | I13.2 | Hypertensive heart and chronic kidney disease with heart failure and with stage 5 chronic kidney disease, or end stage renal disease |
| ICD10CM | I50.1 | Left ventricular failure, unspecified |
| ICD10CM | I50.20 | Unspecified systolic (congestive) heart failure |
| ICD10CM | I50.21 | Acute systolic (congestive) heart failure |
| ICD10CM | I50.22 | Chronic systolic (congestive) heart failure |
| ICD10CM | I50.23 | Acute on chronic systolic (congestive) heart failure |
| ICD10CM | I50.30 | Unspecified diastolic (congestive) heart failure |
| ICD10CM | I50.31 | Acute diastolic (congestive) heart failure |
| ICD10CM | I50.32 | Chronic diastolic (congestive) heart failure |
| ICD10CM | I50.33 | Acute on chronic diastolic (congestive) heart failure |
| ICD10CM | I50.40 | Unspecified combined systolic (congestive) and diastolic (congestive) heart failure |
| ICD10CM | I50.41 | Acute combined systolic (congestive) and diastolic (congestive) heart failure |
| ICD10CM | I50.42 | Chronic combined systolic (congestive) and diastolic (congestive) heart failure |

Continued on next page

**Table10 – continued from previous page**

| CodeSystem | ConceptCode | ConceptName |
|---|---|---|
| ICD10CM | I50.43 | Acute on chronic combined systolic (congestive) and diastolic (congestive) heart failure |
| ICD10CM | I50.81 | Right heart failure |
| ICD10CM | I50.82 | Biventricular heart failure |
| ICD10CM | I50.83 | High output heart failure |
| ICD10CM | I50.84 | End stage heart failure |
| ICD10CM | I50.89 | Other heart failure |
| ICD10CM | I50.9 | Heart failure, unspecified |
| ICD10CM | I50.810 | Right heart failure, unspecified |
| ICD10CM | I50.811 | Acute right heart failure |
| ICD10CM | I50.812 | Chronic right heart failure |
| ICD10CM | I50.813 | Acute on chronic right heart failure |
| ICD10CM | I50.814 | Right heart failure due to left heart failure |
| SNOMED CT | 364006 | Acute left-sided heart failure |
| SNOMED CT | 5148006 | Hypertensive heart disease with congestive heart failure |
| SNOMED CT | 5375005 | Chronic left-sided congestive heart failure |
| SNOMED CT | 10091002 | High output heart failure |
| SNOMED CT | 10633002 | Acute congestive heart failure |
| SNOMED CT | 25544003 | Low output heart failure |
| SNOMED CT | 871617000 | Low output heart failure due to and following Fontan operation |
| SNOMED CT | 7411000175102 | Chronic heart failure co-occurrent with normal ejection fraction |
| SNOMED CT | 15781000119107 | Hypertensive heart AND chronic kidney disease with congestive heart failure |
| SNOMED CT | 23341000119109 | Congestive heart failure with right heart failure |
| SNOMED CT | 67431000119105 | Congestive heart failure stage D |
| SNOMED CT | 67441000119101 | Congestive heart failure stage C |
| SNOMED CT | 72481000119103 | Congestive heart failure as early postoperative complication |
| SNOMED CT | 101281000119107 | Congestive heart failure due to cardiomyopathy |
| SNOMED CT | 120851000119104 | Systolic heart failure stage D |
| SNOMED CT | 120861000119102 | Systolic heart failure stage C |
| SNOMED CT | 120871000119108 | Systolic heart failure stage B |
| SNOMED CT | 120881000119106 | Diastolic heart failure stage D |
| SNOMED CT | 120891000119109 | Diastolic heart failure stage C |
| SNOMED CT | 120901000119108 | Diastolic heart failure stage B |
| SNOMED CT | 153931000119109 | Acute combined systolic and diastolic heart failure |
| SNOMED CT | 153941000119100 | Chronic combined systolic and diastolic heart failure |
| SNOMED CT | 153951000119103 | Acute on chronic combined systolic and diastolic heart failure |
| SNOMED CT | 15629541000119106 | Congestive heart failure stage C due to ischemic cardiomyopathy |

Continued on next page

**Table10 – continued from previous page**

| CodeSystem | ConceptCode | ConceptName |
|---|---|---|
| SNOMED CT | 15629591000119103 | Congestive heart failure stage B due to ischaemic cardiomyopathy |
| SNOMED CT | 15629641000119107 | Systolic heart failure stage B due to ischemic cardiomyopathy |
| SNOMED CT | 15629741000119102 | Systolic heart failure stage C due to ischemic cardiomyopathy |
| SNOMED CT | 15964701000119109 | Saddle embolus of pulmonary artery with acute cor pulmonale |
| SNOMED CT | 194767001 | Benign hypertensive heart disease with congestive cardiac failure |
| SNOMED CT | 194779001 | Hypertensive heart and renal disease with (congestive) heart failure |
| SNOMED CT | 194781004 | Hypertensive heart and renal disease with both (congestive) heart failure and renal failure |
| SNOMED CT | 195111005 | Decompensated cardiac failure |
| SNOMED CT | 195112003 | Compensated cardiac failure |
| SNOMED CT | 195114002 | Acute left ventricular failure |
| SNOMED CT | 206586007 | Congenital cardiac failure |
| SNOMED CT | 233924009 | Heart failure as a complication of care |
| SNOMED CT | 314206003 | Refractory heart failure |
| SNOMED CT | 410431009 | Cardiorespiratory failure |
| SNOMED CT | 417996009 | Systolic heart failure |
| SNOMED CT | 418304008 | Diastolic heart failure |
| SNOMED CT | 424404003 | Decompensated chronic heart failure |
| SNOMED CT | 426263006 | Congestive heart failure due to left ventricular systolic dysfunction |
| SNOMED CT | 426611007 | Congestive heart failure due to valvular disease |
| SNOMED CT | 42343007 | Congestive heart failure |
| SNOMED CT | 43736008 | Rheumatic left ventricular failure |
| SNOMED CT | 44088000 | Low cardiac output syndrome |
| SNOMED CT | 44313006 | Right heart failure secondary to left heart failure |
| SNOMED CT | 46113002 | Hypertensive heart failure |
| SNOMED CT | 48447003 | Chronic heart failure |
| SNOMED CT | 56675007 | Acute heart failure |
| SNOMED CT | 74960003 | Acute left-sided congestive heart failure |
| SNOMED CT | 82523003 | Congestive rheumatic heart failure |
| SNOMED CT | 441481004 | Chronic systolic heart failure |
| SNOMED CT | 441530006 | Chronic diastolic heart failure |
| SNOMED CT | 443253003 | Acute on chronic systolic heart failure |
| SNOMED CT | 443254009 | Acute systolic heart failure |
| SNOMED CT | 443343001 | Acute diastolic heart failure |
| SNOMED CT | 443344007 | Acute on chronic diastolic heart failure |
| SNOMED CT | 446221000 | Heart failure with normal ejection fraction |

Continued on next page

**Table10 – continued from previous page**

| CodeSystem | ConceptCode | ConceptName |
|---|---|---|
| SNOMED CT | 471880001 | Heart failure due to end stage congenital heart disease |
| SNOMED CT | 698296002 | Acute exacerbation of chronic congestive heart failure |
| SNOMED CT | 698594003 | Symptomatic congestive heart failure |
| SNOMED CT | 703272007 | Heart failure with reduced ejection fraction |
| SNOMED CT | 703273002 | Heart failure with reduced ejection fraction due to coronary artery disease |
| SNOMED CT | 703274008 | Heart failure with reduced ejection fraction due to myocarditis |
| SNOMED CT | 703275009 | Heart failure with reduced ejection fraction due to cardiomyopathy |
| SNOMED CT | 703276005 | Heart failure with reduced ejection fraction due to heart valve disease |
| SNOMED CT | 717840005 | Congestive heart failure stage B |
| SNOMED CT | 83105008 | Malignant hypertensive heart disease with congestive heart failure |
| SNOMED CT | 84114007 | Heart failure |
| SNOMED CT | 85232009 | Left heart failure |
| SNOMED CT | 88805009 | Chronic congestive heart failure |
| SNOMED CT | 90727007 | Pleural effusion due to congestive heart failure |
| SNOMED CT | 92506005 | Biventricular congestive heart failure |
| SNOMED CT | 111283005 | Chronic left-sided heart failure |
| SNOMED CT | 1204460007 | Myocardial dysfunction with sepsis |
| SNOMED CT | 5053004 | Cardiac insufficiency due to prosthesis |
| SNOMED CT | 10335000 | Chronic right-sided heart failure |
| SNOMED CT | 16253001 | Dilated peripartum cardiomyopathy |
| SNOMED CT | 24841007 | Cardiorespiratory failure during AND/OR resulting from a procedure |
| SNOMED CT | 33644002 | Postvalvulotomy syndrome |
| SNOMED CT | 1204206001 | Left ventricular failure with normal ejection fraction due to cardiomyopathy |
| SNOMED CT | 1204203009 | Left ventricular failure with normal ejection fraction due to coronary artery disease |
| SNOMED CT | 1204204003 | Left ventricular failure with normal ejection fraction due to myocarditis |
| SNOMED CT | 1204200007 | Left ventricular failure with normal ejection fraction due to valvular heart disease |
| SNOMED CT | 1204462004 | Left ventricular failure with sepsis |
| SNOMED CT | 1208846006 | Right ventricular failure due to pulmonary disease |
| SNOMED CT | 1208848007 | Right ventricular failure due to pulmonary vascular disease |
| SNOMED CT | 1208850004 | Right ventricular failure due to right ventricular infarction |
| SNOMED CT | 1208843003 | Right ventricular failure due to valvular heart disease |
| SNOMED CT | 1204468000 | Right ventricular failure with sepsis |
| SNOMED CT | 788950000 | Heart failure with mid range ejection fraction |

Continued on next page

**Table10 – continued from previous page**

| CodeSystem | ConceptCode | ConceptName |
|---|---|---|
| SNOMED CT | 7371000175103 | Reduced ejection fraction co-occurrent and due to acute on chronic heart failure |
| SNOMED CT | 7381000175100 | Reduced ejection fraction co-occurrent and due to chronic heart failure |
| SNOMED CT | 7391000175102 | Reduced ejection fraction co-occurrent and due to acute heart failure |
| SNOMED CT | 7401000175100 | Acute on chronic heart failure co-occurrent with normal ejection fraction |
| SNOMED CT | 7421000175106 | Acute heart failure co-occurrent with normal ejection fraction |
| SNOMED CT | 96311000119109 | Exacerbation of congestive heart failure |
| SNOMED CT | 285211000119102 | Congestive heart failure as post-operative complication of cardiac surgery |
| SNOMED CT | 285221000119109 | Congestive heart failure as post-operative complication of non-cardiac surgery |
| SNOMED CT | 16838951000119100 | Acute on chronic right-sided congestive heart failure |
| SNOMED CT | 13839000 | Right ventricular obstruction - failure syndrome |
| SNOMED CT | 78862003 | Cardiopathia nigra |
| SNOMED CT | 1264003007 | Postcardiotomy acute right ventricular failure |
| SNOMED CT | 359617009 | Acute right-sided heart failure |
| SNOMED CT | 367363000 | Right ventricular failure |
| SNOMED CT | 426012001 | Right heart failure due to pulmonary hypertension |
| SNOMED CT | 45650007 | Kyphoscoliotic heart disease |
| SNOMED CT | 49584005 | Acute cor pulmonale |
| SNOMED CT | 55565007 | Cardiac failure after obstetrical surgery AND/OR other procedure including delivery |
| SNOMED CT | 60856006 | Cardiac insufficiency following cardiac surgery |
| SNOMED CT | 62377009 | Peripartum cardiomyopathy in puerperium |
| SNOMED CT | 66989003 | Chronic right-sided congestive heart failure |
| SNOMED CT | 71892000 | Cardiac asthma |
| SNOMED CT | 79955004 | Chronic cardiopulmonary disease |
| SNOMED CT | 80479009 | Acute right-sided congestive heart failure |
| SNOMED CT | 445236007 | Cardiorenal syndrome |
| SNOMED CT | 462172006 | Fetal heart failure |
| SNOMED CT | 462174007 | Fetal heart failure with myocardial hypertrophy |
| SNOMED CT | 462175008 | Fetal heart failure with redistribution of cardiac output |
| SNOMED CT | 609507007 | Induced termination of pregnancy complicated by cardiac failure |
| SNOMED CT | 704242009 | Fetal heart failure due to extracardiac disease |
| SNOMED CT | 722095005 | Acute kidney injury due to circulatory failure |
| SNOMED CT | 722919003 | Neonatal cardiac failure due to decreased left ventricular output |
| SNOMED CT | 724550005 | Neonatal cardiac failure due to pulmonary overperfusion |

Continued on next page

**Table10 – continued from previous page**

| CodeSystem | ConceptCode | ConceptName |
|---|---|---|
| SNOMED CT | 276514007 | Neonatal cardiac failure |
| SNOMED CT | 277638005 | Sepsis-associated left ventricular failure |
| SNOMED CT | 277639002 | Sepsis-associated right ventricular failure |
| SNOMED CT | 1010444007 | Legal abortion complicated by cardiac failure |
| SNOMED CT | 1010447000 | Illegal abortion complicated by cardiac failure |
| SNOMED CT | 15964661000119102 | Acute cor pulmonale due to septic pulmonary embolism |
| SNOMED CT | 898208007 | Heart failure due to thyrotoxicosis |
| SNOMED CT | 83291003 | Right heart failure due to pulmonary hypertension |
| SNOMED CT | 89819002 | Cardiac insufficiency during AND/OR resulting from a procedure |

##### 0.4.8 Hyperlipidemia

Table 11: Concept codes used to identify hyperlipidemia conditions.

| CodeSystem | ConceptCode | ConceptName |
|---|---|---|
| ICD10CM | E78.0 | Pure hypercholesterolemia |
| ICD10CM | E78.00 | Pure hypercholesterolemia, unspecified |
| ICD10CM | E78.01 | Familial hypercholesterolemia |
| ICD10CM | E78.1 | Pure hyperglyceridemia |
| ICD10CM | E78.2 | Mixed hyperlipidemia |
| ICD10CM | E78.3 | Hyperchylomicronemia |
| ICD10CM | E78.4 | Other hyperlipidemia |
| ICD10CM | E78.41 | Elevated Lipoprotein(a) |
| ICD10CM | E78.49 | Other hyperlipidemia |
| ICD10CM | E78.5 | Hyperlipidemia, unspecified |
| SNOMED CT | 13644009 | Hypercholesterolemia |
| SNOMED CT | 33513003 | Familial apoC-II deficiency |
| SNOMED CT | 34349009 | Familial type 5 hyperlipoproteinemia |
| SNOMED CT | 34528009 | Familial hypertriglyceridemia |
| SNOMED CT | 773649005 | Transient infantile hypertriglyceridemia and hepatosteatosis |
| SNOMED CT | 773726000 | Hypercholesterolemia due to cholesterol 7alpha-hydroxylase deficiency |
| SNOMED CT | 701000119103 | Mixed hyperlipidemia due to type 2 diabetes mellitus |
| SNOMED CT | 1571000119104 | Mixed hyperlipidemia due to type 1 diabetes mellitus |
| SNOMED CT | 15771000119109 | Familial hyperalphalipoproteinemia |
| SNOMED CT | 114831000119107 | Hyperlipidemia caused by steroid |
| SNOMED CT | 137931000119102 | Hyperlipidemia due to type 2 diabetes mellitus |
| SNOMED CT | 137941000119106 | Hyperlipidemia due to type 1 diabetes mellitus |

Continued on next page

**Table11 – continued from previous page**

| CodeSystem | ConceptCode | ConceptName |
|---|---|---|
| SNOMED CT | 403829002 | Familial hypercholesterolemia due to heterozygous low density lipoprotein receptor mutation |
| SNOMED CT | 403830007 | Familial hypercholesterolemia due to homozygous low density lipoprotein receptor mutation |
| SNOMED CT | 767133009 | Compound heterozygous familial hypercholesterolemia |
| SNOMED CT | 190774002 | Hyperlipidemia, group A |
| SNOMED CT | 238040008 | Familial combined hyperlipidemia |
| SNOMED CT | 238076009 | Primary hypercholesterolemia |
| SNOMED CT | 238077000 | Polygenic hypercholesterolemia |
| SNOMED CT | 238078005 | Familial hypercholesterolemia - homozygous |
| SNOMED CT | 238079002 | Familial hypercholesterolemia - heterozygous |
| SNOMED CT | 238080004 | Hyperalphalipoproteinemia |
| SNOMED CT | 238081000 | Familial defective apolipoprotein B-100 |
| SNOMED CT | 238082007 | Secondary hypercholesterolemia |
| SNOMED CT | 238083002 | Primary hypertriglyceridemia |
| SNOMED CT | 238084008 | Very low density lipoprotinemia |
| SNOMED CT | 238085009 | Fredrickson type IV hyperlipoproteinemia |
| SNOMED CT | 238087001 | Secondary hypertriglyceridemia |
| SNOMED CT | 238088006 | Primary combined hyperlipidemia |
| SNOMED CT | 238089003 | Secondary combined hyperlipidemia |
| SNOMED CT | 397915002 | Fredrickson type IIa hyperlipoproteinemia |
| SNOMED CT | 398036000 | Familial hypercholesterolemia |
| SNOMED CT | 402473001 | Sporadic primary hypertriglyceridemia |
| SNOMED CT | 402474007 | Primary polygenic type IIb combined hyperlipidemia |
| SNOMED CT | 402475008 | Primary acquired chylomicronemia |
| SNOMED CT | 402725005 | Hyperlipidemia with lipid deposition in skin |
| SNOMED CT | 402726006 | Primary chylomicronemia |
| SNOMED CT | 402727002 | Secondary hyperlipidemia |
| SNOMED CT | 402785008 | Primary genetic hyperlipidemia |
| SNOMED CT | 402786009 | Chylomicronemia syndrome |
| SNOMED CT | 402787000 | Primary genetic mixed hyperlipidemia |
| SNOMED CT | 403827000 | Familial lipoprotein lipase deficiency with type I phenotype |
| SNOMED CT | 403828005 | Familial lipoprotein lipase deficiency with type V phenotype |
| SNOMED CT | 403831006 | Familial Combined Hypercholesterolaemia |
| SNOMED CT | 426161002 | Chemically induced hyperlipidemia |
| SNOMED CT | 55822004 | Hyperlipidemia |
| SNOMED CT | 445261005 | Posttransplant hyperlipidemia |
| SNOMED CT | 267432004 | Pure hypercholesterolemia |
| SNOMED CT | 267433009 | Pure hyperglyceridemia |
| SNOMED CT | 267434003 | Mixed hyperlipidemia |
| SNOMED CT | 267435002 | Familial hyperchylomicronemia |
| SNOMED CT | 299465007 | Familial multiple lipoprotein-type hyperlipidemia |
| SNOMED CT | 302870006 | Hypertriglyceridemia |

Continued on next page

**Table11 – continued from previous page**

| CodeSystem | ConceptCode | ConceptName |
|---|---|---|
| SNOMED CT | 129589009 | Endogenous hyperlipidemia |
| SNOMED CT | 129590000 | Exogenous hyperlipidemia |
| SNOMED CT | 129591001 | Mixed hypercholesterolemia and hypertriglyceridemia |
| SNOMED CT | 1197489003 | Familial chylomicronemia syndrome |
| SNOMED CT | 1208738002 | TMEM199 congenital disorder of glycosylation |
| SNOMED CT | 275598004 | Familial hyperlipoproteinemia, type I |

##### 0.4.9 Hypertension

Table 12: Concept codes used to identify hypertension conditions.

| CodeSystem | ConceptCode | ConceptName |
|---|---|---|
| ICD10CM | I10 | Essential (primary) hypertension |
| ICD10CM | I11.0 | Hypertensive heart disease with heart failure |
| ICD10CM | I11.9 | Hypertensive heart disease without heart failure |
| ICD10CM | I12.0 | Hypertensive chronic kidney disease with stage 5 chronic kidney disease or end stage renal disease |
| ICD10CM | I12.9 | Hypertensive chronic kidney disease with stage 1 through stage 4 chronic kidney disease, or unspecified chronic kidney disease |
| ICD10CM | I13.0 | Hypertensive heart and chronic kidney disease with heart failure and stage 1 through stage 4 chronic kidney disease, or unspecified chronic kidney disease |
| ICD10CM | I13.1 | Hypertensive heart and chronic kidney disease without heart failure |
| ICD10CM | I13.10 | Hypertensive heart and chronic kidney disease without heart failure, with stage 1 through stage 4 chronic kidney disease, or unspecified chronic kidney disease |
| ICD10CM | I13.11 | Hypertensive heart and chronic kidney disease without heart failure, with stage 5 chronic kidney disease, or end stage renal disease |
| ICD10CM | I13.2 | Hypertensive heart and chronic kidney disease with heart failure and with stage 5 chronic kidney disease, or end stage renal disease |
| ICD10CM | I15 | Secondary hypertension |
| ICD10CM | I15.0 | Renovascular hypertension |
| ICD10CM | I15.1 | Hypertension secondary to other renal disorders |
| ICD10CM | I15.2 | Hypertension secondary to endocrine disorders |
| ICD10CM | I15.8 | Other secondary hypertension |
| ICD10CM | I15.9 | Secondary hypertension, unspecified |
| ICD10CM | I16 | Hypertensive crisis |

Continued on next page

**Table12 – continued from previous page**

| CodeSystem | ConceptCode | ConceptName |
|---|---|---|
| ICD10CM | I16.0 | Hypertensive urgency |
| ICD10CM | I16.1 | Hypertensive emergency |
| ICD10CM | I16.9 | Hypertensive crisis, unspecified |
| ICD9CM | 401.0 | Malignant essential hypertension |
| ICD9CM | 401.1 | Benign essential hypertension |
| ICD9CM | 401.9 | Unspecified essential hypertension |
| ICD9CM | 402.00 | Malignant hypertensive heart disease without heart failure |
| ICD9CM | 402.01 | Malignant hypertensive heart disease with heart failure |
| ICD9CM | 402.10 | Benign hypertensive heart disease without heart failure |
| ICD9CM | 402.11 | Benign hypertensive heart disease with heart failure |
| ICD9CM | 402.90 | Unspecified hypertensive heart disease without heart failure |
| ICD9CM | 402.91 | Unspecified hypertensive heart disease with heart failure |
| ICD9CM | 403.00 | Hypertensive chronic kidney disease, malignant, with chronic kidney disease stage I through stage IV, or unspecified |
| ICD9CM | 403.01 | Hypertensive chronic kidney disease, malignant, with chronic kidney disease stage V or end stage renal disease |
| ICD9CM | 403.10 | Hypertensive chronic kidney disease, benign, with chronic kidney disease stage I through stage IV, or unspecified |
| ICD9CM | 403.11 | Hypertensive chronic kidney disease, benign, with chronic kidney disease stage V or end stage renal disease |
| ICD9CM | 403.90 | Hypertensive chronic kidney disease, unspecified, with chronic kidney disease stage I through stage IV, or unspecified |
| ICD9CM | 403.91 | Hypertensive chronic kidney disease, unspecified, with chronic kidney disease stage V or end stage renal disease |
| ICD9CM | 404.00 | Hypertensive heart and chronic kidney disease, malignant, without heart failure and with chronic kidney disease stage I through stage IV, or unspecified |
| ICD9CM | 404.01 | Hypertensive heart and chronic kidney disease, malignant, with heart failure and with chronic kidney disease stage I through stage IV, or unspecified |
| ICD9CM | 404.02 | Hypertensive heart and chronic kidney disease, malignant, without heart failure and with chronic kidney disease stage V or end stage renal disease |
| ICD9CM | 404.03 | Hypertensive heart and chronic kidney disease, malignant, with heart failure and with chronic kidney disease stage V or end stage renal disease |
| ICD9CM | 404.10 | Hypertensive heart and chronic kidney disease, benign, without heart failure and with chronic kidney disease stage I through stage IV, or unspecified |

Continued on next page

**Table12 – continued from previous page**

| CodeSystem | ConceptCode | ConceptName |
|---|---|---|
| ICD9CM | 404.11 | Hypertensive heart and chronic kidney disease, benign, with heart failure and with chronic kidney disease stage I through stage IV, or unspecified |
| ICD9CM | 404.12 | Hypertensive heart and chronic kidney disease, benign, without heart failure and with chronic kidney disease stage V or end stage renal disease |
| ICD9CM | 404.13 | Hypertensive heart and chronic kidney disease, benign, with heart failure and chronic kidney disease stage V or end stage renal disease |
| ICD9CM | 404.90 | Hypertensive heart and chronic kidney disease, unspecified, without heart failure and with chronic kidney disease stage I through stage IV, or unspecified |
| ICD9CM | 404.91 | Hypertensive heart and chronic kidney disease, unspecified, with heart failure and with chronic kidney disease stage I through stage IV, or unspecified |
| ICD9CM | 404.92 | Hypertensive heart and chronic kidney disease, unspecified, without heart failure and with chronic kidney disease stage V or end stage renal disease |
| ICD9CM | 404.93 | Hypertensive heart and chronic kidney disease, unspecified, with heart failure and chronic kidney disease stage V or end stage renal disease |
| ICD9CM | 405.01 | Malignant renovascular hypertension |
| ICD9CM | 405.09 | Other malignant secondary hypertension |
| ICD9CM | 405.11 | Benign renovascular hypertension |
| ICD9CM | 405.19 | Other benign secondary hypertension |
| ICD9CM | 405.91 | Unspecified renovascular hypertension |
| ICD9CM | 405.99 | Other unspecified secondary hypertension |
| SNOMED CT | 1201005 | Benign essential hypertension |
| SNOMED CT | 10725009 | Benign hypertension |
| SNOMED CT | 14973001 | Renal sclerosis with hypertension |
| SNOMED CT | 26078007 | Hypertension secondary to renal disease complicating AND/OR reason for care during childbirth |
| SNOMED CT | 28119000 | Renal hypertension |
| SNOMED CT | 31992008 | Secondary hypertension |
| SNOMED CT | 762463000 | Diastolic hypertension and systolic hypertension |
| SNOMED CT | 284981000119102 | Chronic kidney disease stage 2 due to benign hypertension |
| SNOMED CT | 284991000119104 | Chronic kidney disease stage 3 due to benign hypertension |
| SNOMED CT | 461301000124109 | Resistant hypertensive disorder |
| SNOMED CT | 1078301000112109 | Multiple drug intolerant hypertension |
| SNOMED CT | 194783001 | Malignant secondary renovascular hypertension |
| SNOMED CT | 194785008 | Benign secondary hypertension |
| SNOMED CT | 194788005 | Hypertension secondary to endocrine disorder |
| SNOMED CT | 194791005 | Hypertension secondary to drug |

Continued on next page

**Table12 – continued from previous page**

| CodeSystem | ConceptCode | ConceptName |
|---|---|---|
| SNOMED CT | 199008003 | Pre-existing secondary hypertension complicating pregnancy, childbirth and puerperium |
| SNOMED CT | 169465000 | Hypertension induced by oral contraceptive pill |
| SNOMED CT | 371125006 | Labile essential hypertension |
| SNOMED CT | 38341003 | Hypertensive disorder |
| SNOMED CT | 39018007 | Renal arterial hypertension |
| SNOMED CT | 39727004 | Hypertension secondary to renal disease complicating AND/OR reason for care during puerperium |
| SNOMED CT | 46481004 | Low-renin essential hypertension |
| SNOMED CT | 48146000 | Diastolic hypertension |
| SNOMED CT | 48552006 | Hypertension secondary to renal disease complicating AND/OR reason for care during pregnancy |
| SNOMED CT | 56218007 | Systolic hypertension |
| SNOMED CT | 57684003 | Parenchymal renal hypertension |
| SNOMED CT | 59621000 | Essential hypertension |
| SNOMED CT | 59720008 | Sustained diastolic hypertension |
| SNOMED CT | 65518004 | Labile diastolic hypertension |
| SNOMED CT | 73410007 | Benign secondary renovascular hypertension |
| SNOMED CT | 74451002 | Secondary diastolic hypertension |
| SNOMED CT | 78975002 | Malignant essential hypertension |
| SNOMED CT | 427889009 | Hypertension associated with transplantation |
| SNOMED CT | 428575007 | Hypertension secondary to kidney transplant |
| SNOMED CT | 429457004 | Systolic essential hypertension |
| SNOMED CT | 89242004 | Malignant secondary hypertension |
| SNOMED CT | 111438007 | Hypertension secondary to renal disease in obstetric context |
| SNOMED CT | 123799005 | Renovascular hypertension |
| SNOMED CT | 123800009 | Goldblatt hypertension |
| SNOMED CT | 8218002 | Chronic hypertension complicating AND/OR reason for care during childbirth |
| SNOMED CT | 8762007 | Chronic hypertension in obstetric context |
| SNOMED CT | 9901000 | Essential hypertension complicating AND/OR reason for care during puerperium |
| SNOMED CT | 10562009 | Malignant hypertension complicating AND/OR reason for care during childbirth |
| SNOMED CT | 15394000 | Toxemia of pregnancy |
| SNOMED CT | 18416000 | Essential hypertension complicating AND/OR reason for care during childbirth |
| SNOMED CT | 19769006 | High-renin essential hypertension |
| SNOMED CT | 23130000 | Paroxysmal hypertension |
| SNOMED CT | 23717007 | Benign essential hypertension complicating AND/OR reason for care during pregnancy |
| SNOMED CT | 23786008 | Malignant hypertension complicating AND/OR reason for care during puerperium |

Continued on next page

**Table12 – continued from previous page**

| CodeSystem | ConceptCode | ConceptName |
|---|---|---|
| SNOMED CT | 24042004 | Chronic hypertension complicating AND/OR reason for care during puerperium |
| SNOMED CT | 29259002 | Malignant hypertension complicating AND/OR reason for care during pregnancy |
| SNOMED CT | 31407004 | Pre-existing hypertension complicating AND/OR reason for care during puerperium |
| SNOMED CT | 34694006 | Pre-existing hypertension complicating AND/OR reason for care during childbirth |
| SNOMED CT | 35303009 | Benign essential hypertension complicating AND/OR reason for care during puerperium |
| SNOMED CT | 1204139007 | Hypertension due to congenital adrenal hyperplasia |
| SNOMED CT | 1208845005 | Secondary hypertension due to congenital heart disorder |
| SNOMED CT | 1208839002 | Secondary hypertension due to renal tubular disorder |
| SNOMED CT | 198941007 | Hypertension complicating pregnancy, childbirth and the puerperium |
| SNOMED CT | 198942000 | Benign essential hypertension complicating pregnancy, childbirth and the puerperium |
| SNOMED CT | 198944004 | Benign essential hypertension complicating pregnancy, childbirth and the puerperium - delivered |
| SNOMED CT | 198945003 | Benign essential hypertension complicating pregnancy, childbirth and the puerperium - delivered with postnatal complication |
| SNOMED CT | 198946002 | Benign essential hypertension complicating pregnancy, childbirth and the puerperium - not delivered |
| SNOMED CT | 198947006 | Benign essential hypertension complicating pregnancy, childbirth and the puerperium with postnatal complication |
| SNOMED CT | 198949009 | Renal hypertension complicating pregnancy, childbirth and the puerperium |
| SNOMED CT | 198951008 | Renal hypertension complicating pregnancy, childbirth and the puerperium - delivered |
| SNOMED CT | 198952001 | Renal hypertension complicating pregnancy, childbirth and the puerperium - delivered with postnatal complication |
| SNOMED CT | 198953006 | Renal hypertension complicating pregnancy, childbirth and the puerperium - not delivered |
| SNOMED CT | 198965005 | Transient hypertension of pregnancy - delivered |
| SNOMED CT | 198966006 | Transient hypertension of pregnancy - delivered with postnatal complication |
| SNOMED CT | 198967002 | Transient hypertension of pregnancy - not delivered |
| SNOMED CT | 198968007 | Transient hypertension of pregnancy with postnatal complication |
| SNOMED CT | 198983002 | Severe pre-eclampsia - delivered |
| SNOMED CT | 198984008 | Severe pre-eclampsia - delivered with postnatal complication |

Continued on next page

**Table12 – continued from previous page**

| CodeSystem | ConceptCode | ConceptName |
|---|---|---|
| SNOMED CT | 198985009 | Severe pre-eclampsia - not delivered |
| SNOMED CT | 198986005 | Severe pre-eclampsia with postnatal complication |
| SNOMED CT | 198997005 | Pre-eclampsia or eclampsia with pre-existing hypertension |
| SNOMED CT | 198999008 | Pre-eclampsia or eclampsia with pre-existing hypertension - delivered |
| SNOMED CT | 199000005 | Pre-eclampsia or eclampsia with pre-existing hypertension - delivered with postnatal complication |
| SNOMED CT | 199002002 | Pre-eclampsia or eclampsia with pre-existing hypertension - not delivered |
| SNOMED CT | 199003007 | Pre-eclampsia or eclampsia with pre-existing hypertension with postnatal complication |
| SNOMED CT | 199005000 | Pre-existing hypertension complicating pregnancy, childbirth and puerperium |
| SNOMED CT | 199007008 | Pre-existing hypertensive heart and renal disease complicating pregnancy, childbirth and the puerperium |
| SNOMED CT | 765182005 | Postpartum pre-eclampsia |
| SNOMED CT | 766937004 | Hypertension due to gain-of-function mutation in mineralocorticoid receptor |
| SNOMED CT | 871642009 | Hypertension due to aortic arch obstruction |
| SNOMED CT | 541000119105 | Hypertension complicating pregnancy, childbirth and the puerperium, antepartum |
| SNOMED CT | 5501000119106 | Postoperative hypertension |
| SNOMED CT | 40511000119107 | Postpartum pre-existing essential hypertension |
| SNOMED CT | 40521000119100 | Postpartum pregnancy-induced hypertension |
| SNOMED CT | 71421000119105 | Hypertension in chronic kidney disease due to type 2 diabetes mellitus |
| SNOMED CT | 71701000119105 | Hypertension in chronic kidney disease due to type 1 diabetes mellitus |
| SNOMED CT | 82771000119102 | Hypertension complicating pregnancy |
| SNOMED CT | 118781000119108 | Pre-existing hypertensive chronic kidney disease in mother complicating pregnancy |
| SNOMED CT | 127991000119101 | Hypertension concurrent and due to end stage renal disease on dialysis due to type 2 diabetes mellitus |
| SNOMED CT | 128001000119105 | Hypertension concurrent and due to end stage renal disease on dialysis due to type 1 diabetes mellitus |
| SNOMED CT | 132721000119104 | Hypertensive emergency |
| SNOMED CT | 140101000119109 | Hypertension in chronic kidney disease stage 5 due to type 2 diabetes mellitus |
| SNOMED CT | 140111000119107 | Hypertension in chronic kidney disease stage 4 due to type 2 diabetes mellitus |
| SNOMED CT | 140121000119100 | Hypertension in chronic kidney disease stage 3 due to type 2 diabetes mellitus |

Continued on next page

**Table12 – continued from previous page**

| CodeSystem | ConceptCode | ConceptName |
|---|---|---|
| SNOMED CT | 140131000119102 | Hypertension in chronic kidney disease stage 2 due to type 2 diabetes mellitus |
| SNOMED CT | 367821000119106 | Page kidney |
| SNOMED CT | 434711000124103 | Perioperative hypertension |
| SNOMED CT | 10752641000119102 | Eclampsia with pre-existing hypertension in childbirth |
| SNOMED CT | 10757401000119104 | Pre-existing hypertensive heart and chronic kidney disease in mother complicating childbirth |
| SNOMED CT | 16229371000119106 | Labile systemic arterial hypertension |
| SNOMED CT | 48194001 | GH - Gestational hypertension |
| SNOMED CT | 95605009 | Hemolysis-elevated liver enzymes-low platelet count syndrome |
| SNOMED CT | 206596003 | Neonatal hypertension |
| SNOMED CT | 237279007 | Transient hypertension of pregnancy |
| SNOMED CT | 237281009 | Moderate proteinuric hypertension of pregnancy |
| SNOMED CT | 237282002 | Impending eclampsia |
| SNOMED CT | 367390009 | Hypertension in the obstetric context |
| SNOMED CT | 397748008 | Hypertension with albuminuria |
| SNOMED CT | 398254007 | Toxemia of pregnancy |
| SNOMED CT | 37618003 | Chronic hypertension complicating AND/OR reason for care during pregnancy |
| SNOMED CT | 41114007 | Mild pre-eclampsia |
| SNOMED CT | 46764007 | Severe pre-eclampsia |
| SNOMED CT | 52698002 | Transient hypertension |
| SNOMED CT | 63287004 | Benign essential hypertension in obstetric context |
| SNOMED CT | 65402008 | Pre-existing hypertension complicating AND/OR reason for care during pregnancy |
| SNOMED CT | 67359005 | Pre-eclampsia added to pre-existing hypertension |
| SNOMED CT | 69909000 | Eclampsia added to pre-existing hypertension |
| SNOMED CT | 70272006 | Malignant hypertension |
| SNOMED CT | 71874008 | Benign essential hypertension complicating AND/OR reason for care during childbirth |
| SNOMED CT | 72022006 | Essential hypertension in obstetric context |
| SNOMED CT | 78808002 | Essential hypertension complicating AND/OR reason for care during pregnancy |
| SNOMED CT | 81626002 | Malignant hypertension in obstetric context |
| SNOMED CT | 429198000 | Exertional hypertension |
| SNOMED CT | 443482000 | Hypertensive urgency |
| SNOMED CT | 472749004 | Coronary sinus hypertension as complication of procedure |
| SNOMED CT | 697929007 | Intermittent hypertension |
| SNOMED CT | 697930002 | Labile hypertension due to being in a clinical environment |
| SNOMED CT | 698638005 | Pregnancy induced hypertension with pulmonary edema |
| SNOMED CT | 698640000 | Hypertension in the puerperium with pulmonary edema |

Continued on next page

**Table12 – continued from previous page**

| CodeSystem | ConceptCode | ConceptName |
|---|---|---|
| SNOMED CT | 704667004 | Hypertension concurrent and due to end stage renal disease on dialysis |
| SNOMED CT | 706882009 | Hypertensive crisis |
| SNOMED CT | 712832005 | Supine hypertension |
| SNOMED CT | 720568003 | Brachydactyly and arterial hypertension syndrome |
| SNOMED CT | 288250001 | Maternal hypertension |
| SNOMED CT | 307632004 | Non-proteinuric hypertension of pregnancy |
| SNOMED CT | 84094009 | Rebound hypertension |
| SNOMED CT | 86041002 | Pre-existing hypertension in obstetric context |

##### 0.4.10 Major Depressive Disorder

Table 13: Concept codes used to identify major depression conditions.

| CodeSystem | ConceptCode | ConceptName |
|---|---|---|
| ICD10CM | F32.0 | Major depressive disorder, single episode, mild |
| ICD10CM | F32.1 | Major depressive disorder, single episode, moderate |
| ICD10CM | F32.2 | Major depressive disorder, single episode, severe without psychotic features |
| ICD10CM | F32.3 | Major depressive disorder, single episode, severe with psychotic features |
| ICD10CM | F32.4 | Major depressive disorder, single episode, in partial remission |
| ICD10CM | F32.9 | Major depressive disorder, single episode, unspecified |
| ICD10CM | F33.0 | Major depressive disorder, recurrent, mild |
| ICD10CM | F33.1 | Major depressive disorder, recurrent, moderate |
| ICD10CM | F33.2 | Major depressive disorder, recurrent severe without psychotic features |
| ICD10CM | F33.3 | Major depressive disorder, recurrent, severe with psychotic symptoms |
| ICD10CM | F33.41 | Major depressive disorder, recurrent, in partial remission |
| ICD10CM | F33.9 | Major depressive disorder, recurrent, unspecified |
| SNOMED CT | 832007 | Moderate major depression |
| SNOMED CT | 2618002 | Chronic recurrent major depressive disorder |
| SNOMED CT | 14183003 | Chronic major depressive disorder, single episode |
| SNOMED CT | 15193003 | Severe recurrent major depression with psychotic features, mood-incongruent |
| SNOMED CT | 15639000 | Moderate major depression, single episode |
| SNOMED CT | 18818009 | Moderate recurrent major depression |

Continued on next page

**Table13 – continued from previous page**

| CodeSystem | ConceptCode | ConceptName |
|---|---|---|
| SNOMED CT | 20250007 | Severe major depression, single episode, with psychotic features, mood-incongruent |
| SNOMED CT | 25922000 | Major depressive disorder, single episode with postpartum onset |
| SNOMED CT | 28475009 | Severe recurrent major depression with psychotic features |
| SNOMED CT | 33078009 | Severe recurrent major depression with psychotic features, mood-congruent |
| SNOMED CT | 33736005 | Severe major depression with psychotic features, mood-congruent |
| SNOMED CT | 251000119105 | Severe major depression, single episode |
| SNOMED CT | 281000119103 | Severe recurrent major depression |
| SNOMED CT | 10811121000119102 | Major depressive disorder in mother complicating childbirth |
| SNOMED CT | 10811161000119107 | Major depressive disorder in mother complicating pregnancy |
| SNOMED CT | 16264621000119109 | Recurrent mild major depressive disorder co-occurrent with anxiety |
| SNOMED CT | 16264901000119109 | Recurrent moderate major depressive disorder co-occurrent with anxiety |
| SNOMED CT | 16265951000119109 | Mild major depressive disorder co-occurrent with anxiety single episode |
| SNOMED CT | 16266831000119100 | Moderate major depressive disorder co-occurrent with anxiety single episode |
| SNOMED CT | 16266991000119108 | Severe major depressive disorder co-occurrent with anxiety single episode |
| SNOMED CT | 191604000 | Single major depressive episode, severe, with psychosis |
| SNOMED CT | 191610000 | Recurrent major depressive episodes, mild |
| SNOMED CT | 191611001 | Recurrent major depressive episodes, moderate |
| SNOMED CT | 191613003 | Recurrent major depressive episodes, severe, with psychosis |
| SNOMED CT | 319768000 | Recurrent major depressive disorder with melancholic features |
| SNOMED CT | 320751009 | Major depression, melancholic type |
| SNOMED CT | 370143000 | Major depressive disorder |
| SNOMED CT | 36474008 | Severe recurrent major depression without psychotic features |
| SNOMED CT | 36923009 | Major depression, single episode |
| SNOMED CT | 38694004 | Recurrent major depressive disorder with atypical features |
| SNOMED CT | 39809009 | Recurrent major depressive disorder with catatonic features |
| SNOMED CT | 40379007 | Mild recurrent major depression |
| SNOMED CT | 42925002 | Major depressive disorder, single episode with atypical features |
| SNOMED CT | 60099002 | Severe major depression with psychotic features, mood-incongruent |

Continued on next page

**Table13 – continued from previous page**

| CodeSystem | ConceptCode | ConceptName |
|---|---|---|
| SNOMED CT | 63778009 | Major depressive disorder, single episode with melancholic features |
| SNOMED CT | 66344007 | Recurrent major depression |
| SNOMED CT | 69392006 | Major depressive disorder, single episode with catatonic features |
| SNOMED CT | 71336009 | Recurrent major depressive disorder with postpartum onset |
| SNOMED CT | 73867007 | Severe major depression with psychotic features |
| SNOMED CT | 75084000 | Severe major depression without psychotic features |
| SNOMED CT | 76441001 | Severe major depression, single episode, without psychotic features |
| SNOMED CT | 77911002 | Severe major depression, single episode, with psychotic features, mood-congruent |
| SNOMED CT | 79298009 | Mild major depression, single episode |
| SNOMED CT | 430852001 | Severe major depression, single episode, with psychotic features |
| SNOMED CT | 450714000 | Severe major depression |
| SNOMED CT | 719592004 | Moderately severe major depression |
| SNOMED CT | 720451004 | Minimal recurrent major depression |
| SNOMED CT | 720452006 | Moderately severe recurrent major depression |
| SNOMED CT | 720453001 | Moderately severe major depression single episode |
| SNOMED CT | 720454007 | Minimal major depression single episode |
| SNOMED CT | 720455008 | Minimal major depression |
| SNOMED CT | 726772006 | Major depression with psychotic features |
| SNOMED CT | 268621008 | Recurrent major depressive episodes |
| SNOMED CT | 87512008 | Mild major depression |
| SNOMED CT | 19527009 | Single episode of major depression in full remission |
| SNOMED CT | 30605009 | Major depression in partial remission |
| SNOMED CT | 33135002 | Recurrent major depression in partial remission |
| SNOMED CT | 104851000119103 | Postpartum major depression in remission |
| SNOMED CT | 16264821000119108 | Recurrent severe major depressive disorder co-occurrent with anxiety |
| SNOMED CT | 16265061000119105 | Recurrent major depressive disorder co-occurrent with anxiety in full remission |
| SNOMED CT | 16265301000119106 | Recurrent major depressive disorder in partial remission co-occurrent with anxiety |
| SNOMED CT | 42810003 | Major depression in remission |
| SNOMED CT | 46244001 | Recurrent major depression in full remission |
| SNOMED CT | 63412003 | Major depression in full remission |
| SNOMED CT | 68019004 | Recurrent major depression in remission |
| SNOMED CT | 70747007 | Major depression single episode, in partial remission |

##### 0.4.11 Myocardial Infarction

Table 14: Concept codes used to identify acute myocardial infarction conditions.

| CodeSystem | ConceptCode | ConceptName |
|---|---|---|
| ICD10CM | I21.01 | ST elevation (STEMI) myocardial infarction involving left main coronary artery |
| ICD10CM | I21.02 | ST elevation (STEMI) myocardial infarction involving left anterior descending coronary artery |
| ICD10CM | I21.09 | ST elevation (STEMI) myocardial infarction involving other coronary artery of anterior wall |
| ICD10CM | I21.11 | ST elevation (STEMI) myocardial infarction involving right coronary artery |
| ICD10CM | I21.19 | ST elevation (STEMI) myocardial infarction involving other coronary artery of inferior wall |
| ICD10CM | I21.21 | ST elevation (STEMI) myocardial infarction involving left circumflex coronary artery |
| ICD10CM | I21.29 | ST elevation (STEMI) myocardial infarction involving other sites |
| ICD10CM | I21.3 | ST elevation (STEMI) myocardial infarction of unspecified site |
| ICD10CM | I21.4 | Non-ST elevation (NSTEMI) myocardial infarction |
| ICD10CM | I21.9 | Acute myocardial infarction, unspecified |
| ICD10CM | I21.A1 | Myocardial infarction type 2 |
| ICD10CM | I21.A9 | Other myocardial infarction type |
| SNOMED CT | 282006 | Acute myocardial infarction of basal-lateral wall |
| SNOMED CT | 10273003 | Acute infarction of papillary muscle |
| SNOMED CT | 15990001 | Acute myocardial infarction of posterolateral wall |
| SNOMED CT | 30277009 | Rupture of ventricle due to acute myocardial infarction |
| SNOMED CT | 17531000119105 | Acute myocardial infarction due to left coronary artery occlusion |
| SNOMED CT | 23311000119105 | Acute myocardial infarction due to right coronary artery occlusion |
| SNOMED CT | 285981000119103 | Acute ST segment elevation myocardial infarction involving left anterior descending coronary artery |
| SNOMED CT | 15962541000119106 | Acute ST segment elevation myocardial infarction of anteroapical wall |
| SNOMED CT | 15713081000119108 | Acute ST segment elevation myocardial infarction involving left main coronary artery |
| SNOMED CT | 15713121000119105 | Acute STEMI (ST elevation myocardial infarction) due to RCA (right coronary artery) occlusion |
| SNOMED CT | 194802003 | True posterior myocardial infarction |
| SNOMED CT | 194809007 | Acute myocardial infarction of atrium |
| SNOMED CT | 233825009 | Acute Q wave infarction - anteroseptal |
| SNOMED CT | 233826005 | Acute non-Q wave infarction - anteroseptal |
| SNOMED CT | 233827001 | Acute Q wave infarction - anterolateral |

Continued on next page

**Table14 – continued from previous page**

| CodeSystem | ConceptCode | ConceptName |
|---|---|---|
| SNOMED CT | 233828006 | Acute non-Q wave infarction - anterolateral |
| SNOMED CT | 233829003 | Acute Q wave infarction - inferior |
| SNOMED CT | 233830008 | Acute non-Q wave infarction - inferior |
| SNOMED CT | 233831007 | Acute Q wave infarction - inferolateral |
| SNOMED CT | 233832000 | Acute non-Q wave infarction - inferolateral |
| SNOMED CT | 233833005 | Acute Q wave infarction - lateral |
| SNOMED CT | 233834004 | Acute non-Q wave infarction - lateral |
| SNOMED CT | 233835003 | Acute widespread myocardial infarction |
| SNOMED CT | 233836002 | Acute Q wave infarction - widespread |
| SNOMED CT | 233837006 | Acute non-Q wave infarction - widespread |
| SNOMED CT | 233838001 | Acute posterior myocardial infarction |
| SNOMED CT | 401303003 | Acute ST segment elevation myocardial infarction |
| SNOMED CT | 401314000 | Acute non-ST segment elevation myocardial infarction |
| SNOMED CT | 52035003 | Acute anteroapical myocardial infarction |
| SNOMED CT | 54329005 | Acute myocardial infarction of anterior wall |
| SNOMED CT | 57054005 | Acute myocardial infarction |
| SNOMED CT | 58612006 | Acute myocardial infarction of lateral wall |
| SNOMED CT | 59063002 | Acute myocardial infarction of apical-lateral wall |
| SNOMED CT | 62695002 | Acute anteroseptal myocardial infarction |
| SNOMED CT | 64627002 | Acute myocardial infarction of high lateral wall |
| SNOMED CT | 65547006 | Acute myocardial infarction of inferolateral wall |
| SNOMED CT | 70211005 | Acute myocardial infarction of anterolateral wall |
| SNOMED CT | 70422006 | Acute subendocardial infarction |
| SNOMED CT | 70998009 | Acute myocardial infarction of posterobasal wall |
| SNOMED CT | 73795002 | Acute myocardial infarction of inferior wall |
| SNOMED CT | 76593002 | Acute myocardial infarction of inferoposterior wall |
| SNOMED CT | 79009004 | Acute myocardial infarction of septum |
| SNOMED CT | 703164000 | Acute anterior ST segment elevation myocardial infarction |
| SNOMED CT | 304914007 | Acute Q wave myocardial infarction |
| SNOMED CT | 307140009 | Acute non-Q wave infarction |
| SNOMED CT | 1163440003 | Postoperative acute myocardial infarction |
| SNOMED CT | 1208872002 | Subsequent anterior non-ST segment elevation myocardial infarction |
| SNOMED CT | 836293000 | Acute myocardial infarction of right ventricle |
| SNOMED CT | 836294006 | Acute myocardial infarction of apex of heart |
| SNOMED CT | 836295007 | Acute myocardial infarction of inferolateral wall with posterior extension |
| SNOMED CT | 840309000 | Acute ST segment elevation myocardial infarction due to proximal left anterior descending coronary artery occlusion |
| SNOMED CT | 840312002 | Acute ST segment elevation myocardial infarction due to mid left anterior descending coronary artery occlusion |
| SNOMED CT | 840316004 | Acute ST segment elevation myocardial infarction due to distal left anterior descending coronary artery occlusion |

Continued on next page

**Table14 – continued from previous page**

| CodeSystem | ConceptCode | ConceptName |
|---|---|---|
| SNOMED CT | 840609007 | Acute ST segment elevation myocardial infarction due to occlusion of anterior descending branch of left coronary artery |
| SNOMED CT | 840680009 | Acute ST segment elevation myocardial infarction due to occlusion of septal branch of anterior descending branch of left coronary artery |
| SNOMED CT | 846668006 | Acute ST segment elevation myocardial infarction due to occlusion of diagonal branch of anterior descending branch of left coronary artery |
| SNOMED CT | 846683001 | Acute ST segment elevation myocardial infarction due to occlusion of intermediate artery |
| SNOMED CT | 868214006 | Acute ST segment elevation myocardial infarction due to occlusion of proximal portion of right coronary artery |
| SNOMED CT | 868217004 | Acute ST segment elevation myocardial infarction due to occlusion of distal portion of right coronary artery |
| SNOMED CT | 868220007 | Acute ST segment elevation myocardial infarction due to occlusion of mid portion of right coronary artery |
| SNOMED CT | 868224003 | Acute ST segment elevation myocardial infarction due to occlusion of marginal branch of right coronary artery |
| SNOMED CT | 868225002 | Acute ST segment elevation myocardial infarction due to occlusion of posterior descending branch of right coronary artery |
| SNOMED CT | 868226001 | Acute ST segment elevation myocardial infarction due to occlusion of posterior lateral branch of right coronary artery |
| SNOMED CT | 12238111000119106 | Acute ST segment elevation myocardial infarction of inferolateral wall |
| SNOMED CT | 12238151000119107 | Acute ST segment elevation myocardial infarction of inferoposterior wall |
| SNOMED CT | 15712841000119100 | Acute ST segment elevation myocardial infarction of posterolateral wall |
| SNOMED CT | 15712881000119105 | Acute ST segment elevation myocardial infarction of anterolateral wall |
| SNOMED CT | 15712921000119103 | Acute ST segment elevation myocardial infarction of lateral wall |
| SNOMED CT | 15712961000119108 | Acute ST segment elevation myocardial infarction of anteroseptal wall |
| SNOMED CT | 15713041000119103 | Acute ST segment elevation myocardial infarction of posterior wall |
| SNOMED CT | 15713161000119100 | Acute ST segment elevation myocardial infarction of septum |
| SNOMED CT | 15713201000119105 | Acute ST segment elevation myocardial infarction of posterobasal wall |
| SNOMED CT | 15963181000119104 | Acute ST segment elevation myocardial infarction due to occlusion of circumflex coronary artery |

Continued on next page

**Table14 – continued from previous page**

| CodeSystem | ConceptCode | ConceptName |
|---|---|---|
| SNOMED CT | 1204151009 | Acute inferior non-ST segment elevation myocardial infarction of right ventricle |
| SNOMED CT | 1204154001 | Acute anterior non-ST segment elevation myocardial infarction with right ventricular involvement |
| SNOMED CT | 1204155000 | Acute anterior non-ST segment elevation myocardial infarction |
| SNOMED CT | 1204152002 | Acute inferior non-ST segment elevation myocardial infarction |
| SNOMED CT | 1204222000 | Acute non-ST segment elevation myocardial infarction of right ventricle |
| SNOMED CT | 15713001000119100 | Acute ST segment elevation myocardial infarction of atrium |
| SNOMED CT | 44811000087108 | Acute ST segment elevation myocardial infarction due to distal left circumflex coronary artery occlusion |
| SNOMED CT | 44821000087100 | Acute ST segment elevation myocardial infarction due to mid left circumflex coronary artery occlusion |
| SNOMED CT | 44831000087103 | Acute ST segment elevation myocardial infarction due to obtuse marginal branch of left circumflex coronary artery occlusion |
| SNOMED CT | 44841000087109 | Acute ST segment elevation myocardial infarction due to posterolateral branch of left circumflex coronary artery occlusion |
| SNOMED CT | 44851000087107 | Acute ST segment elevation myocardial infarction due to proximal left circumflex coronary artery occlusion |
| SNOMED CT | 703165004 | Acute ST segment elevation myocardial infarction of anterior wall involving right ventricle |
| SNOMED CT | 703212004 | Acute myocardial infarction during procedure |
| SNOMED CT | 703213009 | Acute ST segment elevation myocardial infarction of inferior wall |
| SNOMED CT | 703251009 | Acute myocardial infarction of inferior wall involving right ventricle |
| SNOMED CT | 703252002 | Acute myocardial infarction of anterior wall involving right ventricle |
| SNOMED CT | 703253007 | Acute ST segment elevation myocardial infarction of inferior wall involving right ventricle |
| SNOMED CT | 896689003 | Acute myocardial infarction due to occlusion of circumflex branch of left coronary artery |
| SNOMED CT | 896691006 | Acute ST segment elevation myocardial infarction due to occlusion of circumflex branch of left coronary artery |
| SNOMED CT | 896696001 | Acute ST segment elevation myocardial infarction of apex of heart |
| SNOMED CT | 896697005 | Acute ST segment elevation myocardial infarction of right ventricle |

##### 0.4.12 Osteoporosis

Table 15: Concept codes used to identify osteoporosis conditions.

| CodeSystem | ConceptCode | ConceptName |
|---|---|---|
| ICD10CM | M81.6 | Localized osteoporosis [Lequesne] |
| ICD10CM | M81.0 | Age-related osteoporosis without current pathological fracture |
| ICD10CM | M81.8 | Other osteoporosis without current pathological fracture |
| ICD9CM | 733.01 | Senile osteoporosis |
| ICD9CM | 733.02 | Idiopathic osteoporosis |
| ICD9CM | 733.03 | Disuse osteoporosis |
| ICD9CM | 733.09 | Other osteoporosis |
| SNOMED CT | 3345002 | Idiopathic osteoporosis |
| SNOMED CT | 14651005 | Drug-induced osteoporosis |
| SNOMED CT | 32369003 | Menopausal osteoporosis |
| SNOMED CT | 735617003 | Premenopausal idiopathic osteoporosis |
| SNOMED CT | 735618008 | Osteoporosis due to malabsorption |
| SNOMED CT | 739301006 | Osteoporosis co-occurrent and due to multiple myeloma |
| SNOMED CT | 1515008 | Massive osteolysis |
| SNOMED CT | 18040001 | Type II osteoporosis |
| SNOMED CT | 203429007 | Idiopathic generalized osteoporosis |
| SNOMED CT | 203433000 | Postoophorectomy osteoporosis |
| SNOMED CT | 203434006 | Post-surgical malabsorption osteoporosis |
| SNOMED CT | 203435007 | Localized osteoporosis - Lequesne |
| SNOMED CT | 203437004 | Osteoporosis in endocrine disorders |
| SNOMED CT | 203438009 | Vertebral osteoporosis |
| SNOMED CT | 203444008 | Postoophorectomy osteoporosis with pathological fracture |
| SNOMED CT | 203446005 | Post-surgical malabsorption osteoporosis with pathological fracture |
| SNOMED CT | 203657009 | Osteoporotic kyphosis |
| SNOMED CT | 240155001 | Adult idiopathic generalized osteoporosis |
| SNOMED CT | 240157009 | Secondary generalized osteoporosis |
| SNOMED CT | 240158004 | Regional migrating osteoporosis |
| SNOMED CT | 240159007 | Transient osteoporosis of hip |
| SNOMED CT | 240161003 | Disappearing bone disease |
| SNOMED CT | 240162005 | Secondary localized osteoporosis |
| SNOMED CT | 240164006 | Post-irradiation osteoporosis |
| SNOMED CT | 240198002 | Osteoporotic vertebral collapse |
| SNOMED CT | 311806008 | Osteoporotic collapse of cervical vertebra |
| SNOMED CT | 390833005 | Osteoporosis due to corticosteroid |
| SNOMED CT | 53174001 | Disuse osteoporosis |
| SNOMED CT | 64859006 | Osteoporosis |
| SNOMED CT | 699528002 | Transient osteoporosis |
| SNOMED CT | 703264005 | Secondary osteoporosis |

Continued on next page

**Table15 – continued from previous page**

| CodeSystem | ConceptCode | ConceptName |
|---|---|---|
| SNOMED CT | 268028001 | Localized disuse osteoporosis |
| SNOMED CT | 276661002 | Primary osteoporosis |
| SNOMED CT | 281387004 | Regional osteoporosis |
| SNOMED CT | 102447009 | Postmenopausal osteoporosis |
| SNOMED CT | 109346000 | Osteoporotic bone marrow defect |
| SNOMED CT | 15743005 | Posttraumatic osteoporosis |
| SNOMED CT | 454201000124109 | Pathological fracture of left ulna due to osteoporosis |
| SNOMED CT | 11304761000119103 | Pathological fracture of right tibia due to osteoporosis |
| SNOMED CT | 11305081000119109 | Pathological fracture of right fibula due to osteoporosis |
| SNOMED CT | 11305121000119106 | Pathological fracture of left fibula due to osteoporosis |
| SNOMED CT | 11305721000119107 | Pathological fracture of right humerus due to secondary osteoporosis |
| SNOMED CT | 11305761000119102 | Pathological fracture of left humerus due to secondary osteoporosis |
| SNOMED CT | 11305801000119105 | Pathological fracture of right clavicle due to osteoporosis |
| SNOMED CT | 11305841000119107 | Pathological fracture of left clavicle due to osteoporosis |
| SNOMED CT | 11306041000119109 | Pathological fracture of right clavicle due to secondary osteoporosis |
| SNOMED CT | 11309321000119105 | Pathological fracture of right foot due to osteoporosis |
| SNOMED CT | 11309361000119100 | Pathological fracture of left foot due to osteoporosis |
| SNOMED CT | 11309641000119108 | Pathological fracture of right foot due to secondary osteoporosis |
| SNOMED CT | 11309681000119103 | Pathological fracture of left foot due to secondary osteoporosis |
| SNOMED CT | 11309801000119107 | Pathological fracture of right femur due to osteoporosis |
| SNOMED CT | 11309841000119109 | Pathological fracture of left femur due to osteoporosis |
| SNOMED CT | 11310121000119102 | Pathological fracture of right femur due to secondary osteoporosis |
| SNOMED CT | 11310161000119107 | Pathological fracture of left femur due to secondary osteoporosis |
| SNOMED CT | 11310201000119102 | Pathological fracture of right ankle due to osteoporosis |
| SNOMED CT | 11310241000119100 | Pathological fracture of left ankle due to osteoporosis |
| SNOMED CT | 11311361000119103 | Secondary osteoporotic fracture of thoracic vertebra |
| SNOMED CT | 11311441000119109 | Secondary osteoporotic fracture of lumbar vertebra |
| SNOMED CT | 11311521000119104 | Secondary osteoporotic fracture of cervical vertebra |
| SNOMED CT | 11311601000119109 | Osteoporotic fracture of vertebra |
| SNOMED CT | 11311641000119106 | Osteoporotic fracture of sacral vertebra |
| SNOMED CT | 11311801000119105 | Secondary osteoporotic fracture of vertebra |
| SNOMED CT | 11311841000119107 | Secondary osteoporotic fracture of sacral vertebra |
| SNOMED CT | 11312241000119101 | Pathological fracture of right hand due to osteoporosis |
| SNOMED CT | 11312281000119106 | Pathological fracture of left hand due to osteoporosis |
| SNOMED CT | 11312721000119102 | Pathological fracture of right radius due to osteoporosis |
| SNOMED CT | 11312761000119107 | Pathological fracture of left radius due to osteoporosis |

Continued on next page

**Table15 – continued from previous page**

| CodeSystem | ConceptCode | ConceptName |
|---|---|---|
| SNOMED CT | 11313281000119104 | Pathological fracture of right scapula due to osteoporosis |
| SNOMED CT | 11313321000119109 | Pathological fracture of left scapula due to osteoporosis |
| SNOMED CT | 11314841000119105 | Secondary osteoporotic fracture |
| SNOMED CT | 11314881000119100 | Pathological fracture of right hip due to osteoporosis |
| SNOMED CT | 11314921000119107 | Pathological fracture of left hip due to osteoporosis |
| SNOMED CT | 11315121000119108 | Pathological fracture of proximal end of right femur due to secondary osteoporosis |
| SNOMED CT | 11315161000119103 | Pathological fracture of left hip due to secondary osteoporosis |
| SNOMED CT | 11874341000119106 | Osteoporotic fracture of left rib |
| SNOMED CT | 11874381000119101 | Osteoporotic fracture of right rib |
| SNOMED CT | 16280471000119103 | Pathological fracture of left tibia due to osteoporosis |
| SNOMED CT | 11305481000119104 | Osteoporotic fracture of right humerus |
| SNOMED CT | 11305521000119104 | Osteoporotic fracture of left humerus |
| SNOMED CT | 11312681000119108 | Osteoporotic fracture of right ulna |
| SNOMED CT | 203445009 | Osteoporosis of disuse with pathological fracture |
| SNOMED CT | 203447001 | Drug-induced osteoporosis with pathological fracture |
| SNOMED CT | 203448006 | Idiopathic osteoporosis with pathological fracture |
| SNOMED CT | 203450003 | Osteoporotic fracture of lumbar vertebra |
| SNOMED CT | 203451004 | Osteoporotic fracture of thoracic vertebra |
| SNOMED CT | 203452006 | Osteoporotic fracture of cervical vertebra |
| SNOMED CT | 203453001 | Postmenopausal osteoporosis with pathological fracture |
| SNOMED CT | 240154002 | Idiopathic osteoporosis in pregnancy |
| SNOMED CT | 240156000 | Juvenile idiopathic generalized osteoporosis |
| SNOMED CT | 240160002 | Transient osteoporosis of hip in pregnancy |
| SNOMED CT | 311890007 | Osteoporotic collapse of lumbar vertebra |
| SNOMED CT | 311891006 | Osteoporotic collapse of thoracic vertebra |
| SNOMED CT | 431031001 | Osteoporotic fracture of neck of femur |
| SNOMED CT | 443165006 | Osteoporotic fracture |
| SNOMED CT | 704282002 | Secondary osteoporotic fracture of ulna |
| SNOMED CT | 704283007 | Secondary osteoporotic fracture of tibia |
| SNOMED CT | 704284001 | Secondary osteoporotic fracture of scapula |
| SNOMED CT | 704285000 | Secondary osteoporotic fracture of radius |
| SNOMED CT | 704286004 | Secondary osteoporotic fracture of humerus |
| SNOMED CT | 704287008 | Secondary osteoporotic fracture of proximal femur |
| SNOMED CT | 704288003 | Secondary osteoporotic fracture of hand |
| SNOMED CT | 704289006 | Secondary osteoporotic fracture of foot |
| SNOMED CT | 704290002 | Secondary osteoporotic fracture of fibula |
| SNOMED CT | 704291003 | Secondary osteoporotic fracture of femur |
| SNOMED CT | 704292005 | Secondary osteoporotic fracture of clavicle |
| SNOMED CT | 704293000 | Secondary osteoporotic fracture of ankle |
| SNOMED CT | 704328003 | Osteoporotic fracture of ankle |
| SNOMED CT | 704329006 | Osteoporotic fracture of clavicle |

Continued on next page

**Table15 – continued from previous page**

| CodeSystem | ConceptCode | ConceptName |
|---|---|---|
| SNOMED CT | 704330001 | Osteoporotic fracture of femur |
| SNOMED CT | 704331002 | Osteoporotic fracture of fibula |
| SNOMED CT | 704332009 | Osteoporotic fracture of bone of foot |
| SNOMED CT | 704333004 | Osteoporotic fracture of hand |
| SNOMED CT | 704334005 | Osteoporotic fracture of proximal femur |
| SNOMED CT | 704335006 | Osteoporotic fracture of humerus |
| SNOMED CT | 704336007 | Osteoporotic fracture of radius |
| SNOMED CT | 704337003 | Osteoporotic fracture of scapula |
| SNOMED CT | 704338008 | Osteoporotic fracture of tibia |
| SNOMED CT | 704339000 | Osteoporotic fracture of ulna |
| SNOMED CT | 707419009 | Osteoporosis due to cystic fibrosis |
| SNOMED CT | 897308007 | Chronic kidney disease with osteoporosis |

#### 0.5 Anti-Diabetic Medications (ADMs)

##### 0.5.1 Metformin

Table 16: Concept codes used to identify metformin medications.

| CodeSystem | ConceptCode | ConceptName |
|---|---|---|
| RxNorm | 6809 | metformin |
| RxNorm | 1243830 | Metformin / sitagliptin Extended Release Tablet [Janumet 100/1000] |
| RxNorm | 1243847 | Metformin / sitagliptin Extended Release Tablet [Janumet 50/500] |
| RxNorm | 1243023 | Linagliptin / Metformin Oral Tablet [Jentadueto 2.5/1000] |
| RxNorm | 1243030 | Linagliptin / Metformin Oral Tablet [Jentadueto 2.5/500] |
| RxNorm | 340678 | Metformin 850 MG Extended Release Tablet |
| RxNorm | 368567 | Glipizide / Metformin Oral Tablet [metaglip] |
| RxNorm | 352301 | Glipizide 5 MG / Metformin 500 MG Oral Tablet [metaglip] |
| RxNorm | 352302 | Glipizide 2.5 MG / Metformin 250 MG Oral Tablet [metaglip] |
| RxNorm | 368169 | Glyburide / Metformin Oral Tablet [Glucovance] |
| RxNorm | 352300 | Glipizide 2.5 MG / Metformin 500 MG Oral Tablet [metaglip] |
| RxNorm | 368568 | Metformin / rosiglitazone Oral Tablet [avandamet] |
| RxNorm | 446358 | Metformin 12 hour Extended Release Tablet |
| RxNorm | 404646 | Metformin 1000 MG / rosiglitazone 2 MG Oral Tablet [avandamet] |
| RxNorm | 352296 | Metformin 500 MG / rosiglitazone 4 MG Oral Tablet [avandamet] |

Continued on next page

**Table16 – continued from previous page**

| CodeSystem | ConceptCode | ConceptName |
|---|---|---|
| RxNorm | 404647 | Metformin 1000 MG / rosiglitazone 4 MG Oral Tablet [avandamet] |
| RxNorm | 352294 | Metformin 500 MG / rosiglitazone 1 MG Oral Tablet [avandamet] |
| RxNorm | 375561 | Metformin 24 Hour Extended Release Tablet |
| RxNorm | 369995 | Metformin Extended Release Tablet [Glucophage XR] |
| RxNorm | 284467 | Metformin 500 MG Extended Release Tablet [Glucophage XR] |
| RxNorm | 433928 | Metformin 750 MG 24 Hour Extended Release Tablet |
| RxNorm | 404477 | Metformin 750 MG Extended Release Tablet [Glucophage XR] |
| RxNorm | 352295 | Metformin 500 MG / rosiglitazone 2 MG Oral Tablet [avandamet] |
| RxNorm | 575998 | Glipizide 2.5 MG / Metformin 250 MG [metaglip] |
| RxNorm | 574900 | Metformin 500 MG [Glucophage XR] |
| RxNorm | 575996 | Glipizide 2.5 MG / Metformin 500 MG [metaglip] |
| RxNorm | 541776 | Metformin 500 MG / ropinirole 2 MG [avandamet] |
| RxNorm | 575991 | Metformin 500 MG / rosiglitazone 1 MG [avandamet] |
| RxNorm | 575993 | Metformin 500 MG / rosiglitazone 4 MG [avandamet] |
| RxNorm | 575992 | Metformin 500 MG / rosiglitazone 2 MG [avandamet] |
| RxNorm | 576550 | Metformin 1000 MG / rosiglitazone 4 MG [avandamet] |
| RxNorm | 576428 | Metformin 750 MG [Glucophage XR] |
| RxNorm | 541778 | Metformin 500 MG / ropinirole 2 MG Oral Tablet [avandamet] |
| RxNorm | 575997 | Glipizide 5 MG / Metformin 500 MG [metaglip] |
| RxNorm | 576549 | Metformin 1000 MG / rosiglitazone 2 MG [avandamet] |
| RxNorm | 602413 | Metformin / pioglitazone Oral Tablet [Actoplus Met] |
| RxNorm | 705136 | Metformin 1000 MG / sitagliptin 50 MG Oral Tablet [Janumet] |
| RxNorm | 602416 | Metformin 850 MG / pioglitazone 15 MG Oral Tablet [Actoplus Met] |
| RxNorm | 645111 | Metformin Oral Tablet [Glumetza] |
| RxNorm | 705134 | Metformin 1000 MG / sitagliptin 50 MG [Janumet] |
| RxNorm | 705135 | Metformin / sitagliptin Oral Tablet [Janumet] |
| RxNorm | 706114 | Metformin / rosiglitazone Oral Tablet [Avandamet] |
| RxNorm | 705138 | Metformin 500 MG / sitagliptin 50 MG Oral Tablet [Janumet] |
| RxNorm | 731425 | Metformin / pioglitazone Oral Tablet [Actoplus Met 15/850] |
| RxNorm | 748763 | Metformin 500 MG / sitagliptin 64.3 MG Oral Tablet |
| RxNorm | 602414 | Metformin 500 MG / pioglitazone 15 MG Oral Tablet [Actoplus Met] |
| RxNorm | 757607 | Metformin / sitagliptin Oral Tablet [Janumet 50 mg/500 mg] |

Continued on next page

**Table16 – continued from previous page**

| CodeSystem | ConceptCode | ConceptName |
|---|---|---|
| RxNorm | 602412 | Metformin 500 MG / pioglitazone 15 MG [Actoplus Met] |
| RxNorm | 759753 | Metformin / rosiglitazone Oral Tablet [Adandamet 2/500] |
| RxNorm | 759754 | Metformin 500 MG / rosiglitazone 2 MG Oral Tablet [Adandamet 2/500] |
| RxNorm | 806291 | Metformin / rosiglitazone Oral Tablet [Avandamet 4/500] |
| RxNorm | 806295 | Metformin / rosiglitazone Oral Tablet [Avandamet 2/500] |
| RxNorm | 806283 | Metformin / rosiglitazone Oral Tablet [Avandamet 4/1000] |
| RxNorm | 748765 | Metformin 500 MG / sitagliptin 64.3 MG Oral Tablet [Janumet] |
| RxNorm | 805674 | Metformin / repaglinide Oral Tablet [PrandiMet 2/500] |
| RxNorm | 645112 | Metformin 500 MG Oral Tablet [Glumetza] |
| RxNorm | 705137 | Metformin 500 MG / sitagliptin 50 MG [Janumet] |
| RxNorm | 759752 | Metformin 500 MG / rosiglitazone 2 MG [Adandamet 2/500] |
| RxNorm | 602415 | Metformin 850 MG / pioglitazone 15 MG [Actoplus Met] |
| RxNorm | 849593 | Glipizide / Metformin Oral Tablet [Metaglip 2.5 MG/500 MG] |
| RxNorm | 748764 | Metformin 500 MG / sitagliptin 64.3 MG [Janumet] |
| RxNorm | 806275 | Metformin / rosiglitazone Oral Tablet [Avandamet 1/500] |
| RxNorm | 860996 | 24 HR Metformin hydrochloride 1000 MG Extended Release Oral Tablet |
| RxNorm | 849589 | Glipizide / Metformin Oral Tablet [Metaglip 5 MG/500 MG] |
| RxNorm | 861746 | Glyburide / Metformin Oral Tablet [Glucovance 1.25 MG/250 MG] |
| RxNorm | 861751 | Glyburide / Metformin Oral Tablet [Glucovance 2.5 MG/500 MG] |
| RxNorm | 899999 | Metformin / pioglitazone Extended Release Tablet [Actoplus Met 30/1000] |
| RxNorm | 1043581 | Metformin / saxagliptin Extended Release Tablet [Kombiglyze 5/500] |
| RxNorm | 1043573 | Metformin / saxagliptin Extended Release Tablet [Kombiglyze 5/1000] |
| RxNorm | 1185627 | Metaglip 5 MG/500 MG Oral Product |
| RxNorm | 1243031 | Jentadueto 2.5/500 Oral Product |
| RxNorm | 1169924 | Actoplus Met 15/850 Oral Product |
| RxNorm | 1243025 | Jentadueto 2.5/1000 Pill |
| RxNorm | 1008476 | metformin / ropinirole |
| RxNorm | 1043561 | metformin / saxagliptin Extended Release Oral Tablet |
| RxNorm | 1043562 | metformin / saxagliptin |
| RxNorm | 1043563 | 24 HR metformin hydrochloride 1000 MG / saxagliptin 2.5 MG Extended Release Oral Tablet |

Continued on next page

**Table16 – continued from previous page**

| CodeSystem | ConceptCode | ConceptName |
|---|---|---|
| RxNorm | 1043565 | metformin hydrochloride 1000 MG / saxagliptin 2.5 MG [Kombiglyze] |
| RxNorm | 1043566 | metformin / saxagliptin Extended Release Oral Tablet [Kombiglyze] |
| RxNorm | 1043567 | 24 HR metformin hydrochloride 1000 MG / saxagliptin 2.5 MG Extended Release Oral Tablet [Kombiglyze] |
| RxNorm | 1043568 | metformin hydrochloride 1000 MG / saxagliptin 2.5 MG Extended Release Oral Tablet |
| RxNorm | 1043569 | metformin hydrochloride 1000 MG / saxagliptin 2.5 MG Extended Release Oral Tablet [Kombiglyze] |
| RxNorm | 1043570 | 24 HR metformin hydrochloride 1000 MG / saxagliptin 5 MG Extended Release Oral Tablet |
| RxNorm | 1043572 | metformin hydrochloride 1000 MG / saxagliptin 5 MG [Kombiglyze] |
| RxNorm | 1043574 | 24 HR metformin hydrochloride 1000 MG / saxagliptin 5 MG Extended Release Oral Tablet [Kombiglyze] |
| RxNorm | 1043575 | metformin hydrochloride 1000 MG / saxagliptin 5 MG Extended Release Oral Tablet |
| RxNorm | 1043576 | metformin hydrochloride 1000 MG / saxagliptin 5 MG Extended Release Oral Tablet [Kombiglyze] |
| RxNorm | 1043578 | 24 HR metformin hydrochloride 500 MG / saxagliptin 5 MG Extended Release Oral Tablet |
| RxNorm | 1043580 | metformin hydrochloride 500 MG / saxagliptin 5 MG [Kombiglyze] |
| RxNorm | 1043582 | 24 HR metformin hydrochloride 500 MG / saxagliptin 5 MG Extended Release Oral Tablet [Kombiglyze] |
| RxNorm | 1043583 | metformin hydrochloride 500 MG / saxagliptin 5 MG Extended Release Oral Tablet |
| RxNorm | 1043584 | metformin hydrochloride 500 MG / saxagliptin 5 MG Extended Release Oral Tablet [Kombiglyze] |
| RxNorm | 105376 | metformin hydrochloride 500 MG Oral Tablet [Glucamet] |
| RxNorm | 105377 | metformin hydrochloride 850 MG Oral Tablet [Glucamet] |
| RxNorm | 1156197 | glyburide / metformin Pill |
| RxNorm | 1155467 | chlorpropamide / metformin Oral Product |
| RxNorm | 1155468 | chlorpropamide / metformin Pill |
| RxNorm | 1161597 | metformin / pioglitazone Oral Product |
| RxNorm | 1161598 | metformin / pioglitazone Pill |
| RxNorm | 1161599 | metformin / repaglinide Oral Product |
| RxNorm | 1161600 | metformin / repaglinide Pill |
| RxNorm | 1161601 | metformin / ropinirole Oral Product |
| RxNorm | 1161602 | metformin / ropinirole Pill |
| RxNorm | 1161603 | metformin / rosiglitazone Oral Product |
| RxNorm | 1161604 | metformin / rosiglitazone Pill |

Continued on next page

**Table16 – continued from previous page**

| CodeSystem | ConceptCode | ConceptName |
|---|---|---|
| RxNorm | 1161605 | metformin / saxagliptin Oral Product |
| RxNorm | 1161606 | metformin / saxagliptin Pill |
| RxNorm | 1161607 | metformin / sitagliptin Oral Product |
| RxNorm | 1161608 | metformin / sitagliptin Pill |
| RxNorm | 1161609 | metformin Oral Liquid Product |
| RxNorm | 1161610 | metformin Oral Product |
| RxNorm | 1161611 | metformin Pill |
| RxNorm | 1165205 | glipizide / metformin Oral Product |
| RxNorm | 1165206 | glipizide / metformin Pill |
| RxNorm | 1165845 | glyburide / metformin Oral Product |
| RxNorm | 1171242 | Glucamet Oral Product |
| RxNorm | 1171243 | Glucamet Pill |
| RxNorm | 1171244 | Glucophage Oral Product |
| RxNorm | 1171245 | Glucophage Pill |
| RxNorm | 1171248 | Glucovance Oral Product |
| RxNorm | 1171249 | Glucovance Pill |
| RxNorm | 1171254 | Glumetza Oral Product |
| RxNorm | 1171255 | Glumetza Pill |
| RxNorm | 1169920 | Actoplus Met Oral Product |
| RxNorm | 1169923 | Actoplus Met Pill |
| RxNorm | 1175016 | Avandamet Oral Product |
| RxNorm | 1175021 | Avandamet Pill |
| RxNorm | 1172629 | Fortamet Oral Product |
| RxNorm | 1172630 | Fortamet Pill |
| RxNorm | 1172860 | Kombiglyze Pill |
| RxNorm | 1172861 | Kombiglyze Oral Product |
| RxNorm | 1167810 | Janumet Oral Product |
| RxNorm | 1167811 | Janumet Pill |
| RxNorm | 1184627 | PrandiMet Oral Product |
| RxNorm | 1184628 | PrandiMet Pill |
| RxNorm | 1182890 | Orabet Metformin Oral Product |
| RxNorm | 1182891 | Orabet Metformin Pill |
| RxNorm | 1185049 | Metaglip Oral Product |
| RxNorm | 1185325 | Riomet Oral Liquid Product |
| RxNorm | 1185326 | Riomet Oral Product |
| RxNorm | 1185624 | Metaglip Pill |
| RxNorm | 1185653 | Metforming Oral Product |
| RxNorm | 1185654 | Metforming Pill |
| RxNorm | 1243016 | linagliptin / metformin Oral Product |
| RxNorm | 1243017 | linagliptin / metformin Pill |
| RxNorm | 1243018 | linagliptin / metformin Oral Tablet |
| RxNorm | 1243019 | linagliptin / metformin |

Continued on next page

**Table16 – continued from previous page**

| CodeSystem | ConceptCode | ConceptName |
|---|---|---|
| RxNorm | 1243020 | linagliptin 2.5 MG / metformin hydrochloride 1000 MG Oral Tablet |
| RxNorm | 1243022 | linagliptin 2.5 MG / metformin hydrochloride 1000 MG [Jentadueto] |
| RxNorm | 1243026 | linagliptin 2.5 MG / metformin hydrochloride 1000 MG Oral Tablet [Jentadueto] |
| RxNorm | 1243027 | linagliptin 2.5 MG / metformin hydrochloride 500 MG Oral Tablet |
| RxNorm | 1243029 | linagliptin 2.5 MG / metformin hydrochloride 500 MG [Jentadueto] |
| RxNorm | 1243033 | linagliptin 2.5 MG / metformin hydrochloride 500 MG Oral Tablet [Jentadueto] |
| RxNorm | 1243034 | linagliptin 2.5 MG / metformin hydrochloride 850 MG Oral Tablet |
| RxNorm | 1243036 | linagliptin 2.5 MG / metformin hydrochloride 850 MG [Jentadueto] |
| RxNorm | 1243037 | linagliptin / metformin Oral Tablet [Jentadueto] |
| RxNorm | 1243038 | Jentadueto Oral Product |
| RxNorm | 1243039 | Jentadueto Pill |
| RxNorm | 1243040 | linagliptin 2.5 MG / metformin hydrochloride 850 MG Oral Tablet [Jentadueto] |
| RxNorm | 1243826 | metformin / sitagliptin Extended Release Oral Tablet |
| RxNorm | 1243827 | 24 HR metformin hydrochloride 1000 MG / sitagliptin 100 MG Extended Release Oral Tablet |
| RxNorm | 1243829 | metformin hydrochloride 1000 MG / sitagliptin 100 MG [Janumet] |
| RxNorm | 1243833 | 24 HR metformin hydrochloride 1000 MG / sitagliptin 100 MG Extended Release Oral Tablet [Janumet] |
| RxNorm | 1243834 | metformin hydrochloride 1000 MG / sitagliptin 100 MG Extended Release Oral Tablet |
| RxNorm | 1243835 | metformin hydrochloride 1000 MG / sitagliptin 100 MG Extended Release Oral Tablet [Janumet] |
| RxNorm | 1243839 | metformin / sitagliptin Extended Release Oral Tablet [Janumet] |
| RxNorm | 1243842 | 24 HR metformin hydrochloride 1000 MG / sitagliptin 50 MG Extended Release Oral Tablet |
| RxNorm | 1243843 | 24 HR metformin hydrochloride 1000 MG / sitagliptin 50 MG Extended Release Oral Tablet [Janumet] |
| RxNorm | 1243844 | metformin hydrochloride 1000 MG / sitagliptin 50 MG Extended Release Oral Tablet |
| RxNorm | 1243845 | metformin hydrochloride 1000 MG / sitagliptin 50 MG Extended Release Oral Tablet [Janumet] |

Continued on next page

**Table16 – continued from previous page**

| CodeSystem | ConceptCode | ConceptName |
|---|---|---|
| RxNorm | 1243846 | 24 HR metformin hydrochloride 500 MG / sitagliptin 50 MG Extended Release Oral Tablet |
| RxNorm | 1243848 | 24 HR metformin hydrochloride 500 MG / sitagliptin 50 MG Extended Release Oral Tablet [Janumet] |
| RxNorm | 1243849 | metformin hydrochloride 500 MG / sitagliptin 50 MG Extended Release Oral Tablet |
| RxNorm | 1243850 | metformin hydrochloride 500 MG / sitagliptin 50 MG Extended Release Oral Tablet [Janumet] |
| RxNorm | 1368381 | alogliptin / metformin Oral Product |
| RxNorm | 1368382 | alogliptin / metformin Pill |
| RxNorm | 1368383 | alogliptin / metformin Oral Tablet |
| RxNorm | 1368384 | alogliptin / metformin |
| RxNorm | 1368385 | alogliptin 12.5 MG / metformin hydrochloride 1000 MG Oral Tablet |
| RxNorm | 1368387 | alogliptin 12.5 MG / metformin hydrochloride 1000 MG [Kazano] |
| RxNorm | 1368391 | alogliptin 12.5 MG / metformin hydrochloride 1000 MG Oral Tablet [Kazano] |
| RxNorm | 1368392 | alogliptin 12.5 MG / metformin hydrochloride 500 MG Oral Tablet |
| RxNorm | 1368394 | alogliptin 12.5 MG / metformin hydrochloride 500 MG [Kazano] |
| RxNorm | 1368395 | alogliptin / metformin Oral Tablet [Kazano] |
| RxNorm | 1368396 | Kazano Oral Product |
| RxNorm | 1368397 | Kazano Pill |
| RxNorm | 1368398 | alogliptin 12.5 MG / metformin hydrochloride 500 MG Oral Tablet [Kazano] |
| RxNorm | 1372692 | Kazano |
| RxNorm | 1372706 | Jentadueto |
| RxNorm | 1372716 | PrandiMet |
| RxNorm | 1372730 | Kombiglyze |
| RxNorm | 1372738 | Janumet |
| RxNorm | 1486436 | dapagliflozin / metformin |
| RxNorm | 151825 | Glucamet |
| RxNorm | 151827 | Glucophage |
| RxNorm | 1545146 | canagliflozin / metformin Oral Product |
| RxNorm | 1545147 | canagliflozin / metformin Pill |
| RxNorm | 1545148 | canagliflozin / metformin Oral Tablet |
| RxNorm | 1545149 | canagliflozin / metformin |
| RxNorm | 1545150 | canagliflozin 150 MG / metformin hydrochloride 1000 MG Oral Tablet |
| RxNorm | 1545151 | Invokamet |

Continued on next page

**Table16 – continued from previous page**

| CodeSystem | ConceptCode | ConceptName |
|---|---|---|
| RxNorm | 1545152 | canagliflozin 150 MG / metformin hydrochloride 1000 MG [Invokamet] |
| RxNorm | 1545153 | canagliflozin / metformin Oral Tablet [Invokamet] |
| RxNorm | 1545154 | Invokamet Oral Product |
| RxNorm | 1545155 | Invokamet Pill |
| RxNorm | 1545156 | canagliflozin 150 MG / metformin hydrochloride 1000 MG Oral Tablet [Invokamet] |
| RxNorm | 1545157 | canagliflozin 150 MG / metformin hydrochloride 500 MG Oral Tablet |
| RxNorm | 1545158 | canagliflozin 150 MG / metformin hydrochloride 500 MG [Invokamet] |
| RxNorm | 1545159 | canagliflozin 150 MG / metformin hydrochloride 500 MG Oral Tablet [Invokamet] |
| RxNorm | 1545161 | canagliflozin 50 MG / metformin hydrochloride 1000 MG Oral Tablet |
| RxNorm | 1545162 | canagliflozin 50 MG / metformin hydrochloride 1000 MG [Invokamet] |
| RxNorm | 1545163 | canagliflozin 50 MG / metformin hydrochloride 1000 MG Oral Tablet [Invokamet] |
| RxNorm | 1545164 | canagliflozin 50 MG / metformin hydrochloride 500 MG Oral Tablet |
| RxNorm | 1545165 | canagliflozin 50 MG / metformin hydrochloride 500 MG [Invokamet] |
| RxNorm | 1545166 | canagliflozin 50 MG / metformin hydrochloride 500 MG Oral Tablet [Invokamet] |
| RxNorm | 152161 | Orabet Metformin |
| RxNorm | 1593057 | dapagliflozin / metformin Extended Release Oral Tablet |
| RxNorm | 1593058 | 24 HR dapagliflozin 10 MG / metformin hydrochloride 1000 MG Extended Release Oral Tablet |
| RxNorm | 1593059 | dapagliflozin 10 MG / metformin hydrochloride 1000 MG Extended Release Oral Tablet |
| RxNorm | 1593068 | 24 HR dapagliflozin 10 MG / metformin hydrochloride 500 MG Extended Release Oral Tablet |
| RxNorm | 1593069 | dapagliflozin 10 MG / metformin hydrochloride 500 MG Extended Release Oral Tablet |
| RxNorm | 1593070 | 24 HR dapagliflozin 5 MG / metformin hydrochloride 1000 MG Extended Release Oral Tablet |
| RxNorm | 1593071 | dapagliflozin 5 MG / metformin hydrochloride 1000 MG Extended Release Oral Tablet |
| RxNorm | 1593072 | 24 HR dapagliflozin 5 MG / metformin hydrochloride 500 MG Extended Release Oral Tablet |
| RxNorm | 1593073 | dapagliflozin 5 MG / metformin hydrochloride 500 MG Extended Release Oral Tablet |

Continued on next page

**Table16 – continued from previous page**

| CodeSystem | ConceptCode | ConceptName |
|---|---|---|
| RxNorm | 1593774 | dapagliflozin / metformin Extended Release Oral Tablet [Xigduo] |
| RxNorm | 1593775 | 24 HR dapagliflozin 10 MG / metformin hydrochloride 1000 MG Extended Release Oral Tablet [Xigduo] |
| RxNorm | 1593776 | dapagliflozin 10 MG / metformin hydrochloride 1000 MG Extended Release Oral Tablet [Xigduo] |
| RxNorm | 1593826 | dapagliflozin 10 MG / metformin hydrochloride 500 MG [Xigduo] |
| RxNorm | 1593827 | dapagliflozin 10 MG / metformin hydrochloride 500 MG Extended Release Oral Tablet [Xigduo] |
| RxNorm | 1593828 | dapagliflozin 5 MG / metformin hydrochloride 1000 MG [Xigduo] |
| RxNorm | 1593829 | dapagliflozin 5 MG / metformin hydrochloride 1000 MG Extended Release Oral Tablet [Xigduo] |
| RxNorm | 1593830 | dapagliflozin 5 MG / metformin hydrochloride 500 MG [Xigduo] |
| RxNorm | 1593831 | 24 HR dapagliflozin 5 MG / metformin hydrochloride 500 MG Extended Release Oral Tablet [Xigduo] |
| RxNorm | 1593832 | dapagliflozin 5 MG / metformin hydrochloride 500 MG Extended Release Oral Tablet [Xigduo] |
| RxNorm | 1593833 | 24 HR dapagliflozin 5 MG / metformin hydrochloride 1000 MG Extended Release Oral Tablet [Xigduo] |
| RxNorm | 1593835 | 24 HR dapagliflozin 10 MG / metformin hydrochloride 500 MG Extended Release Oral Tablet [Xigduo] |
| RxNorm | 1592709 | dapagliflozin / metformin Oral Product |
| RxNorm | 1592710 | dapagliflozin / metformin Pill |
| RxNorm | 1592713 | Xigduo |
| RxNorm | 1592716 | Xigduo Oral Product |
| RxNorm | 1592717 | Xigduo Pill |
| RxNorm | 1592722 | dapagliflozin 10 MG / metformin hydrochloride 1000 MG [Xigduo] |
| RxNorm | 1664311 | empagliflozin / metformin Oral Product |
| RxNorm | 1664312 | empagliflozin / metformin Pill |
| RxNorm | 1664313 | empagliflozin / metformin Oral Tablet |
| RxNorm | 1664314 | empagliflozin / metformin |
| RxNorm | 1664315 | empagliflozin 5 MG / metformin hydrochloride 500 MG Oral Tablet |
| RxNorm | 1664316 | Synjardy |
| RxNorm | 1664317 | empagliflozin 5 MG / metformin hydrochloride 500 MG [Synjardy] |
| RxNorm | 1664318 | empagliflozin / metformin Oral Tablet [Synjardy] |
| RxNorm | 1664319 | Synjardy Oral Product |
| RxNorm | 1664320 | Synjardy Pill |

Continued on next page

**Table16 – continued from previous page**

| CodeSystem | ConceptCode | ConceptName |
|---|---|---|
| RxNorm | 1664321 | empagliflozin 5 MG / metformin hydrochloride 500 MG Oral Tablet [Synjardy] |
| RxNorm | 1664323 | empagliflozin 12.5 MG / metformin hydrochloride 500 MG Oral Tablet |
| RxNorm | 1664324 | empagliflozin 12.5 MG / metformin hydrochloride 500 MG [Synjardy] |
| RxNorm | 1664325 | empagliflozin 12.5 MG / metformin hydrochloride 500 MG Oral Tablet [Synjardy] |
| RxNorm | 1664326 | empagliflozin 5 MG / metformin hydrochloride 1000 MG Oral Tablet |
| RxNorm | 1664327 | empagliflozin 5 MG / metformin hydrochloride 1000 MG [Synjardy] |
| RxNorm | 1664328 | empagliflozin 5 MG / metformin hydrochloride 1000 MG Oral Tablet [Synjardy] |
| RxNorm | 1665367 | empagliflozin 12.5 MG / metformin hydrochloride 1000 MG Oral Tablet |
| RxNorm | 1665368 | empagliflozin 12.5 MG / metformin hydrochloride 1000 MG [Synjardy] |
| RxNorm | 1665369 | empagliflozin 12.5 MG / metformin hydrochloride 1000 MG Oral Tablet [Synjardy] |
| RxNorm | 1796088 | linagliptin / metformin Extended Release Oral Tablet |
| RxNorm | 1796089 | 24 HR linagliptin 2.5 MG / metformin hydrochloride 1000 MG Extended Release Oral Tablet |
| RxNorm | 1796090 | linagliptin / metformin Extended Release Oral Tablet [Jentadueto] |
| RxNorm | 1796091 | 24 HR linagliptin 2.5 MG / metformin hydrochloride 1000 MG Extended Release Oral Tablet [Jentadueto] |
| RxNorm | 1796092 | linagliptin 2.5 MG / metformin hydrochloride 1000 MG Extended Release Oral Tablet |
| RxNorm | 1796093 | linagliptin 2.5 MG / metformin hydrochloride 1000 MG Extended Release Oral Tablet [Jentadueto] |
| RxNorm | 1796094 | 24 HR linagliptin 5 MG / metformin hydrochloride 1000 MG Extended Release Oral Tablet |
| RxNorm | 1796095 | linagliptin 5 MG / metformin hydrochloride 1000 MG [Jentadueto] |
| RxNorm | 1796096 | 24 HR linagliptin 5 MG / metformin hydrochloride 1000 MG Extended Release Oral Tablet [Jentadueto] |
| RxNorm | 1796097 | linagliptin 5 MG / metformin hydrochloride 1000 MG Extended Release Oral Tablet |
| RxNorm | 1796098 | linagliptin 5 MG / metformin hydrochloride 1000 MG Extended Release Oral Tablet [Jentadueto] |
| RxNorm | 1807888 | Modified 24 HR metformin hydrochloride 1000 MG Extended Release Oral Tablet |

Continued on next page

**Table16 – continued from previous page**

| CodeSystem | ConceptCode | ConceptName |
|---|---|---|
| RxNorm | 1807894 | Osmotic 24 HR metformin hydrochloride 1000 MG Extended Release Oral Tablet |
| RxNorm | 1807915 | Modified 24 HR metformin hydrochloride 500 MG Extended Release Oral Tablet |
| RxNorm | 1807917 | Osmotic 24 HR metformin hydrochloride 500 MG Extended Release Oral Tablet |
| RxNorm | 1810996 | canagliflozin / metformin Extended Release Oral Tablet |
| RxNorm | 1810997 | 24 HR canagliflozin 150 MG / metformin hydrochloride 1000 MG Extended Release Oral Tablet |
| RxNorm | 1810998 | canagliflozin / metformin Extended Release Oral Tablet [Invokamet] |
| RxNorm | 1810999 | 24 HR canagliflozin 150 MG / metformin hydrochloride 1000 MG Extended Release Oral Tablet [Invokamet] |
| RxNorm | 1811000 | canagliflozin 150 MG / metformin hydrochloride 1000 MG Extended Release Oral Tablet |
| RxNorm | 1811001 | canagliflozin 150 MG / metformin hydrochloride 1000 MG Extended Release Oral Tablet [Invokamet] |
| RxNorm | 1811002 | 24 HR canagliflozin 150 MG / metformin hydrochloride 500 MG Extended Release Oral Tablet |
| RxNorm | 1811003 | 24 HR canagliflozin 150 MG / metformin hydrochloride 500 MG Extended Release Oral Tablet [Invokamet] |
| RxNorm | 1811004 | canagliflozin 150 MG / metformin hydrochloride 500 MG Extended Release Oral Tablet |
| RxNorm | 1811005 | canagliflozin 150 MG / metformin hydrochloride 500 MG Extended Release Oral Tablet [Invokamet] |
| RxNorm | 1811006 | 24 HR canagliflozin 50 MG / metformin hydrochloride 1000 MG Extended Release Oral Tablet |
| RxNorm | 1811007 | 24 HR canagliflozin 50 MG / metformin hydrochloride 1000 MG Extended Release Oral Tablet [Invokamet] |
| RxNorm | 1862684 | empagliflozin / metformin Extended Release Oral Tablet |
| RxNorm | 1862685 | 24 HR empagliflozin 10 MG / metformin hydrochloride 1000 MG Extended Release Oral Tablet |
| RxNorm | 1862686 | empagliflozin 10 MG / metformin hydrochloride 1000 MG [Synjardy] |
| RxNorm | 1862687 | empagliflozin / metformin Extended Release Oral Tablet [Synjardy] |
| RxNorm | 1862688 | 24 HR empagliflozin 10 MG / metformin hydrochloride 1000 MG Extended Release Oral Tablet [Synjardy] |
| RxNorm | 1862689 | empagliflozin 10 MG / metformin hydrochloride 1000 MG Extended Release Oral Tablet |
| RxNorm | 1862690 | empagliflozin 10 MG / metformin hydrochloride 1000 MG Extended Release Oral Tablet [Synjardy] |

Continued on next page

**Table16 – continued from previous page**

| CodeSystem | ConceptCode | ConceptName |
|---|---|---|
| RxNorm | 1862691 | 24 HR empagliflozin 12.5 MG / metformin hydrochloride 1000 MG Extended Release Oral Tablet |
| RxNorm | 1862692 | 24 HR empagliflozin 12.5 MG / metformin hydrochloride 1000 MG Extended Release Oral Tablet [Synjardy] |
| RxNorm | 1862693 | empagliflozin 12.5 MG / metformin hydrochloride 1000 MG Extended Release Oral Tablet |
| RxNorm | 1811008 | canagliflozin 50 MG / metformin hydrochloride 1000 MG Extended Release Oral Tablet |
| RxNorm | 1811009 | canagliflozin 50 MG / metformin hydrochloride 1000 MG Extended Release Oral Tablet [Invokamet] |
| RxNorm | 1811010 | 24 HR canagliflozin 50 MG / metformin hydrochloride 500 MG Extended Release Oral Tablet |
| RxNorm | 1811011 | 24 HR canagliflozin 50 MG / metformin hydrochloride 500 MG Extended Release Oral Tablet [Invokamet] |
| RxNorm | 1811012 | canagliflozin 50 MG / metformin hydrochloride 500 MG Extended Release Oral Tablet |
| RxNorm | 1811013 | canagliflozin 50 MG / metformin hydrochloride 500 MG Extended Release Oral Tablet [Invokamet] |
| RxNorm | 1862694 | empagliflozin 12.5 MG / metformin hydrochloride 1000 MG Extended Release Oral Tablet [Synjardy] |
| RxNorm | 1862695 | 24 HR empagliflozin 25 MG / metformin hydrochloride 1000 MG Extended Release Oral Tablet |
| RxNorm | 1862696 | empagliflozin 25 MG / metformin hydrochloride 1000 MG [Synjardy] |
| RxNorm | 1862697 | 24 HR empagliflozin 25 MG / metformin hydrochloride 1000 MG Extended Release Oral Tablet [Synjardy] |
| RxNorm | 1862698 | empagliflozin 25 MG / metformin hydrochloride 1000 MG Extended Release Oral Tablet |
| RxNorm | 1862699 | empagliflozin 25 MG / metformin hydrochloride 1000 MG Extended Release Oral Tablet [Synjardy] |
| RxNorm | 1862700 | 24 HR empagliflozin 5 MG / metformin hydrochloride 1000 MG Extended Release Oral Tablet |
| RxNorm | 1862701 | 24 HR empagliflozin 5 MG / metformin hydrochloride 1000 MG Extended Release Oral Tablet [Synjardy] |
| RxNorm | 1862702 | empagliflozin 5 MG / metformin hydrochloride 1000 MG Extended Release Oral Tablet |
| RxNorm | 1862703 | empagliflozin 5 MG / metformin hydrochloride 1000 MG Extended Release Oral Tablet [Synjardy] |
| RxNorm | 1940496 | 24 HR dapagliflozin 2.5 MG / metformin hydrochloride 1000 MG Extended Release Oral Tablet |
| RxNorm | 1940497 | dapagliflozin 2.5 MG / metformin hydrochloride 1000 MG [Xigduo] |

Continued on next page

**Table16 – continued from previous page**

| CodeSystem | ConceptCode | ConceptName |
|---|---|---|
| RxNorm | 1940498 | 24 HR dapagliflozin 2.5 MG / metformin hydrochloride 1000 MG Extended Release Oral Tablet [Xigduo] |
| RxNorm | 1940499 | dapagliflozin 2.5 MG / metformin hydrochloride 1000 MG Extended Release Oral Tablet |
| RxNorm | 1940500 | dapagliflozin 2.5 MG / metformin hydrochloride 1000 MG Extended Release Oral Tablet [Xigduo] |
| RxNorm | 1992681 | ertugliflozin / metformin Oral Product |
| RxNorm | 1992682 | ertugliflozin / metformin Pill |
| RxNorm | 1992683 | ertugliflozin / metformin Oral Tablet |
| RxNorm | 1992684 | ertugliflozin / metformin |
| RxNorm | 1992685 | ertugliflozin 2.5 MG / metformin hydrochloride 1000 MG Oral Tablet |
| RxNorm | 1992686 | Segluromet |
| RxNorm | 1992687 | ertugliflozin 2.5 MG / metformin hydrochloride 1000 MG [Segluromet] |
| RxNorm | 1992688 | ertugliflozin / metformin Oral Tablet [Segluromet] |
| RxNorm | 1992689 | Segluromet Oral Product |
| RxNorm | 1992690 | Segluromet Pill |
| RxNorm | 1992691 | ertugliflozin 2.5 MG / metformin hydrochloride 1000 MG Oral Tablet [Segluromet] |
| RxNorm | 1992693 | ertugliflozin 2.5 MG / metformin hydrochloride 500 MG Oral Tablet |
| RxNorm | 1992694 | ertugliflozin 2.5 MG / metformin hydrochloride 500 MG [Segluromet] |
| RxNorm | 1992695 | ertugliflozin 2.5 MG / metformin hydrochloride 500 MG Oral Tablet [Segluromet] |
| RxNorm | 1992698 | ertugliflozin 7.5 MG / metformin hydrochloride 1000 MG Oral Tablet |
| RxNorm | 1992699 | ertugliflozin 7.5 MG / metformin hydrochloride 1000 MG [Segluromet] |
| RxNorm | 1992700 | ertugliflozin 7.5 MG / metformin hydrochloride 1000 MG Oral Tablet [Segluromet] |
| RxNorm | 1992701 | ertugliflozin 7.5 MG / metformin hydrochloride 500 MG Oral Tablet |
| RxNorm | 1992702 | ertugliflozin 7.5 MG / metformin hydrochloride 500 MG [Segluromet] |
| RxNorm | 1992703 | ertugliflozin 7.5 MG / metformin hydrochloride 500 MG Oral Tablet [Segluromet] |
| RxNorm | 204045 | metformin hydrochloride 500 MG Oral Tablet [Orabet Metformin] |
| RxNorm | 204047 | metformin hydrochloride 850 MG Oral Tablet [Orabet Metformin] |
| RxNorm | 2117292 | dapagliflozin / metformin / saxagliptin |

Continued on next page

**Table16 – continued from previous page**

| CodeSystem | ConceptCode | ConceptName |
|---|---|---|
| RxNorm | 802051 | metformin Extended Release Oral Tablet [Glucophage] |
| RxNorm | 802646 | metformin / repaglinide |
| RxNorm | 802742 | metformin / repaglinide Oral Tablet |
| RxNorm | 849585 | glipizide / metformin Oral Tablet [Metaglip] |
| RxNorm | 805670 | metformin / repaglinide Oral Tablet [PrandiMet] |
| RxNorm | 806287 | metformin / rosiglitazone Oral Tablet [Avandamet] |
| RxNorm | 861730 | metformin hydrochloride 250 MG |
| RxNorm | 861731 | glipizide 2.5 MG / metformin hydrochloride 250 MG Oral Tablet |
| RxNorm | 861732 | glipizide 2.5 MG / metformin hydrochloride 250 MG [Metaglip] |
| RxNorm | 861733 | glipizide 2.5 MG / metformin hydrochloride 250 MG Oral Tablet [Metaglip] |
| RxNorm | 861736 | glipizide 2.5 MG / metformin hydrochloride 500 MG Oral Tablet |
| RxNorm | 861737 | glipizide 2.5 MG / metformin hydrochloride 500 MG [Metaglip] |
| RxNorm | 861738 | glipizide 2.5 MG / metformin hydrochloride 500 MG Oral Tablet [Metaglip] |
| RxNorm | 861740 | glipizide 5 MG / metformin hydrochloride 500 MG Oral Tablet |
| RxNorm | 861741 | glipizide 5 MG / metformin hydrochloride 500 MG [Metaglip] |
| RxNorm | 861742 | glipizide 5 MG / metformin hydrochloride 500 MG Oral Tablet [Metaglip] |
| RxNorm | 861743 | glyburide 1.25 MG / metformin hydrochloride 250 MG Oral Tablet |
| RxNorm | 861745 | glyburide 1.25 MG / metformin hydrochloride 250 MG [Glucovance] |
| RxNorm | 861747 | glyburide 1.25 MG / metformin hydrochloride 250 MG Oral Tablet [Glucovance] |
| RxNorm | 861748 | glyburide 2.5 MG / metformin hydrochloride 500 MG Oral Tablet |
| RxNorm | 861750 | glyburide 2.5 MG / metformin hydrochloride 500 MG [Glucovance] |
| RxNorm | 861752 | glyburide 2.5 MG / metformin hydrochloride 500 MG Oral Tablet [Glucovance] |
| RxNorm | 861753 | glyburide 5 MG / metformin hydrochloride 500 MG Oral Tablet |
| RxNorm | 861755 | glyburide 5 MG / metformin hydrochloride 500 MG [Glucovance] |
| RxNorm | 861756 | glyburide / metformin Oral Tablet [Glucovance] |

Continued on next page

**Table16 – continued from previous page**

| CodeSystem | ConceptCode | ConceptName |
|---|---|---|
| RxNorm | 861757 | glyburide 5 MG / metformin hydrochloride 500 MG Oral Tablet [Glucovance] |
| RxNorm | 861760 | metformin hydrochloride 1000 MG / rosiglitazone 2 MG Oral Tablet |
| RxNorm | 861761 | metformin hydrochloride 1000 MG / rosiglitazone 2 MG [Avandamet] |
| RxNorm | 861762 | metformin hydrochloride 1000 MG / rosiglitazone 2 MG Oral Tablet [Avandamet] |
| RxNorm | 861763 | metformin hydrochloride 1000 MG / rosiglitazone 4 MG Oral Tablet |
| RxNorm | 861764 | metformin hydrochloride 1000 MG / rosiglitazone 4 MG [Avandamet] |
| RxNorm | 875864 | metformin hydrochloride 500 MG [Glucamet] |
| RxNorm | 875865 | metformin hydrochloride 850 MG [Glucamet] |
| RxNorm | 861765 | metformin hydrochloride 1000 MG / rosiglitazone 4 MG Oral Tablet [Avandamet] |
| RxNorm | 861769 | metformin hydrochloride 1000 MG / sitagliptin 50 MG Oral Tablet |
| RxNorm | 861770 | metformin hydrochloride 1000 MG / sitagliptin 50 MG [Janumet] |
| RxNorm | 861771 | metformin hydrochloride 1000 MG / sitagliptin 50 MG Oral Tablet [Janumet] |
| RxNorm | 861783 | metformin hydrochloride 500 MG / pioglitazone 15 MG Oral Tablet |
| RxNorm | 861784 | metformin hydrochloride 500 MG / pioglitazone 15 MG [Actoplus Met] |
| RxNorm | 861785 | metformin hydrochloride 500 MG / pioglitazone 15 MG Oral Tablet [Actoplus Met] |
| RxNorm | 861787 | metformin hydrochloride 500 MG / repaglinide 1 MG Oral Tablet |
| RxNorm | 861788 | metformin hydrochloride 500 MG / repaglinide 1 MG [PrandiMet] |
| RxNorm | 861789 | metformin hydrochloride 500 MG / repaglinide 1 MG Oral Tablet [PrandiMet] |
| RxNorm | 861790 | metformin hydrochloride 500 MG / repaglinide 2 MG Oral Tablet |
| RxNorm | 861791 | metformin hydrochloride 500 MG / repaglinide 2 MG [PrandiMet] |
| RxNorm | 861792 | metformin hydrochloride 500 MG / repaglinide 2 MG Oral Tablet [PrandiMet] |
| RxNorm | 861795 | metformin hydrochloride 500 MG / rosiglitazone 1 MG Oral Tablet |

Continued on next page

**Table16 – continued from previous page**

| CodeSystem | ConceptCode | ConceptName |
|---|---|---|
| RxNorm | 861796 | metformin hydrochloride 500 MG / rosiglitazone 1 MG [Avandamet] |
| RxNorm | 861797 | metformin hydrochloride 500 MG / rosiglitazone 1 MG Oral Tablet [Avandamet] |
| RxNorm | 861806 | metformin hydrochloride 500 MG / rosiglitazone 2 MG Oral Tablet |
| RxNorm | 861807 | metformin hydrochloride 500 MG / rosiglitazone 2 MG [Avandamet] |
| RxNorm | 861808 | metformin hydrochloride 500 MG / rosiglitazone 2 MG Oral Tablet [Avandamet] |
| RxNorm | 861816 | metformin hydrochloride 500 MG / rosiglitazone 4 MG Oral Tablet |
| RxNorm | 861817 | metformin hydrochloride 500 MG / rosiglitazone 4 MG [Avandamet] |
| RxNorm | 861818 | metformin hydrochloride 500 MG / rosiglitazone 4 MG Oral Tablet [Avandamet] |
| RxNorm | 861819 | metformin hydrochloride 500 MG / sitagliptin 50 MG Oral Tablet |
| RxNorm | 861820 | metformin hydrochloride 500 MG / sitagliptin 50 MG [Janumet] |
| RxNorm | 861821 | metformin hydrochloride 500 MG / sitagliptin 50 MG Oral Tablet [Janumet] |
| RxNorm | 861822 | metformin hydrochloride 850 MG / pioglitazone 15 MG Oral Tablet |
| RxNorm | 861823 | metformin hydrochloride 850 MG / pioglitazone 15 MG [Actoplus Met] |
| RxNorm | 861824 | metformin hydrochloride 850 MG / pioglitazone 15 MG Oral Tablet [Actoplus Met] |
| RxNorm | 876009 | metformin hydrochloride 500 MG [Orabet Metformin] |
| RxNorm | 876010 | metformin hydrochloride 850 MG [Orabet Metformin] |
| RxNorm | 876033 | metformin hydrochloride 850 MG [Metforming] |
| RxNorm | 860974 | metformin hydrochloride 500 MG |
| RxNorm | 860975 | 24 HR metformin hydrochloride 500 MG Extended Release Oral Tablet |
| RxNorm | 860976 | metformin hydrochloride 500 MG [Glucophage] |
| RxNorm | 860977 | 24 HR metformin hydrochloride 500 MG Extended Release Oral Tablet [Glucophage] |
| RxNorm | 860978 | metformin hydrochloride 500 MG Extended Release Oral Tablet |
| RxNorm | 860979 | metformin hydrochloride 500 MG Extended Release Oral Tablet [Glucophage] |
| RxNorm | 860980 | metformin hydrochloride 750 MG |

Continued on next page

**Table16 – continued from previous page**

| CodeSystem | ConceptCode | ConceptName |
|---|---|---|
| RxNorm | 860981 | 24 HR metformin hydrochloride 750 MG Extended Release Oral Tablet |
| RxNorm | 860982 | metformin hydrochloride 750 MG [Glucophage] |
| RxNorm | 860983 | 24 HR metformin hydrochloride 750 MG Extended Release Oral Tablet [Glucophage] |
| RxNorm | 860984 | metformin hydrochloride 750 MG Extended Release Oral Tablet |
| RxNorm | 860985 | metformin hydrochloride 750 MG Extended Release Oral Tablet [Glucophage] |
| RxNorm | 860995 | metformin hydrochloride 1000 MG |
| RxNorm | 860997 | metformin hydrochloride 1000 MG [Fortamet] |
| RxNorm | 860998 | Osmotic 24 HR metformin hydrochloride 1000 MG Extended Release Oral Tablet [Fortamet] |
| RxNorm | 860999 | metformin hydrochloride 1000 MG Extended Release Oral Tablet |
| RxNorm | 861000 | metformin hydrochloride 1000 MG Extended Release Oral Tablet [Fortamet] |
| RxNorm | 861001 | metformin hydrochloride 500 MG [Fortamet] |
| RxNorm | 861002 | Osmotic 24 HR metformin hydrochloride 500 MG Extended Release Oral Tablet [Fortamet] |
| RxNorm | 861003 | metformin hydrochloride 500 MG Extended Release Oral Tablet [Fortamet] |
| RxNorm | 861004 | metformin hydrochloride 1000 MG Oral Tablet |
| RxNorm | 861005 | metformin hydrochloride 1000 MG [Glucophage] |
| RxNorm | 861006 | metformin hydrochloride 1000 MG Oral Tablet [Glucophage] |
| RxNorm | 861007 | metformin hydrochloride 500 MG Oral Tablet |
| RxNorm | 861008 | metformin hydrochloride 500 MG Oral Tablet [Glucophage] |
| RxNorm | 861009 | metformin hydrochloride 850 MG |
| RxNorm | 861010 | metformin hydrochloride 850 MG Oral Tablet |
| RxNorm | 861011 | metformin hydrochloride 850 MG [Glucophage] |
| RxNorm | 861012 | metformin hydrochloride 850 MG Oral Tablet [Glucophage] |
| RxNorm | 861014 | metformin hydrochloride 1000 MG [Glumetza] |
| RxNorm | 861015 | Modified 24 HR metformin hydrochloride 1000 MG Extended Release Oral Tablet [Glumetza] |
| RxNorm | 861016 | metformin hydrochloride 1000 MG Extended Release Oral Tablet [Glumetza] |
| RxNorm | 861017 | metformin hydrochloride 500 MG [Glumetza] |
| RxNorm | 861018 | Modified 24 HR metformin hydrochloride 500 MG Extended Release Oral Tablet [Glumetza] |
| RxNorm | 861019 | metformin hydrochloride 500 MG Extended Release Oral Tablet [Glumetza] |
| RxNorm | 861020 | metformin hydrochloride 625 MG |

Continued on next page

**Table16 – continued from previous page**

| CodeSystem | ConceptCode | ConceptName |
|---|---|---|
| RxNorm | 861021 | metformin hydrochloride 625 MG Oral Tablet |
| RxNorm | 861022 | metformin hydrochloride 625 MG [Glucophage] |
| RxNorm | 861023 | metformin hydrochloride 625 MG Oral Tablet [Glucophage] |
| RxNorm | 861024 | metformin hydrochloride 100 MG/ML |
| RxNorm | 861025 | metformin hydrochloride 100 MG/ML Oral Solution |
| RxNorm | 861026 | metformin hydrochloride 100 MG/ML [Riomet] |
| RxNorm | 861027 | metformin hydrochloride 100 MG/ML Oral Solution [Riomet] |
| RxNorm | 899988 | metformin / pioglitazone Extended Release Oral Tablet |
| RxNorm | 899989 | 24 HR metformin hydrochloride 1000 MG / pioglitazone 15 MG Extended Release Oral Tablet |
| RxNorm | 899991 | metformin hydrochloride 1000 MG / pioglitazone 15 MG [Actoplus Met] |
| RxNorm | 899992 | metformin / pioglitazone Extended Release Oral Tablet [Actoplus Met] |
| RxNorm | 899993 | 24 HR metformin hydrochloride 1000 MG / pioglitazone 15 MG Extended Release Oral Tablet [Actoplus Met] |
| RxNorm | 899994 | metformin hydrochloride 1000 MG / pioglitazone 15 MG Extended Release Oral Tablet |
| RxNorm | 899995 | metformin hydrochloride 1000 MG / pioglitazone 15 MG Extended Release Oral Tablet [Actoplus Met] |
| RxNorm | 899996 | 24 HR metformin hydrochloride 1000 MG / pioglitazone 30 MG Extended Release Oral Tablet |
| RxNorm | 899998 | metformin hydrochloride 1000 MG / pioglitazone 30 MG [Actoplus Met] |
| RxNorm | 900000 | 24 HR metformin hydrochloride 1000 MG / pioglitazone 30 MG Extended Release Oral Tablet [Actoplus Met] |
| RxNorm | 900001 | metformin hydrochloride 1000 MG / pioglitazone 30 MG Extended Release Oral Tablet |
| RxNorm | 900002 | metformin hydrochloride 1000 MG / pioglitazone 30 MG Extended Release Oral Tablet [Actoplus Met] |
| RxNorm | 977566 | metformin hydrochloride 850 MG Extended Release Oral Tablet |
| RxNorm | 1007411 | chlorpropamide / metformin |
| RxNorm | 351269 | Metformin 500 MG / rosiglitazone 2 MG Oral Tablet |
| RxNorm | 349554 | Metformin 500 MG / rosiglitazone 4 MG Oral Tablet |
| RxNorm | 351268 | Metformin 500 MG / rosiglitazone 1 MG Oral Tablet |
| RxNorm | 541765 | Metformin 1000 MG Extended Release Tablet |
| RxNorm | 284433 | Glyburide 5 MG / Metformin 500 MG Oral Tablet [Glucovance] |
| RxNorm | 311572 | Metformin 850 MG Oral Tablet |
| RxNorm | 311571 | Metformin 500 MG Extended Release Tablet |
| RxNorm | 213454 | Metformin 1000 MG Oral Tablet [Glucophage] |

Continued on next page

**Table16 – continued from previous page**

| CodeSystem | ConceptCode | ConceptName |
|---|---|---|
| RxNorm | 284432 | Glyburide 2.5 MG / Metformin 500 MG Oral Tablet [Glucovance] |
| RxNorm | 647242 | Metformin 500 MG Extended Release Tablet [Glumetza] |
| RxNorm | 541811 | Metformin 500 MG Extended Release Tablet [Fortamet] |
| RxNorm | 828382 | Metformin 750 MG Extended Release Tablet [Glucophage] |
| RxNorm | 316257 | Metformin 850 MG |
| RxNorm | 541769 | Metformin 1000 MG Extended Release Tablet [Fortamet] |
| RxNorm | 351273 | Glipizide 2.5 MG / Metformin 500 MG Oral Tablet |
| RxNorm | 311569 | Metformin 1000 MG Oral Tablet |
| RxNorm | 358336 | Metformin 500 MG 24 Hour Extended Release Tablet |
| RxNorm | 351275 | Glipizide 2.5 MG / Metformin 250 MG Oral Tablet |
| RxNorm | 284431 | Glyburide 1.25 MG / Metformin 250 MG Oral Tablet [Glucovance] |
| RxNorm | 731426 | Metformin 850 MG / pioglitazone 15 MG Oral Tablet [Actoplus Met 15/850] |
| RxNorm | 351274 | Glipizide 5 MG / Metformin 500 MG Oral Tablet |
| RxNorm | 731443 | Metformin 500 MG / pioglitazone 15 MG Oral Tablet [Actoplus Met 15/500] |
| RxNorm | 574869 | Glyburide 1.25 MG / Metformin 250 MG [Glucovance] |
| RxNorm | 566228 | Metformin 500 MG [Orabet] |
| RxNorm | 566230 | Metformin 850 MG [Orabet] |
| RxNorm | 574871 | Glyburide 5 MG / Metformin 500 MG [Glucovance] |
| RxNorm | 403819 | Metformin 750 MG Extended Release Tablet |
| RxNorm | 403968 | Metformin 100 MG/ML Oral Solution |
| RxNorm | 403915 | Metformin 1000 MG / rosiglitazone 2 MG Oral Tablet |
| RxNorm | 541767 | Metformin 1000 MG [Fortamet] |
| RxNorm | 828303 | Metformin 500 MG Extended Release Tablet [Glucophage] |
| RxNorm | 404727 | Metformin 100 MG/ML Oral Solution [Riomet] |
| RxNorm | 574870 | Glyburide 2.5 MG / Metformin 500 MG [Glucovance] |
| RxNorm | 406061 | Metformin 750 MG |
| RxNorm | 403916 | Metformin 1000 MG / rosiglitazone 4 MG Oral Tablet |
| RxNorm | 406081 | Metformin 100 MG/ML |
| RxNorm | 577095 | Metformin 850 MG / pioglitazone 15 MG Oral Tablet |
| RxNorm | 577094 | Metformin 500 MG / pioglitazone 15 MG Oral Tablet |
| RxNorm | 583193 | Metformin 850 MG [Metforming] |
| RxNorm | 576612 | Metformin 100 MG/ML [Riomet] |
| RxNorm | 654546 | Metformin 625 MG Oral Tablet |
| RxNorm | 731441 | Metformin 500 MG / pioglitazone 15 MG [Actoplus Met 15/500] |
| RxNorm | 654545 | Metformin 625 MG |
| RxNorm | 310538 | Glyburide 5 MG / Metformin 500 MG Oral Tablet |
| RxNorm | 700518 | Metformin 1000 MG / sitagliptin 50 MG Oral Tablet |
| RxNorm | 645110 | Metformin 500 MG [Glumetza] |

Continued on next page

**Table16 – continued from previous page**

| CodeSystem | ConceptCode | ConceptName |
|---|---|---|
| RxNorm | 731424 | Metformin 850 MG / pioglitazone 15 MG [Actoplus Met 15/850] |
| RxNorm | 700517 | Metformin 500 MG / sitagliptin 50 MG Oral Tablet |
| RxNorm | 310535 | Glyburide 2.5 MG / Metformin 500 MG Oral Tablet |
| RxNorm | 310533 | Glyburide 1.25 MG / Metformin 250 MG Oral Tablet |
| RxNorm | 566229 | Metformin 500 MG [Glucophage] |
| RxNorm | 311570 | Metformin 500 MG Oral Tablet |
| RxNorm | 573342 | Metformin 1000 MG [Glucophage] |
| RxNorm | 566231 | Metformin 850 MG [Glucophage] |
| RxNorm | 757606 | Metformin 500 MG / sitagliptin 50 MG [Janumet 50 mg/500 mg] |
| RxNorm | 204046 | Metformin 500 MG Oral Tablet [Glucophage] |
| RxNorm | 204048 | Metformin 850 MG Oral Tablet [Glucophage] |
| RxNorm | 476506 | Metformin 1000 MG 24 Hour Extended Release Tablet |
| RxNorm | 757602 | Metformin 1000 MG / sitagliptin 50 MG [Janumet 50 mg/1000 mg] |
| RxNorm | 757608 | Metformin 500 MG / sitagliptin 50 MG Oral Tablet [Janumet 50 mg/500 mg] |
| RxNorm | 757604 | Metformin 1000 MG / sitagliptin 50 MG Oral Tablet [Janumet 50 mg/1000 mg] |
| RxNorm | 541810 | Metformin 500 MG [Fortamet] |
| RxNorm | 802743 | Metformin 500 MG / repaglinide 1 MG Oral Tablet |
| RxNorm | 802050 | Metformin 750 MG [Glucophage] |
| RxNorm | 801708 | Metformin 1000 MG Extended Release Tablet [Glumetza] |
| RxNorm | 564040 | Metformin 850 MG [Glucamet] |
| RxNorm | 805669 | Metformin 500 MG / repaglinide 1 MG [PrandiMet 1/500] |
| RxNorm | 564039 | Metformin 500 MG [Glucamet] |
| RxNorm | 801707 | Metformin 1000 MG [Glumetza] |
| RxNorm | 805673 | Metformin 500 MG / repaglinide 2 MG [PrandiMet 2/500] |
| RxNorm | 806290 | Metformin 500 MG / rosiglitazone 4 MG [Avandamet 4/500] |
| RxNorm | 805671 | Metformin 500 MG / repaglinide 1 MG Oral Tablet [PrandiMet 1/500] |
| RxNorm | 806288 | Metformin 1000 MG / rosiglitazone 2 MG Oral Tablet [Avandamet 2/1000] |
| RxNorm | 806296 | Metformin 500 MG / rosiglitazone 2 MG Oral Tablet [Avandamet 2/500] |
| RxNorm | 806276 | Metformin 500 MG / rosiglitazone 1 MG Oral Tablet [Avandamet 1/500] |
| RxNorm | 806284 | Metformin 1000 MG / rosiglitazone 4 MG Oral Tablet [Avandamet 4/1000] |
| RxNorm | 806294 | Metformin 500 MG / rosiglitazone 2 MG [Avandamet 2/500] |
| RxNorm | 805675 | Metformin 500 MG / repaglinide 2 MG Oral Tablet [PrandiMet 2/500] |

Continued on next page

**Table16 – continued from previous page**

| CodeSystem | ConceptCode | ConceptName |
|---|---|---|
| RxNorm | 806292 | Metformin 500 MG / rosiglitazone 4 MG Oral Tablet [Avandamet 4/500] |
| RxNorm | 806282 | Metformin 1000 MG / rosiglitazone 4 MG [Avandamet 4/1000] |
| RxNorm | 806286 | Metformin 1000 MG / rosiglitazone 2 MG [Avandamet 2/1000] |
| RxNorm | 802744 | Metformin 500 MG / repaglinide 2 MG Oral Tablet |
| RxNorm | 806274 | Metformin 500 MG / rosiglitazone 1 MG [Avandamet 1/500] |
| RxNorm | 1184630 | PrandiMet 2/500 Pill |
| RxNorm | 1185628 | Metaglip 5 MG/500 MG Pill |
| RxNorm | 1169921 | Actoplus Met 15/1000 Pill |
| RxNorm | 1175023 | Avandamet 4/1000 Pill |
| RxNorm | 1175018 | Avandamet 2/1000 Oral Product |
| RxNorm | 1169927 | Actoplus Met 30/1000 Pill |
| RxNorm | 1175019 | Avandamet 2/1000 Pill |
| RxNorm | 1175024 | Avandamet 4/500 Oral Product |
| RxNorm | 1169922 | Actoplus Met 15/500 Oral Product |
| RxNorm | 1173547 | Kombiglyze 5/500 Oral Product |
| RxNorm | 1185625 | Metaglip 2.5 MG/500 MG Oral Product |
| RxNorm | 1171251 | Glucovance 2.5 MG/500 MG Pill |
| RxNorm | 1172859 | Kombiglyze 2.5/1000 Oral Product |
| RxNorm | 1169926 | Actoplus Met 30/1000 Oral Product |
| RxNorm | 1173546 | Kombiglyze 5/1000 Pill |
| RxNorm | 1175025 | Avandamet 4/500 Pill |
| RxNorm | 1243832 | Janumet 100/1000 Pill |
| RxNorm | 1175022 | Avandamet 4/1000 Oral Product |
| RxNorm | 1184629 | PrandiMet 2/500 Oral Product |
| RxNorm | 1175020 | Avandamet 2/500 Oral Product |
| RxNorm | 1185626 | Metaglip 2.5 MG/500 MG Pill |
| RxNorm | 1175017 | Avandamet 1/500 Pill |
| RxNorm | 1171253 | Glucovance 5 MG/500 MG Pill |
| RxNorm | 1171250 | Glucovance 2.5 MG/500 MG Oral Product |
| RxNorm | 1167812 | Janumet 50/500 Oral Product |
| RxNorm | 1243032 | Jentadueto 2.5/500 Pill |
| RxNorm | 1169925 | Actoplus Met 15/850 Pill |
| RxNorm | 1173548 | Kombiglyze 5/500 Pill |
| RxNorm | 1167813 | Janumet 50/500 Pill |
| RxNorm | 1171252 | Glucovance 5 MG/500 MG Oral Product |
| RxNorm | 1243024 | Jentadueto 2.5/1000 Oral Product |
| RxNorm | 1243831 | Janumet 100/1000 Oral Product |
| RxNorm | 2200517 | metformin Extended Release Suspension |
| RxNorm | 2200518 | metformin hydrochloride 100 MG/ML Extended Release Suspension |

Continued on next page

**Table16 – continued from previous page**

| CodeSystem | ConceptCode | ConceptName |
|---|---|---|
| RxNorm | 2200519 | metformin Extended Release Suspension [Riomet] |
| RxNorm | 2200520 | metformin hydrochloride 100 MG/ML Extended Release Suspension [Riomet] |
| RxNorm | 2281864 | empagliflozin / linagliptin / metformin |
| RxNorm | 2371722 | dapagliflozin / metformin / saxagliptin Oral Product |
| RxNorm | 2371723 | dapagliflozin / metformin / saxagliptin Pill |
| RxNorm | 2371724 | dapagliflozin / metformin / saxagliptin Extended Release Oral Tablet |
| RxNorm | 2371725 | 24 HR dapagliflozin 10 MG / metformin hydrochloride 1000 MG / saxagliptin 5 MG Extended Release Oral Tablet |
| RxNorm | 2371726 | Qternmet |
| RxNorm | 2371727 | dapagliflozin 10 MG / metformin hydrochloride 1000 MG / saxagliptin 5 MG [Qternmet] |
| RxNorm | 2371728 | dapagliflozin / metformin / saxagliptin Extended Release Oral Tablet [Qternmet] |
| RxNorm | 2371729 | Qternmet Oral Product |
| RxNorm | 2371730 | Qternmet Pill |
| RxNorm | 2371731 | 24 HR dapagliflozin 10 MG / metformin hydrochloride 1000 MG / saxagliptin 5 MG Extended Release Oral Tablet [Qternmet] |
| RxNorm | 2371732 | dapagliflozin 10 MG / metformin hydrochloride 1000 MG / saxagliptin 5 MG Extended Release Oral Tablet |
| RxNorm | 2371733 | dapagliflozin 10 MG / metformin hydrochloride 1000 MG / saxagliptin 5 MG Extended Release Oral Tablet [Qternmet] |
| RxNorm | 2371734 | 24 HR dapagliflozin 2.5 MG / metformin hydrochloride 1000 MG / saxagliptin 2.5 MG Extended Release Oral Tablet |
| RxNorm | 2371735 | dapagliflozin 2.5 MG / metformin hydrochloride 1000 MG / saxagliptin 2.5 MG [Qternmet] |
| RxNorm | 2371736 | 24 HR dapagliflozin 2.5 MG / metformin hydrochloride 1000 MG / saxagliptin 2.5 MG Extended Release Oral Tablet [Qternmet] |
| RxNorm | 2371737 | dapagliflozin 2.5 MG / metformin hydrochloride 1000 MG / saxagliptin 2.5 MG Extended Release Oral Tablet |
| RxNorm | 2371738 | dapagliflozin 2.5 MG / metformin hydrochloride 1000 MG / saxagliptin 2.5 MG Extended Release Oral Tablet [Qternmet] |
| RxNorm | 2371740 | 24 HR dapagliflozin 5 MG / metformin hydrochloride 1000 MG / saxagliptin 2.5 MG Extended Release Oral Tablet |
| RxNorm | 2371741 | dapagliflozin 5 MG / metformin hydrochloride 1000 MG / saxagliptin 2.5 MG [Qternmet] |
| RxNorm | 2371742 | 24 HR dapagliflozin 5 MG / metformin hydrochloride 1000 MG / saxagliptin 2.5 MG Extended Release Oral Tablet [Qternmet] |

Continued on next page

**Table16 – continued from previous page**

| CodeSystem | ConceptCode | ConceptName |
|---|---|---|
| RxNorm | 2371743 | dapagliflozin 5 MG / metformin hydrochloride 1000 MG / saxagliptin 2.5 MG Extended Release Oral Tablet |
| RxNorm | 2371744 | dapagliflozin 5 MG / metformin hydrochloride 1000 MG / saxagliptin 2.5 MG Extended Release Oral Tablet [Qternmet] |
| RxNorm | 2371745 | 24 HR dapagliflozin 5 MG / metformin hydrochloride 1000 MG / saxagliptin 5 MG Extended Release Oral Tablet |
| RxNorm | 2371746 | dapagliflozin 5 MG / metformin hydrochloride 1000 MG / saxagliptin 5 MG [Qternmet] |
| RxNorm | 2371747 | 24 HR dapagliflozin 5 MG / metformin hydrochloride 1000 MG / saxagliptin 5 MG Extended Release Oral Tablet [Qternmet] |
| RxNorm | 2371748 | dapagliflozin 5 MG / metformin hydrochloride 1000 MG / saxagliptin 5 MG Extended Release Oral Tablet |
| RxNorm | 2371749 | dapagliflozin 5 MG / metformin hydrochloride 1000 MG / saxagliptin 5 MG Extended Release Oral Tablet [Qternmet] |
| RxNorm | 2359276 | empagliflozin / linagliptin / metformin Oral Product |
| RxNorm | 2359277 | empagliflozin / linagliptin / metformin Pill |
| RxNorm | 2359278 | empagliflozin / linagliptin / metformin Extended Release Oral Tablet |
| RxNorm | 2359279 | 24 HR empagliflozin 10 MG / linagliptin 5 MG / metformin hydrochloride 1000 MG Extended Release Oral Tablet |
| RxNorm | 2359280 | Trijardy |
| RxNorm | 2359281 | empagliflozin 10 MG / linagliptin 5 MG / metformin hydrochloride 1000 MG [Trijardy] |
| RxNorm | 2359282 | empagliflozin / linagliptin / metformin Extended Release Oral Tablet [Trijardy] |
| RxNorm | 2359283 | Trijardy Oral Product |
| RxNorm | 2359284 | Trijardy Pill |
| RxNorm | 2359285 | 24 HR empagliflozin 10 MG / linagliptin 5 MG / metformin hydrochloride 1000 MG Extended Release Oral Tablet [Trijardy] |
| RxNorm | 2359286 | empagliflozin 10 MG / linagliptin 5 MG / metformin hydrochloride 1000 MG Extended Release Oral Tablet |
| RxNorm | 2359287 | empagliflozin 10 MG / linagliptin 5 MG / metformin hydrochloride 1000 MG Extended Release Oral Tablet [Trijardy] |
| RxNorm | 2359288 | 24 HR empagliflozin 12.5 MG / linagliptin 2.5 MG / metformin hydrochloride 1000 MG Extended Release Oral Tablet |
| RxNorm | 2359289 | empagliflozin 12.5 MG / linagliptin 2.5 MG / metformin hydrochloride 1000 MG [Trijardy] |

Continued on next page

**Table16 – continued from previous page**

| CodeSystem | ConceptCode | ConceptName |
|---|---|---|
| RxNorm | 2359290 | 24 HR empagliflozin 12.5 MG / linagliptin 2.5 MG / metformin hydrochloride 1000 MG Extended Release Oral Tablet [Trijardy] |
| RxNorm | 2359291 | empagliflozin 12.5 MG / linagliptin 2.5 MG / metformin hydrochloride 1000 MG Extended Release Oral Tablet |
| RxNorm | 2359292 | empagliflozin 12.5 MG / linagliptin 2.5 MG / metformin hydrochloride 1000 MG Extended Release Oral Tablet [Trijardy] |
| RxNorm | 2359351 | 24 HR empagliflozin 25 MG / linagliptin 5 MG / metformin hydrochloride 1000 MG Extended Release Oral Tablet |
| RxNorm | 2359352 | empagliflozin 25 MG / linagliptin 5 MG / metformin hydrochloride 1000 MG [Trijardy] |
| RxNorm | 2359353 | 24 HR empagliflozin 25 MG / linagliptin 5 MG / metformin hydrochloride 1000 MG Extended Release Oral Tablet [Trijardy] |
| RxNorm | 2359354 | empagliflozin 25 MG / linagliptin 5 MG / metformin hydrochloride 1000 MG Extended Release Oral Tablet |
| RxNorm | 2359355 | empagliflozin 25 MG / linagliptin 5 MG / metformin hydrochloride 1000 MG Extended Release Oral Tablet [Trijardy] |
| RxNorm | 2359356 | 24 HR empagliflozin 5 MG / linagliptin 2.5 MG / metformin hydrochloride 1000 MG Extended Release Oral Tablet |
| RxNorm | 2359357 | empagliflozin 5 MG / linagliptin 2.5 MG / metformin hydrochloride 1000 MG [Trijardy] |
| RxNorm | 2359358 | 24 HR empagliflozin 5 MG / linagliptin 2.5 MG / metformin hydrochloride 1000 MG Extended Release Oral Tablet [Trijardy] |
| RxNorm | 2359359 | empagliflozin 5 MG / linagliptin 2.5 MG / metformin hydrochloride 1000 MG Extended Release Oral Tablet |
| RxNorm | 2359360 | empagliflozin 5 MG / linagliptin 2.5 MG / metformin hydrochloride 1000 MG Extended Release Oral Tablet [Trijardy] |
| RxNorm | 246522 | chlorpropamide 125 MG / metformin 400 MG Oral Tablet |
| RxNorm | 250919 | glyburide 2.5 MG / metformin 400 MG Oral Tablet |
| RxNorm | 284743 | Glucovance |
| RxNorm | 285129 | glyburide / metformin |
| RxNorm | 330861 | metformin 250 MG |
| RxNorm | 316255 | metformin 1000 MG |
| RxNorm | 316256 | metformin 500 MG |
| RxNorm | 332809 | metformin 400 MG |
| RxNorm | 352381 | glipizide / metformin |
| RxNorm | 352450 | Avandamet |
| RxNorm | 352764 | Metaglip |

Continued on next page

**Table16 – continued from previous page**

| CodeSystem | ConceptCode | ConceptName |
|---|---|---|
| RxNorm | 368254 | metformin Oral Tablet [Glucophage] |
| RxNorm | 361841 | metformin Oral Tablet [Orabet Metformin] |
| RxNorm | 368526 | metformin Oral Tablet [Glucamet] |
| RxNorm | 371466 | chlorpropamide / metformin Oral Tablet |
| RxNorm | 374635 | glyburide / metformin Oral Tablet |
| RxNorm | 372803 | metformin Oral Tablet |
| RxNorm | 372804 | metformin Extended Release Oral Tablet |
| RxNorm | 378729 | metformin / rosiglitazone Oral Tablet |
| RxNorm | 378730 | glipizide / metformin Oral Tablet |
| RxNorm | 405304 | Riomet |
| RxNorm | 406082 | metformin Oral Solution |
| RxNorm | 406257 | metformin Oral Solution [Riomet] |
| RxNorm | 431724 | 12 HR metformin hydrochloride 850 MG Extended Release Oral Tablet |
| RxNorm | 432366 | chlorpropamide 125 MG / metformin 513 MG Oral Tablet |
| RxNorm | 428759 | metformin 250 MG Oral Tablet |
| RxNorm | 432780 | chlorpropamide 125 MG / metformin 500 MG Oral Tablet |
| RxNorm | 429841 | glyburide 5 MG / metformin 1000 MG Oral Tablet |
| RxNorm | 438507 | metformin 513 MG |
| RxNorm | 541766 | Fortamet |
| RxNorm | 541768 | metformin Extended Release Oral Tablet [Fortamet] |
| RxNorm | 541774 | metformin / ropinirole Oral Tablet |
| RxNorm | 541775 | metformin 500 MG / ropinirole 2 MG Oral Tablet |
| RxNorm | 583192 | Metforming |
| RxNorm | 583194 | metformin Oral Tablet [Metforming] |
| RxNorm | 583195 | metformin hydrochloride 850 MG Oral Tablet [Metforming] |
| RxNorm | 602411 | Actoplus Met |
| RxNorm | 607999 | metformin / pioglitazone |
| RxNorm | 614348 | metformin / rosiglitazone |
| RxNorm | 577093 | metformin / pioglitazone Oral Tablet |
| RxNorm | 645109 | Glumetza |
| RxNorm | 647241 | metformin Extended Release Oral Tablet [Glumetza] |
| RxNorm | 700516 | metformin / sitagliptin Oral Tablet |
| RxNorm | 729717 | metformin / sitagliptin |
| RxNorm | 731442 | metformin / pioglitazone Oral Tablet [Actoplus Met] |
| RxNorm | 757603 | metformin / sitagliptin Oral Tablet [Janumet] |

##### 0.5.2 Insulin

Table 17: Concept codes used to identify insulin medications.

| CodeSystem | ConceptCode | ConceptName |
|---|---|---|
| RxNorm | 253182 | insulin, regular, human |
| RxNorm | 1544487 | Regular Insulin, Human 0.7 MG/ACTUAT |
| RxNorm | 1544489 | Regular Insulin, Human 0.7 MG/ACTUAT [Afrezza] |
| RxNorm | 1543204 | Regular Insulin, Human 0.35 MG/ACTUAT [Afrezza] |
| RxNorm | 1543201 | Regular Insulin, Human Inhalant Powder |
| RxNorm | 1543205 | Regular Insulin, Human Inhalant Powder [Afrezza] |
| RxNorm | 1543199 | Regular Insulin, Human 0.35 MG/ACTUAT |
| RxNorm | 380934 | Insulin, Isophane, Human / Insulin, Regular, Human Injectable Suspension [Human Mixtard Penfill] |
| RxNorm | 379754 | Insulin, Isophane, Human / Insulin, Regular, Human Injectable Suspension [Novolin 70/30 PenFill] |
| RxNorm | 365578 | Insulin, Isophane, Human / Insulin, Regular, Human Injectable Suspension [Humulin M1] |
| RxNorm | 365580 | Insulin, Isophane, Human / Insulin, Regular, Human Injectable Suspension [Pur-In Mix 50/50] |
| RxNorm | 365577 | Insulin, Isophane, Human / Insulin, Regular, Human Injectable Suspension [Humulin M2] |
| RxNorm | 365575 | Insulin, Isophane, Human / Insulin, Regular, Human Injectable Suspension [Humulin M5] |
| RxNorm | 379741 | Insulin, Isophane, Human / Insulin, Regular, Human Injectable Suspension [Human Mixtard 50] |
| RxNorm | 365673 | Insulin, Isophane, Human / Insulin, Regular, Human Injectable Suspension [Humulin 70/30] |
| RxNorm | 311023 | Insulin, Isophane, Human 70 UNT/ML / Insulin, Regular, Human 30 UNT/ML Injectable Suspension [Novolin 70/30 PenFill] |
| RxNorm | 379742 | Insulin, Isophane, Human / Insulin, Regular, Human Injectable Suspension [Human Mixtard 30 ge] |
| RxNorm | 365582 | Insulin, Isophane, Human / Insulin, Regular, Human Injectable Suspension [Humulin M3] |
| RxNorm | 365576 | Insulin, Isophane, Human / Insulin, Regular, Human Injectable Suspension [Humulin M4] |
| RxNorm | 365684 | Insulin, Isophane, Human / Insulin, Regular, Human Injectable Suspension [Pur-In Mix 25/75] |
| RxNorm | 575139 | Insulin, Isophane, Human 70 UNT/ML / Insulin, Regular, Human 30 UNT/ML [Novolin 70/30 PenFill] |
| RxNorm | 724341 | Insulin, Isophane, Human / Insulin, Regular, Human Injectable Suspension [ReliOn/Novolin 70/30] |
| RxNorm | 727909 | 1.5 ML Insulin, Isophane, Human 70 UNT/ML / Insulin, Regular, Human 30 UNT/ML Prefilled Syringe |
| RxNorm | 727917 | 3 ML Insulin, Regular, Human 100 UNT/ML Prefilled Syringe |

Continued on next page

**Table17 – continued from previous page**

| CodeSystem | ConceptCode | ConceptName |
|---|---|---|
| RxNorm | 727770 | 3 ML Insulin, Isophane, Human 50 UNT/ML / Insulin, Regular, Human 50 UNT/ML Prefilled Syringe |
| RxNorm | 727766 | 3 ML Insulin, Isophane, Human 85 UNT/ML / Insulin, Regular, Human 15 UNT/ML Prefilled Syringe |
| RxNorm | 727626 | Insulin, Isophane, Human / Insulin, Regular, Human Prefilled Syringe |
| RxNorm | 723552 | Insulin, Regular, Human Injectable Solution [ReliOn R/Novolin] |
| RxNorm | 727895 | Insulin, Isophane, Human / Insulin, Regular, Human Prefilled Syringe [Humulin 70/30] |
| RxNorm | 727916 | Insulin, Regular, Human Prefilled Syringe |
| RxNorm | 727769 | 3 ML Insulin, Isophane, Human 60 UNT/ML / Insulin, Regular, Human 40 UNT/ML Prefilled Syringe |
| RxNorm | 727918 | 1.5 ML Insulin, Regular, Human 100 UNT/ML Prefilled Syringe |
| RxNorm | 727627 | 3 ML Insulin, Isophane, Human 70 UNT/ML / Insulin, Regular, Human 30 UNT/ML Prefilled Syringe |
| RxNorm | 727755 | 3 ML Insulin, Isophane, Human 90 UNT/ML / Insulin, Regular, Human 10 UNT/ML Prefilled Syringe |
| RxNorm | 727628 | Insulin, Isophane, Human / Insulin, Regular, Human Prefilled Syringe [Novolin 70/30] |
| RxNorm | 727896 | 3 ML Insulin, Isophane, Human 70 UNT/ML / Insulin, Regular, Human 30 UNT/ML Prefilled Syringe [Humulin 70/30] |
| RxNorm | 724342 | Insulin, Isophane, Human 70 UNT/ML / Insulin, Regular, Human 30 UNT/ML Injectable Suspension [ReliOn/Novolin 70/30] |
| RxNorm | 727629 | 3 ML Insulin, Isophane, Human 70 UNT/ML / Insulin, Regular, Human 30 UNT/ML Prefilled Syringe [Novolin 70/30] |
| RxNorm | 727768 | 3 ML Insulin, Isophane, Human 75 UNT/ML / Insulin, Regular, Human 25 UNT/ML Prefilled Syringe |
| RxNorm | 723553 | Insulin, Regular, Human 100 UNT/ML Injectable Solution [ReliOn R/Novolin] |
| RxNorm | 727910 | 1.5 ML Insulin, Isophane, Human 70 UNT/ML / Insulin, Regular, Human 30 UNT/ML Prefilled Syringe [Novolin 70/30] |
| RxNorm | 723551 | Insulin, Regular, Human 100 UNT/ML [ReliOn R/Novolin] |
| RxNorm | 727767 | 3 ML Insulin, Isophane, Human 80 UNT/ML / Insulin, Regular, Human 20 UNT/ML Prefilled Syringe |
| RxNorm | 724340 | Insulin, Isophane, Human 70 UNT/ML / Insulin, Regular, Human 30 UNT/ML [ReliOn/Novolin 70/30] |
| RxNorm | 1172687 | Humulin M2 Injectable Product |
| RxNorm | 1172688 | Humulin M3 Injectable Product |
| RxNorm | 1008501 | insulin isophane / insulin, regular, human |

Continued on next page

**Table17 – continued from previous page**

| CodeSystem | ConceptCode | ConceptName |
|---|---|---|
| RxNorm | 108813 | insulin isophane, human 90 UNT/ML / insulin, regular, human 10 UNT/ML Injectable Suspension [Human Mixtard] |
| RxNorm | 108814 | insulin isophane, human 80 UNT/ML / insulin, regular, human 20 UNT/ML Injectable Suspension [Human Mixtard] |
| RxNorm | 108815 | insulin isophane, human 70 UNT/ML / insulin, regular, human 30 UNT/ML Injectable Suspension [Human Mixtard] |
| RxNorm | 108816 | insulin isophane, human 50 UNT/ML / insulin, regular, human 50 UNT/ML Injectable Suspension [Human Mixtard] |
| RxNorm | 108822 | insulin isophane, human 60 UNT/ML / insulin, regular, human 40 UNT/ML Injectable Suspension [Human Mixtard] |
| RxNorm | 2644769 | {9 (insulin, regular, human 12 UNT Inhalation Powder [Afrezza]) / 9 (insulin, regular, human 4 UNT Inhalation Powder [Afrezza]) / 9 (insulin, regular, human 8 UNT Inhalation Powder [Afrezza]) } Pack [Afrezza Sample Pack] |
| RxNorm | 2644768 | {9 (insulin, regular, human 12 UNT Inhalation Powder) / 9 (insulin, regular, human 4 UNT Inhalation Powder) / 9 (insulin, regular, human 8 UNT Inhalation Powder) } Pack |
| RxNorm | 11160 | Velosulin |
| RxNorm | 106892 | insulin isophane, human 70 UNT/ML / insulin, regular, human 30 UNT/ML Injectable Suspension [Humulin] |
| RxNorm | 106894 | insulin isophane, human 85 UNT/ML / insulin, regular, human 15 UNT/ML Injectable Suspension [Pur-In Mix] |
| RxNorm | 106895 | insulin isophane, human 75 UNT/ML / insulin, regular, human 25 UNT/ML Injectable Suspension [Pur-In Mix] |
| RxNorm | 106896 | insulin isophane, human 50 UNT/ML / insulin, regular, human 50 UNT/ML Injectable Suspension [Pur-In Mix] |
| RxNorm | 106899 | insulin isophane, human 90 UNT/ML / insulin, regular, human 10 UNT/ML Injectable Suspension [Humulin] |
| RxNorm | 106900 | insulin isophane, human 80 UNT/ML / insulin, regular, human 20 UNT/ML Injectable Suspension [Humulin] |
| RxNorm | 106901 | insulin isophane, human 60 UNT/ML / insulin, regular, human 40 UNT/ML Injectable Suspension [Humulin] |
| RxNorm | 108407 | insulin isophane, human 50 UNT/ML / insulin, regular, human 50 UNT/ML Injectable Suspension [Humulin] |
| RxNorm | 1164824 | insulin, regular, human Injectable Product |
| RxNorm | 1171292 | Human Actrapid Injectable Product |
| RxNorm | 1171293 | Human Actrapid Penfill Injectable Product |
| RxNorm | 1171296 | Human Mixtard Injectable Product |
| RxNorm | 1171997 | Humulin Injectable Product |
| RxNorm | 1172692 | Humulin R Injectable Product |
| RxNorm | 1172693 | Humulin S Injectable Product |
| RxNorm | 1183566 | Pur-In Mix Injectable Product |
| RxNorm | 1183568 | Pur-In Neutral Injectable Product |

Continued on next page

**Table17 – continued from previous page**

| CodeSystem | ConceptCode | ConceptName |
|---|---|---|
| RxNorm | 1178125 | Novolin Injectable Product |
| RxNorm | 1178128 | Novolin R Injectable Product |
| RxNorm | 1187762 | Velosulin Injectable Product |
| RxNorm | 1359720 | insulin isophane, human 70 UNT/ML / insulin, regular, human 30 UNT/ML Prefilled Syringe [Humulin] |
| RxNorm | 1360172 | insulin isophane, human 70 UNT/ML / insulin, regular, human 30 UNT/ML Prefilled Syringe |
| RxNorm | 1360226 | insulin isophane, human 70 UNT/ML / insulin, regular, human 30 UNT/ML Prefilled Syringe [Novolin] |
| RxNorm | 1360482 | insulin, regular, human 100 UNT/ML Prefilled Syringe |
| RxNorm | 1360435 | insulin, regular, human 100 UNT/ML Prefilled Syringe [Novolin R] |
| RxNorm | 1372685 | Human Mixtard |
| RxNorm | 1372723 | Novolin |
| RxNorm | 1372744 | Humulin |
| RxNorm | 1372761 | Pur-In Mix |
| RxNorm | 150659 | insulin, regular, human 100 UNT/ML Injectable Solution [Pur-In Neutral] |
| RxNorm | 150831 | insulin, regular, human 100 UNT/ML Injectable Solution [Human Actrapid Penfill] |
| RxNorm | 150973 | insulin, regular, human 100 UNT/ML Injectable Solution [Human Actrapid] |
| RxNorm | 150974 | insulin, regular, human 100 UNT/ML Injectable Solution [Humulin S] |
| RxNorm | 1544488 | insulin, regular, human 8 UNT Inhalation Powder |
| RxNorm | 1544490 | insulin, regular, human 8 UNT Inhalation Powder [Afrezza] |
| RxNorm | 1544568 | {60 (insulin, regular, human 4 UNT Inhalation Powder) / 30 (insulin, regular, human 8 UNT Inhalation Powder) } Pack |
| RxNorm | 1544569 | {60 (insulin, regular, human 4 UNT Inhalation Powder [Afrezza]) / 30 (insulin, regular, human 8 UNT Inhalation Powder [Afrezza]) } Pack [Afrezza 90 - 60 (4 UNT), 30 (8 UNT)] |
| RxNorm | 1544570 | {30 (insulin, regular, human 4 UNT Inhalation Powder) / 60 (insulin, regular, human 8 UNT Inhalation Powder) } Pack |
| RxNorm | 1544571 | {30 (insulin, regular, human 4 UNT Inhalation Powder [Afrezza]) / 60 (insulin, regular, human 8 UNT Inhalation Powder [Afrezza]) } Pack [Afrezza 90 - 30 (4 UNT), 60 (8 UNT)] |
| RxNorm | 1543200 | insulin, regular, human Inhalant Product |
| RxNorm | 1543202 | insulin, regular, human 4 UNT Inhalation Powder |
| RxNorm | 1543203 | Afrezza |

Continued on next page

**Table17 – continued from previous page**

| CodeSystem | ConceptCode | ConceptName |
|---|---|---|
| RxNorm | 1543206 | Afrezza Inhalant Product |
| RxNorm | 1543207 | insulin, regular, human 4 UNT Inhalation Powder [Afrezza] |
| RxNorm | 152644 | Human Actrapid |
| RxNorm | 1650256 | insulin, regular, human 4 UNT |
| RxNorm | 1650260 | insulin, regular, human 4 UNT [Afrezza] |
| RxNorm | 1650262 | insulin, regular, human 8 UNT |
| RxNorm | 1650264 | insulin, regular, human 8 UNT [Afrezza] |
| RxNorm | 1653899 | insulin isophane / insulin, regular, human Injectable Product |
| RxNorm | 1654651 | insulin isophane / insulin, regular, human Pen Injector |
| RxNorm | 1654855 | insulin isophane / insulin, regular, human Pen Injector [Humulin] |
| RxNorm | 1654857 | insulin isophane, human 70 UNT/ML / insulin, regular, human 30 UNT/ML Pen Injector |
| RxNorm | 1654858 | insulin isophane, human 70 UNT/ML / insulin, regular, human 30 UNT/ML Pen Injector [Humulin] |
| RxNorm | 1654909 | insulin, regular, human 12 UNT |
| RxNorm | 1654910 | insulin, regular, human 12 UNT Inhalation Powder |
| RxNorm | 1654911 | insulin, regular, human 12 UNT [Afrezza] |
| RxNorm | 1654912 | insulin, regular, human 12 UNT Inhalation Powder [Afrezza] |
| RxNorm | 1656705 | {30 (insulin, regular, human 12 UNT Inhalation Powder) / 60 (insulin, regular, human 8 UNT Inhalation Powder) } Pack |
| RxNorm | 1656706 | {30 (insulin, regular, human 12 UNT Inhalation Powder [Afrezza]) / 60 (insulin, regular, human 8 UNT Inhalation Powder [Afrezza]) } Pack [Afrezza 90 - 60 (8 UNT), 30 (12 UNT)] |
| RxNorm | 1731314 | insulin, regular, human Pen Injector |
| RxNorm | 1731315 | 3 ML insulin, regular, human 500 UNT/ML Pen Injector |
| RxNorm | 1731316 | insulin, regular, human Pen Injector [Humulin R] |
| RxNorm | 1731317 | 3 ML insulin, regular, human 500 UNT/ML Pen Injector [Humulin R] |
| RxNorm | 1731318 | insulin, regular, human 500 UNT/ML Pen Injector |
| RxNorm | 1731319 | insulin, regular, human 500 UNT/ML Pen Injector [Humulin R] |
| RxNorm | 1798387 | {90 (insulin, regular, human 4 UNT Inhalation Powder) / 90 (insulin, regular, human 8 UNT Inhalation Powder) } Pack |
| RxNorm | 1798388 | {90 (insulin, regular, human 4 UNT Inhalation Powder [Afrezza]) / 90 (insulin, regular, human 8 UNT Inhalation Powder [Afrezza]) } Pack [Afrezza Titration Pack] |

Continued on next page

**Table17 – continued from previous page**

| CodeSystem | ConceptCode | ConceptName |
|---|---|---|
| RxNorm | 1862101 | {60 (insulin, regular, human 12 UNT Inhalation Powder) / 60 (insulin, regular, human 4 UNT Inhalation Powder) / 60 (insulin, regular, human 8 UNT Inhalation Powder) } Pack |
| RxNorm | 1862102 | {60 (insulin, regular, human 12 UNT Inhalation Powder [Afrezza]) / 60 (insulin, regular, human 4 UNT Inhalation Powder [Afrezza]) / 60 (insulin, regular, human 8 UNT Inhalation Powder [Afrezza]) } Pack [Afrezza 180 Titration Pack- 60 (4 UNT), 60 (8 UNT), 60 (12 UNT)] |
| RxNorm | 203209 | Human Actrapid Penfill |
| RxNorm | 2049379 | insulin isophane / insulin, regular, human Pen Injector [Novolin] |
| RxNorm | 2049380 | 3 ML insulin isophane, human 70 UNT/ML / insulin, regular, human 30 UNT/ML Pen Injector [Novolin] |
| RxNorm | 2049381 | insulin isophane, human 70 UNT/ML / insulin, regular, human 30 UNT/ML Pen Injector [Novolin] |
| RxNorm | 2100028 | {90 (insulin, regular, human 12 UNT Inhalation Powder) / 90 (insulin, regular, human 8 UNT Inhalation Powder) } Pack |
| RxNorm | 2100029 | {90 (insulin, regular, human 12 UNT Inhalation Powder [Afrezza]) / 90 (insulin, regular, human 8 UNT Inhalation Powder [Afrezza]) } Pack [Afrezza 180 - 90 (8 UNT), 90 (12 UNT)] |
| RxNorm | 2108525 | insulin, regular, human Inhalation Powder |
| RxNorm | 2108527 | insulin, regular, human Inhalation Powder [Afrezza] |
| RxNorm | 847186 | insulin isophane / insulin, regular, human Prefilled Syringe |
| RxNorm | 847187 | 3 ML insulin isophane, human 70 UNT/ML / insulin, regular, human 30 UNT/ML Pen Injector |
| RxNorm | 847188 | insulin isophane / insulin, regular, human Prefilled Syringe [Humulin] |
| RxNorm | 847189 | 3 ML insulin isophane, human 70 UNT/ML / insulin, regular, human 30 UNT/ML Pen Injector [Humulin] |
| RxNorm | 847194 | insulin isophane / insulin, regular, human Prefilled Syringe [Novolin] |
| RxNorm | 847202 | insulin, regular, human Prefilled Syringe |
| RxNorm | 847203 | 3 ML insulin, regular, human 100 UNT/ML Prefilled Syringe |
| RxNorm | 847204 | insulin, regular, human Prefilled Syringe [Novolin R] |
| RxNorm | 847205 | 3 ML insulin, regular, human 100 UNT/ML Prefilled Syringe [Novolin R] |
| RxNorm | 847256 | 1.5 ML insulin isophane, human 70 UNT/ML / insulin, regular, human 30 UNT/ML Prefilled Syringe |
| RxNorm | 847257 | 1.5 ML insulin isophane, human 70 UNT/ML / insulin, regular, human 30 UNT/ML Prefilled Syringe [Novolin] |

Continued on next page

**Table17 – continued from previous page**

| CodeSystem | ConceptCode | ConceptName |
|---|---|---|
| RxNorm | 847343 | 1.5 ML insulin isophane, human 70 UNT/ML / insulin, regular, human 30 UNT/ML Prefilled Syringe [Humulin] |
| RxNorm | 847417 | 1.5 ML insulin, regular, human 100 UNT/ML Prefilled Syringe |
| RxNorm | 92881 | Humulin R |
| RxNorm | 93560 | Novolin R |
| RxNorm | 575133 | Insulin, Isophane, Human 50 UNT/ML / Insulin, Regular, Human 50 UNT/ML [Humulin 50/50] |
| RxNorm | 106893 | Insulin, Isophane, Human 70 UNT/ML / Insulin, Regular, Human 30 UNT/ML Injectable Suspension [Human Mixtard 30] |
| RxNorm | 108812 | Insulin, Isophane, Human 70 UNT/ML / Insulin, Regular, Human 30 UNT/ML Injectable Suspension [Human Mixtard 30 ge] |
| RxNorm | 847195 | 3 ML insulin human, isophane 70 UNT/ML / Regular Insulin, Human 30 UNT/ML Prefilled Syringe [Novolin] |
| RxNorm | 213441 | Insulin, Isophane, Human 70 UNT/ML / Insulin, Regular, Human 30 UNT/ML Injectable Suspension [Humulin 70/30] |
| RxNorm | 153122 | Insulin, Isophane, Human 50 UNT/ML / Insulin, Regular, Human 50 UNT/ML Injectable Suspension [Human Mixtard 50] |
| RxNorm | 311016 | Insulin, Isophane, Human 50 UNT/ML / Insulin, Regular, Human 50 UNT/ML Injectable Suspension [Humulin 50/50] |
| RxNorm | 564604 | Insulin, Isophane, Human 50 UNT/ML / Insulin, Regular, Human 50 UNT/ML [Human Mixtard Penfill] |
| RxNorm | 564394 | Insulin, Isophane, Human 70 UNT/ML / Insulin, Regular, Human 30 UNT/ML [Human Mixtard 30] |
| RxNorm | 564600 | Insulin, Isophane, Human 70 UNT/ML / Insulin, Regular, Human 30 UNT/ML [Human Mixtard 30 ge] |
| RxNorm | 564393 | Insulin, Isophane, Human 70 UNT/ML / Insulin, Regular, Human 30 UNT/ML [Humulin M3] |
| RxNorm | 1156396 | NPH Insulin, Human / Regular Insulin, Human Injectable Product |
| RxNorm | 1172689 | Humulin M4 Injectable Product |
| RxNorm | 1171298 | Human Mixtard 50 Injectable Product |
| RxNorm | 1172690 | Humulin M5 Injectable Product |
| RxNorm | 1183567 | Pur-In Mix 50/50 Injectable Product |
| RxNorm | 1183565 | Pur-In Mix 15/85 Injectable Product |
| RxNorm | 1172683 | Humulin 70/30 Injectable Product |
| RxNorm | 1172686 | Humulin M1 Injectable Product |
| RxNorm | 1171299 | Human Mixtard Penfill Injectable Product |
| RxNorm | 1171297 | Human Mixtard 30 ge Injectable Product |

Continued on next page

**Table17 – continued from previous page**

| CodeSystem | ConceptCode | ConceptName |
|---|---|---|
| RxNorm | 213442 | insulin isophane, human 70 UNT/ML / insulin, regular, human 30 UNT/ML Injectable Suspension [Novolin] |
| RxNorm | 2179742 | insulin, regular, human 1 UNT/ML |
| RxNorm | 2179743 | insulin, regular, human Injection |
| RxNorm | 2179744 | 100 ML insulin, regular, human 1 UNT/ML Injection |
| RxNorm | 2179745 | Myxredlin |
| RxNorm | 2179746 | insulin, regular, human 1 UNT/ML [Myxredlin] |
| RxNorm | 2179747 | insulin, regular, human Injection [Myxredlin] |
| RxNorm | 2179748 | Myxredlin Injectable Product |
| RxNorm | 2179749 | 100 ML insulin, regular, human 1 UNT/ML Injection [Myxredlin] |
| RxNorm | 2179750 | insulin, regular, human 1 UNT/ML Injection |
| RxNorm | 2179751 | insulin, regular, human 1 UNT/ML Injection [Myxredlin] |
| RxNorm | 2206090 | 3 ML insulin, regular, human 100 UNT/ML Pen Injector |
| RxNorm | 2206091 | insulin, regular, human Pen Injector [Novolin R] |
| RxNorm | 2206092 | 3 ML insulin, regular, human 100 UNT/ML Pen Injector [Novolin R] |
| RxNorm | 2206093 | insulin, regular, human 100 UNT/ML Pen Injector |
| RxNorm | 2206094 | insulin, regular, human 100 UNT/ML Pen Injector [Novolin R] |
| RxNorm | 247511 | insulin isophane, human 90 UNT/ML / insulin, regular, human 10 UNT/ML Injectable Suspension |
| RxNorm | 247512 | insulin isophane, human 80 UNT/ML / insulin, regular, human 20 UNT/ML Injectable Suspension |
| RxNorm | 247513 | insulin isophane, human 60 UNT/ML / insulin, regular, human 40 UNT/ML Injectable Suspension |
| RxNorm | 249026 | insulin, regular, human 40 UNT/ML Injectable Solution |
| RxNorm | 245264 | insulin isophane, human 85 UNT/ML / insulin, regular, human 15 UNT/ML Injectable Suspension |
| RxNorm | 245265 | insulin isophane, human 50 UNT/ML / insulin, regular, human 50 UNT/ML Injectable Suspension |
| RxNorm | 249220 | insulin, regular, human 500 UNT/ML Injectable Solution |
| RxNorm | 311033 | insulin, regular, human 100 UNT/ML Injectable Solution [Novolin R] |
| RxNorm | 311034 | insulin, regular, human 100 UNT/ML Injectable Solution |
| RxNorm | 311035 | insulin, regular, human 100 UNT/ML Injectable Solution [Velosulin] |
| RxNorm | 311036 | insulin, regular, human 100 UNT/ML Injectable Solution [Humulin R] |
| RxNorm | 311048 | insulin isophane, human 70 UNT/ML / insulin, regular, human 30 UNT/ML Injectable Suspension |
| RxNorm | 340325 | insulin, regular, human 100 UNT/ML |
| RxNorm | 340326 | insulin, regular, human 40 UNT/ML |

Continued on next page

**Table17 – continued from previous page**

| CodeSystem | ConceptCode | ConceptName |
|---|---|---|
| RxNorm | 340327 | insulin, regular, human 500 UNT/ML |
| RxNorm | 351859 | insulin, regular, human 500 UNT/ML Injectable Solution [Humulin R] |
| RxNorm | 362622 | insulin, regular, human Injectable Solution [Humulin R] |
| RxNorm | 363120 | insulin, regular, human Injectable Solution [Velosulin] |
| RxNorm | 363221 | insulin, regular, human Injectable Solution [Novolin R] |
| RxNorm | 343083 | insulin, regular, human 30 UNT/ML |
| RxNorm | 343258 | insulin, regular, human 50 UNT/ML |
| RxNorm | 360894 | insulin, regular, human 25 UNT/ML |
| RxNorm | 359125 | insulin, regular, human 15 UNT/ML |
| RxNorm | 359126 | insulin, regular, human 10 UNT/ML |
| RxNorm | 359127 | insulin, regular, human 20 UNT/ML |
| RxNorm | 365668 | insulin isophane / insulin, regular, human Injectable Suspension [Pur-In Mix] |
| RxNorm | 365672 | insulin isophane / insulin, regular, human Injectable Suspension [Novolin] |
| RxNorm | 365679 | insulin isophane / insulin, regular, human Injectable Suspension [Humulin] |
| RxNorm | 392660 | insulin isophane, human 75 UNT/ML / insulin, regular, human 25 UNT/ML Injectable Suspension |
| RxNorm | 378857 | insulin isophane / insulin, regular, human Injectable Suspension |
| RxNorm | 376915 | insulin, regular, human Injectable Solution |
| RxNorm | 412978 | insulin isophane, human 16 UNT/ML / insulin, regular, human 24 UNT/ML Injectable Suspension |
| RxNorm | 379734 | insulin, regular, human Injectable Solution [Humulin S] |
| RxNorm | 379740 | insulin isophane / insulin, regular, human Injectable Suspension [Human Mixtard] |
| RxNorm | 379744 | insulin, regular, human Injectable Solution [Human Actrapid] |
| RxNorm | 379747 | insulin, regular, human Injectable Solution [Human Actrapid Penfill] |
| RxNorm | 385895 | Pur-In Neutral |
| RxNorm | 385896 | insulin, regular, human Injectable Solution [Pur-In Neutral] |
| RxNorm | 412453 | insulin, regular, human 5 UNT/ML Injectable Solution |
| RxNorm | 415088 | insulin isophane, human 28 UNT/ML / insulin, regular, human 12 UNT/ML Injectable Suspension |
| RxNorm | 415089 | insulin isophane, human 30 UNT/ML / insulin, regular, human 10 UNT/ML Injectable Suspension |
| RxNorm | 415090 | insulin isophane, human 34 UNT/ML / insulin, regular, human 6 UNT/ML Injectable Suspension |
| RxNorm | 415184 | insulin isophane, human 32 UNT/ML / insulin, regular, human 8 UNT/ML Injectable Suspension |

Continued on next page

**Table17 – continued from previous page**

| CodeSystem | ConceptCode | ConceptName |
|---|---|---|
| RxNorm | 415185 | insulin isophane, human 36 UNT/ML / insulin, regular, human 4 UNT/ML Injectable Suspension |
| RxNorm | 440399 | insulin, regular, human 24 UNT/ML |
| RxNorm | 440650 | insulin, regular, human 12 UNT/ML |
| RxNorm | 440653 | insulin, regular, human 6 UNT/ML |
| RxNorm | 440654 | insulin, regular, human 5 UNT/ML |
| RxNorm | 451437 | insulin, regular, human 8 UNT/ML |
| RxNorm | 451439 | insulin, regular, human 4 UNT/ML |
| RxNorm | 564395 | insulin isophane, human 85 UNT/ML / insulin, regular, human 15 UNT/ML [Pur-In Mix] |
| RxNorm | 564396 | insulin isophane, human 75 UNT/ML / insulin, regular, human 25 UNT/ML [Pur-In Mix] |
| RxNorm | 564397 | insulin isophane, human 50 UNT/ML / insulin, regular, human 50 UNT/ML [Pur-In Mix] |
| RxNorm | 564399 | insulin isophane, human 90 UNT/ML / insulin, regular, human 10 UNT/ML [Humulin] |
| RxNorm | 564400 | insulin isophane, human 80 UNT/ML / insulin, regular, human 20 UNT/ML [Humulin] |
| RxNorm | 564401 | insulin isophane, human 60 UNT/ML / insulin, regular, human 40 UNT/ML [Humulin] |
| RxNorm | 5459 | Humulin S |
| RxNorm | 564531 | insulin isophane, human 50 UNT/ML / insulin, regular, human 50 UNT/ML [Humulin] |
| RxNorm | 564601 | insulin isophane, human 90 UNT/ML / insulin, regular, human 10 UNT/ML [Human Mixtard] |
| RxNorm | 564602 | insulin isophane, human 80 UNT/ML / insulin, regular, human 20 UNT/ML [Human Mixtard] |
| RxNorm | 564603 | insulin isophane, human 70 UNT/ML / insulin, regular, human 30 UNT/ML [Human Mixtard] |
| RxNorm | 564605 | insulin isophane, human 60 UNT/ML / insulin, regular, human 40 UNT/ML [Human Mixtard] |
| RxNorm | 564766 | insulin, regular, human 100 UNT/ML [Pur-In Neutral] |
| RxNorm | 564820 | insulin, regular, human 100 UNT/ML [Human Actrapid Penfill] |
| RxNorm | 564881 | insulin, regular, human 100 UNT/ML [Human Actrapid] |
| RxNorm | 564882 | insulin, regular, human 100 UNT/ML [Humulin S] |
| RxNorm | 565176 | insulin isophane, human 50 UNT/ML / insulin, regular, human 50 UNT/ML [Human Mixtard] |
| RxNorm | 575146 | insulin, regular, human 100 UNT/ML [Novolin R] |
| RxNorm | 575147 | insulin, regular, human 100 UNT/ML [Velosulin] |
| RxNorm | 575148 | insulin, regular, human 100 UNT/ML [Humulin R] |
| RxNorm | 575628 | insulin, regular, human 500 UNT/ML [Humulin R] |

Continued on next page

**Table17 – continued from previous page**

| CodeSystem | ConceptCode | ConceptName |
|---|---|---|
| RxNorm | 573330 | insulin isophane, human 70 UNT/ML / insulin, regular, human 30 UNT/ML [Humulin] |
| RxNorm | 573331 | insulin isophane, human 70 UNT/ML / insulin, regular, human 30 UNT/ML [Novolin] |

##### 0.5.3 Sulfonylurea

Table 18: Concept codes used to identify Sulfonylurea medications.

| CodeSystem | ConceptCode | ConceptName |
|---|---|---|
| RxNorm | 10633 | tolazamide |
| RxNorm | 25789 | glimepiride |
| RxNorm | 4815 | glyburide |
| RxNorm | 4821 | glipizide |
| RxNorm | 368567 | Glipizide / Metformin Oral Tablet [metaglip] |
| RxNorm | 352301 | Glipizide 5 MG / Metformin 500 MG Oral Tablet [metaglip] |
| RxNorm | 352302 | Glipizide 2.5 MG / Metformin 250 MG Oral Tablet [metaglip] |
| RxNorm | 379801 | Glipizide 24 Hour Extended Release Tablet [Glucotrol XL] |
| RxNorm | 369372 | Glyburide Oral Tablet [Glynase Pres-Tab] |
| RxNorm | 379805 | Glipizide 24 Hour Extended Release Tablet |
| RxNorm | 368169 | Glyburide / Metformin Oral Tablet [Glucovance] |
| RxNorm | 352300 | Glipizide 2.5 MG / Metformin 500 MG Oral Tablet [metaglip] |
| RxNorm | 541898 | Glyburide 1.5 MG Oral Tablet [Glyguride] |
| RxNorm | 542065 | Glyburide 6 MG [Gyburide] |
| RxNorm | 575998 | Glipizide 2.5 MG / Metformin 250 MG [metaglip] |
| RxNorm | 575996 | Glipizide 2.5 MG / Metformin 500 MG [metaglip] |
| RxNorm | 541896 | Glyburide 1.5 MG [Glyguride] |
| RxNorm | 542067 | Glyburide 6 MG Oral Tablet [Gyburide] |
| RxNorm | 542066 | Glyburide Oral Tablet [Gyburide] |
| RxNorm | 541897 | Glyburide Oral Tablet [Glyguride] |
| RxNorm | 575997 | Glipizide 5 MG / Metformin 500 MG [metaglip] |
| RxNorm | 615211 | glimepiride / rosiglitazone Oral Tablet [Avandaryl] |
| RxNorm | 668563 | glimepiride 4 MG / pioglitazone 30 MG Oral Tablet [Duetact] |
| RxNorm | 706933 | glimepiride 2 MG / rosiglitazone 8 MG Oral Tablet [Avandaryl] |
| RxNorm | 668555 | glimepiride / pioglitazone Oral Tablet [Duetact] |
| RxNorm | 615918 | glimepiride 4 MG / rosiglitazone 4 MG [Avandaryl] |

Continued on next page

**Table18 – continued from previous page**

| CodeSystem | ConceptCode | ConceptName |
|---|---|---|
| RxNorm | 615212 | glimepiride 1 MG / rosiglitazone 4 MG Oral Tablet [Avandaryl] |
| RxNorm | 668554 | glimepiride 2 MG / pioglitazone 30 MG [Duetact] |
| RxNorm | 668562 | glimepiride 4 MG / pioglitazone 30 MG [Duetact] |
| RxNorm | 700836 | Glipizide 5 MG Extended Release Tablet |
| RxNorm | 706932 | glimepiride 2 MG / rosiglitazone 8 MG [Avandaryl] |
| RxNorm | 731456 | glimepiride / pioglitazone Oral Tablet [Duetact 30/4] |
| RxNorm | 615214 | glimepiride 2 MG / rosiglitazone 4 MG Oral Tablet [Avandaryl] |
| RxNorm | 615919 | glimepiride 4 MG / rosiglitazone 4 MG Oral Tablet [Avandaryl] |
| RxNorm | 615210 | glimepiride 1 MG / rosiglitazone 4 MG [Avandaryl] |
| RxNorm | 668556 | glimepiride 2 MG / pioglitazone 30 MG Oral Tablet [Duetact] |
| RxNorm | 847723 | glimepiride / rosiglitazone Oral Tablet [Avandaryl 8/4] |
| RxNorm | 847719 | glimepiride / rosiglitazone Oral Tablet [Avandaryl 8/2] |
| RxNorm | 707181 | glimepiride 4 MG / rosiglitazone 8 MG [Avandaryl] |
| RxNorm | 615213 | glimepiride 2 MG / rosiglitazone 4 MG [Avandaryl] |
| RxNorm | 707182 | glimepiride 4 MG / rosiglitazone 8 MG Oral Tablet [Avandaryl] |
| RxNorm | 849593 | Glipizide / Metformin Oral Tablet [Metaglip 2.5 MG/500 MG] |
| RxNorm | 847715 | glimepiride / rosiglitazone Oral Tablet [Avandaryl 4/1] |
| RxNorm | 847711 | glimepiride / rosiglitazone Oral Tablet [Avandaryl 4/2] |
| RxNorm | 849589 | Glipizide / Metformin Oral Tablet [Metaglip 5 MG/500 MG] |
| RxNorm | 861746 | Glyburide / Metformin Oral Tablet [Glucovance 1.25 MG/250 MG] |
| RxNorm | 861751 | Glyburide / Metformin Oral Tablet [Glucovance 2.5 MG/500 MG] |
| RxNorm | 1185627 | Metaglip 5 MG/500 MG Oral Product |
| RxNorm | 1175663 | Avandaryl 8/2 Pill |
| RxNorm | 1175657 | Avandaryl 4/1 Pill |
| RxNorm | 1175661 | Avandaryl 4/4 Pill |
| RxNorm | 1008873 | glyburide / phenformin |
| RxNorm | 102845 | glyburide 5 MG Oral Tablet [Daonil] |
| RxNorm | 105371 | glyburide 2.5 MG Oral Tablet [Calabren] |
| RxNorm | 105372 | glyburide 5 MG Oral Tablet [Calabren] |
| RxNorm | 105373 | glipizide 5 MG Oral Tablet [Minidiab] |
| RxNorm | 1157121 | tolazamide Oral Product |
| RxNorm | 1157122 | tolazamide Pill |
| RxNorm | 1156197 | glyburide / metformin Pill |
| RxNorm | 1156198 | glyburide / phenformin Oral Product |

Continued on next page

**Table18 – continued from previous page**

| CodeSystem | ConceptCode | ConceptName |
|---|---|---|
| RxNorm | 1156199 | glyburide / phenformin Pill |
| RxNorm | 1156200 | glyburide Oral Product |
| RxNorm | 1156201 | glyburide Pill |
| RxNorm | 1157240 | glimepiride / pioglitazone Oral Product |
| RxNorm | 1157241 | glimepiride / pioglitazone Pill |
| RxNorm | 1157242 | glimepiride / rosiglitazone Oral Product |
| RxNorm | 1157243 | glimepiride / rosiglitazone Pill |
| RxNorm | 1157244 | glimepiride Oral Product |
| RxNorm | 1157245 | glimepiride Pill |
| RxNorm | 1166403 | Duetact Oral Product |
| RxNorm | 1166404 | Duetact Pill |
| RxNorm | 1165205 | glipizide / metformin Oral Product |
| RxNorm | 1165206 | glipizide / metformin Pill |
| RxNorm | 1165207 | glipizide Oral Product |
| RxNorm | 1165208 | glipizide Pill |
| RxNorm | 1165845 | glyburide / metformin Oral Product |
| RxNorm | 1170663 | Amaryl Oral Product |
| RxNorm | 1170664 | Amaryl Pill |
| RxNorm | 1170866 | Calabren Oral Product |
| RxNorm | 1170867 | Calabren Pill |
| RxNorm | 1171148 | Euglucon Oral Product |
| RxNorm | 1171149 | Euglucon Pill |
| RxNorm | 1171233 | Glibenese Oral Product |
| RxNorm | 1171234 | Glibenese Pill |
| RxNorm | 1171246 | Glucotrol Oral Product |
| RxNorm | 1171247 | Glucotrol Pill |
| RxNorm | 1171248 | Glucovance Oral Product |
| RxNorm | 1171249 | Glucovance Pill |
| RxNorm | 1171929 | Glycron Oral Product |
| RxNorm | 1171930 | Glycron Pill |
| RxNorm | 1171933 | Glynase Oral Product |
| RxNorm | 1171934 | Glynase Pill |
| RxNorm | 1171919 | Glyburase Oral Product |
| RxNorm | 1171920 | Glyburase Pill |
| RxNorm | 1175658 | Avandaryl Oral Product |
| RxNorm | 1175659 | Avandaryl Pill |
| RxNorm | 1175878 | Diabeta Oral Product |
| RxNorm | 1175879 | Diabeta Pill |
| RxNorm | 1175880 | Diabetamide Oral Product |
| RxNorm | 1175881 | Diabetamide Pill |
| RxNorm | 1168162 | Daonil Oral Product |
| RxNorm | 1168163 | Daonil Pill |
| RxNorm | 1179291 | Minidiab Oral Product |

Continued on next page

**Table18 – continued from previous page**

| CodeSystem | ConceptCode | ConceptName |
|---|---|---|
| RxNorm | 1179292 | Minidiab Pill |
| RxNorm | 1185049 | Metaglip Oral Product |
| RxNorm | 1179740 | Malix Oral Product |
| RxNorm | 1179741 | Malix Pill |
| RxNorm | 1177973 | Tolinase Oral Product |
| RxNorm | 1177974 | Tolinase Pill |
| RxNorm | 1186169 | Libanil Oral Product |
| RxNorm | 1186170 | Libanil Pill |
| RxNorm | 1178082 | Micronase Oral Product |
| RxNorm | 1178083 | Micronase Pill |
| RxNorm | 1178368 | Semi-Daonil Oral Product |
| RxNorm | 1178369 | Semi-Daonil Pill |
| RxNorm | 1185624 | Metaglip Pill |
| RxNorm | 1361492 | glimepiride 6 MG |
| RxNorm | 1361493 | glimepiride 6 MG Oral Tablet |
| RxNorm | 1361494 | glimepiride 8 MG |
| RxNorm | 1361495 | glimepiride 8 MG Oral Tablet |
| RxNorm | 151466 | Calabren |
| RxNorm | 151615 | Diabetamide |
| RxNorm | 151725 | Euglucon |
| RxNorm | 151822 | Glibenese |
| RxNorm | 153842 | glimepiride 3 MG Oral Tablet |
| RxNorm | 153843 | glimepiride 1 MG Oral Tablet [Amaryl] |
| RxNorm | 153844 | glimepiride 3 MG Oral Tablet [Amaryl] |
| RxNorm | 153845 | glimepiride 4 MG Oral Tablet [Amaryl] |
| RxNorm | 152651 | Malix |
| RxNorm | 153591 | glimepiride 2 MG Oral Tablet [Amaryl] |
| RxNorm | 153592 | Amaryl |
| RxNorm | 197737 | glyburide 1.25 MG Oral Tablet |
| RxNorm | 199245 | glimepiride 1 MG Oral Tablet |
| RxNorm | 199246 | glimepiride 2 MG Oral Tablet |
| RxNorm | 199247 | glimepiride 4 MG Oral Tablet |
| RxNorm | 198291 | tolazamide 100 MG Oral Tablet |
| RxNorm | 198292 | tolazamide 250 MG Oral Tablet |
| RxNorm | 198293 | tolazamide 500 MG Oral Tablet |
| RxNorm | 203289 | Micronase |
| RxNorm | 203295 | Diabeta |
| RxNorm | 203296 | Daonil |
| RxNorm | 201921 | glipizide 5 MG Oral Tablet [Glibenese] |
| RxNorm | 201922 | glipizide 2.5 MG Oral Tablet [Minidiab] |
| RxNorm | 203679 | Minidiab |
| RxNorm | 203680 | Glucotrol |
| RxNorm | 201056 | glyburide 2.5 MG Oral Tablet [Libanil] |

Continued on next page

**Table18 – continued from previous page**

| CodeSystem | ConceptCode | ConceptName |
|---|---|---|
| RxNorm | 201057 | glyburide 2.5 MG Oral Tablet [Semi-Daonil] |
| RxNorm | 201058 | glyburide 2.5 MG Oral Tablet [Euglucon] |
| RxNorm | 201059 | glyburide 2.5 MG Oral Tablet [Malix] |
| RxNorm | 201060 | glyburide 2.5 MG Oral Tablet [Diabetamide] |
| RxNorm | 201061 | glyburide 5 MG Oral Tablet [Libanil] |
| RxNorm | 201062 | glyburide 5 MG Oral Tablet [Euglucon] |
| RxNorm | 201063 | glyburide 5 MG Oral Tablet [Malix] |
| RxNorm | 201064 | glyburide 5 MG Oral Tablet [Diabetamide] |
| RxNorm | 205828 | glipizide 5 MG Oral Tablet [Glucotrol] |
| RxNorm | 205830 | glipizide 10 MG Oral Tablet [Glucotrol] |
| RxNorm | 205872 | glyburide 1.25 MG Oral Tablet [Diabeta] |
| RxNorm | 205873 | glyburide 1.25 MG Oral Tablet [Micronase] |
| RxNorm | 205875 | glyburide 2.5 MG Oral Tablet [Diabeta] |
| RxNorm | 205876 | glyburide 2.5 MG Oral Tablet [Micronase] |
| RxNorm | 205879 | glyburide 5 MG Oral Tablet [Diabeta] |
| RxNorm | 205880 | glyburide 5 MG Oral Tablet [Micronase] |
| RxNorm | 207953 | tolazamide 100 MG Oral Tablet [Tolinase] |
| RxNorm | 207954 | tolazamide 250 MG Oral Tablet [Tolinase] |
| RxNorm | 207955 | tolazamide 500 MG Oral Tablet [Tolinase] |
| RxNorm | 844809 | glipizide 2.5 MG Extended Release Oral Tablet |
| RxNorm | 844824 | glipizide 5 MG Extended Release Oral Tablet |
| RxNorm | 844827 | glipizide 10 MG Extended Release Oral Tablet |
| RxNorm | 849585 | glipizide / metformin Oral Tablet [Metaglip] |
| RxNorm | 847708 | glimepiride 4 MG / rosiglitazone 4 MG Oral Tablet [Avandaryl] |
| RxNorm | 847706 | glimepiride 4 MG / rosiglitazone 4 MG [Avandaryl] |
| RxNorm | 847707 | glimepiride / rosiglitazone Oral Tablet [Avandaryl] |
| RxNorm | 847710 | glimepiride 2 MG / rosiglitazone 4 MG [Avandaryl] |
| RxNorm | 847712 | glimepiride 2 MG / rosiglitazone 4 MG Oral Tablet [Avandaryl] |
| RxNorm | 847714 | glimepiride 1 MG / rosiglitazone 4 MG [Avandaryl] |
| RxNorm | 847716 | glimepiride 1 MG / rosiglitazone 4 MG Oral Tablet [Avandaryl] |
| RxNorm | 847718 | glimepiride 2 MG / rosiglitazone 8 MG [Avandaryl] |
| RxNorm | 847720 | glimepiride 2 MG / rosiglitazone 8 MG Oral Tablet [Avandaryl] |
| RxNorm | 847722 | glimepiride 4 MG / rosiglitazone 8 MG [Avandaryl] |
| RxNorm | 847724 | glimepiride 4 MG / rosiglitazone 8 MG Oral Tablet [Avandaryl] |
| RxNorm | 861731 | glipizide 2.5 MG / metformin hydrochloride 250 MG Oral Tablet |
| RxNorm | 861732 | glipizide 2.5 MG / metformin hydrochloride 250 MG [Metaglip] |

Continued on next page

**Table18 – continued from previous page**

| CodeSystem | ConceptCode | ConceptName |
|---|---|---|
| RxNorm | 861733 | glipizide 2.5 MG / metformin hydrochloride 250 MG Oral Tablet [Metaglip] |
| RxNorm | 861736 | glipizide 2.5 MG / metformin hydrochloride 500 MG Oral Tablet |
| RxNorm | 861737 | glipizide 2.5 MG / metformin hydrochloride 500 MG [Metaglip] |
| RxNorm | 861738 | glipizide 2.5 MG / metformin hydrochloride 500 MG Oral Tablet [Metaglip] |
| RxNorm | 861740 | glipizide 5 MG / metformin hydrochloride 500 MG Oral Tablet |
| RxNorm | 861741 | glipizide 5 MG / metformin hydrochloride 500 MG [Metaglip] |
| RxNorm | 861742 | glipizide 5 MG / metformin hydrochloride 500 MG Oral Tablet [Metaglip] |
| RxNorm | 861743 | glyburide 1.25 MG / metformin hydrochloride 250 MG Oral Tablet |
| RxNorm | 861745 | glyburide 1.25 MG / metformin hydrochloride 250 MG [Glucovance] |
| RxNorm | 861747 | glyburide 1.25 MG / metformin hydrochloride 250 MG Oral Tablet [Glucovance] |
| RxNorm | 861748 | glyburide 2.5 MG / metformin hydrochloride 500 MG Oral Tablet |
| RxNorm | 861750 | glyburide 2.5 MG / metformin hydrochloride 500 MG [Glucovance] |
| RxNorm | 861752 | glyburide 2.5 MG / metformin hydrochloride 500 MG Oral Tablet [Glucovance] |
| RxNorm | 861753 | glyburide 5 MG / metformin hydrochloride 500 MG Oral Tablet |
| RxNorm | 861755 | glyburide 5 MG / metformin hydrochloride 500 MG [Glucovance] |
| RxNorm | 861756 | glyburide / metformin Oral Tablet [Glucovance] |
| RxNorm | 861757 | glyburide 5 MG / metformin hydrochloride 500 MG Oral Tablet [Glucovance] |
| RxNorm | 881404 | Glynase |
| RxNorm | 881405 | glyburide 1.5 MG [Glynase] |
| RxNorm | 881406 | glyburide Oral Tablet [Glynase] |
| RxNorm | 881407 | glyburide 1.5 MG Oral Tablet [Glynase] |
| RxNorm | 881408 | glyburide 3 MG [Glynase] |
| RxNorm | 881409 | glyburide 3 MG Oral Tablet [Glynase] |
| RxNorm | 881410 | glyburide 6 MG [Glynase] |
| RxNorm | 881411 | glyburide 6 MG Oral Tablet [Glynase] |
| RxNorm | 865567 | glipizide Extended Release Oral Tablet [Glucotrol] |

Continued on next page

**Table18 – continued from previous page**

| CodeSystem | ConceptCode | ConceptName |
|---|---|---|
| RxNorm | 865568 | 24 HR glipizide 10 MG Extended Release Oral Tablet [Glucotrol] |
| RxNorm | 865569 | glipizide 10 MG Extended Release Oral Tablet [Glucotrol] |
| RxNorm | 865570 | glipizide 2.5 MG [Glucotrol] |
| RxNorm | 865571 | 24 HR glipizide 2.5 MG Extended Release Oral Tablet [Glucotrol] |
| RxNorm | 865572 | glipizide 2.5 MG Extended Release Oral Tablet [Glucotrol] |
| RxNorm | 865573 | 24 HR glipizide 5 MG Extended Release Oral Tablet [Glucotrol] |
| RxNorm | 865574 | glipizide 5 MG Extended Release Oral Tablet [Glucotrol] |
| RxNorm | 93312 | glyburide Oral Tablet [Micronase] |
| RxNorm | 284433 | Glyburide 5 MG / Metformin 500 MG Oral Tablet [Glucovance] |
| RxNorm | 284432 | Glyburide 2.5 MG / Metformin 500 MG Oral Tablet [Glucovance] |
| RxNorm | 351273 | Glipizide 2.5 MG / Metformin 500 MG Oral Tablet |
| RxNorm | 351275 | Glipizide 2.5 MG / Metformin 250 MG Oral Tablet |
| RxNorm | 261311 | Glipizide 2.5 MG 24 Hour Extended Release Tablet [Glucotrol XL] |
| RxNorm | 205829 | Glipizide 5 MG 24 Hour Extended Release Tablet [Glucotrol XL] |
| RxNorm | 284431 | Glyburide 1.25 MG / Metformin 250 MG Oral Tablet [Glucovance] |
| RxNorm | 205881 | Glyburide 6 MG Oral Tablet [Glynase Pres-Tab] |
| RxNorm | 205877 | Glyburide 3 MG Oral Tablet [Glynase Pres-Tab] |
| RxNorm | 205831 | Glipizide 10 MG 24 Hour Extended Release Tablet [Glucotrol XL] |
| RxNorm | 205874 | Glyburide 1.5 MG Oral Tablet [Glynase Pres-Tab] |
| RxNorm | 351274 | Glipizide 5 MG / Metformin 500 MG Oral Tablet |
| RxNorm | 566719 | Glipizide 5 MG [Glucotrol XL] |
| RxNorm | 566721 | Glipizide 10 MG [Glucotrol XL] |
| RxNorm | 574533 | Glipizide 2.5 MG [Glucotrol XL] |
| RxNorm | 566763 | Glyburide 1.5 MG [Glynase Pres-Tab] |
| RxNorm | 574869 | Glyburide 1.25 MG / Metformin 250 MG [Glucovance] |
| RxNorm | 566770 | Glyburide 6 MG [Glynase Pres-Tab] |
| RxNorm | 574871 | Glyburide 5 MG / Metformin 500 MG [Glucovance] |
| RxNorm | 566766 | Glyburide 3 MG [Glynase Pres-Tab] |
| RxNorm | 574870 | Glyburide 2.5 MG / Metformin 500 MG [Glucovance] |
| RxNorm | 310538 | Glyburide 5 MG / Metformin 500 MG Oral Tablet |
| RxNorm | 310535 | Glyburide 2.5 MG / Metformin 500 MG Oral Tablet |
| RxNorm | 310533 | Glyburide 1.25 MG / Metformin 250 MG Oral Tablet |
| RxNorm | 1185628 | Metaglip 5 MG/500 MG Pill |
| RxNorm | 1175664 | Avandaryl 8/4 Oral Product |

Continued on next page

**Table18 – continued from previous page**

| CodeSystem | ConceptCode | ConceptName |
|---|---|---|
| RxNorm | 1166405 | Duetact 30/4 Oral Product |
| RxNorm | 1185625 | Metaglip 2.5 MG/500 MG Oral Product |
| RxNorm | 1171251 | Glucovance 2.5 MG/500 MG Pill |
| RxNorm | 1166406 | Duetact 30/4 Pill |
| RxNorm | 1185626 | Metaglip 2.5 MG/500 MG Pill |
| RxNorm | 1175660 | Avandaryl 4/4 Oral Product |
| RxNorm | 1175662 | Avandaryl 8/2 Oral Product |
| RxNorm | 1171253 | Glucovance 5 MG/500 MG Pill |
| RxNorm | 1171250 | Glucovance 2.5 MG/500 MG Oral Product |
| RxNorm | 1175665 | Avandaryl 8/4 Pill |
| RxNorm | 1171252 | Glucovance 5 MG/500 MG Oral Product |
| RxNorm | 1175656 | Avandaryl 4/1 Oral Product |
| RxNorm | 220338 | Tolinase |
| RxNorm | 225613 | Semi-Daonil |
| RxNorm | 246391 | glyburide 2.5 MG / phenformin 25 MG Oral Tablet |
| RxNorm | 246524 | glyburide 5 MG / phenformin 50 MG Oral Tablet |
| RxNorm | 250919 | glyburide 2.5 MG / metformin 400 MG Oral Tablet |
| RxNorm | 284743 | Glucovance |
| RxNorm | 261351 | glyburide 4.5 MG Oral Tablet [Glycron] |
| RxNorm | 252960 | glyburide 4.5 MG Oral Tablet |
| RxNorm | 285129 | glyburide / metformin |
| RxNorm | 261532 | Glycron |
| RxNorm | 261974 | glyburide 3 MG Oral Tablet [Glycron] |
| RxNorm | 260286 | glyburide 1.5 MG Oral Tablet [Glycron] |
| RxNorm | 260287 | glyburide 6 MG Oral Tablet [Glycron] |
| RxNorm | 310488 | glipizide 10 MG Oral Tablet |
| RxNorm | 310489 | 24 HR glipizide 2.5 MG Extended Release Oral Tablet |
| RxNorm | 310490 | glipizide 5 MG Oral Tablet |
| RxNorm | 310534 | glyburide 2.5 MG Oral Tablet |
| RxNorm | 310536 | glyburide 3 MG Oral Tablet |
| RxNorm | 310537 | glyburide 5 MG Oral Tablet |
| RxNorm | 310539 | glyburide 6 MG Oral Tablet |
| RxNorm | 313418 | tolazamide 250 MG Oral Capsule |
| RxNorm | 314000 | glyburide 1.5 MG Oral Tablet |
| RxNorm | 314006 | 24 HR glipizide 5 MG Extended Release Oral Tablet |
| RxNorm | 315107 | 24 HR glipizide 10 MG Extended Release Oral Tablet |
| RxNorm | 316832 | tolazamide 100 MG |
| RxNorm | 316833 | tolazamide 250 MG |
| RxNorm | 316834 | tolazamide 500 MG |
| RxNorm | 315978 | glimepiride 1 MG |
| RxNorm | 315979 | glimepiride 4 MG |
| RxNorm | 315980 | glipizide 5 MG |
| RxNorm | 315987 | glyburide 1.25 MG |

Continued on next page

**Table18 – continued from previous page**

| CodeSystem | ConceptCode | ConceptName |
|---|---|---|
| RxNorm | 315988 | glyburide 1.5 MG |
| RxNorm | 315989 | glyburide 2.5 MG |
| RxNorm | 315990 | glyburide 3 MG |
| RxNorm | 315991 | glyburide 5 MG |
| RxNorm | 315992 | glyburide 6 MG |
| RxNorm | 317379 | glipizide 10 MG |
| RxNorm | 317637 | glimepiride 2 MG |
| RxNorm | 331496 | glyburide 4.5 MG |
| RxNorm | 330349 | glipizide 2.5 MG |
| RxNorm | 352381 | glipizide / metformin |
| RxNorm | 352764 | Metaglip |
| RxNorm | 367762 | glimepiride Oral Tablet [Amaryl] |
| RxNorm | 368204 | glyburide Oral Tablet [Glycron] |
| RxNorm | 369304 | tolazamide Oral Tablet [Tolinase] |
| RxNorm | 369373 | glyburide Oral Tablet [Diabeta] |
| RxNorm | 369500 | glipizide Oral Tablet [Glucotrol] |
| RxNorm | 369557 | glyburide Oral Tablet [Semi-Daonil] |
| RxNorm | 369562 | glyburide Oral Tablet [Daonil] |
| RxNorm | 374635 | glyburide / metformin Oral Tablet |
| RxNorm | 375952 | tolazamide Oral Capsule |
| RxNorm | 379803 | glipizide Oral Tablet [Glibenese] |
| RxNorm | 379804 | glipizide 2.5 MG Oral Tablet |
| RxNorm | 380849 | glimepiride 3 MG |
| RxNorm | 372319 | glimepiride Oral Tablet |
| RxNorm | 372320 | glipizide Oral Tablet |
| RxNorm | 372333 | glyburide Oral Tablet |
| RxNorm | 372334 | glyburide / phenformin Oral Tablet |
| RxNorm | 378730 | glipizide / metformin Oral Tablet |
| RxNorm | 374149 | tolazamide Oral Tablet |
| RxNorm | 379559 | glyburide Oral Tablet [Euglucon] |
| RxNorm | 379565 | glyburide Oral Tablet [Diabetamide] |
| RxNorm | 379568 | glyburide Oral Tablet [Malix] |
| RxNorm | 379570 | glyburide Oral Tablet [Calabren] |
| RxNorm | 379571 | Libanil |
| RxNorm | 379572 | glyburide Oral Tablet [Libanil] |
| RxNorm | 379802 | glipizide Oral Tablet [Minidiab] |
| RxNorm | 429841 | glyburide 5 MG / metformin 1000 MG Oral Tablet |
| RxNorm | 430102 | glyburide 3.5 MG Oral Tablet |
| RxNorm | 430103 | glyburide 1.75 MG Oral Tablet |
| RxNorm | 430104 | glyburide 1 MG Oral Tablet |
| RxNorm | 440285 | glyburide 1.75 MG |
| RxNorm | 440286 | glyburide 1 MG |
| RxNorm | 440287 | glyburide 3.5 MG |

Continued on next page

**Table18 – continued from previous page**

| CodeSystem | ConceptCode | ConceptName |
|---|---|---|
| RxNorm | 563154 | glyburide 5 MG [Daonil] |
| RxNorm | 566056 | glipizide 5 MG [Glibenese] |
| RxNorm | 566057 | glipizide 2.5 MG [Minidiab] |
| RxNorm | 565327 | glimepiride 2 MG [Amaryl] |
| RxNorm | 565408 | glimepiride 1 MG [Amaryl] |
| RxNorm | 565409 | glimepiride 3 MG [Amaryl] |
| RxNorm | 565410 | glimepiride 4 MG [Amaryl] |
| RxNorm | 568684 | tolazamide 100 MG [Tolinase] |
| RxNorm | 568685 | tolazamide 250 MG [Tolinase] |
| RxNorm | 568686 | tolazamide 500 MG [Tolinase] |
| RxNorm | 564036 | glyburide 2.5 MG [Calabren] |
| RxNorm | 564037 | glyburide 5 MG [Calabren] |
| RxNorm | 564038 | glipizide 5 MG [Minidiab] |
| RxNorm | 566718 | glipizide 5 MG [Glucotrol] |
| RxNorm | 566720 | glipizide 10 MG [Glucotrol] |
| RxNorm | 566761 | glyburide 1.25 MG [Diabeta] |
| RxNorm | 566762 | glyburide 1.25 MG [Micronase] |
| RxNorm | 566764 | glyburide 2.5 MG [Diabeta] |
| RxNorm | 566765 | glyburide 2.5 MG [Micronase] |
| RxNorm | 566768 | glyburide 5 MG [Diabeta] |
| RxNorm | 566769 | glyburide 5 MG [Micronase] |
| RxNorm | 565667 | glyburide 2.5 MG [Libanil] |
| RxNorm | 565668 | glyburide 2.5 MG [Semi-Daonil] |
| RxNorm | 565669 | glyburide 2.5 MG [Euglucon] |
| RxNorm | 565670 | glyburide 2.5 MG [Malix] |
| RxNorm | 565671 | glyburide 2.5 MG [Diabetamide] |
| RxNorm | 565672 | glyburide 5 MG [Libanil] |
| RxNorm | 565673 | glyburide 5 MG [Euglucon] |
| RxNorm | 565674 | glyburide 5 MG [Malix] |
| RxNorm | 565675 | glyburide 5 MG [Diabetamide] |
| RxNorm | 574612 | glyburide 3 MG [Glycron] |
| RxNorm | 606253 | glimepiride / rosiglitazone |
| RxNorm | 602543 | glimepiride / rosiglitazone Oral Tablet |
| RxNorm | 602544 | glimepiride 1 MG / rosiglitazone 4 MG Oral Tablet |
| RxNorm | 602549 | glimepiride 2 MG / rosiglitazone 4 MG Oral Tablet |
| RxNorm | 602550 | glimepiride 4 MG / rosiglitazone 4 MG Oral Tablet |
| RxNorm | 607816 | Avandaryl |
| RxNorm | 574089 | glyburide 1.5 MG [Glycron] |
| RxNorm | 574090 | glyburide 6 MG [Glycron] |
| RxNorm | 574571 | glyburide 4.5 MG [Glycron] |
| RxNorm | 669980 | Glyburase |
| RxNorm | 669981 | glyburide 1.25 MG [Glyburase] |
| RxNorm | 669982 | glyburide Oral Tablet [Glyburase] |

Continued on next page

**Table18 – continued from previous page**

| CodeSystem | ConceptCode | ConceptName |
|---|---|---|
| RxNorm | 669983 | glyburide 1.25 MG Oral Tablet [Glyburase] |
| RxNorm | 669984 | glyburide 2.5 MG [Glyburase] |
| RxNorm | 669985 | glyburide 2.5 MG Oral Tablet [Glyburase] |
| RxNorm | 669986 | glyburide 5 MG [Glyburase] |
| RxNorm | 669987 | glyburide 5 MG Oral Tablet [Glyburase] |
| RxNorm | 647208 | Duetact |
| RxNorm | 647235 | glimepiride / pioglitazone |
| RxNorm | 647236 | glimepiride / pioglitazone Oral Tablet |
| RxNorm | 647237 | glimepiride 2 MG / pioglitazone 30 MG Oral Tablet |
| RxNorm | 647239 | glimepiride 4 MG / pioglitazone 30 MG Oral Tablet |
| RxNorm | 700835 | glipizide Extended Release Oral Tablet |
| RxNorm | 706895 | glimepiride 2 MG / rosiglitazone 8 MG Oral Tablet |
| RxNorm | 706896 | glimepiride 4 MG / rosiglitazone 8 MG Oral Tablet |
| RxNorm | 731455 | glimepiride 4 MG / pioglitazone 30 MG [Duetact] |
| RxNorm | 731457 | glimepiride 4 MG / pioglitazone 30 MG Oral Tablet [Duetact] |
| RxNorm | 731461 | glimepiride 2 MG / pioglitazone 30 MG [Duetact] |
| RxNorm | 731462 | glimepiride / pioglitazone Oral Tablet [Duetact] |
| RxNorm | 731463 | glimepiride 2 MG / pioglitazone 30 MG Oral Tablet [Duetact] |

##### 0.5.4 SGLT2 Inhibitors

Table 19: Concept codes used to identify SGLT2i medications.

| CodeSystem | ConceptCode | ConceptName |
|---|---|---|
| RxNorm | 1488564 | dapagliflozin |
| RxNorm | 1545653 | empagliflozin |
| RxNorm | 1546031 | canagliflozin anhydrous |
| RxNorm | 1373458 | canagliflozin |
| RxNorm | 1373459 | canagliflozin 100 MG |
| RxNorm | 1373460 | canagliflozin Oral Product |
| RxNorm | 1373461 | canagliflozin Pill |
| RxNorm | 1373462 | canagliflozin Oral Tablet |
| RxNorm | 1373463 | canagliflozin 100 MG Oral Tablet |
| RxNorm | 1373464 | Invokana |
| RxNorm | 1373465 | canagliflozin 100 MG [Invokana] |
| RxNorm | 1373466 | canagliflozin Oral Tablet [Invokana] |
| RxNorm | 1373467 | Invokana Oral Product |
| RxNorm | 1373468 | Invokana Pill |
| RxNorm | 1373469 | canagliflozin 100 MG Oral Tablet [Invokana] |

Continued on next page

**Table19 – continued from previous page**

| CodeSystem | ConceptCode | ConceptName |
|---|---|---|
| RxNorm | 1373470 | canagliflozin 300 MG |
| RxNorm | 1373471 | canagliflozin 300 MG Oral Tablet |
| RxNorm | 1373472 | canagliflozin 300 MG [Invokana] |
| RxNorm | 1373473 | canagliflozin 300 MG Oral Tablet [Invokana] |
| RxNorm | 1486436 | dapagliflozin / metformin |
| RxNorm | 1486972 | Farxiga |
| RxNorm | 1486975 | Farxiga Oral Product |
| RxNorm | 1486976 | Farxiga Pill |
| RxNorm | 1486977 | dapagliflozin 10 MG Oral Tablet [Farxiga] |
| RxNorm | 1486981 | dapagliflozin 5 MG Oral Tablet [Farxiga] |
| RxNorm | 1488565 | dapagliflozin 10 MG |
| RxNorm | 1488566 | dapagliflozin Oral Product |
| RxNorm | 1488567 | dapagliflozin Pill |
| RxNorm | 1488568 | dapagliflozin Oral Tablet |
| RxNorm | 1488569 | dapagliflozin 10 MG Oral Tablet |
| RxNorm | 1488573 | dapagliflozin 5 MG |
| RxNorm | 1488574 | dapagliflozin 5 MG Oral Tablet |
| RxNorm | 1534343 | dapagliflozin Oral Tablet [Farxiga] |
| RxNorm | 1534344 | dapagliflozin 5 MG [Farxiga] |
| RxNorm | 1534397 | dapagliflozin 10 MG [Farxiga] |
| RxNorm | 1545145 | canagliflozin 150 MG |
| RxNorm | 1545146 | canagliflozin / metformin Oral Product |
| RxNorm | 1545147 | canagliflozin / metformin Pill |
| RxNorm | 1545148 | canagliflozin / metformin Oral Tablet |
| RxNorm | 1545149 | canagliflozin / metformin |
| RxNorm | 1545150 | canagliflozin 150 MG / metformin hydrochloride 1000 MG Oral Tablet |
| RxNorm | 1545151 | Invokamet |
| RxNorm | 1545152 | canagliflozin 150 MG / metformin hydrochloride 1000 MG [Invokamet] |
| RxNorm | 1545153 | canagliflozin / metformin Oral Tablet [Invokamet] |
| RxNorm | 1545154 | Invokamet Oral Product |
| RxNorm | 1545155 | Invokamet Pill |
| RxNorm | 1545156 | canagliflozin 150 MG / metformin hydrochloride 1000 MG Oral Tablet [Invokamet] |
| RxNorm | 1545157 | canagliflozin 150 MG / metformin hydrochloride 500 MG Oral Tablet |
| RxNorm | 1545158 | canagliflozin 150 MG / metformin hydrochloride 500 MG [Invokamet] |
| RxNorm | 1545159 | canagliflozin 150 MG / metformin hydrochloride 500 MG Oral Tablet [Invokamet] |
| RxNorm | 1545160 | canagliflozin 50 MG |

Continued on next page

**Table19 – continued from previous page**

| CodeSystem | ConceptCode | ConceptName |
|---|---|---|
| RxNorm | 1545161 | canagliflozin 50 MG / metformin hydrochloride 1000 MG Oral Tablet |
| RxNorm | 1545162 | canagliflozin 50 MG / metformin hydrochloride 1000 MG [Invokamet] |
| RxNorm | 1545163 | canagliflozin 50 MG / metformin hydrochloride 1000 MG Oral Tablet [Invokamet] |
| RxNorm | 1545164 | canagliflozin 50 MG / metformin hydrochloride 500 MG Oral Tablet |
| RxNorm | 1545165 | canagliflozin 50 MG / metformin hydrochloride 500 MG [Invokamet] |
| RxNorm | 1545166 | canagliflozin 50 MG / metformin hydrochloride 500 MG Oral Tablet [Invokamet] |
| RxNorm | 1545654 | empagliflozin 10 MG |
| RxNorm | 1545655 | empagliflozin Oral Product |
| RxNorm | 1545656 | empagliflozin Pill |
| RxNorm | 1545657 | empagliflozin Oral Tablet |
| RxNorm | 1545658 | empagliflozin 10 MG Oral Tablet |
| RxNorm | 1545659 | Jardiance |
| RxNorm | 1545660 | empagliflozin 10 MG [Jardiance] |
| RxNorm | 1545661 | empagliflozin Oral Tablet [Jardiance] |
| RxNorm | 1545662 | Jardiance Oral Product |
| RxNorm | 1545663 | Jardiance Pill |
| RxNorm | 1545664 | empagliflozin 10 MG Oral Tablet [Jardiance] |
| RxNorm | 1545665 | empagliflozin 25 MG |
| RxNorm | 1545666 | empagliflozin 25 MG Oral Tablet |
| RxNorm | 1545667 | empagliflozin 25 MG [Jardiance] |
| RxNorm | 1545668 | empagliflozin 25 MG Oral Tablet [Jardiance] |
| RxNorm | 1593057 | dapagliflozin / metformin Extended Release Oral Tablet |
| RxNorm | 1593058 | 24 HR dapagliflozin 10 MG / metformin hydrochloride 1000 MG Extended Release Oral Tablet |
| RxNorm | 1593059 | dapagliflozin 10 MG / metformin hydrochloride 1000 MG Extended Release Oral Tablet |
| RxNorm | 1593068 | 24 HR dapagliflozin 10 MG / metformin hydrochloride 500 MG Extended Release Oral Tablet |
| RxNorm | 1593069 | dapagliflozin 10 MG / metformin hydrochloride 500 MG Extended Release Oral Tablet |
| RxNorm | 1593070 | 24 HR dapagliflozin 5 MG / metformin hydrochloride 1000 MG Extended Release Oral Tablet |
| RxNorm | 1593071 | dapagliflozin 5 MG / metformin hydrochloride 1000 MG Extended Release Oral Tablet |
| RxNorm | 1593072 | 24 HR dapagliflozin 5 MG / metformin hydrochloride 500 MG Extended Release Oral Tablet |

Continued on next page

**Table19 – continued from previous page**

| CodeSystem | ConceptCode | ConceptName |
|---|---|---|
| RxNorm | 1593073 | dapagliflozin 5 MG / metformin hydrochloride 500 MG Extended Release Oral Tablet |
| RxNorm | 1593774 | dapagliflozin / metformin Extended Release Oral Tablet [Xigduo] |
| RxNorm | 1593775 | 24 HR dapagliflozin 10 MG / metformin hydrochloride 1000 MG Extended Release Oral Tablet [Xigduo] |
| RxNorm | 1593776 | dapagliflozin 10 MG / metformin hydrochloride 1000 MG Extended Release Oral Tablet [Xigduo] |
| RxNorm | 1593826 | dapagliflozin 10 MG / metformin hydrochloride 500 MG [Xigduo] |
| RxNorm | 1593827 | dapagliflozin 10 MG / metformin hydrochloride 500 MG Extended Release Oral Tablet [Xigduo] |
| RxNorm | 1593828 | dapagliflozin 5 MG / metformin hydrochloride 1000 MG [Xigduo] |
| RxNorm | 1593829 | dapagliflozin 5 MG / metformin hydrochloride 1000 MG Extended Release Oral Tablet [Xigduo] |
| RxNorm | 1593830 | dapagliflozin 5 MG / metformin hydrochloride 500 MG [Xigduo] |
| RxNorm | 1593831 | 24 HR dapagliflozin 5 MG / metformin hydrochloride 500 MG Extended Release Oral Tablet [Xigduo] |
| RxNorm | 1593832 | dapagliflozin 5 MG / metformin hydrochloride 500 MG Extended Release Oral Tablet [Xigduo] |
| RxNorm | 1593833 | 24 HR dapagliflozin 5 MG / metformin hydrochloride 1000 MG Extended Release Oral Tablet [Xigduo] |
| RxNorm | 1593835 | 24 HR dapagliflozin 10 MG / metformin hydrochloride 500 MG Extended Release Oral Tablet [Xigduo] |
| RxNorm | 1598392 | empagliflozin / linagliptin |
| RxNorm | 1602106 | empagliflozin / linagliptin Oral Product |
| RxNorm | 1602107 | empagliflozin / linagliptin Pill |
| RxNorm | 1602108 | empagliflozin / linagliptin Oral Tablet |
| RxNorm | 1602109 | empagliflozin 10 MG / linagliptin 5 MG Oral Tablet |
| RxNorm | 1602110 | Glyxambi |
| RxNorm | 1602111 | empagliflozin 10 MG / linagliptin 5 MG [Glyxambi] |
| RxNorm | 1602112 | empagliflozin / linagliptin Oral Tablet [Glyxambi] |
| RxNorm | 1602113 | Glyxambi Oral Product |
| RxNorm | 1602114 | Glyxambi Pill |
| RxNorm | 1602115 | empagliflozin 10 MG / linagliptin 5 MG Oral Tablet [Glyxambi] |
| RxNorm | 1602118 | empagliflozin 25 MG / linagliptin 5 MG Oral Tablet |
| RxNorm | 1602119 | empagliflozin 25 MG / linagliptin 5 MG [Glyxambi] |
| RxNorm | 1602120 | empagliflozin 25 MG / linagliptin 5 MG Oral Tablet [Glyxambi] |
| RxNorm | 1592709 | dapagliflozin / metformin Oral Product |

Continued on next page

**Table19 – continued from previous page**

| CodeSystem | ConceptCode | ConceptName |
|---|---|---|
| RxNorm | 1592710 | dapagliflozin / metformin Pill |
| RxNorm | 1592713 | Xigduo |
| RxNorm | 1592716 | Xigduo Oral Product |
| RxNorm | 1592717 | Xigduo Pill |
| RxNorm | 1592722 | dapagliflozin 10 MG / metformin hydrochloride 1000 MG [Xigduo] |
| RxNorm | 1664310 | empagliflozin 5 MG |
| RxNorm | 1664311 | empagliflozin / metformin Oral Product |
| RxNorm | 1664312 | empagliflozin / metformin Pill |
| RxNorm | 1664313 | empagliflozin / metformin Oral Tablet |
| RxNorm | 1664314 | empagliflozin / metformin |
| RxNorm | 1664315 | empagliflozin 5 MG / metformin hydrochloride 500 MG Oral Tablet |
| RxNorm | 1664316 | Synjardy |
| RxNorm | 1664317 | empagliflozin 5 MG / metformin hydrochloride 500 MG [Synjardy] |
| RxNorm | 1664318 | empagliflozin / metformin Oral Tablet [Synjardy] |
| RxNorm | 1664319 | Synjardy Oral Product |
| RxNorm | 1664320 | Synjardy Pill |
| RxNorm | 1664321 | empagliflozin 5 MG / metformin hydrochloride 500 MG Oral Tablet [Synjardy] |
| RxNorm | 1664322 | empagliflozin 12.5 MG |
| RxNorm | 1664323 | empagliflozin 12.5 MG / metformin hydrochloride 500 MG Oral Tablet |
| RxNorm | 1664324 | empagliflozin 12.5 MG / metformin hydrochloride 500 MG [Synjardy] |
| RxNorm | 1664325 | empagliflozin 12.5 MG / metformin hydrochloride 500 MG Oral Tablet [Synjardy] |
| RxNorm | 1664326 | empagliflozin 5 MG / metformin hydrochloride 1000 MG Oral Tablet |
| RxNorm | 1664327 | empagliflozin 5 MG / metformin hydrochloride 1000 MG [Synjardy] |
| RxNorm | 1664328 | empagliflozin 5 MG / metformin hydrochloride 1000 MG Oral Tablet [Synjardy] |
| RxNorm | 1665367 | empagliflozin 12.5 MG / metformin hydrochloride 1000 MG Oral Tablet |
| RxNorm | 1665368 | empagliflozin 12.5 MG / metformin hydrochloride 1000 MG [Synjardy] |
| RxNorm | 1665369 | empagliflozin 12.5 MG / metformin hydrochloride 1000 MG Oral Tablet [Synjardy] |
| RxNorm | 1727500 | dapagliflozin / saxagliptin |
| RxNorm | 1810996 | canagliflozin / metformin Extended Release Oral Tablet |

Continued on next page

**Table19 – continued from previous page**

| CodeSystem | ConceptCode | ConceptName |
|---|---|---|
| RxNorm | 1810997 | 24 HR canagliflozin 150 MG / metformin hydrochloride 1000 MG Extended Release Oral Tablet |
| RxNorm | 1810998 | canagliflozin / metformin Extended Release Oral Tablet [Invokamet] |
| RxNorm | 1810999 | 24 HR canagliflozin 150 MG / metformin hydrochloride 1000 MG Extended Release Oral Tablet [Invokamet] |
| RxNorm | 1811000 | canagliflozin 150 MG / metformin hydrochloride 1000 MG Extended Release Oral Tablet |
| RxNorm | 1811001 | canagliflozin 150 MG / metformin hydrochloride 1000 MG Extended Release Oral Tablet [Invokamet] |
| RxNorm | 1811002 | 24 HR canagliflozin 150 MG / metformin hydrochloride 500 MG Extended Release Oral Tablet |
| RxNorm | 1811003 | 24 HR canagliflozin 150 MG / metformin hydrochloride 500 MG Extended Release Oral Tablet [Invokamet] |
| RxNorm | 1811004 | canagliflozin 150 MG / metformin hydrochloride 500 MG Extended Release Oral Tablet |
| RxNorm | 1811005 | canagliflozin 150 MG / metformin hydrochloride 500 MG Extended Release Oral Tablet [Invokamet] |
| RxNorm | 1811006 | 24 HR canagliflozin 50 MG / metformin hydrochloride 1000 MG Extended Release Oral Tablet |
| RxNorm | 1811007 | 24 HR canagliflozin 50 MG / metformin hydrochloride 1000 MG Extended Release Oral Tablet [Invokamet] |
| RxNorm | 1862684 | empagliflozin / metformin Extended Release Oral Tablet |
| RxNorm | 1862685 | 24 HR empagliflozin 10 MG / metformin hydrochloride 1000 MG Extended Release Oral Tablet |
| RxNorm | 1862686 | empagliflozin 10 MG / metformin hydrochloride 1000 MG [Synjardy] |
| RxNorm | 1862687 | empagliflozin / metformin Extended Release Oral Tablet [Synjardy] |
| RxNorm | 1862688 | 24 HR empagliflozin 10 MG / metformin hydrochloride 1000 MG Extended Release Oral Tablet [Synjardy] |
| RxNorm | 1862689 | empagliflozin 10 MG / metformin hydrochloride 1000 MG Extended Release Oral Tablet |
| RxNorm | 1862690 | empagliflozin 10 MG / metformin hydrochloride 1000 MG Extended Release Oral Tablet [Synjardy] |
| RxNorm | 1862691 | 24 HR empagliflozin 12.5 MG / metformin hydrochloride 1000 MG Extended Release Oral Tablet |
| RxNorm | 1862692 | 24 HR empagliflozin 12.5 MG / metformin hydrochloride 1000 MG Extended Release Oral Tablet [Synjardy] |
| RxNorm | 1862693 | empagliflozin 12.5 MG / metformin hydrochloride 1000 MG Extended Release Oral Tablet |
| RxNorm | 1811008 | canagliflozin 50 MG / metformin hydrochloride 1000 MG Extended Release Oral Tablet |

Continued on next page

**Table19 – continued from previous page**

| CodeSystem | ConceptCode | ConceptName |
|---|---|---|
| RxNorm | 1811009 | canagliflozin 50 MG / metformin hydrochloride 1000 MG Extended Release Oral Tablet [Invokamet] |
| RxNorm | 1811010 | 24 HR canagliflozin 50 MG / metformin hydrochloride 500 MG Extended Release Oral Tablet |
| RxNorm | 1811011 | 24 HR canagliflozin 50 MG / metformin hydrochloride 500 MG Extended Release Oral Tablet [Invokamet] |
| RxNorm | 1811012 | canagliflozin 50 MG / metformin hydrochloride 500 MG Extended Release Oral Tablet |
| RxNorm | 1811013 | canagliflozin 50 MG / metformin hydrochloride 500 MG Extended Release Oral Tablet [Invokamet] |
| RxNorm | 1862694 | empagliflozin 12.5 MG / metformin hydrochloride 1000 MG Extended Release Oral Tablet [Synjardy] |
| RxNorm | 1862695 | 24 HR empagliflozin 25 MG / metformin hydrochloride 1000 MG Extended Release Oral Tablet |
| RxNorm | 1862696 | empagliflozin 25 MG / metformin hydrochloride 1000 MG [Synjardy] |
| RxNorm | 1862697 | 24 HR empagliflozin 25 MG / metformin hydrochloride 1000 MG Extended Release Oral Tablet [Synjardy] |
| RxNorm | 1862698 | empagliflozin 25 MG / metformin hydrochloride 1000 MG Extended Release Oral Tablet |
| RxNorm | 1862699 | empagliflozin 25 MG / metformin hydrochloride 1000 MG Extended Release Oral Tablet [Synjardy] |
| RxNorm | 1862700 | 24 HR empagliflozin 5 MG / metformin hydrochloride 1000 MG Extended Release Oral Tablet |
| RxNorm | 1862701 | 24 HR empagliflozin 5 MG / metformin hydrochloride 1000 MG Extended Release Oral Tablet [Synjardy] |
| RxNorm | 1862702 | empagliflozin 5 MG / metformin hydrochloride 1000 MG Extended Release Oral Tablet |
| RxNorm | 1862703 | empagliflozin 5 MG / metformin hydrochloride 1000 MG Extended Release Oral Tablet [Synjardy] |
| RxNorm | 1925495 | dapagliflozin / saxagliptin Oral Product |
| RxNorm | 1925496 | dapagliflozin / saxagliptin Pill |
| RxNorm | 1925497 | dapagliflozin / saxagliptin Oral Tablet |
| RxNorm | 1925498 | dapagliflozin 10 MG / saxagliptin 5 MG Oral Tablet |
| RxNorm | 1925499 | Qtern |
| RxNorm | 1925500 | dapagliflozin 10 MG / saxagliptin 5 MG [Qtern] |
| RxNorm | 1925501 | dapagliflozin / saxagliptin Oral Tablet [Qtern] |
| RxNorm | 1925502 | Qtern Oral Product |
| RxNorm | 1925503 | Qtern Pill |
| RxNorm | 1925504 | dapagliflozin 10 MG / saxagliptin 5 MG Oral Tablet [Qtern] |
| RxNorm | 1940495 | dapagliflozin 2.5 MG |
| RxNorm | 1940496 | 24 HR dapagliflozin 2.5 MG / metformin hydrochloride 1000 MG Extended Release Oral Tablet |

Continued on next page

**Table19 – continued from previous page**

| CodeSystem | ConceptCode | ConceptName |
|---|---|---|
| RxNorm | 1940497 | dapagliflozin 2.5 MG / metformin hydrochloride 1000 MG [Xigduo] |
| RxNorm | 1940498 | 24 HR dapagliflozin 2.5 MG / metformin hydrochloride 1000 MG Extended Release Oral Tablet [Xigduo] |
| RxNorm | 1940499 | dapagliflozin 2.5 MG / metformin hydrochloride 1000 MG Extended Release Oral Tablet |
| RxNorm | 1940500 | dapagliflozin 2.5 MG / metformin hydrochloride 1000 MG Extended Release Oral Tablet [Xigduo] |
| RxNorm | 2117292 | dapagliflozin / metformin / saxagliptin |
| RxNorm | 2169274 | dapagliflozin 5 MG / saxagliptin 5 MG Oral Tablet |
| RxNorm | 2169275 | dapagliflozin 5 MG / saxagliptin 5 MG [Qtern] |
| RxNorm | 2169276 | dapagliflozin 5 MG / saxagliptin 5 MG Oral Tablet [Qtern] |
| RxNorm | 2281864 | empagliflozin / linagliptin / metformin |
| RxNorm | 2371722 | dapagliflozin / metformin / saxagliptin Oral Product |
| RxNorm | 2371723 | dapagliflozin / metformin / saxagliptin Pill |
| RxNorm | 2371724 | dapagliflozin / metformin / saxagliptin Extended Release Oral Tablet |
| RxNorm | 2371725 | 24 HR dapagliflozin 10 MG / metformin hydrochloride 1000 MG / saxagliptin 5 MG Extended Release Oral Tablet |
| RxNorm | 2371726 | Qternmet |
| RxNorm | 2371727 | dapagliflozin 10 MG / metformin hydrochloride 1000 MG / saxagliptin 5 MG [Qternmet] |
| RxNorm | 2371728 | dapagliflozin / metformin / saxagliptin Extended Release Oral Tablet [Qternmet] |
| RxNorm | 2371729 | Qternmet Oral Product |
| RxNorm | 2371730 | Qternmet Pill |
| RxNorm | 2371731 | 24 HR dapagliflozin 10 MG / metformin hydrochloride 1000 MG / saxagliptin 5 MG Extended Release Oral Tablet [Qternmet] |
| RxNorm | 2371732 | dapagliflozin 10 MG / metformin hydrochloride 1000 MG / saxagliptin 5 MG Extended Release Oral Tablet |
| RxNorm | 2371733 | dapagliflozin 10 MG / metformin hydrochloride 1000 MG / saxagliptin 5 MG Extended Release Oral Tablet [Qternmet] |
| RxNorm | 2371734 | 24 HR dapagliflozin 2.5 MG / metformin hydrochloride 1000 MG / saxagliptin 2.5 MG Extended Release Oral Tablet |
| RxNorm | 2371735 | dapagliflozin 2.5 MG / metformin hydrochloride 1000 MG / saxagliptin 2.5 MG [Qternmet] |
| RxNorm | 2371736 | 24 HR dapagliflozin 2.5 MG / metformin hydrochloride 1000 MG / saxagliptin 2.5 MG Extended Release Oral Tablet [Qternmet] |
| RxNorm | 2371737 | dapagliflozin 2.5 MG / metformin hydrochloride 1000 MG / saxagliptin 2.5 MG Extended Release Oral Tablet |

Continued on next page

**Table19 – continued from previous page**

| CodeSystem | ConceptCode | ConceptName |
|---|---|---|
| RxNorm | 2371738 | dapagliflozin 2.5 MG / metformin hydrochloride 1000 MG / saxagliptin 2.5 MG Extended Release Oral Tablet [Qternmet] |
| RxNorm | 2371740 | 24 HR dapagliflozin 5 MG / metformin hydrochloride 1000 MG / saxagliptin 2.5 MG Extended Release Oral Tablet |
| RxNorm | 2371741 | dapagliflozin 5 MG / metformin hydrochloride 1000 MG / saxagliptin 2.5 MG [Qternmet] |
| RxNorm | 2371742 | 24 HR dapagliflozin 5 MG / metformin hydrochloride 1000 MG / saxagliptin 2.5 MG Extended Release Oral Tablet [Qternmet] |
| RxNorm | 2371743 | dapagliflozin 5 MG / metformin hydrochloride 1000 MG / saxagliptin 2.5 MG Extended Release Oral Tablet |
| RxNorm | 2371744 | dapagliflozin 5 MG / metformin hydrochloride 1000 MG / saxagliptin 2.5 MG Extended Release Oral Tablet [Qternmet] |
| RxNorm | 2371745 | 24 HR dapagliflozin 5 MG / metformin hydrochloride 1000 MG / saxagliptin 5 MG Extended Release Oral Tablet |
| RxNorm | 2371746 | dapagliflozin 5 MG / metformin hydrochloride 1000 MG / saxagliptin 5 MG [Qternmet] |
| RxNorm | 2371747 | 24 HR dapagliflozin 5 MG / metformin hydrochloride 1000 MG / saxagliptin 5 MG Extended Release Oral Tablet [Qternmet] |
| RxNorm | 2371748 | dapagliflozin 5 MG / metformin hydrochloride 1000 MG / saxagliptin 5 MG Extended Release Oral Tablet |
| RxNorm | 2371749 | dapagliflozin 5 MG / metformin hydrochloride 1000 MG / saxagliptin 5 MG Extended Release Oral Tablet [Qternmet] |
| RxNorm | 2359276 | empagliflozin / linagliptin / metformin Oral Product |
| RxNorm | 2359277 | empagliflozin / linagliptin / metformin Pill |
| RxNorm | 2359278 | empagliflozin / linagliptin / metformin Extended Release Oral Tablet |
| RxNorm | 2359279 | 24 HR empagliflozin 10 MG / linagliptin 5 MG / metformin hydrochloride 1000 MG Extended Release Oral Tablet |
| RxNorm | 2359280 | Trijardy |
| RxNorm | 2359281 | empagliflozin 10 MG / linagliptin 5 MG / metformin hydrochloride 1000 MG [Trijardy] |
| RxNorm | 2359282 | empagliflozin / linagliptin / metformin Extended Release Oral Tablet [Trijardy] |
| RxNorm | 2359283 | Trijardy Oral Product |
| RxNorm | 2359284 | Trijardy Pill |
| RxNorm | 2359285 | 24 HR empagliflozin 10 MG / linagliptin 5 MG / metformin hydrochloride 1000 MG Extended Release Oral Tablet [Trijardy] |

Continued on next page

**Table19 – continued from previous page**

| CodeSystem | ConceptCode | ConceptName |
|---|---|---|
| RxNorm | 2359286 | empagliflozin 10 MG / linagliptin 5 MG / metformin hydrochloride 1000 MG Extended Release Oral Tablet |
| RxNorm | 2359287 | empagliflozin 10 MG / linagliptin 5 MG / metformin hydrochloride 1000 MG Extended Release Oral Tablet [Trijardy] |
| RxNorm | 2359288 | 24 HR empagliflozin 12.5 MG / linagliptin 2.5 MG / metformin hydrochloride 1000 MG Extended Release Oral Tablet |
| RxNorm | 2359289 | empagliflozin 12.5 MG / linagliptin 2.5 MG / metformin hydrochloride 1000 MG [Trijardy] |
| RxNorm | 2359290 | 24 HR empagliflozin 12.5 MG / linagliptin 2.5 MG / metformin hydrochloride 1000 MG Extended Release Oral Tablet [Trijardy] |
| RxNorm | 2359291 | empagliflozin 12.5 MG / linagliptin 2.5 MG / metformin hydrochloride 1000 MG Extended Release Oral Tablet |
| RxNorm | 2359292 | empagliflozin 12.5 MG / linagliptin 2.5 MG / metformin hydrochloride 1000 MG Extended Release Oral Tablet [Trijardy] |
| RxNorm | 2359351 | 24 HR empagliflozin 25 MG / linagliptin 5 MG / metformin hydrochloride 1000 MG Extended Release Oral Tablet |
| RxNorm | 2359352 | empagliflozin 25 MG / linagliptin 5 MG / metformin hydrochloride 1000 MG [Trijardy] |
| RxNorm | 2359353 | 24 HR empagliflozin 25 MG / linagliptin 5 MG / metformin hydrochloride 1000 MG Extended Release Oral Tablet [Trijardy] |
| RxNorm | 2359354 | empagliflozin 25 MG / linagliptin 5 MG / metformin hydrochloride 1000 MG Extended Release Oral Tablet |
| RxNorm | 2359355 | empagliflozin 25 MG / linagliptin 5 MG / metformin hydrochloride 1000 MG Extended Release Oral Tablet [Trijardy] |
| RxNorm | 2359356 | 24 HR empagliflozin 5 MG / linagliptin 2.5 MG / metformin hydrochloride 1000 MG Extended Release Oral Tablet |
| RxNorm | 2359357 | empagliflozin 5 MG / linagliptin 2.5 MG / metformin hydrochloride 1000 MG [Trijardy] |
| RxNorm | 2359358 | 24 HR empagliflozin 5 MG / linagliptin 2.5 MG / metformin hydrochloride 1000 MG Extended Release Oral Tablet [Trijardy] |
| RxNorm | 2359359 | empagliflozin 5 MG / linagliptin 2.5 MG / metformin hydrochloride 1000 MG Extended Release Oral Tablet |
| RxNorm | 2359360 | empagliflozin 5 MG / linagliptin 2.5 MG / metformin hydrochloride 1000 MG Extended Release Oral Tablet [Trijardy] |

##### 0.5.5 DPP-4

Table 20: Concept codes used to identify DPP-4 medications.

| CodeSystem | ConceptCode | ConceptName |
|---|---|---|
| RxNorm | 1100699 | linagliptin |
| RxNorm | 1368001 | alogliptin |
| RxNorm | 1546030 | saxagliptin anhydrous |
| RxNorm | 593411 | sitagliptin |
| RxNorm | 1243830 | Metformin / sitagliptin Extended Release Tablet [Janumet 100/1000] |
| RxNorm | 1243847 | Metformin / sitagliptin Extended Release Tablet [Janumet 50/500] |
| RxNorm | 1243023 | Linagliptin / Metformin Oral Tablet [Jentadueto 2.5/1000] |
| RxNorm | 1243030 | Linagliptin / Metformin Oral Tablet [Jentadueto 2.5/500] |
| RxNorm | 705136 | Metformin 1000 MG / sitagliptin 50 MG Oral Tablet [Janumet] |
| RxNorm | 705134 | Metformin 1000 MG / sitagliptin 50 MG [Janumet] |
| RxNorm | 705135 | Metformin / sitagliptin Oral Tablet [Janumet] |
| RxNorm | 705138 | Metformin 500 MG / sitagliptin 50 MG Oral Tablet [Janumet] |
| RxNorm | 748763 | Metformin 500 MG / sitagliptin 64.3 MG Oral Tablet |
| RxNorm | 757607 | Metformin / sitagliptin Oral Tablet [Janumet 50 mg/500 mg] |
| RxNorm | 748762 | sitagliptin 64.3 MG |
| RxNorm | 748765 | Metformin 500 MG / sitagliptin 64.3 MG Oral Tablet [Janumet] |
| RxNorm | 705137 | Metformin 500 MG / sitagliptin 50 MG [Janumet] |
| RxNorm | 748764 | Metformin 500 MG / sitagliptin 64.3 MG [Janumet] |
| RxNorm | 1043581 | Metformin / saxagliptin Extended Release Tablet [Kombiglyze 5/500] |
| RxNorm | 1043573 | Metformin / saxagliptin Extended Release Tablet [Kombiglyze 5/1000] |
| RxNorm | 1189807 | Simvastatin / sitagliptin Oral Tablet [Juvisync 100/10] |
| RxNorm | 1189824 | Simvastatin / sitagliptin Oral Tablet [Juvisync 100/40] |
| RxNorm | 1243031 | Jentadueto 2.5/500 Oral Product |
| RxNorm | 1243025 | Jentadueto 2.5/1000 Pill |
| RxNorm | 1043561 | metformin / saxagliptin Extended Release Oral Tablet |
| RxNorm | 1043562 | metformin / saxagliptin |
| RxNorm | 1043563 | 24 HR metformin hydrochloride 1000 MG / saxagliptin 2.5 MG Extended Release Oral Tablet |
| RxNorm | 1043565 | metformin hydrochloride 1000 MG / saxagliptin 2.5 MG [Kombiglyze] |
| RxNorm | 1043566 | metformin / saxagliptin Extended Release Oral Tablet [Kombiglyze] |

Continued on next page

**Table20 – continued from previous page**

| CodeSystem | ConceptCode | ConceptName |
|---|---|---|
| RxNorm | 1043567 | 24 HR metformin hydrochloride 1000 MG / saxagliptin 2.5 MG Extended Release Oral Tablet [Kombiglyze] |
| RxNorm | 1043568 | metformin hydrochloride 1000 MG / saxagliptin 2.5 MG Extended Release Oral Tablet |
| RxNorm | 1043569 | metformin hydrochloride 1000 MG / saxagliptin 2.5 MG Extended Release Oral Tablet [Kombiglyze] |
| RxNorm | 1043570 | 24 HR metformin hydrochloride 1000 MG / saxagliptin 5 MG Extended Release Oral Tablet |
| RxNorm | 1043572 | metformin hydrochloride 1000 MG / saxagliptin 5 MG [Kombiglyze] |
| RxNorm | 1043574 | 24 HR metformin hydrochloride 1000 MG / saxagliptin 5 MG Extended Release Oral Tablet [Kombiglyze] |
| RxNorm | 1043575 | metformin hydrochloride 1000 MG / saxagliptin 5 MG Extended Release Oral Tablet |
| RxNorm | 1043576 | metformin hydrochloride 1000 MG / saxagliptin 5 MG Extended Release Oral Tablet [Kombiglyze] |
| RxNorm | 1043578 | 24 HR metformin hydrochloride 500 MG / saxagliptin 5 MG Extended Release Oral Tablet |
| RxNorm | 1043580 | metformin hydrochloride 500 MG / saxagliptin 5 MG [Kombiglyze] |
| RxNorm | 1043582 | 24 HR metformin hydrochloride 500 MG / saxagliptin 5 MG Extended Release Oral Tablet [Kombiglyze] |
| RxNorm | 1043583 | metformin hydrochloride 500 MG / saxagliptin 5 MG Extended Release Oral Tablet |
| RxNorm | 1043584 | metformin hydrochloride 500 MG / saxagliptin 5 MG Extended Release Oral Tablet [Kombiglyze] |
| RxNorm | 1100700 | linagliptin 5 MG |
| RxNorm | 1100701 | linagliptin Oral Tablet |
| RxNorm | 1100702 | linagliptin 5 MG Oral Tablet |
| RxNorm | 1100703 | Tradjenta |
| RxNorm | 1100704 | linagliptin 5 MG [Tradjenta] |
| RxNorm | 1100705 | linagliptin Oral Tablet [Tradjenta] |
| RxNorm | 1100706 | linagliptin 5 MG Oral Tablet [Tradjenta] |
| RxNorm | 1158518 | saxagliptin Oral Product |
| RxNorm | 1158519 | saxagliptin Pill |
| RxNorm | 1159662 | sitagliptin Oral Product |
| RxNorm | 1159663 | sitagliptin Pill |
| RxNorm | 1161605 | metformin / saxagliptin Oral Product |
| RxNorm | 1161606 | metformin / saxagliptin Pill |
| RxNorm | 1161607 | metformin / sitagliptin Oral Product |
| RxNorm | 1161608 | metformin / sitagliptin Pill |
| RxNorm | 1164670 | linagliptin Oral Product |
| RxNorm | 1164671 | linagliptin Pill |

Continued on next page

**Table20 – continued from previous page**

| CodeSystem | ConceptCode | ConceptName |
|---|---|---|
| RxNorm | 1181729 | Onglyza Oral Product |
| RxNorm | 1181730 | Onglyza Pill |
| RxNorm | 1172860 | Kombiglyze Pill |
| RxNorm | 1172861 | Kombiglyze Oral Product |
| RxNorm | 1167810 | Janumet Oral Product |
| RxNorm | 1167811 | Janumet Pill |
| RxNorm | 1167814 | Januvia Oral Product |
| RxNorm | 1167815 | Januvia Pill |
| RxNorm | 1179163 | Tradjenta Oral Product |
| RxNorm | 1179164 | Tradjenta Pill |
| RxNorm | 1189800 | simvastatin / sitagliptin Oral Product |
| RxNorm | 1189801 | simvastatin / sitagliptin Pill |
| RxNorm | 1189802 | simvastatin / sitagliptin Oral Tablet |
| RxNorm | 1189803 | simvastatin / sitagliptin |
| RxNorm | 1189804 | simvastatin 10 MG / sitagliptin 100 MG Oral Tablet |
| RxNorm | 1189806 | simvastatin 10 MG / sitagliptin 100 MG [Juvisync] |
| RxNorm | 1189808 | simvastatin 20 MG / sitagliptin 100 MG Oral Tablet |
| RxNorm | 1189810 | simvastatin 20 MG / sitagliptin 100 MG [Juvisync] |
| RxNorm | 1189811 | simvastatin / sitagliptin Oral Tablet [Juvisync] |
| RxNorm | 1189812 | Juvisync Oral Product |
| RxNorm | 1189813 | Juvisync Pill |
| RxNorm | 1189814 | simvastatin 20 MG / sitagliptin 100 MG Oral Tablet [Juvisync] |
| RxNorm | 1189818 | simvastatin 10 MG / sitagliptin 100 MG Oral Tablet [Juvisync] |
| RxNorm | 1189821 | simvastatin 40 MG / sitagliptin 100 MG Oral Tablet |
| RxNorm | 1189823 | simvastatin 40 MG / sitagliptin 100 MG [Juvisync] |
| RxNorm | 1189827 | simvastatin 40 MG / sitagliptin 100 MG Oral Tablet [Juvisync] |
| RxNorm | 1243015 | linagliptin 2.5 MG |
| RxNorm | 1243016 | linagliptin / metformin Oral Product |
| RxNorm | 1243017 | linagliptin / metformin Pill |
| RxNorm | 1243018 | linagliptin / metformin Oral Tablet |
| RxNorm | 1243019 | linagliptin / metformin |
| RxNorm | 1243020 | linagliptin 2.5 MG / metformin hydrochloride 1000 MG Oral Tablet |
| RxNorm | 1243022 | linagliptin 2.5 MG / metformin hydrochloride 1000 MG [Jentadueto] |
| RxNorm | 1243026 | linagliptin 2.5 MG / metformin hydrochloride 1000 MG Oral Tablet [Jentadueto] |
| RxNorm | 1243027 | linagliptin 2.5 MG / metformin hydrochloride 500 MG Oral Tablet |

Continued on next page

**Table20 – continued from previous page**

| CodeSystem | ConceptCode | ConceptName |
|---|---|---|
| RxNorm | 1243029 | linagliptin 2.5 MG / metformin hydrochloride 500 MG [Jentadueto] |
| RxNorm | 1243033 | linagliptin 2.5 MG / metformin hydrochloride 500 MG Oral Tablet [Jentadueto] |
| RxNorm | 1243034 | linagliptin 2.5 MG / metformin hydrochloride 850 MG Oral Tablet |
| RxNorm | 1243036 | linagliptin 2.5 MG / metformin hydrochloride 850 MG [Jentadueto] |
| RxNorm | 1243037 | linagliptin / metformin Oral Tablet [Jentadueto] |
| RxNorm | 1243038 | Jentadueto Oral Product |
| RxNorm | 1243039 | Jentadueto Pill |
| RxNorm | 1243040 | linagliptin 2.5 MG / metformin hydrochloride 850 MG Oral Tablet [Jentadueto] |
| RxNorm | 1243826 | metformin / sitagliptin Extended Release Oral Tablet |
| RxNorm | 1243827 | 24 HR metformin hydrochloride 1000 MG / sitagliptin 100 MG Extended Release Oral Tablet |
| RxNorm | 1243829 | metformin hydrochloride 1000 MG / sitagliptin 100 MG [Janumet] |
| RxNorm | 1243833 | 24 HR metformin hydrochloride 1000 MG / sitagliptin 100 MG Extended Release Oral Tablet [Janumet] |
| RxNorm | 1243834 | metformin hydrochloride 1000 MG / sitagliptin 100 MG Extended Release Oral Tablet |
| RxNorm | 1243835 | metformin hydrochloride 1000 MG / sitagliptin 100 MG Extended Release Oral Tablet [Janumet] |
| RxNorm | 1243839 | metformin / sitagliptin Extended Release Oral Tablet [Janumet] |
| RxNorm | 1243842 | 24 HR metformin hydrochloride 1000 MG / sitagliptin 50 MG Extended Release Oral Tablet |
| RxNorm | 1243843 | 24 HR metformin hydrochloride 1000 MG / sitagliptin 50 MG Extended Release Oral Tablet [Janumet] |
| RxNorm | 1243844 | metformin hydrochloride 1000 MG / sitagliptin 50 MG Extended Release Oral Tablet |
| RxNorm | 1243845 | metformin hydrochloride 1000 MG / sitagliptin 50 MG Extended Release Oral Tablet [Janumet] |
| RxNorm | 1243846 | 24 HR metformin hydrochloride 500 MG / sitagliptin 50 MG Extended Release Oral Tablet |
| RxNorm | 1243848 | 24 HR metformin hydrochloride 500 MG / sitagliptin 50 MG Extended Release Oral Tablet [Janumet] |
| RxNorm | 1243849 | metformin hydrochloride 500 MG / sitagliptin 50 MG Extended Release Oral Tablet |
| RxNorm | 1243850 | metformin hydrochloride 500 MG / sitagliptin 50 MG Extended Release Oral Tablet [Janumet] |
| RxNorm | 1312409 | simvastatin 10 MG / sitagliptin 50 MG Oral Tablet |

Continued on next page

**Table20 – continued from previous page**

| CodeSystem | ConceptCode | ConceptName |
|---|---|---|
| RxNorm | 1312411 | simvastatin 10 MG / sitagliptin 50 MG [Juvisync] |
| RxNorm | 1312415 | simvastatin 10 MG / sitagliptin 50 MG Oral Tablet [Juvisync] |
| RxNorm | 1312416 | simvastatin 20 MG / sitagliptin 50 MG Oral Tablet |
| RxNorm | 1312418 | simvastatin 20 MG / sitagliptin 50 MG [Juvisync] |
| RxNorm | 1312422 | simvastatin 20 MG / sitagliptin 50 MG Oral Tablet [Juvisync] |
| RxNorm | 1312423 | simvastatin 40 MG / sitagliptin 50 MG Oral Tablet |
| RxNorm | 1312425 | simvastatin 40 MG / sitagliptin 50 MG [Juvisync] |
| RxNorm | 1312429 | simvastatin 40 MG / sitagliptin 50 MG Oral Tablet [Juvisync] |
| RxNorm | 1368002 | alogliptin 25 MG |
| RxNorm | 1368003 | alogliptin Oral Product |
| RxNorm | 1368004 | alogliptin Pill |
| RxNorm | 1368005 | alogliptin Oral Tablet |
| RxNorm | 1368006 | alogliptin 25 MG Oral Tablet |
| RxNorm | 1368007 | Nesina |
| RxNorm | 1368008 | alogliptin 25 MG [Nesina] |
| RxNorm | 1368009 | alogliptin Oral Tablet [Nesina] |
| RxNorm | 1368010 | Nesina Oral Product |
| RxNorm | 1368011 | Nesina Pill |
| RxNorm | 1368012 | alogliptin 25 MG Oral Tablet [Nesina] |
| RxNorm | 1368017 | alogliptin 6.25 MG |
| RxNorm | 1368018 | alogliptin 6.25 MG Oral Tablet |
| RxNorm | 1368019 | alogliptin 6.25 MG [Nesina] |
| RxNorm | 1368020 | alogliptin 6.25 MG Oral Tablet [Nesina] |
| RxNorm | 1368033 | alogliptin 12.5 MG |
| RxNorm | 1368034 | alogliptin 12.5 MG Oral Tablet |
| RxNorm | 1368035 | alogliptin 12.5 MG [Nesina] |
| RxNorm | 1368036 | alogliptin 12.5 MG Oral Tablet [Nesina] |
| RxNorm | 1368381 | alogliptin / metformin Oral Product |
| RxNorm | 1368382 | alogliptin / metformin Pill |
| RxNorm | 1368383 | alogliptin / metformin Oral Tablet |
| RxNorm | 1368384 | alogliptin / metformin |
| RxNorm | 1368385 | alogliptin 12.5 MG / metformin hydrochloride 1000 MG Oral Tablet |
| RxNorm | 1368387 | alogliptin 12.5 MG / metformin hydrochloride 1000 MG [Kazano] |
| RxNorm | 1368391 | alogliptin 12.5 MG / metformin hydrochloride 1000 MG Oral Tablet [Kazano] |
| RxNorm | 1368392 | alogliptin 12.5 MG / metformin hydrochloride 500 MG Oral Tablet |

Continued on next page

**Table20 – continued from previous page**

| CodeSystem | ConceptCode | ConceptName |
|---|---|---|
| RxNorm | 1368394 | alogliptin 12.5 MG / metformin hydrochloride 500 MG [Kazano] |
| RxNorm | 1368395 | alogliptin / metformin Oral Tablet [Kazano] |
| RxNorm | 1368396 | Kazano Oral Product |
| RxNorm | 1368397 | Kazano Pill |
| RxNorm | 1368398 | alogliptin 12.5 MG / metformin hydrochloride 500 MG Oral Tablet [Kazano] |
| RxNorm | 1368399 | alogliptin / pioglitazone Oral Product |
| RxNorm | 1368400 | alogliptin / pioglitazone Pill |
| RxNorm | 1368401 | alogliptin / pioglitazone Oral Tablet |
| RxNorm | 1368402 | alogliptin / pioglitazone |
| RxNorm | 1368403 | alogliptin 12.5 MG / pioglitazone 15 MG Oral Tablet |
| RxNorm | 1368405 | alogliptin 12.5 MG / pioglitazone 15 MG [Oseni] |
| RxNorm | 1368409 | alogliptin 12.5 MG / pioglitazone 15 MG Oral Tablet [Oseni] |
| RxNorm | 1368410 | alogliptin 12.5 MG / pioglitazone 30 MG Oral Tablet |
| RxNorm | 1368412 | alogliptin 12.5 MG / pioglitazone 30 MG [Oseni] |
| RxNorm | 1368416 | alogliptin 12.5 MG / pioglitazone 30 MG Oral Tablet [Oseni] |
| RxNorm | 1368417 | alogliptin 12.5 MG / pioglitazone 45 MG Oral Tablet |
| RxNorm | 1368419 | alogliptin 12.5 MG / pioglitazone 45 MG [Oseni] |
| RxNorm | 1368423 | alogliptin 12.5 MG / pioglitazone 45 MG Oral Tablet [Oseni] |
| RxNorm | 1368424 | alogliptin 25 MG / pioglitazone 15 MG Oral Tablet |
| RxNorm | 1368426 | alogliptin 25 MG / pioglitazone 15 MG [Oseni] |
| RxNorm | 1368430 | alogliptin 25 MG / pioglitazone 15 MG Oral Tablet [Oseni] |
| RxNorm | 1368431 | alogliptin 25 MG / pioglitazone 30 MG Oral Tablet |
| RxNorm | 1368433 | alogliptin 25 MG / pioglitazone 30 MG [Oseni] |
| RxNorm | 1368434 | alogliptin / pioglitazone Oral Tablet [Oseni] |
| RxNorm | 1368435 | Oseni Oral Product |
| RxNorm | 1368436 | Oseni Pill |
| RxNorm | 1368437 | alogliptin 25 MG / pioglitazone 30 MG Oral Tablet [Oseni] |
| RxNorm | 1368438 | alogliptin 25 MG / pioglitazone 45 MG Oral Tablet |
| RxNorm | 1368440 | alogliptin 25 MG / pioglitazone 45 MG [Oseni] |
| RxNorm | 1368444 | alogliptin 25 MG / pioglitazone 45 MG Oral Tablet [Oseni] |
| RxNorm | 1372692 | Kazano |
| RxNorm | 1372706 | Jentadueto |
| RxNorm | 1372717 | Oseni |
| RxNorm | 1372730 | Kombiglyze |
| RxNorm | 1372738 | Janumet |
| RxNorm | 1372754 | Juvisync |
| RxNorm | 1598392 | empagliflozin / linagliptin |
| RxNorm | 1602106 | empagliflozin / linagliptin Oral Product |
| RxNorm | 1602107 | empagliflozin / linagliptin Pill |
| RxNorm | 1602108 | empagliflozin / linagliptin Oral Tablet |
| RxNorm | 1602109 | empagliflozin 10 MG / linagliptin 5 MG Oral Tablet |

Continued on next page

**Table20 – continued from previous page**

| CodeSystem | ConceptCode | ConceptName |
|---|---|---|
| RxNorm | 1602110 | Glyxambi |
| RxNorm | 1602111 | empagliflozin 10 MG / linagliptin 5 MG [Glyxambi] |
| RxNorm | 1602112 | empagliflozin / linagliptin Oral Tablet [Glyxambi] |
| RxNorm | 1602113 | Glyxambi Oral Product |
| RxNorm | 1602114 | Glyxambi Pill |
| RxNorm | 1602115 | empagliflozin 10 MG / linagliptin 5 MG Oral Tablet [Glyxambi] |
| RxNorm | 1602118 | empagliflozin 25 MG / linagliptin 5 MG Oral Tablet |
| RxNorm | 1602119 | empagliflozin 25 MG / linagliptin 5 MG [Glyxambi] |
| RxNorm | 1602120 | empagliflozin 25 MG / linagliptin 5 MG Oral Tablet [Glyxambi] |
| RxNorm | 1727500 | dapagliflozin / saxagliptin |
| RxNorm | 1796088 | linagliptin / metformin Extended Release Oral Tablet |
| RxNorm | 1796089 | 24 HR linagliptin 2.5 MG / metformin hydrochloride 1000 MG Extended Release Oral Tablet |
| RxNorm | 1796090 | linagliptin / metformin Extended Release Oral Tablet [Jentadueto] |
| RxNorm | 1796091 | 24 HR linagliptin 2.5 MG / metformin hydrochloride 1000 MG Extended Release Oral Tablet [Jentadueto] |
| RxNorm | 1796092 | linagliptin 2.5 MG / metformin hydrochloride 1000 MG Extended Release Oral Tablet |
| RxNorm | 1796093 | linagliptin 2.5 MG / metformin hydrochloride 1000 MG Extended Release Oral Tablet [Jentadueto] |
| RxNorm | 1796094 | 24 HR linagliptin 5 MG / metformin hydrochloride 1000 MG Extended Release Oral Tablet |
| RxNorm | 1796095 | linagliptin 5 MG / metformin hydrochloride 1000 MG [Jentadueto] |
| RxNorm | 1796096 | 24 HR linagliptin 5 MG / metformin hydrochloride 1000 MG Extended Release Oral Tablet [Jentadueto] |
| RxNorm | 1796097 | linagliptin 5 MG / metformin hydrochloride 1000 MG Extended Release Oral Tablet |
| RxNorm | 1796098 | linagliptin 5 MG / metformin hydrochloride 1000 MG Extended Release Oral Tablet [Jentadueto] |
| RxNorm | 1925495 | dapagliflozin / saxagliptin Oral Product |
| RxNorm | 1925496 | dapagliflozin / saxagliptin Pill |
| RxNorm | 1925497 | dapagliflozin / saxagliptin Oral Tablet |
| RxNorm | 1925498 | dapagliflozin 10 MG / saxagliptin 5 MG Oral Tablet |
| RxNorm | 1925499 | Qtern |
| RxNorm | 1925500 | dapagliflozin 10 MG / saxagliptin 5 MG [Qtern] |
| RxNorm | 1925501 | dapagliflozin / saxagliptin Oral Tablet [Qtern] |
| RxNorm | 1925502 | Qtern Oral Product |
| RxNorm | 1925503 | Qtern Pill |
| RxNorm | 1925504 | dapagliflozin 10 MG / saxagliptin 5 MG Oral Tablet [Qtern] |

Continued on next page

**Table20 – continued from previous page**

| CodeSystem | ConceptCode | ConceptName |
|---|---|---|
| RxNorm | 1992822 | ertugliflozin / sitagliptin Oral Product |
| RxNorm | 1992823 | ertugliflozin / sitagliptin Pill |
| RxNorm | 1992824 | ertugliflozin / sitagliptin Oral Tablet |
| RxNorm | 1992825 | ertugliflozin / sitagliptin |
| RxNorm | 1992826 | ertugliflozin 15 MG / sitagliptin 100 MG Oral Tablet |
| RxNorm | 1992827 | Steglujan |
| RxNorm | 1992828 | ertugliflozin 15 MG / sitagliptin 100 MG [Steglujan] |
| RxNorm | 1992829 | ertugliflozin / sitagliptin Oral Tablet [Steglujan] |
| RxNorm | 1992830 | Steglujan Oral Product |
| RxNorm | 1992831 | Steglujan Pill |
| RxNorm | 1992832 | ertugliflozin 15 MG / sitagliptin 100 MG Oral Tablet [Steglujan] |
| RxNorm | 1992835 | ertugliflozin 5 MG / sitagliptin 100 MG Oral Tablet |
| RxNorm | 1992836 | ertugliflozin 5 MG / sitagliptin 100 MG [Steglujan] |
| RxNorm | 1992837 | ertugliflozin 5 MG / sitagliptin 100 MG Oral Tablet [Steglujan] |
| RxNorm | 2117292 | dapagliflozin / metformin / saxagliptin |
| RxNorm | 857974 | saxagliptin |
| RxNorm | 858034 | saxagliptin 5 MG |
| RxNorm | 858035 | saxagliptin Oral Tablet |
| RxNorm | 858036 | saxagliptin 5 MG Oral Tablet |
| RxNorm | 858037 | Onglyza |
| RxNorm | 858038 | saxagliptin 5 MG [Onglyza] |
| RxNorm | 858039 | saxagliptin Oral Tablet [Onglyza] |
| RxNorm | 858040 | saxagliptin 5 MG Oral Tablet [Onglyza] |
| RxNorm | 858041 | saxagliptin 2.5 MG |
| RxNorm | 858042 | saxagliptin 2.5 MG Oral Tablet |
| RxNorm | 858043 | saxagliptin 2.5 MG [Onglyza] |
| RxNorm | 858044 | saxagliptin 2.5 MG Oral Tablet [Onglyza] |
| RxNorm | 861769 | metformin hydrochloride 1000 MG / sitagliptin 50 MG Oral Tablet |
| RxNorm | 861770 | metformin hydrochloride 1000 MG / sitagliptin 50 MG [Janumet] |
| RxNorm | 861771 | metformin hydrochloride 1000 MG / sitagliptin 50 MG Oral Tablet [Janumet] |
| RxNorm | 861819 | metformin hydrochloride 500 MG / sitagliptin 50 MG Oral Tablet |
| RxNorm | 861820 | metformin hydrochloride 500 MG / sitagliptin 50 MG [Janumet] |
| RxNorm | 861821 | metformin hydrochloride 500 MG / sitagliptin 50 MG Oral Tablet [Janumet] |
| RxNorm | 700518 | Metformin 1000 MG / sitagliptin 50 MG Oral Tablet |
| RxNorm | 700517 | Metformin 500 MG / sitagliptin 50 MG Oral Tablet |

Continued on next page

**Table20 – continued from previous page**

| CodeSystem | ConceptCode | ConceptName |
|---|---|---|
| RxNorm | 757606 | Metformin 500 MG / sitagliptin 50 MG [Janumet 50 mg/500 mg] |
| RxNorm | 757602 | Metformin 1000 MG / sitagliptin 50 MG [Janumet 50 mg/1000 mg] |
| RxNorm | 757608 | Metformin 500 MG / sitagliptin 50 MG Oral Tablet [Janumet 50 mg/500 mg] |
| RxNorm | 757604 | Metformin 1000 MG / sitagliptin 50 MG Oral Tablet [Janumet 50 mg/1000 mg] |
| RxNorm | 1173547 | Kombiglyze 5/500 Oral Product |
| RxNorm | 1189825 | Juvisync 100/40 Oral Product |
| RxNorm | 1172859 | Kombiglyze 2.5/1000 Oral Product |
| RxNorm | 1173546 | Kombiglyze 5/1000 Pill |
| RxNorm | 1243832 | Janumet 100/1000 Pill |
| RxNorm | 1189816 | Juvisync 100/10 Oral Product |
| RxNorm | 1189826 | Juvisync 100/40 Pill |
| RxNorm | 1189817 | Juvisync 100/10 Pill |
| RxNorm | 1167812 | Janumet 50/500 Oral Product |
| RxNorm | 1243032 | Jentadueto 2.5/500 Pill |
| RxNorm | 1173548 | Kombiglyze 5/500 Pill |
| RxNorm | 1167813 | Janumet 50/500 Pill |
| RxNorm | 1243024 | Jentadueto 2.5/1000 Oral Product |
| RxNorm | 1243831 | Janumet 100/1000 Oral Product |
| RxNorm | 2169274 | dapagliflozin 5 MG / saxagliptin 5 MG Oral Tablet |
| RxNorm | 2169275 | dapagliflozin 5 MG / saxagliptin 5 MG [Qtern] |
| RxNorm | 2169276 | dapagliflozin 5 MG / saxagliptin 5 MG Oral Tablet [Qtern] |
| RxNorm | 2281864 | empagliflozin / linagliptin / metformin |
| RxNorm | 2371722 | dapagliflozin / metformin / saxagliptin Oral Product |
| RxNorm | 2371723 | dapagliflozin / metformin / saxagliptin Pill |
| RxNorm | 2371724 | dapagliflozin / metformin / saxagliptin Extended Release Oral Tablet |
| RxNorm | 2371725 | 24 HR dapagliflozin 10 MG / metformin hydrochloride 1000 MG / saxagliptin 5 MG Extended Release Oral Tablet |
| RxNorm | 2371726 | Qternmet |
| RxNorm | 2371727 | dapagliflozin 10 MG / metformin hydrochloride 1000 MG / saxagliptin 5 MG [Qternmet] |
| RxNorm | 2371728 | dapagliflozin / metformin / saxagliptin Extended Release Oral Tablet [Qternmet] |
| RxNorm | 2371729 | Qternmet Oral Product |
| RxNorm | 2371730 | Qternmet Pill |
| RxNorm | 2371731 | 24 HR dapagliflozin 10 MG / metformin hydrochloride 1000 MG / saxagliptin 5 MG Extended Release Oral Tablet [Qternmet] |

Continued on next page

**Table20 – continued from previous page**

| CodeSystem | ConceptCode | ConceptName |
|---|---|---|
| RxNorm | 2371732 | dapagliflozin 10 MG / metformin hydrochloride 1000 MG / saxagliptin 5 MG Extended Release Oral Tablet |
| RxNorm | 2371733 | dapagliflozin 10 MG / metformin hydrochloride 1000 MG / saxagliptin 5 MG Extended Release Oral Tablet [Qternmet] |
| RxNorm | 2371734 | 24 HR dapagliflozin 2.5 MG / metformin hydrochloride 1000 MG / saxagliptin 2.5 MG Extended Release Oral Tablet |
| RxNorm | 2371735 | dapagliflozin 2.5 MG / metformin hydrochloride 1000 MG / saxagliptin 2.5 MG [Qternmet] |
| RxNorm | 2371736 | 24 HR dapagliflozin 2.5 MG / metformin hydrochloride 1000 MG / saxagliptin 2.5 MG Extended Release Oral Tablet [Qternmet] |
| RxNorm | 2371737 | dapagliflozin 2.5 MG / metformin hydrochloride 1000 MG / saxagliptin 2.5 MG Extended Release Oral Tablet |
| RxNorm | 2371738 | dapagliflozin 2.5 MG / metformin hydrochloride 1000 MG / saxagliptin 2.5 MG Extended Release Oral Tablet [Qternmet] |
| RxNorm | 2371740 | 24 HR dapagliflozin 5 MG / metformin hydrochloride 1000 MG / saxagliptin 2.5 MG Extended Release Oral Tablet |
| RxNorm | 2371741 | dapagliflozin 5 MG / metformin hydrochloride 1000 MG / saxagliptin 2.5 MG [Qternmet] |
| RxNorm | 2371742 | 24 HR dapagliflozin 5 MG / metformin hydrochloride 1000 MG / saxagliptin 2.5 MG Extended Release Oral Tablet [Qternmet] |
| RxNorm | 2371743 | dapagliflozin 5 MG / metformin hydrochloride 1000 MG / saxagliptin 2.5 MG Extended Release Oral Tablet |
| RxNorm | 2371744 | dapagliflozin 5 MG / metformin hydrochloride 1000 MG / saxagliptin 2.5 MG Extended Release Oral Tablet [Qternmet] |
| RxNorm | 2371745 | 24 HR dapagliflozin 5 MG / metformin hydrochloride 1000 MG / saxagliptin 5 MG Extended Release Oral Tablet |
| RxNorm | 2371746 | dapagliflozin 5 MG / metformin hydrochloride 1000 MG / saxagliptin 5 MG [Qternmet] |
| RxNorm | 2371747 | 24 HR dapagliflozin 5 MG / metformin hydrochloride 1000 MG / saxagliptin 5 MG Extended Release Oral Tablet [Qternmet] |
| RxNorm | 2371748 | dapagliflozin 5 MG / metformin hydrochloride 1000 MG / saxagliptin 5 MG Extended Release Oral Tablet |
| RxNorm | 2371749 | dapagliflozin 5 MG / metformin hydrochloride 1000 MG / saxagliptin 5 MG Extended Release Oral Tablet [Qternmet] |
| RxNorm | 2359276 | empagliflozin / linagliptin / metformin Oral Product |
| RxNorm | 2359277 | empagliflozin / linagliptin / metformin Pill |
| RxNorm | 2359278 | empagliflozin / linagliptin / metformin Extended Release Oral Tablet |

Continued on next page

**Table20 – continued from previous page**

| CodeSystem | ConceptCode | ConceptName |
|---|---|---|
| RxNorm | 2359279 | 24 HR empagliflozin 10 MG / linagliptin 5 MG / metformin hydrochloride 1000 MG Extended Release Oral Tablet |
| RxNorm | 2359280 | Trijardy |
| RxNorm | 2359281 | empagliflozin 10 MG / linagliptin 5 MG / metformin hydrochloride 1000 MG [Trijardy] |
| RxNorm | 2359282 | empagliflozin / linagliptin / metformin Extended Release Oral Tablet [Trijardy] |
| RxNorm | 2359283 | Trijardy Oral Product |
| RxNorm | 2359284 | Trijardy Pill |
| RxNorm | 2359285 | 24 HR empagliflozin 10 MG / linagliptin 5 MG / metformin hydrochloride 1000 MG Extended Release Oral Tablet [Trijardy] |
| RxNorm | 2359286 | empagliflozin 10 MG / linagliptin 5 MG / metformin hydrochloride 1000 MG Extended Release Oral Tablet |
| RxNorm | 2359287 | empagliflozin 10 MG / linagliptin 5 MG / metformin hydrochloride 1000 MG Extended Release Oral Tablet [Trijardy] |
| RxNorm | 2359288 | 24 HR empagliflozin 12.5 MG / linagliptin 2.5 MG / metformin hydrochloride 1000 MG Extended Release Oral Tablet |
| RxNorm | 2359289 | empagliflozin 12.5 MG / linagliptin 2.5 MG / metformin hydrochloride 1000 MG [Trijardy] |
| RxNorm | 2359290 | 24 HR empagliflozin 12.5 MG / linagliptin 2.5 MG / metformin hydrochloride 1000 MG Extended Release Oral Tablet [Trijardy] |
| RxNorm | 2359291 | empagliflozin 12.5 MG / linagliptin 2.5 MG / metformin hydrochloride 1000 MG Extended Release Oral Tablet |
| RxNorm | 2359292 | empagliflozin 12.5 MG / linagliptin 2.5 MG / metformin hydrochloride 1000 MG Extended Release Oral Tablet [Trijardy] |
| RxNorm | 2359351 | 24 HR empagliflozin 25 MG / linagliptin 5 MG / metformin hydrochloride 1000 MG Extended Release Oral Tablet |
| RxNorm | 2359352 | empagliflozin 25 MG / linagliptin 5 MG / metformin hydrochloride 1000 MG [Trijardy] |
| RxNorm | 2359353 | 24 HR empagliflozin 25 MG / linagliptin 5 MG / metformin hydrochloride 1000 MG Extended Release Oral Tablet [Trijardy] |
| RxNorm | 2359354 | empagliflozin 25 MG / linagliptin 5 MG / metformin hydrochloride 1000 MG Extended Release Oral Tablet |
| RxNorm | 2359355 | empagliflozin 25 MG / linagliptin 5 MG / metformin hydrochloride 1000 MG Extended Release Oral Tablet [Trijardy] |

Continued on next page

**Table20 – continued from previous page**

| CodeSystem | ConceptCode | ConceptName |
|---|---|---|
| RxNorm | 2359356 | 24 HR empagliflozin 5 MG / linagliptin 2.5 MG / metformin hydrochloride 1000 MG Extended Release Oral Tablet |
| RxNorm | 2359357 | empagliflozin 5 MG / linagliptin 2.5 MG / metformin hydrochloride 1000 MG [Trijardy] |
| RxNorm | 2359358 | 24 HR empagliflozin 5 MG / linagliptin 2.5 MG / metformin hydrochloride 1000 MG Extended Release Oral Tablet [Trijardy] |
| RxNorm | 2359359 | empagliflozin 5 MG / linagliptin 2.5 MG / metformin hydrochloride 1000 MG Extended Release Oral Tablet |
| RxNorm | 2359360 | empagliflozin 5 MG / linagliptin 2.5 MG / metformin hydrochloride 1000 MG Extended Release Oral Tablet [Trijardy] |
| RxNorm | 665031 | sitagliptin 100 MG |
| RxNorm | 665032 | sitagliptin Oral Tablet |
| RxNorm | 665033 | sitagliptin 100 MG Oral Tablet |
| RxNorm | 665034 | sitagliptin 100 MG [Januvia] |
| RxNorm | 665035 | sitagliptin Oral Tablet [Januvia] |
| RxNorm | 665036 | sitagliptin 100 MG Oral Tablet [Januvia] |
| RxNorm | 665037 | sitagliptin 25 MG |
| RxNorm | 665038 | sitagliptin 25 MG Oral Tablet |
| RxNorm | 665039 | sitagliptin 25 MG [Januvia] |
| RxNorm | 665040 | sitagliptin 25 MG Oral Tablet [Januvia] |
| RxNorm | 665041 | sitagliptin 50 MG |
| RxNorm | 665042 | sitagliptin 50 MG Oral Tablet |
| RxNorm | 665043 | sitagliptin 50 MG [Januvia] |
| RxNorm | 665044 | sitagliptin 50 MG Oral Tablet [Januvia] |
| RxNorm | 638596 | Januvia |
| RxNorm | 700516 | metformin / sitagliptin Oral Tablet |
| RxNorm | 729717 | metformin / sitagliptin |
| RxNorm | 757603 | metformin / sitagliptin Oral Tablet [Janumet] |

#### 0.6 Anti-Obesity Medications (AOMS)

##### 0.6.1 Orlistat

Table 21: Concept codes used to identify orlistat medications.

| CodeSystem | ConceptCode | ConceptName |
|---|---|---|
| RxNorm | 1159582 | orlistat Oral Product |
| RxNorm | 1159583 | orlistat Pill |
| RxNorm | 1168454 | Alli Oral Product |
| RxNorm | 1168455 | Alli Pill |

Continued on next page

**Table21 – continued from previous page**

| CodeSystem | ConceptCode | ConceptName |
|---|---|---|
| RxNorm | 1186415 | Xenical Oral Product |
| RxNorm | 1186416 | Xenical Pill |
| RxNorm | 226917 | orlistat 120 MG Oral Capsule [Xenical] |
| RxNorm | 226918 | Xenical |
| RxNorm | 314153 | orlistat 120 MG Oral Capsule |
| RxNorm | 330389 | orlistat 120 MG |
| RxNorm | 366436 | orlistat Oral Capsule [Xenical] |
| RxNorm | 373151 | orlistat Oral Capsule |
| RxNorm | 37925 | orlistat |
| RxNorm | 574045 | orlistat 120 MG [Xenical] |
| RxNorm | 692875 | orlistat 60 MG |
| RxNorm | 692876 | orlistat 60 MG Oral Capsule |
| RxNorm | 708361 | Alli |
| RxNorm | 723844 | orlistat 60 MG [Alli] |
| RxNorm | 723845 | orlistat Oral Capsule [Alli] |
| RxNorm | 723846 | orlistat 60 MG Oral Capsule [Alli] |
| SNOMED CT | 768105006 | Orlistat-containing product in oral dose form |
| SNOMED CT | 777002008 | Orlistat only product |
| SNOMED CT | 780075002 | Orlistat only product in oral dose form |
| SNOMED CT | 317893003 | Orlistat 120mg capsule |
| SNOMED CT | 426638000 | Orlistat 60mg capsule |
| SNOMED CT | 387007000 | Orlistat |
| SNOMED CT | 116093009 | Orlistat-containing product |

##### 0.6.2 Phentermine Topiramate

Table 22: Concept codes used to identify phentermine topiramate medications.

| CodeSystem | ConceptCode | ConceptName |
|---|---|---|
| RxNorm | 1302868 | Phentermine 11.25 MG / topiramate 69 MG Extended Release Capsule |
| RxNorm | 1302827 | 24 HR phentermine 7.5 MG / topiramate 46 MG Extended Release Oral Capsule |
| RxNorm | 1302839 | 24 HR phentermine 3.75 MG / topiramate 23 MG Extended Release Oral Capsule |
| RxNorm | 1302850 | 24 HR phentermine 15 MG / topiramate 92 MG Extended Release Oral Capsule |
| RxNorm | 1313059 | 24 HR phentermine 11.25 MG / topiramate 69 MG Extended Release Oral Capsule |

Continued on next page

**Table22 – continued from previous page**

| CodeSystem | ConceptCode | ConceptName |
|---|---|---|
| RxNorm | 1302833 | 24 HR phentermine 7.5 MG / topiramate 46 MG Extended Release Oral Capsule [Qsymia] |
| RxNorm | 1302845 | 24 HR phentermine 3.75 MG / topiramate 23 MG Extended Release Oral Capsule [Qsymia] |
| RxNorm | 1302856 | 24 HR phentermine 15 MG / topiramate 92 MG Extended Release Oral Capsule [Qsymia] |
| RxNorm | 1313061 | 24 HR phentermine 11.25 MG / topiramate 69 MG Extended Release Oral Capsule [Qsymia] |

#### 0.7 Adverse Events

##### 0.7.1 Bowel Obstruction

Table 23: Concept codes used to identify bowel obstruction adverse events.

| CodeSystem | ConceptCode | ConceptName |
|---|---|---|
| SNOMED CT | 197044000 | Intestinal obstruction without mention of hernia |
| SNOMED CT | 197067003 | (Intestinal adhesions with obstruction) or (other intestinal obstruction) |
| SNOMED CT | 197071000 | Other intestinal obstruction NOS |
| SNOMED CT | 197072007 | Intestinal obstruction NOS |
| SNOMED CT | 197078006 | Acute intestinal obstruction |
| SNOMED CT | 197079003 | Subacute intestinal obstruction |
| SNOMED CT | 197080000 | Obstruction NOS: [intestinal] or [colonic] or [large bowel] or [small bowel] or [subacute intestinal] |
| SNOMED CT | 197541001 | [X]Other and unspecified intestinal obstruction |
| SNOMED CT | 213232001 | Intestinal obstruction as a complication of care NOS |
| SNOMED CT | 233662009 | Distal intestinal obstruction syndrome |
| SNOMED CT | 235801003 | Subacute intestinal obstruction NOS |
| SNOMED CT | 235802005 | Small bowel obstruction NOS |
| SNOMED CT | 235803000 | Large bowel obstruction NOS |
| SNOMED CT | 235805007 | Extrinsic intestinal obstruction |
| SNOMED CT | 155777003 | (Intestinal obstruction NOS) or (intestinal stricture) |
| SNOMED CT | 16834011000119109 | Partial obstruction of intestine |
| SNOMED CT | 16834071000119101 | Complete obstruction of lumen of small intestine |
| SNOMED CT | 46420000 | Intestinal luminal obstruction |
| SNOMED CT | 81060008 | IO - Intestinal obstruction |
| SNOMED CT | 429196001 | Partial obstruction of small bowel |
| SNOMED CT | 710572000 | Intestinal obstruction co-occurrent and due to decreased peristalsis |
| SNOMED CT | 266456007 | Other intestinal obstruction |

Continued on next page

**Table23 – continued from previous page**

| CodeSystem | ConceptCode | ConceptName |
|---|---|---|
| SNOMED CT | 266523009 | (Intestinal obstruction NOS) or (intestinal stricture) |
| SNOMED CT | 278526005 | Intestinal luminal obstruction |
| SNOMED CT | 281254000 | Large bowel obstruction |
| SNOMED CT | 281255004 | Small bowel obstruction |
| SNOMED CT | 67745001 | Recurrent intestinal obstruction |
| SNOMED CT | 77437008 | Intestinal obstruction due to a procedure |
| SNOMED CT | 91580003 | Mural thickening of intestine causing obstruction |
| SNOMED CT | 92825002 | Closed-loop obstruction of intestinal tract |
| SNOMED CT | 1542009 | Omphalocele with obstruction |
| SNOMED CT | 4397001 | Partial congenital duodenal obstruction |
| SNOMED CT | 4410001 | Retroperitoneal hernia with obstruction |
| SNOMED CT | 4998000 | Acute obstructive appendicitis |
| SNOMED CT | 6441003 | Volvulus of colon |
| SNOMED CT | 9359003 | Obstructed bilateral inguinal hernia with gangrene |
| SNOMED CT | 9707006 | Intestinal volvulus |
| SNOMED CT | 9822005 | Umbilical hernia with gangrene AND obstruction |
| SNOMED CT | 10389003 | Acute gastrojejunal ulcer without hemorrhage AND without perforation but with obstruction |
| SNOMED CT | 10897002 | Chronic gastrojejunal ulcer with perforation AND with obstruction |
| SNOMED CT | 11552008 | Complete congenital duodenal obstruction |
| SNOMED CT | 12355008 | Duodenal ulcer with hemorrhage, with perforation AND with obstruction |
| SNOMED CT | 13466009 | Dynamic ileus |
| SNOMED CT | 13916005 | Bilateral recurrent femoral hernia with gangrene AND obstruction |
| SNOMED CT | 15270002 | Obturation obstruction of intestine |
| SNOMED CT | 18169007 | Duodenal ulcer without hemorrhage AND without perforation but with obstruction |
| SNOMED CT | 18269002 | Congenital duodenal obstruction |
| SNOMED CT | 18367003 | Duodenal ulcer with hemorrhage AND obstruction |
| SNOMED CT | 21759003 | Gastrojejunal ulcer with perforation AND obstruction |
| SNOMED CT | 24001002 | Chronic gastrojejunal ulcer with hemorrhage, with perforation and with obstruction |
| SNOMED CT | 25617003 | Congenital duodenal obstruction due to malrotation of intestine |
| SNOMED CT | 26315009 | Congenital obstruction of small intestine |
| SNOMED CT | 28082003 | Chronic duodenal ulcer without hemorrhage AND without perforation but with obstruction |
| SNOMED CT | 28358004 | Acute obstructive appendicitis with perforation AND peritonitis |
| SNOMED CT | 30074005 | Strangulation obstruction of intestine |
| SNOMED CT | 30300009 | Obstructed incisional hernia with gangrene |

Continued on next page

**Table23 – continued from previous page**

| CodeSystem | ConceptCode | ConceptName |
|---|---|---|
| SNOMED CT | 33652004 | Lumbar hernia with obstruction |
| SNOMED CT | 34021006 | Chronic duodenal ulcer with hemorrhage AND obstruction |
| SNOMED CT | 16840521000119100 | Partial obstruction of intestine following procedure |
| SNOMED CT | 16840581000119101 | Complete obstruction of intestine following procedure |
| SNOMED CT | 196658005 | Acute duodenal ulcer with obstruction |
| SNOMED CT | 196666001 | Chronic duodenal ulcer with obstruction |
| SNOMED CT | 196712004 | Acute gastrojejunal ulcer with obstruction |
| SNOMED CT | 196719008 | Chronic gastrojejunal ulcer with obstruction |
| SNOMED CT | 196807006 | Obstructed inguinal hernia |
| SNOMED CT | 196835005 | Obstructed femoral hernia |
| SNOMED CT | 196859000 | Obstructed umbilical hernia |
| SNOMED CT | 196861009 | Paraumbilical hernia with obstruction |
| SNOMED CT | 196884002 | Incisional hernia with obstruction but no gangrene |
| SNOMED CT | 196885001 | Obstructed epigastric hernia |
| SNOMED CT | 196917000 | Obstructed gluteal hernia |
| SNOMED CT | 196956006 | Obstructed Spigelian hernia |
| SNOMED CT | 197006009 | Superior mesenteric artery syndrome |
| SNOMED CT | 197052002 | Multiple intussusception |
| SNOMED CT | 197058003 | Volvulus of small intestine |
| SNOMED CT | 197059006 | Volvulus of the ileocecum |
| SNOMED CT | 197060001 | Sigmoid volvulus |
| SNOMED CT | 197063004 | Enterolith |
| SNOMED CT | 1171359008 | Intestinal obstruction due to adhesion of small intestine |
| SNOMED CT | 1162728008 | Intestinal obstruction due to stricture of intestine |
| SNOMED CT | 1162567000 | Postoperative paralytic ileus |
| SNOMED CT | 206523001 | Meconium ileus |
| SNOMED CT | 206528005 | Meconium plug |
| SNOMED CT | 206529002 | Congenital fecaliths causing obstruction |
| SNOMED CT | 235806008 | Arteriomesenteric compression of duodenojejunal flexure |
| SNOMED CT | 235810006 | Bolus obstruction of intestine |
| SNOMED CT | 235811005 | Cecal volvulus |
| SNOMED CT | 235820001 | Ileo-sigmoid knotting |
| SNOMED CT | 235821002 | Postoperative intestinal obstruction |
| SNOMED CT | 235822009 | Postoperative mechanical intestinal obstruction |
| SNOMED CT | 235833007 | Postoperative ileus |
| SNOMED CT | 235834001 | Infection-induced ileus |
| SNOMED CT | 235835000 | Neural reflex-induced ileus |
| SNOMED CT | 235836004 | Metabolic ileus |
| SNOMED CT | 235837008 | Drug-induced ileus |
| SNOMED CT | 721652009 | Gallstone ileus of small intestine |
| SNOMED CT | 721653004 | Enterolith of small intestine |
| SNOMED CT | 721678000 | Gallstone ileus of large intestine |
| SNOMED CT | 721679008 | Paralytic ileus of large intestine |

Continued on next page

**Table23 – continued from previous page**

| CodeSystem | ConceptCode | ConceptName |
|---|---|---|
| SNOMED CT | 722540008 | Volvulus of large intestine |
| SNOMED CT | 722764006 | Obstructed parastomal hernia |
| SNOMED CT | 722841004 | Perforation of intestine co-occurrent and due to meconium ileus |
| SNOMED CT | 722844007 | Obstruction of small intestine co-occurrent and due to peritoneal adhesions |
| SNOMED CT | 722856008 | Obstruction of large intestine co-occurrent and due to peritoneal adhesions |
| SNOMED CT | 733145007 | Neonatal obstruction of intestine |
| SNOMED CT | 733149001 | Obstructive ileus of small intestine with impaction |
| SNOMED CT | 733447005 | Intestinal obstruction in newborn due to guanylate cyclase 2C deficiency |
| SNOMED CT | 735722005 | Neonatal intestinal perforation with in utero intraluminal obstruction |
| SNOMED CT | 741061001 | Obstructed recurrent right femoral hernia with gangrene |
| SNOMED CT | 741515006 | Obstructed recurrent left femoral hernia with gangrene |
| SNOMED CT | 766942007 | Complete obstruction of colon |
| SNOMED CT | 766943002 | Complete obstruction of intestine |
| SNOMED CT | 767684006 | Obstructed recurrent left inguinal hernia with gangrene |
| SNOMED CT | 767685007 | Obstructed recurrent right inguinal hernia with gangrene |
| SNOMED CT | 767748008 | Obstructed bilateral inguinal hernia |
| SNOMED CT | 767823004 | Obstructed left inguinal hernia with gangrene |
| SNOMED CT | 767824005 | Obstructed right inguinal hernia with gangrene |
| SNOMED CT | 767995003 | Obstructed right femoral hernia with gangrene |
| SNOMED CT | 767996002 | Obstructed left femoral hernia with gangrene |
| SNOMED CT | 772139001 | Obstructed left femoral hernia |
| SNOMED CT | 772140004 | Obstructed right femoral hernia |
| SNOMED CT | 772141000 | Obstructed recurrent right inguinal hernia |
| SNOMED CT | 772142007 | Obstructed recurrent left inguinal hernia |
| SNOMED CT | 772143002 | Obstructed left inguinal hernia |
| SNOMED CT | 772144008 | Obstructed right inguinal hernia |
| SNOMED CT | 772149003 | Obstructed recurrent bilateral inguinal hernia |
| SNOMED CT | 772781000 | Obstructed recurrent bilateral femoral hernia |
| SNOMED CT | 772782007 | Obstructed recurrent right femoral hernia |
| SNOMED CT | 772783002 | Obstructed recurrent left femoral hernia |
| SNOMED CT | 773269009 | Obstructed perineal hernia |
| SNOMED CT | 782504002 | Intestinal obstruction due to recurrent umbilical hernia |
| SNOMED CT | 782505001 | Obstructed recurrent femoral hernia |
| SNOMED CT | 817966005 | Distal intestinal obstruction syndrome due to cystic fibrosis |
| SNOMED CT | 8631000119105 | Intestinal obstruction due to bilateral inguinal hernia |
| SNOMED CT | 8651000119104 | Intestinal obstruction due to bilateral recurrent femoral hernia |

Continued on next page

**Table23 – continued from previous page**

| CodeSystem | ConceptCode | ConceptName |
|---|---|---|
| SNOMED CT | 8661000119102 | Intestinal obstruction due to bilateral recurrent inguinal hernia |
| SNOMED CT | 95001000119103 | Obstructed recurrent inguinal hernia |
| SNOMED CT | 1083751000119104 | Obstructed bilateral femoral hernia |
| SNOMED CT | 1084511000119108 | Intestinal obstruction due to recurrent irreducible inguinal hernia |
| SNOMED CT | 1084521000119101 | Obstructed recurrent irreducible bilateral inguinal hernia |
| SNOMED CT | 1085161000119102 | Intestinal obstruction due to chronic ulcerative pancolitis |
| SNOMED CT | 1085211000119105 | Intestinal obstruction due to chronic ulcerative proctitis |
| SNOMED CT | 1085261000119108 | Intestinal obstruction due to chronic ulcerative rectosigmoiditis |
| SNOMED CT | 1085421000119104 | Intestinal obstruction due to colonic inflammatory polyps |
| SNOMED CT | 1085781000119107 | Intestinal obstruction due to Crohn's disease of large intestine |
| SNOMED CT | 1085831000119104 | Intestinal obstruction due to Crohn's disease of small and large intestine |
| SNOMED CT | 1085881000119103 | Intestinal obstruction due to Crohn's disease of small intestine |
| SNOMED CT | 1085931000119108 | Intestinal obstruction due to Crohn's disease |
| SNOMED CT | 1087271000119105 | Intestinal obstruction co-occurrent and due to incisional hernia |
| SNOMED CT | 1092871000119107 | Intestinal obstruction due to ulcerative colitis |
| SNOMED CT | 16833871000119109 | Partial obstruction of colon |
| SNOMED CT | 84739008 | Perineal hernia with obstruction |
| SNOMED CT | 440663004 | Encapsulating peritoneal sclerosis associated with peritoneal dialysis |
| SNOMED CT | 447320006 | Food bolus obstruction of intestine |
| SNOMED CT | 458422009 | Malrotation of intestine with midgut volvulus |
| SNOMED CT | 698733009 | Intestinal obstruction due to tuberculosis |
| SNOMED CT | 414398007 | Hernia of anterior abdominal wall with obstruction AND gangrene |
| SNOMED CT | 414476004 | Obstructed ventral incisional hernia with gangrene |
| SNOMED CT | 414923000 | Obstructed hernia of anterior abdominal wall |
| SNOMED CT | 414924006 | Obstructed incisional ventral hernia |
| SNOMED CT | 415253006 | Obstructed recurrent hernia of anterior abdominal wall |
| SNOMED CT | 415256003 | Obstructed recurrent incisional hernia of anterior abdominal wall |
| SNOMED CT | 253786009 | Congenital volvulus |
| SNOMED CT | 271575002 | Inspissated stool syndrome |
| SNOMED CT | 276520008 | Transitory ileus of prematurity |
| SNOMED CT | 276521007 | Perinatal intestinal obstruction |
| SNOMED CT | 37976006 | Gallstone ileus |
| SNOMED CT | 40650009 | Obstruction of colon |

Continued on next page

**Table23 – continued from previous page**

| CodeSystem | ConceptCode | ConceptName |
|---|---|---|
| SNOMED CT | 41585009 | Strangulation of colon |
| SNOMED CT | 41916005 | Paraumbilical hernia with gangrene AND obstruction |
| SNOMED CT | 41986000 | Acute duodenal ulcer with hemorrhage, with perforation AND with obstruction |
| SNOMED CT | 42140004 | Obstructed recurrent bilateral inguinal hernia with gangrene |
| SNOMED CT | 42698006 | Gastrojejunal ulcer with hemorrhage AND obstruction |
| SNOMED CT | 45989004 | Obstructed femoral hernia with gangrene |
| SNOMED CT | 47152002 | Gastrojejunal ulcer without hemorrhage AND without perforation but with obstruction |
| SNOMED CT | 49678000 | Obstructed inguinal hernia with gangrene |
| SNOMED CT | 52232007 | Chronic duodenal ileus |
| SNOMED CT | 54798007 | Gastrojejunal ulcer with hemorrhage, with perforation and with obstruction |
| SNOMED CT | 54842003 | Intestinal obstruction by inspissated milk in newborn |
| SNOMED CT | 55525008 | Paralytic ileus |
| SNOMED CT | 56579005 | Chronic gastrojejunal ulcer without hemorrhage AND without perforation but with obstruction |
| SNOMED CT | 57265009 | Congenital obstruction of large intestine |
| SNOMED CT | 58711008 | Acute gastrojejunal ulcer with hemorrhage, with perforation and with obstruction |
| SNOMED CT | 59658000 | Epigastric hernia with gangrene AND obstruction |
| SNOMED CT | 60551006 | Chronic duodenal ulcer with perforation AND obstruction |
| SNOMED CT | 61456005 | Obturator hernia with obstruction |
| SNOMED CT | 62936002 | Acute duodenal ulcer with perforation AND obstruction |
| SNOMED CT | 67766009 | Intestinal adhesions with obstruction |
| SNOMED CT | 68126001 | Obstructed bilateral femoral hernia with gangrene |
| SNOMED CT | 72219001 | Acute gastrojejunal ulcer with perforation AND obstruction |
| SNOMED CT | 72408002 | Acute gastrojejunal ulcer with hemorrhage and obstruction |
| SNOMED CT | 73225005 | Ischiatic hernia with obstruction |
| SNOMED CT | 75342000 | Acute duodenal ulcer without hemorrhage AND without perforation but with obstruction |
| SNOMED CT | 75419006 | Chronic partial afferent loop obstruction |
| SNOMED CT | 77410006 | Duodenal ulcer with perforation AND obstruction |
| SNOMED CT | 79812002 | Volvulus of duodenum |
| SNOMED CT | 16840431000119101 | Partial obstruction of intestine due to peritoneal adhesions |
| SNOMED CT | 1144423005 | Strangulation of small intestine |
| SNOMED CT | 1148568006 | Volvulus of jejunum |
| SNOMED CT | 1197665000 | Enterolith of large intestine |
| SNOMED CT | 1197677005 | Paralytic ileus of small intestine |
| SNOMED CT | 1197679008 | Paralytic ileus of small intestine and colon |
| SNOMED CT | 1197723003 | Postoperative obstruction of large intestine |
| SNOMED CT | 1197725005 | Postoperative obstruction of small intestine |

Continued on next page

**Table23 – continued from previous page**

| CodeSystem | ConceptCode | ConceptName |
|---|---|---|
| SNOMED CT | 82368005 | Transitory ileus of newborn |
| SNOMED CT | 85836000 | Congenital duodenal obstruction due to annular pancreas |
| SNOMED CT | 86092005 | Cystic fibrosis with meconium ileus |
| SNOMED CT | 86258000 | Chronic duodenal ulcer with hemorrhage, with perforation AND with obstruction |
| SNOMED CT | 87756006 | Acute duodenal ulcer with hemorrhage AND obstruction |
| SNOMED CT | 90257004 | Chronic gastrojejunal ulcer with hemorrhage and obstruction |
| SNOMED CT | 95532008 | Obstruction of duodenum |
| SNOMED CT | 95625008 | Paralytic ileus of the newborn |
| SNOMED CT | 95626009 | Spastic ileus of the newborn |
| SNOMED CT | 129601002 | Congenital neurogenic ileus syndrome |
| ICD10CM | K56 | Paralytic ileus and intestinal obstruction without hernia |
| ICD10CM | K56.6 | Other and unspecified intestinal obstruction |
| ICD10CM | K56.60 | Unspecified intestinal obstruction |
| ICD10CM | K56.600 | Partial intestinal obstruction, unspecified as to cause |
| ICD10CM | K56.601 | Complete intestinal obstruction, unspecified as to cause |
| ICD10CM | K56.609 | Unspecified intestinal obstruction, unspecified as to partial versus complete obstruction |
| ICD10CM | K56.69 | Other intestinal obstruction |
| ICD10CM | K56.690 | Other partial intestinal obstruction |
| ICD10CM | K56.691 | Other complete intestinal obstruction |
| ICD10CM | K56.699 | Other intestinal obstruction unspecified as to partial versus complete obstruction |
| ICD10CM | K56.0 | Paralytic ileus |
| ICD10CM | K56.1 | Intussusception |
| ICD10CM | K56.2 | Volvulus |
| ICD10CM | K56.3 | Gallstone ileus |
| ICD10CM | K56.4 | Other impaction of intestine |
| ICD10CM | K56.41 | Fecal impaction |
| ICD10CM | K56.49 | Other impaction of intestine |
| ICD10CM | K56.5 | Intestinal adhesions [bands] with obstruction (postinfection) |
| ICD10CM | K56.50 | Intestinal adhesions [bands], unspecified as to partial versus complete obstruction |
| ICD10CM | K56.51 | Intestinal adhesions [bands], with partial obstruction |
| ICD10CM | K56.52 | Intestinal adhesions [bands] with complete obstruction |
| ICD10CM | K56.7 | Ileus, unspecified |
| ICD9CM | 560.8 | Other specified intestinal obstruction |
| ICD9CM | 560.89 | Other specified intestinal obstruction |
| ICD9CM | 560.9 | Unspecified intestinal obstruction |
| ICD9CM | 560.81 | Intestinal or peritoneal adhesions with obstruction (postoperative) (postinfection) |

Continued on next page

**Table23 – continued from previous page**

| CodeSystem | ConceptCode | ConceptName |
|---|---|---|

##### 0.7.2 Cholecystitis

Table 24: Concept codes used to identify cholecystitis adverse events.

| CodeSystem | ConceptCode | ConceptName |
|---|---|---|
| SNOMED CT | 12932003 | Calculus of common bile duct with chronic cholecystitis with obstruction |
| SNOMED CT | 15082003 | Calculus of bile duct with chronic cholecystitis with obstruction |
| SNOMED CT | 19335008 | Calculus of cystic duct with acute cholecystitis |
| SNOMED CT | 19968009 | Cholecystitis without calculus |
| SNOMED CT | 20824003 | Chronic cholecystitis |
| SNOMED CT | 25924004 | Calculus of gallbladder with cholecystitis |
| SNOMED CT | 29484002 | Cholelithiasis AND cholecystitis without obstruction |
| SNOMED CT | 32067005 | Acute emphysematous cholecystitis |
| SNOMED CT | 34346002 | Acute cholecystitis without calculus |
| SNOMED CT | 197377009 | Gallbladder calculus with acute cholecystitis and no obstruction |
| SNOMED CT | 197378004 | Gallbladder calculus with acute cholecystitis and obstruction |
| SNOMED CT | 197389006 | Bile duct calculus with acute cholecystitis |
| SNOMED CT | 197391003 | Bile duct calculus with acute cholecystitis and obstruction |
| SNOMED CT | 722869007 | Calculus of gallbladder without cholecystitis or cholangitis |
| SNOMED CT | 38871000119104 | Acute and chronic cholecystitis co-occurrent and due to calculus of gallbladder and bile duct |
| SNOMED CT | 40331000119107 | Acute cholecystitis due to biliary calculus |
| SNOMED CT | 435121000124109 | Calculus of gallbladder and bile duct with cholecystitis |
| SNOMED CT | 435131000124107 | Calculus of gallbladder and bile duct with acute cholecystitis |
| SNOMED CT | 435161000124103 | Calculus of gallbladder and bile duct with acute and chronic cholecystitis |
| SNOMED CT | 1084931000119109 | Chronic cholecystitis due to cholelithiasis co-occurrent with choledocholithiasis |
| SNOMED CT | 1084971000119107 | Chronic cholecystitis due to gallbladder calculus with obstruction |
| SNOMED CT | 50450007 | Gallstone AND cholecystitis with obstruction |
| SNOMED CT | 448286002 | Xanthogranulomatous cholecystitis |
| SNOMED CT | 699050007 | Calculus of gallbladder with acute and chronic cholecystitis |
| SNOMED CT | 314949006 | Perforated calculous chronic cholecystitis |

Continued on next page

**Table24 – continued from previous page**

| CodeSystem | ConceptCode | ConceptName |
|---|---|---|
| SNOMED CT | 396335001 | Acute and chronic cholecystitis |
| SNOMED CT | 36483003 | Calculus of bile duct with cholecystitis |
| SNOMED CT | 46341007 | Cholecystitis glandularis proliferans |
| SNOMED CT | 48413001 | Acute hemorrhagic cholecystitis |
| SNOMED CT | 49341008 | Calculus of cystic duct with cholecystitis |
| SNOMED CT | 53928001 | Chronic cholecystitis without calculus |
| SNOMED CT | 59771005 | Calculus of gallbladder with acute cholecystitis |
| SNOMED CT | 60127009 | Calculus of bile duct with acute cholecystitis without obstruction |
| SNOMED CT | 64534006 | Hyperplastic cholecystitis |
| SNOMED CT | 65275009 | Acute cholecystitis |
| SNOMED CT | 68368005 | Calculus of common bile duct with chronic cholecystitis |
| SNOMED CT | 71745007 | Calculus of bile duct with chronic cholecystitis without obstruction |
| SNOMED CT | 72053008 | Calculus of common bile duct with acute cholecystitis without obstruction |
| SNOMED CT | 72533002 | Acute gangrenous cholecystitis |
| SNOMED CT | 74091007 | Calculus of common bile duct with chronic cholecystitis without obstruction |
| SNOMED CT | 76581006 | Cholecystitis |
| SNOMED CT | 81115009 | Calculus of common bile duct with acute cholecystitis with obstruction |
| SNOMED CT | 89251007 | Calculus of common bile duct with acute cholecystitis |
| SNOMED CT | 89628003 | Acute suppurative cholecystitis |
| SNOMED CT | 91316003 | Calculus of bile duct with chronic cholecystitis |
| SNOMED CT | 95558008 | Pneumocholecystitis |
| SNOMED CT | 95559000 | Chronic cholecystitis with calculus |
| SNOMED CT | 1269314004 | Perforation of gallbladder due to chronic cholecystitis with calculus |
| SNOMED CT | 197410004 | Acute angiocholecystitis |
| SNOMED CT | 73125001 | Empyema of gallbladder |
| SNOMED CT | 78754002 | Angiocholecystitis |

##### 0.7.3 Cholelithiasis

Table 25: Concept codes used to identify cholelithiasis adverse events.

| CodeSystem | ConceptCode | ConceptName |
|---|---|---|
| SNOMED CT | 25924004 | Calculus of gallbladder with cholecystitis |
| SNOMED CT | 29484002 | Cholelithiasis AND cholecystitis without obstruction |

Continued on next page

**Table25 – continued from previous page**

| CodeSystem | ConceptCode | ConceptName |
|---|---|---|
| SNOMED CT | 197377009 | Gallbladder calculus with acute cholecystitis and no obstruction |
| SNOMED CT | 197378004 | Gallbladder calculus with acute cholecystitis and obstruction |
| SNOMED CT | 38871000119104 | Acute and chronic cholecystitis co-occurrent and due to calculus of gallbladder and bile duct |
| SNOMED CT | 1084931000119109 | Chronic cholecystitis due to cholelithiasis co-occurrent with choledocholithiasis |
| SNOMED CT | 50450007 | Gallstone AND cholecystitis with obstruction |
| SNOMED CT | 699050007 | Calculus of gallbladder with acute and chronic cholecystitis |
| SNOMED CT | 715577009 | Low phospholipid associated cholelithiasis |
| SNOMED CT | 266474003 | Biliary calculus |
| SNOMED CT | 59771005 | Calculus of gallbladder with acute cholecystitis |
| SNOMED CT | 70342003 | Cholelithiasis without obstruction |
| SNOMED CT | 77528005 | Cholelithiasis with obstruction |
| SNOMED CT | 1356007 | Calculus of common duct with obstruction |
| SNOMED CT | 4661003 | Calculus of bile duct with obstruction |
| SNOMED CT | 6087002 | Calculus of hepatic duct with obstruction |
| SNOMED CT | 7290007 | Impacted gallstone of cystic duct |
| SNOMED CT | 12932003 | Calculus of common bile duct with chronic cholecystitis with obstruction |
| SNOMED CT | 15082003 | Calculus of bile duct with chronic cholecystitis with obstruction |
| SNOMED CT | 19335008 | Calculus of cystic duct with acute cholecystitis |
| SNOMED CT | 1234821002 | Primary intrahepatic lithiasis |
| SNOMED CT | 27123005 | Biliary sludge |
| SNOMED CT | 27697003 | Calculus of hepatic duct without obstruction |
| SNOMED CT | 30093007 | Calculus of bile duct |
| SNOMED CT | 1269314004 | Perforation of gallbladder due to chronic cholecystitis with calculus |
| SNOMED CT | 94611000119105 | Gallbladder calculus during pregnancy |
| SNOMED CT | 1162777000 | Obstruction of ampulla of Vater by calculus |
| SNOMED CT | 197389006 | Bile duct calculus with acute cholecystitis |
| SNOMED CT | 197391003 | Bile duct calculus with acute cholecystitis and obstruction |
| SNOMED CT | 197402000 | Calculus of bile duct with cholangitis |
| SNOMED CT | 235919008 | Gallstone |
| SNOMED CT | 235923000 | Retained bile duct stone |
| SNOMED CT | 168036006 | O/E: gallstone |
| SNOMED CT | 168037002 | O/E: cholesterol gallstone |
| SNOMED CT | 168038007 | O/E: pigment gallstone |
| SNOMED CT | 722869007 | Calculus of gallbladder without cholecystitis or cholangitis |
| SNOMED CT | 735467004 | Cholangitis co-occurrent and due to calculus of gallbladder |
| SNOMED CT | 735734002 | Calculus of bile duct without inflammation of biliary tract |

Continued on next page

**Table25 – continued from previous page**

| CodeSystem | ConceptCode | ConceptName |
|---|---|---|
| SNOMED CT | 830074003 | Cholesterol calculus of gallbladder |
| SNOMED CT | 830075002 | Pigment calculus of gallbladder |
| SNOMED CT | 40331000119107 | Acute cholecystitis due to biliary calculus |
| SNOMED CT | 435121000124109 | Calculus of gallbladder and bile duct with cholecystitis |
| SNOMED CT | 435131000124107 | Calculus of gallbladder and bile duct with acute cholecystitis |
| SNOMED CT | 435161000124103 | Calculus of gallbladder and bile duct with acute and chronic cholecystitis |
| SNOMED CT | 435211000124107 | Calculus of common bile duct with acute pancreatitis |
| SNOMED CT | 10754781000119108 | Gallbladder calculus in mother complicating childbirth |
| SNOMED CT | 711564007 | Cholecystolithiasis with obstruction |
| SNOMED CT | 718309005 | Calculus of common bile duct without obstruction |
| SNOMED CT | 307132003 | Common bile duct calculus |
| SNOMED CT | 312110005 | Gallbladder and bile duct calculi |
| SNOMED CT | 314949006 | Perforated calculous chronic cholecystitis |
| SNOMED CT | 413197000 | Calculus of cystic stump |
| SNOMED CT | 36483003 | Calculus of bile duct with cholecystitis |
| SNOMED CT | 49341008 | Calculus of cystic duct with cholecystitis |
| SNOMED CT | 55479002 | Impacted gallstone of gallbladder |
| SNOMED CT | 59049008 | Calculus of hepatic duct |
| SNOMED CT | 60127009 | Calculus of bile duct with acute cholecystitis without obstruction |
| SNOMED CT | 62809003 | Calculus of cystic duct with obstruction |
| SNOMED CT | 65082000 | Calculus of cystic duct |
| SNOMED CT | 68368005 | Calculus of common bile duct with chronic cholecystitis |
| SNOMED CT | 68394001 | Calculus of bile duct without obstruction |
| SNOMED CT | 68601009 | Calculus of cystic duct without obstruction |
| SNOMED CT | 71745007 | Calculus of bile duct with chronic cholecystitis without obstruction |
| SNOMED CT | 72053008 | Calculus of common bile duct with acute cholecystitis without obstruction |
| SNOMED CT | 74091007 | Calculus of common bile duct with chronic cholecystitis without obstruction |
| SNOMED CT | 81115009 | Calculus of common bile duct with acute cholecystitis with obstruction |
| SNOMED CT | 1162565008 | Calculus of intrahepatic bile duct |
| SNOMED CT | 89251007 | Calculus of common bile duct with acute cholecystitis |
| SNOMED CT | 91316003 | Calculus of bile duct with chronic cholecystitis |
| SNOMED CT | 95559000 | Chronic cholecystitis with calculus |

##### 0.7.4 Gastroenteritis

Table 26: Concept codes used to identify gastroenteritis adverse events.

| CodeSystem | ConceptCode | ConceptName |
|---|---|---|
| SNOMED CT | 12463005 | Infectious gastroenteritis |
| SNOMED CT | 12574004 | Noninfectious gastroenteritis |
| SNOMED CT | 24789006 | Viral gastroenteritis due to Norwalk-like agent |
| SNOMED CT | 25374005 | Gastroenteritis |
| SNOMED CT | 26006005 | Allergic gastroenteritis |
| SNOMED CT | 236063005 | Adenoviral gastroenteritis |
| SNOMED CT | 713186008 | Gastroenteritis caused by drug |
| SNOMED CT | 266081001 | Colitis, enteritis and gastroenteritis presumed infectious |
| SNOMED CT | 266451002 | Allergic gastroenteritis and colitis |
| SNOMED CT | 359613008 | Acute infectious nonbacterial gastroenteritis |
| SNOMED CT | 36789003 | Acute infective gastroenteritis |
| SNOMED CT | 69776003 | Acute gastroenteritis |
| SNOMED CT | 111843007 | Viral gastroenteritis |
| SNOMED CT | 27858009 | Clostridial gastroenteritis |
| SNOMED CT | 33687004 | Dietetic gastroenteritis |
| SNOMED CT | 196731005 | Gastroduodenitis |
| SNOMED CT | 197011006 | Radiation gastroenteritis |
| SNOMED CT | 240332005 | Infantile gastroenteritis |
| SNOMED CT | 735459007 | Gastrointestinal hypersensitivity caused by food |
| SNOMED CT | 765756007 | Benign infantile seizure with mild gastroenteritis syndrome |
| SNOMED CT | 10628871000119101 | Gastroenteritis due to influenza |
| SNOMED CT | 10628911000119103 | Gastroenteritis due to Influenza A virus |
| SNOMED CT | 415822001 | Viral gastroenteritis caused by Rotavirus |
| SNOMED CT | 421454008 | Infectious gastroenteritis with AIDS (acquired immunodeficiency syndrome) |
| SNOMED CT | 421983003 | Noninfectious gastroenteritis with AIDS (acquired immunodeficiency syndrome) |
| SNOMED CT | 142961000119108 | Gastroenteritis caused by H1N1 influenza |
| SNOMED CT | 707222009 | Epidemic gastroenteritis |
| SNOMED CT | 707352004 | Institution-acquired gastroenteritis |
| SNOMED CT | 713180002 | Ischemic gastroenteritis |
| SNOMED CT | 713570009 | Infectious gastroenteritis co-occurrent with human immunodeficiency virus infection |
| SNOMED CT | 308119005 | Infantile viral gastroenteritis |
| SNOMED CT | 312088007 | Fungal gastroenteritis |
| SNOMED CT | 359651008 | Nonbacterial gastroenteritis of infant |
| SNOMED CT | 359804008 | Eosinophilic gastroenteritis |
| SNOMED CT | 409506009 | Hemorrhagic gastroenteritis |
| SNOMED CT | 416482004 | Food-borne gastroenteritis |
| SNOMED CT | 283876006 | Astrovirus gastroenteritis |
| SNOMED CT | 283877002 | Calicivirus gastroenteritis |

Continued on next page

**Table26 – continued from previous page**

| CodeSystem | ConceptCode | ConceptName |
|---|---|---|
| SNOMED CT | 35974005 | Cryptococcal gastroenteritis |
| SNOMED CT | 51338006 | Eosinophilic gastroenteropathy with food sensitivity |
| SNOMED CT | 53954009 | Eosinophilic gastroenteropathy with predominant sub-serosal disease |
| SNOMED CT | 57419008 | Gastroenteritis presumed infectious |
| SNOMED CT | 63924002 | Eosinophilic gastroenteropathy with predominant mucosal disease |
| SNOMED CT | 66160001 | Cryptosporidial gastroenteritis |
| SNOMED CT | 67375008 | Eosinophilic gastroenteropathy with predominant muscle layer disease |
| SNOMED CT | 71583005 | Toxic gastroenteritis |
| Truveta | 1215944 | Gastroenteritis due to severe acute respiratory syndrome coronavirus 2 (SARS-CoV-2) |
| SNOMED CT | 1187631004 | Viral gastroenteritis caused by Sapovirus |
| SNOMED CT | 89030000 | Eosinophilic gastroenteropathy with collagen vascular disease |
| SNOMED CT | 109814008 | Acute ulcerative gastroenteritis complicating pneumonia |

##### 0.7.5 Gastroparesis

Table 27: Concept codes used to identify gastroparesis adverse events.

| CodeSystem | ConceptCode | ConceptName |
|---|---|---|
| SNOMED CT | 34140002 | Diabetic gastroparesis |
| SNOMED CT | 235675006 | Gastroparesis syndrome |
| SNOMED CT | 424989000 | Diabetic gastroparesis associated with type 2 diabetes mellitus |
| SNOMED CT | 425159004 | Diabetic gastroparesis associated with type 1 diabetes mellitus |
| SNOMED CT | 713702000 | Gastroparesis due to type 1 diabetes mellitus |
| SNOMED CT | 713703005 | Gastroparesis due to type 2 diabetes mellitus |
| SNOMED CT | 713704004 | Gastroparesis due to diabetes mellitus |
| SNOMED CT | 353821005 | Idiopathic gastroparesis |
| SNOMED CT | 75994008 | Nondiabetic gastroparesis |
| SNOMED CT | 77164002 | Gastroparesis syndrome |
| SNOMED CT | 235676007 | Delayed gastric emptying following procedure |
| ICD9CM | 536.3 | Gastroparesis |
| ICD10CM | K31.84 | Gastroparesis |

##### 0.7.6 Pancreatitis

Table 28: Concept codes used to identify pancreatitis adverse events.

| CodeSystem | ConceptCode | ConceptName |
|---|---|---|
| SNOMED CT | 4399003 | Acute hemorrhagic pancreatitis |
| SNOMED CT | 7881005 | Acute necrotizing pancreatitis |
| SNOMED CT | 8551005 | Recurrent acute pancreatitis |
| SNOMED CT | 15974001 | Chronic pancreatitis |
| SNOMED CT | 24407009 | Suppurative pancreatitis |
| SNOMED CT | 197456007 | Acute pancreatitis |
| SNOMED CT | 197457003 | Acute pancreatitis unspecified |
| SNOMED CT | 197458008 | Acute recurrent pancreatitis |
| SNOMED CT | 197459000 | Acute hemorrhagic pancreatitis |
| SNOMED CT | 197460005 | Acute suppurative pancreatitis |
| SNOMED CT | 197461009 | Pancreatitis (& [acute NOS]) |
| SNOMED CT | 197462002 | Chronic pancreatitis |
| SNOMED CT | 197565003 | [X]Other chronic pancreatitis |
| SNOMED CT | 233005009 | Painless pancreatitis |
| SNOMED CT | 233870001 | Recurrent pancreatitis |
| SNOMED CT | 234689009 | Relapsing pancreatitis |
| SNOMED CT | 235494005 | Chronic pancreatitis |
| SNOMED CT | 235941008 | Gallstone acute pancreatitis |
| SNOMED CT | 235943006 | Idiopathic acute pancreatitis |
| SNOMED CT | 235944000 | Drug-induced acute pancreatitis |
| SNOMED CT | 235945004 | Acute pancreatitis due to infection |
| SNOMED CT | 235946003 | Viral acute pancreatitis |
| SNOMED CT | 235949005 | Familial acute pancreatitis |
| SNOMED CT | 235951009 | Gallstone chronic pancreatitis |
| SNOMED CT | 235953007 | Idiopathic chronic pancreatitis |
| SNOMED CT | 235954001 | Obstructive chronic pancreatitis |
| SNOMED CT | 235955000 | Drug-induced chronic pancreatitis |
| SNOMED CT | 155834006 | Acute pancreatitis |
| SNOMED CT | 155835007 | Chronic pancreatitis |
| SNOMED CT | 721724009 | Acute on chronic pancreatitis |
| SNOMED CT | 722544004 | Pancreatitis due to pancreatic duct obstruction |
| SNOMED CT | 722871007 | Groove pancreatitis |
| SNOMED CT | 722872000 | Autoimmune pancreatitis type 1 |
| SNOMED CT | 838375006 | Chronic infectious pancreatitis |
| SNOMED CT | 96081000119101 | Acute pancreatitis due to common bile duct calculus |
| SNOMED CT | 435211000124107 | Calculus of common bile duct with acute pancreatitis |
| SNOMED CT | 448542008 | Autoimmune pancreatitis |
| SNOMED CT | 393591004 | Pancreatitis |
| SNOMED CT | 394519007 | Gallstone acute pancreatitis |
| SNOMED CT | 394520001 | Gallstone chronic pancreatitis |
| SNOMED CT | 266476001 | Acute pancreatitis NOS |

Continued on next page

**Table28 – continued from previous page**

| CodeSystem | ConceptCode | ConceptName |
|---|---|---|
| SNOMED CT | 301009006 | Calcific chronic pancreatitis |
| SNOMED CT | 39205007 | Infectious pancreatitis |
| SNOMED CT | 39726008 | Acute pancreatitis |
| SNOMED CT | 44636008 | Interstitial pancreatitis |
| SNOMED CT | 46626002 | Subcutaneous nodular fat necrosis in pancreatitis |
| SNOMED CT | 59154002 | Subacute pancreatitis |
| SNOMED CT | 59329007 | Metabolic pancreatitis |
| SNOMED CT | 75694006 | Pancreatitis |
| SNOMED CT | 95563007 | Gallstone pancreatitis |
| SNOMED CT | 123289004 | Relapsing pancreatitis |
| SNOMED CT | 303002 | Apoplectic pancreatitis |
| SNOMED CT | 10665004 | Mumps pancreatitis |
| SNOMED CT | 1263995002 | Chronic pancreatitis due to gallbladder calculus |
| SNOMED CT | 1197711009 | Acute ischemic pancreatitis |
| SNOMED CT | 234367000 | Pancytopenia with pancreatitis |
| SNOMED CT | 235942001 | Alcohol-induced acute pancreatitis |
| SNOMED CT | 235947007 | Cytomegaloviral pancreatitis |
| SNOMED CT | 235948002 | Postoperative acute pancreatitis |
| SNOMED CT | 235950005 | Traumatic acute pancreatitis |
| SNOMED CT | 235956004 | Familial chronic pancreatitis |
| SNOMED CT | 724540009 | Tropical calcific chronic pancreatitis |
| SNOMED CT | 767291004 | Chronic pancreatitis due to acute alcohol intoxication |
| SNOMED CT | 771446000 | Follicular cholangitis and pancreatitis |
| SNOMED CT | 154211000119108 | Chronic pancreatitis due to chronic alcoholism |
| SNOMED CT | 235952002 | Chronic pancreatitis due to acute alcohol intoxication |
| SNOMED CT | 277537008 | Post-endoscopic retrograde cholangiopancreatography acute pancreatitis |
| SNOMED CT | 399525009 | Inflammation of hepatopancreatic ampulla |
| SNOMED CT | 445507008 | Alcohol-induced pancreatitis |
| SNOMED CT | 1197710005 | Acute pancreatitis due to systemic disease |
| SNOMED CT | 68072000 | Hereditary pancreatitis |
| SNOMED CT | 74973004 | Chronic fibrosing pancreatitis |
| SNOMED CT | 878822006 | Ischemic pancreatitis |
| SNOMED CT | 1197701002 | Type 1 autoimmune pancreatitis with extrapancreatic involvement |
| SNOMED CT | 1197740007 | Autoimmune pancreatitis type 2 |
| ICD10CM | K85 | Acute pancreatitis |
| ICD10CM | K85.0 | Idiopathic acute pancreatitis |
| ICD10CM | K85.00 | Idiopathic acute pancreatitis without necrosis or infection |
| ICD10CM | K85.01 | Idiopathic acute pancreatitis with uninfected necrosis |
| ICD10CM | K85.02 | Idiopathic acute pancreatitis with infected necrosis |
| ICD10CM | K85.1 | Biliary acute pancreatitis |
| ICD10CM | K85.10 | Biliary acute pancreatitis without necrosis or infection |

Continued on next page

**Table28 – continued from previous page**

| CodeSystem | ConceptCode | ConceptName |
|---|---|---|
| ICD10CM | K85.11 | Biliary acute pancreatitis with uninfected necrosis |
| ICD10CM | K85.12 | Biliary acute pancreatitis with infected necrosis |
| ICD10CM | K85.3 | Drug induced acute pancreatitis |
| ICD10CM | K85.30 | Drug induced acute pancreatitis without necrosis or infection |
| ICD10CM | K85.31 | Drug induced acute pancreatitis with uninfected necrosis |
| ICD10CM | K85.32 | Drug induced acute pancreatitis with infected necrosis |
| ICD10CM | K85.8 | Other acute pancreatitis |
| ICD10CM | K85.80 | Other acute pancreatitis without necrosis or infection |
| ICD10CM | K85.81 | Other acute pancreatitis with uninfected necrosis |
| ICD10CM | K85.82 | Other acute pancreatitis with infected necrosis |
| ICD10CM | K85.9 | Acute pancreatitis, unspecified |
| ICD10CM | K85.90 | Acute pancreatitis without necrosis or infection, unspecified |
| ICD10CM | K85.91 | Acute pancreatitis with uninfected necrosis, unspecified |
| ICD10CM | K85.92 | Acute pancreatitis with infected necrosis, unspecified |
| ICD10CM | K86.0 | Alcohol-induced chronic pancreatitis |
| ICD10CM | K86.1 | Other chronic pancreatitis |
| ICD10CM | K85.2 | Alcohol induced acute pancreatitis |
| ICD10CM | K85.20 | Alcohol induced acute pancreatitis without necrosis or infection |
| ICD10CM | K85.21 | Alcohol induced acute pancreatitis with uninfected necrosis |
| ICD10CM | K85.22 | Alcohol induced acute pancreatitis with infected necrosis |
| ICD9CM | 577.0 | Acute pancreatitis |
| ICD9CM | 577.1 | Chronic pancreatitis |

### Contents
